# Population-scale analysis of common and rare genetic variation associated with hearing loss in adults

**DOI:** 10.1101/2021.09.27.21264091

**Authors:** Kavita Praveen, Lee Dobbyn, Lauren Gurski, Ariane H. Ayer, Jeffrey Staples, Shawn Mishra, Yu Bai, Alexandra Kaufman, Arden Moscati, Christian Benner, Esteban Chen, Siying Chen, Alexander Popov, Janell Smith, GHS-REGN DiscovEHR collaboration, Regeneron Genetics Center, Decibel-REGN collaboration, Olle Melander, Marcus Jones, Jonathan Marchini, Suganthi Balasubramanian, Brian Zambrowicz, Meghan Drummond, Aris Baras, Goncalo R. Abecasis, Manuel A. Ferreira, Eli A. Stahl, Giovanni Coppola

## Abstract

Understanding the genetic underpinnings of disabling hearing loss, which affects ∼466 million people worldwide, can provide avenues for new therapeutic target development. We performed a genome-wide association meta-analysis of hearing loss with 125,749 cases and 469,497 controls across five cohorts, including UK Biobank, Geisinger DiscovEHR, the Malmö Diet and Cancer Study, Mount Sinai’s Bio*Me* Personalized Medicine Cohort, and FinnGen. We identified 53 loci affecting hearing loss risk, 15 of which are novel, including common coding variants in *COL9A3 and TMPRSS3*. Through exome-sequencing of 108,415 cases and 329,581 controls from the same cohorts, we identified hearing loss associations with burden of rare coding variants in *FSCN2* (odds ratio [OR] = 1.14, *P* = 1.9 × 10^−15^) and burden of predicted loss-of-function variants in *KLHDC7B* (OR = 2.14, *P* = 5.2 × 10^−30^). We also observed single-variant and gene-burden associations with 11 genes known to cause Mendelian forms of hearing loss, including an increased risk in heterozygous carriers of mutations in the autosomal recessive hearing loss genes *GJB2* (Gly12fs; OR = 1.21, *P* = 4.2 × 10^−11^) and *SLC26A5* (gene burden; OR = 1.96, *P* = 2.8 × 10^−17^). Our results suggest that loss of KLHDC7B function increases risk for hearing loss, and show that Mendelian hearing loss genes contribute to the burden of hearing loss in the adult population, suggesting a shared etiology between common and rare forms of hearing loss. This work illustrates the potential of large-scale exome sequencing to elucidate the genetic architecture of common traits in which risk is modulated by both common and rare variation.

## INTRODUCTION

The loss of hearing can have a debilitating impact on quality of life, requiring major adjustments to day-to-day activities. Significant comorbidities are also associated with hearing loss including social isolation, depression, cognitive impairment, and dementia, which further deteriorate quality of life^1^. Disabling hearing loss is common, with ∼466 million people affected worldwide (World Health Organization: https://www.who.int/health-topics/hearing-loss). While existing technologies – such as hearing aids and cochlear implants – can ameliorate hearing loss, their use is limited by barriers to access, including cost, health policies and regulations, and social stigma associated with device use^2^. Furthermore, while these assistive devices typically provide some benefit, they do not address the chief complaint associated with acquired hearing loss: lack of hearing clarity, particularly in social and work environments (https://www.hearingloss.org/hlaa-pfdd/). The Lancet Commission on dementia prevention, intervention and care has identified untreated hearing loss in middle age as the top modifiable risk factor for dementia, but it is estimated that 67-86% of adults who may benefit from hearing aids do not use them^1,3^. These challenges, combined with a lack of therapeutics to stop or slow hearing loss progression, have contributed to its status as a growing global health issue. Novel therapies based on genetic evidence, therefore, will be crucial in addressing this unmet need.

Hearing loss affects individuals of all ages, but its prevalence increases with age. Approximately 1-2/1,000 babies are born with hearing loss^4^. Mutations in over 150 genes (hereditaryhearingloss.com) account for over 50% of the cases. While autosomal recessive hearing loss is generally pre-lingual and non-progressive, autosomal dominant forms are mostly post-lingual (including adult onset) and progressive. The prevalence of hearing loss increases to 2.8/1,000 in primary school-age children and 3.5/1,000 in adolescents^4,5^. The National Institute on Deafness and other Communication Diseases calculates that by the age of 45 ∼2% of individuals have a disabling hearing loss, and this number increases to 50% in individuals over the age of 75. This increase in prevalence with age reflects a combination of late-onset hearing loss mutations, the cumulative effects of environmental factors such as exposure to noise and ototoxic drugs and, in aging individuals, the degenerative effects of age on the cochlea. These genetic and environmental insults primarily damage the structures of the inner ear, resulting in sensorineural hearing loss^6,7^.

Heritability estimates for age-related hearing loss range as high as 36 and 70%^8–12^ suggesting that genetics, along with environmental factors, play a significant role in determining an individual’s risk for developing hearing loss. Genome-wide association studies (GWAS) of hearing loss in adults have identified 61 common variant loci associated with the trait in Europeans^13–18^. While the majority of these studies have established a common variant contribution to adult hearing loss, there are few reports addressing rare and low-frequency variant contribution^19^.

Recently, Ivarsdottir et al^18^ published association results with hearing loss on ∼50K Icelandic individuals with whole-genome sequence and ∼50K individuals from UKB with exome sequence data, with imputation of larger samples into these variant sets. We have now expanded the rare variant analysis to exome-sequences from ∼295K individuals in UKB and ∼143K individuals from three other datasets. Here, we report findings from genome- and exome-wide association meta analyses with a total of 125,749 cases and 469,497 controls. Our analyses have identified 15 novel susceptibility loci and 15 rare variant associations that provide novel insights into the biology of hearing loss in adults.

## RESULTS

To study common variants, we performed association meta-analyses using genotyping and imputation across five cohorts: Geisinger DiscovEHR study (GHS), the Malmö Diet and Cancer study from Malmö, Sweden (MALMO), Mount Sinai’s Bio*Me* Personalized Medicine Cohort from Mount Sinai Health System, New York (SINAI), UK Biobank (UKB) and an additional study from Finland, FinnGen, for a total of 125,749 cases and 469,497 controls. To study rare and ultra-rare variants, we also generated exome sequence data and performed combined GWAS and exome-wide association study (ExWAS) on a subset of 108,415 cases and 329,581 controls across GHS, MALMO, SINAI, and UKB. Phenotypes were derived from ICD-10 diagnosis codes in GHS, MALMO, SINAI and FinnGen, and combined self-report and ICD-10 codes in UKB (see Methods and Supplementary Table 1 for details). Our genome-wide association analyses included 15,881,489 variants with frequency > 0.1% that were genotyped or imputed with r2 > 0.3 in at least one study, and 2,923,124 coding or essential splice site variants with minor allele count at least five from exome sequencing (111,588 of which overlapped with the imputed).

### Novel common variant associations

We identified 53 independent loci harboring genome-wide significant (*P* < 5 × 10^−8^) common (MAF ≥ 1%) variants associated with hearing loss (Figure 1A, Supplementary Table 2, Supplementary Figure 1), 38 of which are shared with the previously reported 61 hearing loss-associated loci. We observed a genomic control lambda_GC = 1.36 but an LD score regression intercept 1.054 (standard error 0.008), indicating that inflation was largely due to polygenic signal for the hearing loss phenotype.

**Figure 1:**
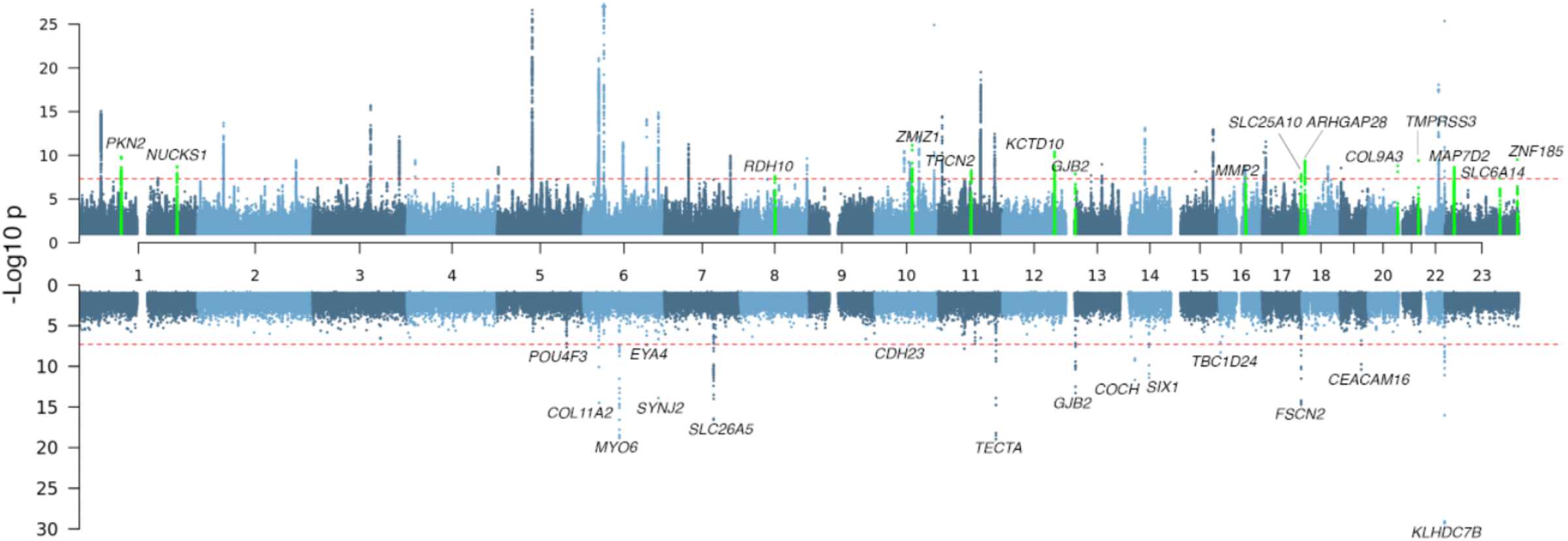
Common (MAF ≥ 0.01) variant (A), and rare (MAF < 0.01) coding single-variant and gene burden (B) associations with hearing loss. The gene labels refer to the nearest gene and colored in green are loci that have not previously been associated with hearing loss in the common variant results (A).

Among the lead variants at the 15 novel loci (Figure 1A, Table 1, Supplementary Figure 2) is rs117887149 that maps close to *GJB2* (Supplementary Figure 2H), which is a predominant cause of congenital hearing loss. While majority of the lead variants in the novel loci lay in intergenic or downstream/upstream regions of genes, at two loci they were within the introns of the following genes: *KCTD10*, a member of the potassium channel tetramerization domain family that is implicated in cardiac development^20,21^ and *MAP7D2*, an axonal cargo transport protein predominantly expressed in the brain^22^.

**Table 1.**
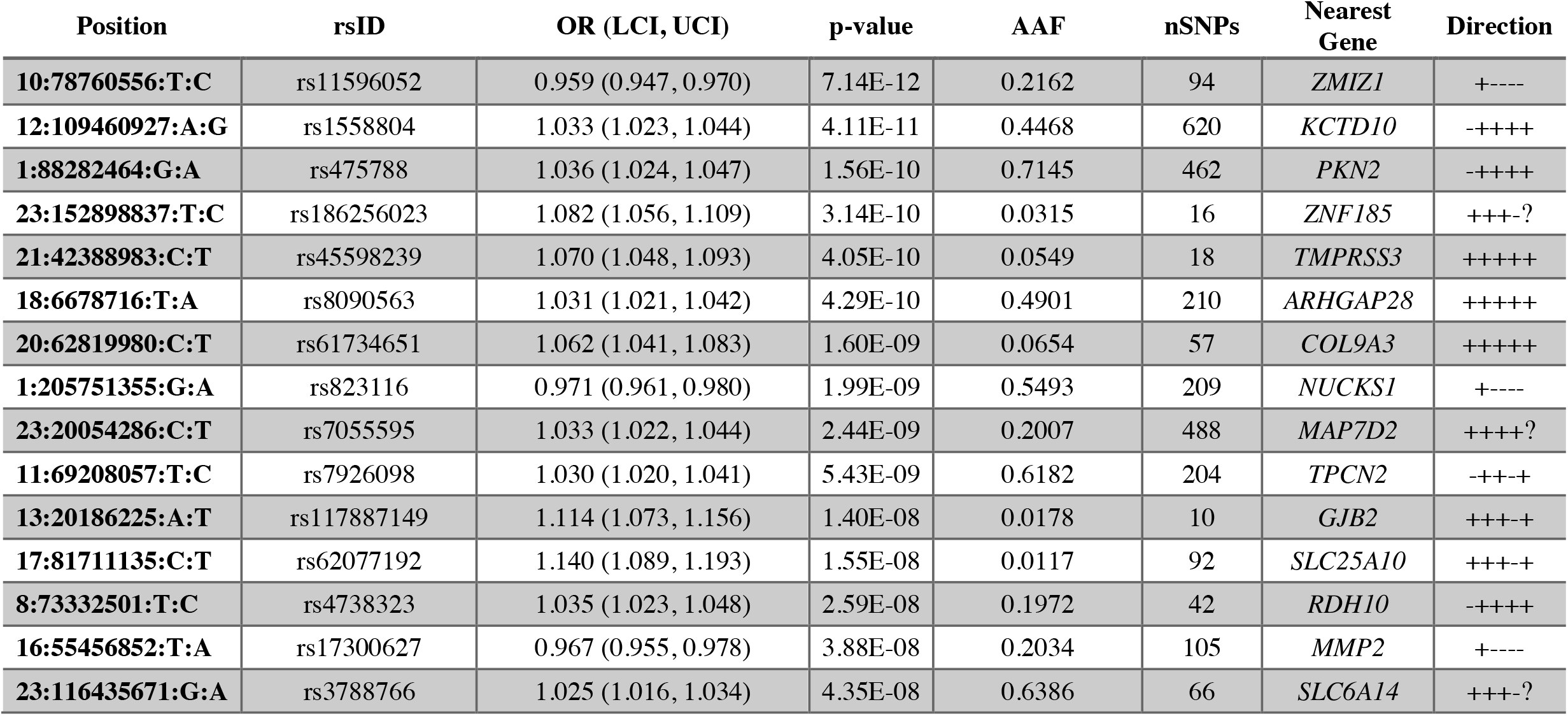
Lead variants at the 15 novel loci associated with hearing loss. The alternate allele frequency (AAF) refers to the allele listed second in the ‘Position’ column. The direction of the effect in each single study (in the order: MALMO, UKB, GHS, SINAI, FinnGen) in the meta-analysis is given in the ‘Direction’ column where ‘+’ indicates increased risk, ‘-’ indicates decreased risk, and ‘?’ indicates that the variant was not present or tested. The nSNPs column indicates the number of genome-wide significant SNPs within each locus.

Top variants at two other novel loci were missense changes and thus also implicate specific genes: in *COL9A3* (Arg103Trp; MAF = 0.066) and *TMPRSS3* (Ala90Thr; MAF = 0.055). Mutations in *COL9A3*, which is highly expressed in the ear, have been implicated in the autosomal recessive Stickler syndrome, in which hearing loss is prominent^23^, and tentatively in non-syndromic hearing loss^24,25^. High-throughput mouse knockout characterization from the International Mouse Phenotyping Consortium (IMPC) indicates that *Col9a3*-null mice also have hearing loss^26^. *TMPRSS3* is a type II membrane serine protease that localizes to the endoplasmic reticulum and plasma membranes, and is expressed in hair cells and supporting cells in the organ of Corti, the spiral ganglion, and the stria vascularis in the ear^27–29^. Mutations in *TMPRSS3* cause congenital and childhood-onset autosomal recessive hearing loss^30^ but there is also evidence for hearing deficits in heterozygous carriers^18^.

The number of associated variants at the 50 autosomal loci ranged from eight to 1,326. In order to assess the presence of multiple independent causal variants at each locus, we ran conditional analyses using GCTA-COJO^31^, which indicated the presence of secondary association signals (joint *P* < 10^−5^) at eight loci (Supplementary Table 3). We also ran FINEMAP Bayesian causal variant inference^32^ to prioritize associated variants as credibly causal at thirty loci that were genome-wide significant in RGC data (excluding FinnGen). The COJO and FINEMAP methods disagreed on the presence of single vs. multiple association signals within five loci, three of which showed subthreshold (10^−5^ < *P* < 10^−4^) secondary associations in COJO. FINEMAP prioritized ten or fewer variants in top causal variant 95% credible sets for the top causal variant at eleven loci, including single putatively causal variants at four loci (Supplementary Table 3): missense variants in *CDH23* (Ala371Thr; rs143282422) and *KLHDC7B* (Val504Met; rs36062310), and intronic variants in *CTBP2* (rs10901863) and *PAFAH1B1* (rs12938775).

### Colocalization with GTEx eQTLs identifies candidate genes driving GWAS signals

To identify genes for which expression regulation might drive the observed association signals, we tested for colocalization of our hearing loss-associated loci with expression quantitative trait loci (eQTL) data for 48 tissues from the Genotype-Tissue Expression (GTEx) project (see URLs) using coloc2^33^. Across all GTEx eQTL tissues tested, we identified 19 genes mapping to 15 loci with evidence (posterior probability of colocalization, PPH4 ≥ 0.5) for colocalization between the hearing loss association and an eQTL signal, in at least one tissue (Supplementary Table 4). Only two of the 15 loci with GWAS-eQTL overlap had multiple eQTL signals for more than one gene (*NUCKS1* and *RAB29* in locus 1-4; *ACADVL, DLG4, CTDNEP1* and *CLDN7* in locus 17-2) making it difficult to prioritize causal genes at these loci based on eQTL data. Since GTEx did not include tissues from the ear, we used single-cell RNA sequencing data that were generated in-house from mouse cochleae to check the expression of the 19 genes in the ear (Supplementary Table 5). Thirteen of the 19 genes showed evidence of expression across 26 inner-ear cell-types and, of these, 12 were expressed in the hair cells. While the majority of genes showed broad expression across the 26 cell types, we noted a subset that were specific to only a few, including *CRIP3* in inner and outer hair cells, and *TCF19* in neurons and immune cells (Supplementary Tables 4 & 5).

### Rare-variant association analysis identifies novel large-effect hearing loss variants in known hearing loss genes

We identified significant (*P* < 5 × 10^−8^) rare variant (MAF < 1%) associations in 25 genes, 15 of which had nonsynonymous variant or gene burden associations (Figure 1B, Table 2, Supplementary Tables 6 & 7). Five of the 15 genes also had significant common variant associations within 1 Mb *(KLHDC7B, SYNJ2, GJB2, EYA4*, and *CDH23)*. After conditioning on independent common variants at each locus (Supplementary Tables 6, 7 & 8), the rare variant and gene burdens remained associated with hearing loss (maximum conditional *P* ≤ 3 × 10^−5^) except for the *CDH23* Asn1103Ser association (conditional *P* = 0.79). Of note, the lead common variant in *CDH23* is also a missense (Ala371Thr) variant (MAF = 1.1%) and is pinpointed by FINEMAP as the only causal variant at that locus with high confidence. Overall, our conditional analyses suggest that rare variant association signals are usually independent of nearby common variant associations.

**Table 2:**
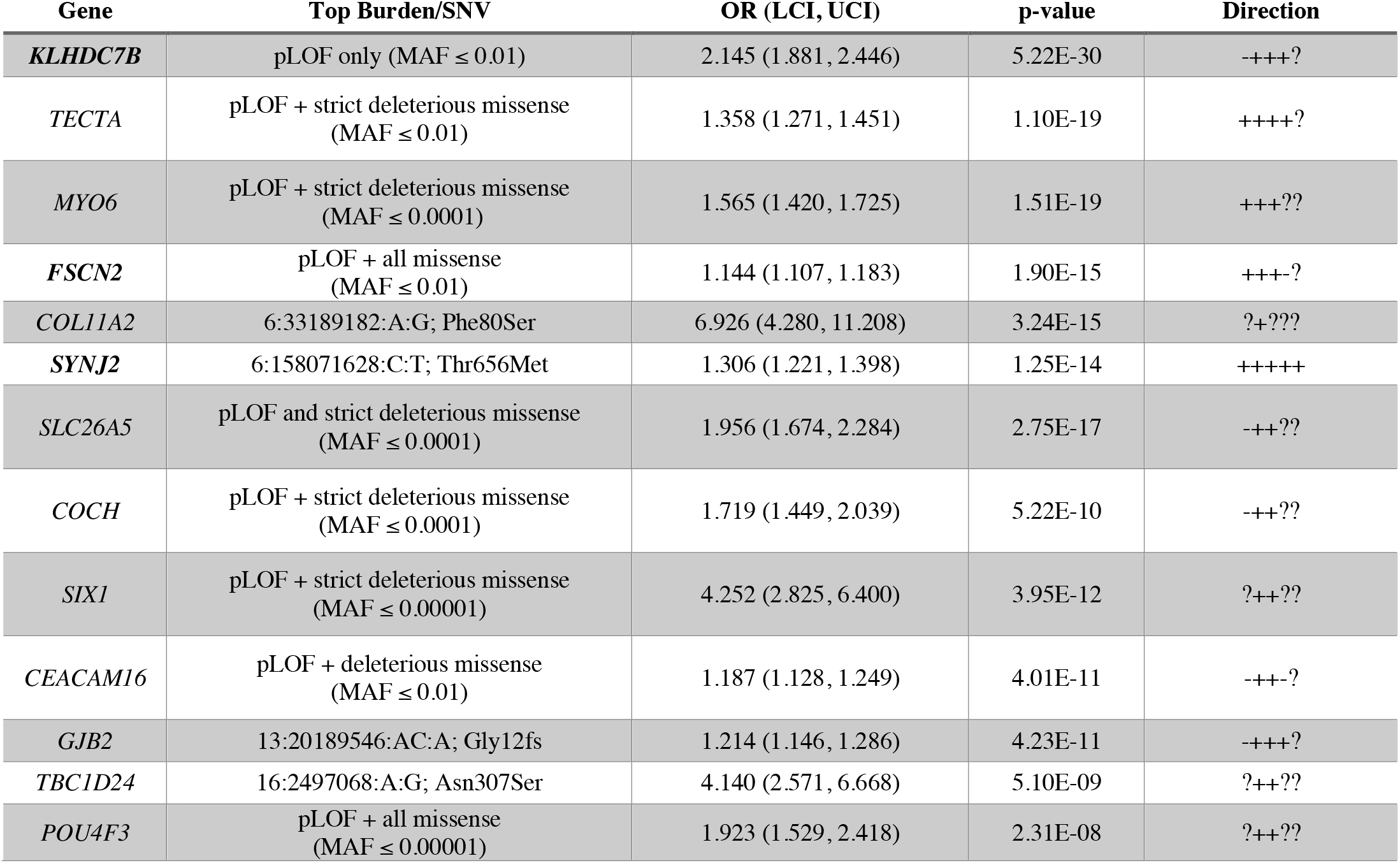
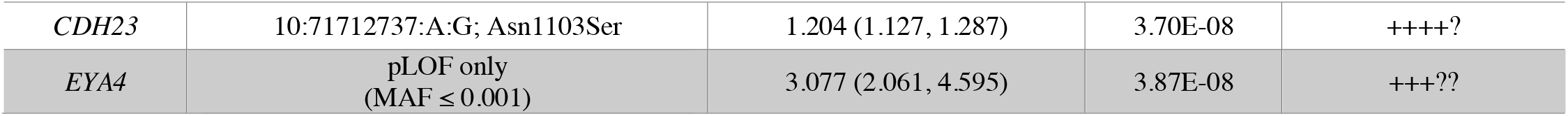
Nonsynonymous rare (minor allele frequency, MAF < 0.01) variants and gene burdens associated (*P* < 5 × 10^−8^) with hearing loss in meta-analysis. The direction of the effect in each single study (in the order: MALMO, UKB, GHS, SINAI, FinnGen) in the meta-analysis is given in the ‘Direction’ column where ‘+’ indicates increased risk, ‘-’ indicates decreased risk and ‘?’ indicates that the variant was not present or tested.

#### Associations with Mendelian hearing loss genes

Of the 14 genes with independent, rare nonsynonymous and/or burden associations, 11 were previously identified as causes of Mendelian forms of hearing loss. These include associations with two genes (*GJB2* and *SLC26A5)* that cause recessive hearing loss, burden associations in seven genes (*MYO6, COCH, TECTA, SIX1, CEACAM16, POU4F3*, and *EYA4*) and single-variant associations in two genes (*TBC1D24* and *COL11A2*) that cause autosomal dominant hearing loss (reviewed in Shearer et. al., 1999^34^). In *MYO6* and *COCH*, we also observed genome-wide significant single-variant associations of large effect size (*MYO6* His246Arg, OR=30.7; *COCH* Cys542Phe, OR=81.4; Supplementary Table 6). Both variants have previously been characterized as pathogenic in family-based genetic analyses^35,36^. Only a minority of variants included in burden tests have been classified as pathogenic by ClinVar (Supplementary Table 9) suggesting that our analysis has detected additional novel, risk-associated variants with variable penetrance in Mendelian hearing loss genes. We also identified single-variant associations in two genes that cause autosomal dominant hearing loss: *TBC1D24* Asn307Ser, also recently reported by Ivarsdottir et al^18^, was implicated as pathogenic in two unrelated families^37^, and *COL11A2* Phe80Ser, which has not yet been classified as pathogenic but is predicted deleterious, lies in a domain (Laminin G-like/NC4) that harbors other mutations causing non-syndromic hearing loss^38^. We performed our analysis under an additive model, which assumes risk effects in heterozygous carriers as well as homozygotes; therefore, it is not surprising to see that the majority of Mendelian genes (9/11) identified can cause hearing loss in heterozygote carriers. However, we also detect associations in two genes that have previously been implicated in recessive hearing loss: *GJB2* Gly12fs (OR = 1.21; *P* = 4 × 10^−11^), and *SLC26A5* Leu46Pro (OR = 1.3; *P* = 3 × 10^−14^) as well as the burden of *SLC26A5* predicted loss-of-function (pLOF) and strict deleterious missense variants (excluding Leu46Pro) (OR = 1.96; *P* = 3 × 10^−17^) (Figure 2). We also observed a suggestive association with *GJB2* Leu90Pro (OR = 1.51, *P* = 4.4 × 10^−5^), another known pathogenic variant in recessive hearing loss. The associations persisted after excluding homozygous carriers and compound heterozygous carriers of rare, coding variants in these genes (Supplementary Table 10), suggesting a previously unappreciated increase in risk for hearing loss in heterozygous carriers of loss-of-function *GJB2*, and missense and pLOF *SLC26A5* variants.

**Figure 2:**
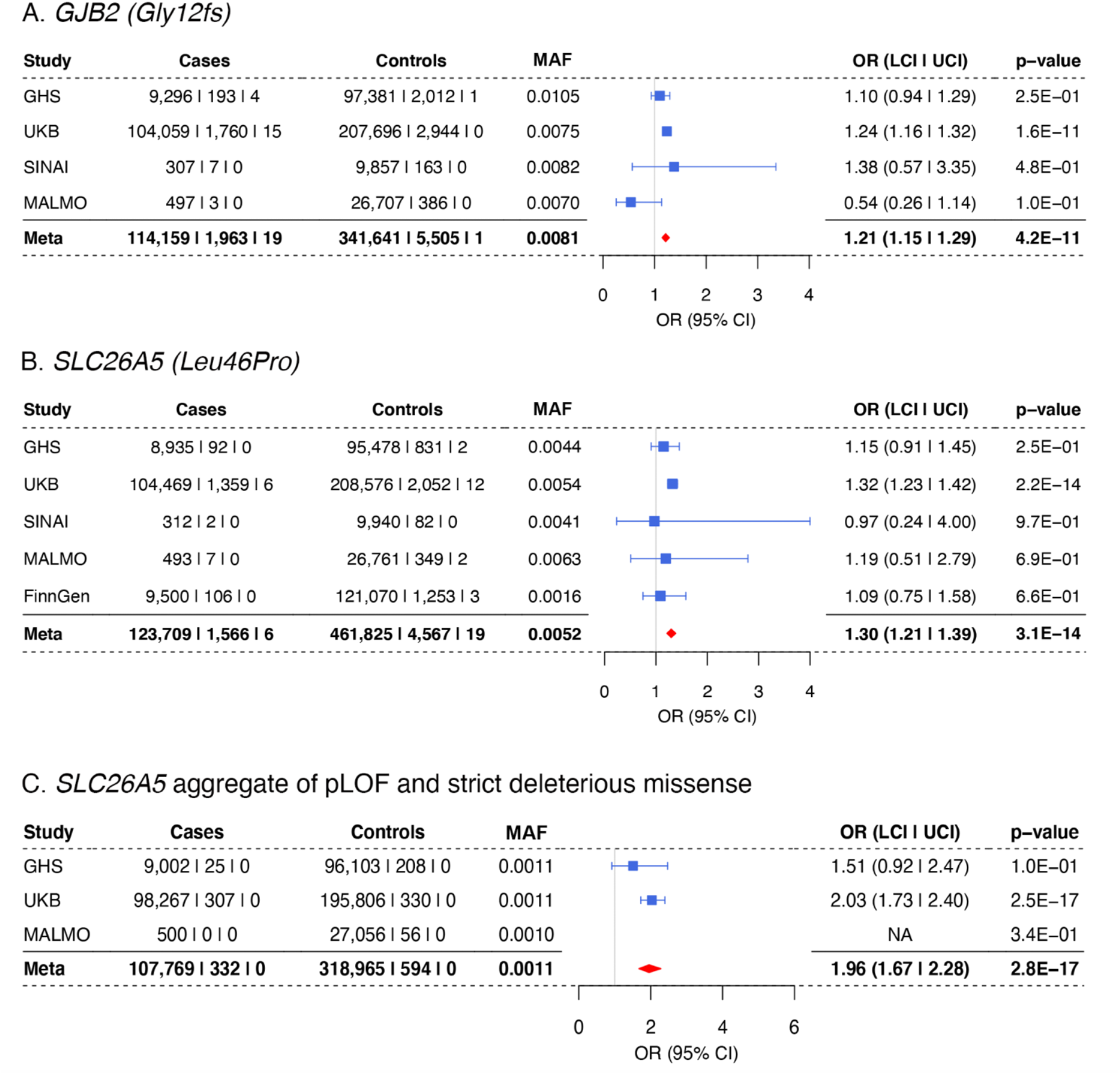
Association of *GJB2* Gly12fs (A), *SLC26A5* Leu46Pro (B) and *SLC26A5* pLOF and strict deleterious missense, MAF ≤ 0.0001 (C) with hearing loss.

#### *Rare variant associations in KLHDC7B, FSCN2*, and *SYNJ2*

The most significant rare coding association in our analysis was the aggregate of 68 pLOF variants (43 frameshift, 23 stop-gain and 2 stop-loss, (Supplementary Table 9)) in *KLHDC7B* (Kelch-like domain containing 7B), with an approximately two-fold increase in risk for hearing loss (OR = 2.14, *P* = 5 × 10^−30^). The main contributors to the gene burden were two frameshift variants, Gly302fs and Lys181fs, that are predicted to truncate the protein near the start of the Kelch domains. These variants were also significantly associated in single-variant tests (Figure 3), and Gly302fs was recently reported in an analysis that included UKB^18^. The association with pLOF variants remained significant after repeating the pLOF burden test conditioning on Gly302fs and Lys181fs (*P* = 8 × 10^−8^; Supplementary Table 7). In addition, we observed a common (MAF = 4%) missense variant (Val504Met) within the last Kelch domain of *KLHDC7B* associated with increased risk for hearing loss (OR = 1.14, *P* = 4 × 10^−26^). This variant was also prioritized as the sole causal variant at this locus by FINEMAP (Supplementary Table 3).

**Figure 3:**
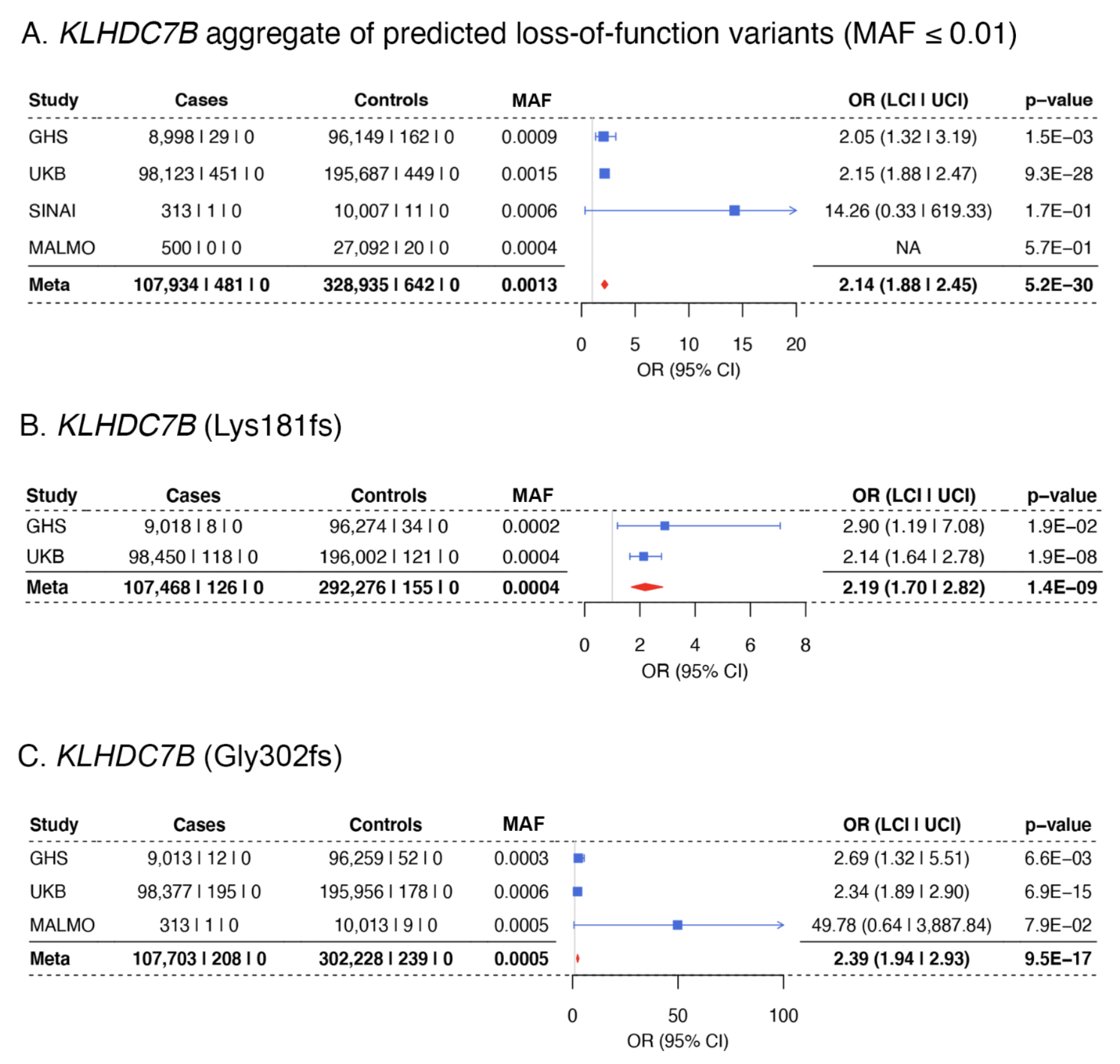
Association of rare variants in *KLHDC7B* with increased risk for hearing loss in meta-analysis. **A**. Association of an aggregate of predicted loss of function variants (MAF ≤ 0.01) in *KLHDC7B* with risk for hearing loss. **B and C**. Two variants were the predominant contributors to the *KLHDC7B* loss-of-function gene burden aggregate, Lys181fs (B) and Gly302fs (C).

We also identified rare coding associations in fascin actin-bundling protein 2 (*FSCN2)* and synaptojanin 2 (*SYNJ2)* with increased risk for hearing loss, both of which were recently observed in UKB^18^ (Figure 4). In *FSCN2*, an actin cross-linking protein^39–41^, an aggregate of pLOF and deleterious missense variants was associated with hearing loss, with majority of the carriers in the burden having the His138Tyr variant. Conditioning on His138Tyr attenuated the significance of the burden test (*P* = 3 × 10^−6^ after conditioning on His138Tyr; Supplementary Table 7) but did not eliminate the signal, suggesting that other variants in *FSCN2* may increase the risk for hearing loss. Mice homozygous for loss-of-function mutations in *Fscn2* present progressive hearing loss starting at 3 weeks and near deafness by 24 weeks due to degeneration of the outer hair cells in the cochlea^41^. In *SYNJ2*, we identified an association with a missense (Thr656Met)^18^ variant that lies in the catalytic domain of this inositol polyphosphate 5-phosphatase^42^. Mice harboring homozygous mutations in *Synj2* that are predicted to reduce protein levels or the 5-phosphatase catalytic activity show progressive high-frequency hearing loss and a degeneration of hair cells that is most profound in the outer hair cells^43,44^.

**Figure 4.**
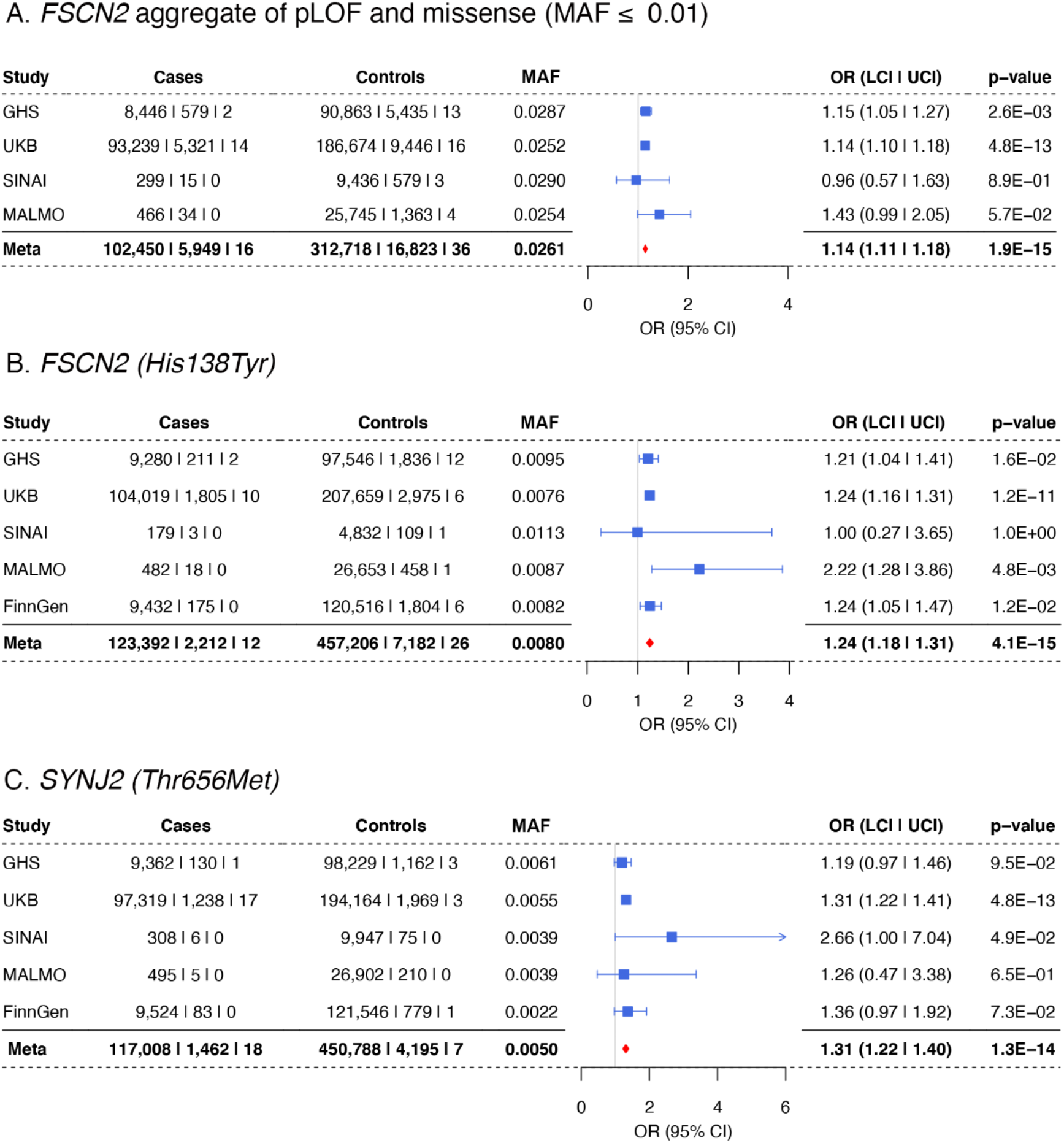
Association of human hearing loss with genes previously implicated in hearing loss in mice. **A**. pLOF and missense (MAF ≤ 0.01) burden association in *FSCN2*. **B**. The His138Tyr variant is the major contributor to the burden. **C**. *SYNJ2* (Thr656Met) association with increased risk for hearing loss.

### GWAS/ExWAS supports a highly polygenic architecture of adult hearing loss

Given that this is the largest sequencing study to date of adult hearing loss, and given the presence of Mendelian hearing loss genes among our common and rare-variant associations, we sought to explore the distribution of effect sizes across allele frequencies (Figure 5). Hearing loss-associated variants span the frequency spectrum and, while we observe a few rare variants of large effect (e.g. *COCH* Cys542Phe and *MYO6* His246Arg), we do not observe any common variants of large effect. We further estimated phenotypic variance explained by the genetic data, or heritability *h*^*2*^_*Tot*_, using LD score regression (LDSC) partitioning into functional categories and stratifying by minor allele frequency^45,46^. We estimated a total heritability (h^2^_Tot_) of 0.089, with contributions from common variation (h^2^_CV_, MAF > 0.05) and low-frequency variation (h^2^_LFV_, 0.001 < MAF ≤ 0.05) of 0.074 and 0.015, respectively (Supplementary Figure 3, Supplementary Table 11). These results indicate that, while the bulk of SNP heritability is derived from common variation, low-frequency variation contributes 16.8% of total SNP heritability.

**Figure 5:**
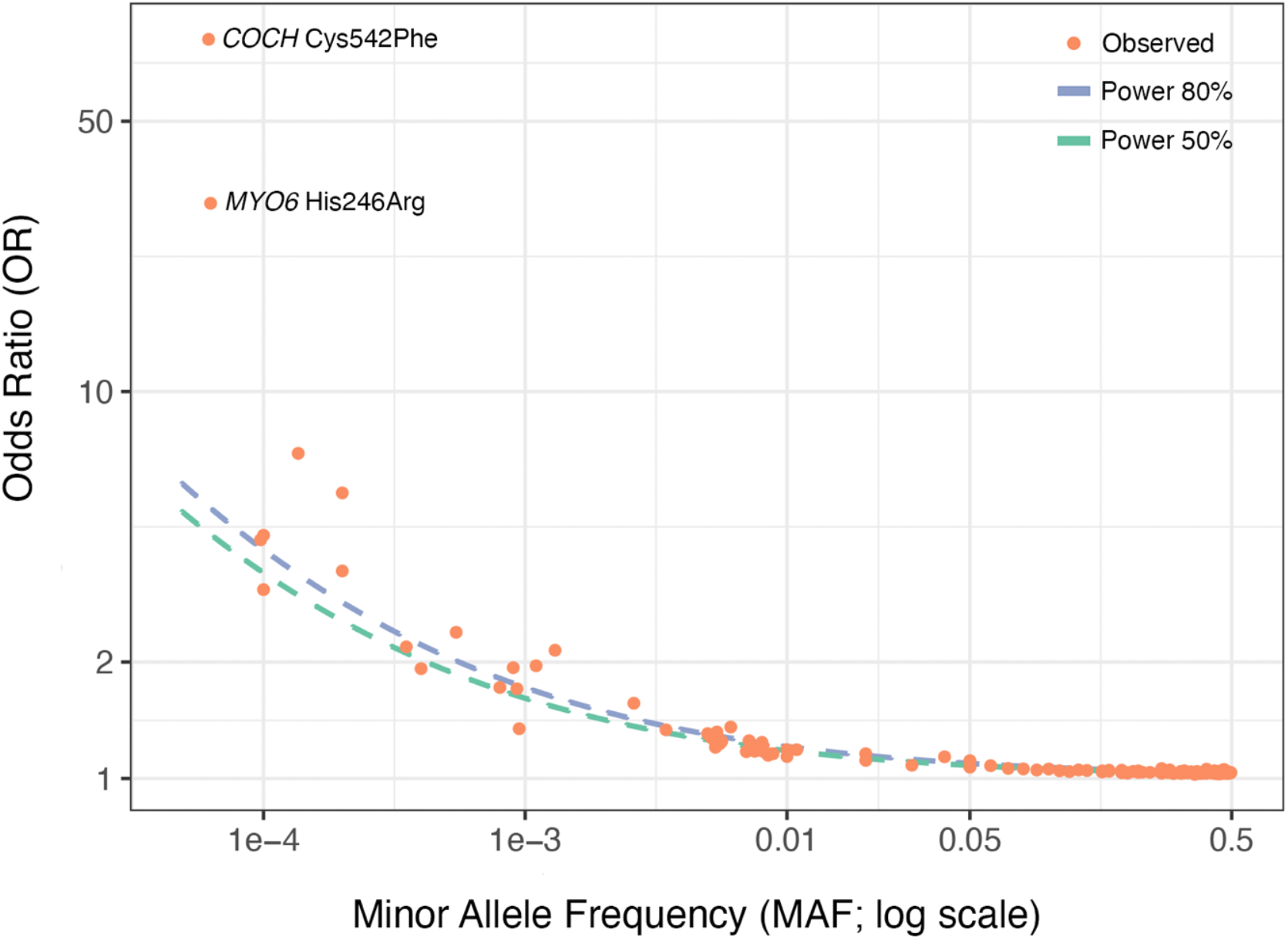
Effect size and allele frequency for variants associated with adult hearing loss. Plotted are odds ratio estimates (on log scale) and minor allele frequencies of genome-wide significant variants and gene burdens (Supplementary Tables 2, 6 & 7). 50% and 80% power curves for the present study are plotted as dotted lines. Notably, almost all points (particularly low-frequency and common variants, MAF > 0.0005) lie very close to the dotted power curves.

## DISCUSSION

We performed combined GWAS and ExWAS using exome sequencing data and identified 53 independent associations, of which 15 were novel. Novel associations included a missense lead variant in *TMPRSS3*, a known cause of Mendelian hearing loss, adding to the tally of Mendelian deafness genes (*EYA4, CDH23, TRIOBP*) showing common coding-variant associations with hearing loss in adult humans. We also identified coding variant/gene burden associations with 15 genes through exome sequencing. We estimated that low-frequency variation contributes a non-negligible portion (16.8%) of SNP heritability for adult hearing loss, consistent with a highly polygenic genetic architecture with rare, low-frequency and common genetic variation for adult hearing loss, and where variants of large effect would be subject to purifying selection.

Notably, the majority of genes implicated by rare-variant and burden associations are already known to cause Mendelian forms of hearing loss; however, the odds ratios for these associations are wide-ranging. At the lower end of the effect spectrum are single-variant associations in *GJB2* and *SLC26A5*, with ORs close to typical for low-frequency GWAS findings (1.2-1.3), followed by *COL11A2* (2.2-7) and, at the highest end of the distribution, *COCH* and *MYO6* (31-81). *COCH* Cys542Phe and *MYO6* His246Arg are known pathogenic variants, and consistent with this, we see almost all carriers presenting hearing loss. The two control carriers for each variant (Supplementary Table 6) could be explained by imperfect ascertainment of the phenotype or, as these mutations cause late-onset and progressive hearing loss, could also reflect a difference in expressivity. While some dilution of the effect sizes may be expected when working with self-reported as opposed to objective measures of hearing loss, the broad range in ORs that we observe suggests that for several of these Mendelian hearing loss genes we are identifying carriers with incompletely penetrant, risk-increasing variants. These results are consistent with the hypothesis that there is a continuum between Mendelian and common forms of hearing loss with the same genes harboring mutations ‘causal’ for the former, and risk-increasing for the latter. Furthermore, compared to epidemiological risk factors for common hearing loss, including noise exposure (OR∼1.5-3^47^), odds ratios of low-frequency and rare genetic factors may be large enough to provide mechanistic insights and to have implications for precision medicine.

Ultimately, to understand the biology of hearing loss and develop testable hypotheses from our association results, we need to identify the genes driving the observed associations at each locus. For common variants, we note the high resolution of FINEMAP to prioritize causal variants in several loci, including pinpointing single variants in four genes, three of which are related to hearing loss (*CDH23*), hearing function (*CTBP2*), or have significant rare variant associations with hearing loss (*KLHDC7B*). While FINEMAP helps prioritize variants, the gene that is impacted by those variants is not necessarily obvious, so we looked for colocalization with GTEx eQTL data and identified 19 genes whose expression may influence hearing loss risk (Supplementary Table 4). Analysis of single-cell expression data from mouse ears showed that 13 of the 19 genes are expressed in the ear. Of note, among these are *LMO7* and *SPTBN1* (BetaII-spectrin). Both genes code for components of the cuticular plate in the hair cells of the ear and knockout mice for either gene develop hearing loss^48,49^. *LMO7* colocalizes with eQTLs for the gene in GTEx skeletal muscle and *SPTBN1* in thyroid tissue, such that decreased expression of these genes is correlated with increased risk for hearing loss in humans.

One of the most compelling ways in which human genetics can establish a role for a gene in disease is when a diverse set of rare nonsynonymous or loss-of-function variants in the same gene show consistent association with disease. Coding variant and gene burden associations that we identified included associations with known pathogenic (e.g. in *MYO6* and *COCH*) and novel variants (*COL11A2* Phe80Ser), in genes that can cause autosomal dominant Mendelian hearing loss. We also identified variants and genes that cause recessive hearing loss (*GJB2* Gly12fs and *SLC26A5* pLOF+strict deleterious missense mutations), associated with increased risk (OR∼1.2-1.3) for hearing loss in heterozygous carriers. Missense mutations in *GJB2* can cause dominant, syndromic hearing loss^50^ but loss-of-function mutations, including Gly12fs, cause recessive non-syndromic hearing loss^51–53^. Consistent with our findings, studies have detected hearing deficiency at high frequencies and a possibly more prominent effect in female adult heterozygous carriers of Gly12fs^53–55^. Mutations in *SLC26A5* cause recessive hearing loss in humans^56^ and mice homozygous for *Slc26a5* null mutations develop hearing loss as early as 5-7 weeks of age^57^. *Slc26a5* heterozygous null mice have a hearing deficiency that is intermediate between the controls and knockouts. While we cannot rule out the possibility of compound heterozygosity with variation in regulatory/promoter regions in heterozygous carriers, our results offer the possibility that heterozygous carriers of loss-of-function variants in *GJB2* and *SLC26A5* may also have increased risk for adult-onset hearing loss.

Three of the 15 coding and burden associations were in genes that have not previously been implicated in Mendelian hearing loss in humans: *KLHDC7B, FSCN2*, and *SYNJ2*. In *KLHDC7B*, we confirmed a previously reported association of the common Val504Met and a rare frameshift variant with increased risk for hearing loss^18,58^, and identified associations of increased risk with a burden of additional pLOF variants. While the biological effect of the missense is difficult to interpret, the association of putative LOFs with increased risk for hearing loss suggests that loss of KLHDC7B function is deleterious for hearing function. Based on the smaller effect of Val504Met (OR = 1.14) compared to the pLOF burden (OR=2.14), we would hypothesize that this more common missense (MAF = 4%) is a hypomorph. *KLHDC7B* (Kelch-like domain containing 7B) is a relatively understudied gene; it has been characterized as a 594-amino-acid protein containing a Kelch domain that is hypermethylated and upregulated in breast cancer cell lines and may influence cell proliferation in MCF-7 cells lines^59,60^. In the mouse ear, RNAseq expression profiling^61^ and our real-time PCR data (Supplementary Figure 4) show *Klhdc7B* expression in the cochlea with enrichment in the outer hair cells^62^. Consistent with the hypothesis from our genetic findings that loss of KLHDC7B function increases risk for hearing loss, initial characterization of *Klhdc7B* null mice by IMPC showed that homozygous carriers have hearing loss^26^. Mouse models *Fscn2* and *Synj2* also develop hearing loss^41,43,44^. Based on our association results, it would also be interesting to test heterozygous null *Klhdc7b, Fscn2*, and *Synj2* animals for increased susceptibility to hearing loss with age or environmental insults such as noise-exposure.

We recognize that our study has several limitations. Our phenotype includes (in UKB) self-reported hearing loss among adults, which is likely to be a heterogeneous mix of early-onset, late-onset, age-related, as well as hearing loss due to environmental insults. In general, greater phenotype precision, including environmental exposure measures, should help future genetic analyses of adult hearing loss. We note that our colocalization analyses utilized GTEx eQTL data across tissues not including ear expression data. Given the high degree of sharing of genetic regulation of expression across tissues^63,64^, the results are likely to point to the causal genes in many instances. eQTL data for ear cell types should help with the interpretation of genetic analyses of adult hearing loss.

In summary, this work contributes to connecting the two ends of the genetic architecture of hearing loss by detecting a common signal in genes known to cause hearing loss in Mendelian fashion and by detecting an additive signal (i.e. increased risk in heterozygous carriers) in genes known to cause autosomal recessive hearing loss. This latter finding also connects young- and adult-onset hearing loss in a single phenotypic spectrum with complex genetic underpinnings, including contributions from rare and common variation.

## METHODS

### Participating cohorts and phenotype data

We performed meta-analysis for hearing loss on a total of 125,749 cases and 469,495 controls of European ancestry from the following cohorts: United Kingdom Biobank (UKB)^65^, the MyCode Community Health Initiative cohort from Geisinger Health System (GHS)^66^, the Mount Sinai Biome cohort (SINAI) (see URLs), the Malmö Diet and Cancer study (MALMO)^67^, and FinnGen R3 (URLs). Hearing loss case-control status in GHS, MALMO and SINAI was defined using ICD-10 code diagnoses from the EHR. Cases were individuals with ICD10 H90.3-H90.8, H91.1or H91.9 diagnoses. Controls were individuals who did not meet the case criteria and did not have a diagnosis for ICD-10 Q16 (congenital malformations of ear causing hearing impairment) or ICD-10 H93.1 (tinnitus). In UKB the phenotype was defined using ICD-10 codes as above or by self-reported hearing loss or complete deafness on the touchscreen questionnaire (Field IDs: 2247 and 2257). Controls in UKB were individuals who did not have an ICD-10 diagnosis for hearing loss or tinnitus and did not self-report hearing loss, deafness or tinnitus (Field IDs: 4803 and non-cancer illness code 1597). The FinnGen analysis used was finngen_r3_H8_CONSENHEARINGLOSS, conductive or sensorineural hearing loss defined by ICD10 codes H90[.0-8], versus controls excluding other ear disorders (H91-H95). Further details are given in the Supplementary Note.

### Genetic data and association analyses

High-coverage whole-exome sequencing was performed at the Regeneron Genetics Center as previously described^68,69^. For SINAI and MALMO, DNA from participants was genotyped on the Global Screening Array (GSA), and for GHS genotyping was done on either the Illumina OmniExpress Exome (OMNI) or GSA array; the datasets (stratified by array for GHS) were imputed to the TOPMed (GHS) or HRC (MALMO and SINAI) reference panels using the University of Michigan Imputation Server or the TOPMed Imputation Server (URLs). Additional details are given in the Supplementary Note. We tested for association with hearing loss genetic variants or their gene burdens using REGENIE v1.0.43^70^. Analyses were adjusted for age, age2, sex, an age-by-sex interaction term, experimental batch-related covariates, and genetic principal components. Cohort-specific statistical analysis details are provided in the Supplementary Note. Results across cohorts were pooled using inverse-variance weighted meta-analysis.

### Fine mapping and follow-on genetic analyses

We defined genome-wide significant loci in our analysis by linkage disequilibrium (r^2^ > 0.1) with lead variants. We defined previously associated loci by their index variants reported in previous hearing loss GWAS^13–17,58,71–75^, and excluded 1 Mb regions surrounding them in the identification of novel loci in our analysis. LD score (LDSC) regression^76^ was used to assess inflation (LDSC intercept) accounting for polygenic signal. Power calculations determined genotype relative risks (GRRs) providing 80 and 50 percent power given specified risk allele frequencies (RAFs, from 10^−6^ to 1) and the numbers of cases and controls in our meta-analysis. Fine mapping analyses included forward stepwise conditional analyses carried out in every locus with GCTA-COJO using a UK Biobank subsample LD reference panel, with independent associations determined using a joint P-value threshold of 1 × 10^−5^ and r^2^ cutoff of 0.9, and FINEMAP^33^ Bayesian causal variant inference, using LD from available individual level data.

For genes whose cis regions overlapped genome-wide significant hearing loss loci, coloc2^75,76^ was used to assess evidence for colocalization between our hearing loss GWAS and GTEx (release v8) cis-eQTL data derived from 48 tissues (URLs), using GWAS and eQTL summary statistics for all common (MAF ≥ 0.01) variants within each gene’s cis-region. Genes with posterior probability of colocalization H4 ≥ 0.5 were determined as having evidence for colocalization, and were visually inspected in eQTL+GWAS regional association plots (Supplementary Figure 5).

Conditional analyses were performed for each cohort using REGENIE’s Firth-corrected logistic regression and the resulting summary statistics were meta-analyzed as described above. Rare variant association analyses conditional on the common variant signal were carried out for four loci with both common (MAF ≥ 0.01) and single rare variant (MAF < 0.01) genome-wide significant signals, by including as covariates the dosages of variants identified in fine mapping analyses. Burden analyses conditional on rare variants were carried out for five genes with significant single rare variant and burden associations.

Assessment of heterozygous effects used association analyses excluding homozygotes as well as individuals carrying pairs of exome sequenced variants (MAF < 2% and MAC > 1) that were called as compound heterozygous mutations (CHMs) or potential CMHs (i.e. unknown phase). CHMs were called by SHAPEIT4 phasing of merged genotype and exome data with scaffolds based on inferred close relatives^77,78^.

Heritability derived from variants partitioned into seven functional categories (coding-synonymous, coding-nonsynonymous, 5-prime-UTR, 3-prime-UTR, splice site, intronic, intergenic) with each category further stratified into a low frequency (0.001 < MAF ≤ 0.05) and common (MAF > 0.05) minor allele frequency bin as in stratified LD score regression was estimated using LD score regression (LDSC) of hearing loss association statistics on LD scores. LD scores were generated from a reference panel of N = 10,000 random UK Biobank European-ancestry samples’ merged imputed and exome data.

### Single-Cell RNA Sequencing and Analysis

All protocols were approved by the Institutional Animal Care and Use Committee in accordance with the Regeneron’s Institutional Animal Care and Use Committee (IACUC). Cochlea and utricles from C57BL/6 mice at post-natal day 7 were micro-dissected and dissociated. Suspensions of 200 cells / uL were subjected to Chromium Single Cell (10xGenomics) library preparation and were sequenced on Illumina NextSeq 500. Cell Ranger Single-Cell Software Suite (10X Genomics, v2.0.0) was used to perform sample de-multiplexing, alignment to MM10 Genome assembly with UCSC gene models, filtering, and UMI counting. PCA, UMAP and clustering analyses used Seurat V3.2 (URLs)^79^. Cluster marker genes as well as canonical cell type-specific genes were used to manually label the cell type for each cluster.

### URLs

International Mouse Phenotyping Consortium: https://www.mousephenotype.org/.

gEAR portal: https://umgear.org/

<u>GTEx:</u> https://www.gtexportal.org/home/

[referred to in METHODS]

Mount Sinai Bio*Me*: https://icahn.mssm.edu/research/ipm/programs/biome-biobank/pioneering

FinnGen: https://www.finngen.fi/

University of Michigan Imputation Server: https://imputationserver.sph.umich.edu/index.html

TOPMed Imputation Server: https://imputation.biodatacatalyst.nhlbi.nih.gov/

MakeScaffold: https://github.com/odelaneau/makeScaffold

SHAPEIT4: https://github.com/odelaneau/shapeit4

GTEx v8 eQTL data: https://www.gtexportal.org/

Seurat V3.2: https://github.com/satijalab/seurat

## Data Availability

All whole-exome sequencing, genotyping chip, and imputed sequence for UKB described in this report are publicly available to registered researchers via the UK Biobank data access protocol. Additional information about registration for access to the data is available at http://www.ukbiobank.ac.uk/register-apply/. Further information about the whole exome sequence is available at http://www.ukbiobank.ac.uk/wp-content/uploads/2019/03/Access_064-UK-Biobank-50k-Exome-Release-FAQ-v3.pdf Detailed information about the chip and imputed sequence is available at: http://www.ukbiobank.ac.uk/wp-content/uploads/2018/03/UKB-Genotyping-and-Imputation-Data-Release-FAQ-v3-2-1.pdf. Geisinger DiscovEHR, Malmo Diet and Cancer study and Mt. Sinai Biome exome sequencing and genotyping data can be made available to qualified, academic, non-commercial researchers upon request via a Data Transfer Agreement with the respective institutions. Summary statistics for FinnGen r3 can be downloaded from https://www.finngen.fi/en/access_results.

## ACKNOWLEDGEMENTS

The authors would like to thank everyone who made this work possible. In particular: the UK Biobank team, their funders, the dedicated professionals from the member institutions who contributed to and supported this work, and most especially the UK Biobank participants, without whom this research would not be possible. The exome sequencing was funded by the UK Biobank Exome Sequencing Consortium (i.e., Bristol Myers Squibb, Regeneron, Biogen, Takeda, Abbvie, Alnylam, AstraZeneca and Pfizer). This research has been conducted using the UK Biobank Resource under application number 2604. We also want to acknowledge the participants and investigators of the FinnGen study. We thank the MyCode Community Health Initiative participants for taking part in the DiscovEHR collaboration.

## Author contributions

All authors contributed to critical review of the manuscript for important intellectual content, and final approval of submission of the manuscript for publication.

Conceptualization: KP, EAS, MD, BZ, AB, GC Data curation: MNC

Analysis: KP, LD, LG, MAF, AHA, JS, AP, AM, SC, EAS

Single-cell sequencing data generation and analysis: SM, YB, JS

Mouse experiments: AK

Project administration: EC, MJ

Datasets: OM

Supervision: MAF, GA, EAS, SB, BZ, GC, AB

Writing – original draft: KP, LD, EAS, GC

