## Supplementary Appendix, Supplementary Figures 1, 3, 4 and Supplementary Note for "Population-scale analysis of common and rare genetic variation associated with hearing loss in adults"

**SUPPLEMENTAL FILE 1.**
**Praveen et al.**

Supplementary Appendix:

#### Regeneron Genetics Center Banner Author List and Contribution Statements

All authors/contributors are listed in alphabetical order.

RGC Management and Leadership Team

Goncalo Abecasis, Aris Baras, Michael Cantor, Giovanni Coppola, Andrew Deubler, Aris Economides, Luca A. Lotta, John D. Overton, Jeffrey G. Reid, Alan Shuldiner, Katia Karalis and Katherine Siminovitch

Contribution: All authors contributed to securing funding, study design and oversight. All authors reviewed the final version of the manuscript.

Sequencing and Lab Operations

Christina Beechert, Caitlin Forsythe, Erin D. Fuller, Zhenhua Gu, Michael Lattari, Alexander Lopez, John D. Overton, Thomas D. Schleicher, Maria Sotiropoulos Padilla, Louis Widom, Sarah E. Wolf, Manasi Pradhan, Kia Manoochehri, Ricardo H. Ulloa.

Contribution: C.B., C.F., A.L., and J.D.O. performed and are responsible for sample genotyping. C.B, C.F., E.D.F., M.L., M.S.P., L.W., S.E.W., A.L., and J.D.O. performed and are responsible for exome sequencing. T.D.S., Z.G., A.L., and J.D.O. conceived and are responsible for laboratory automation. M.S.P., K.M., R.U., and J.D.O are responsible for sample tracking and the library information management system.

Genome Informatics

Xiaodong Bai, Suganthi Balasubramanian, Boris Boutkov, Gisu Eom, Lukas Habegger, Alicia Hawes, Shareef Khalid, Olga Krasheninina, Rouel Lanche, Adam J. Mansfield, Evan K. Maxwell,

Mona Nafde, Sean O'Keeffe, Max Orelus, Razvan Panea, Tommy Polanco, Ayesha Rasool, Jeffrey G. Reid, William Salerno, Jeffrey C. Staples

Contribution: X.B., A.H., O.K., A.M., S.O., R.P., T.P., A.R., W.S. and J.G.R. performed and are responsible for the compute logistics, analysis and infrastructure needed to produce exome and genotype data. G.E., M.O., M.N. and J.G.R. provided compute infrastructure development and operational support. S.B., S.K., and J.G.R. provide variant and gene annotations and their functional interpretation of variants. E.M., J.S., R.L., B.B., A.B., L.H., J.G.R. conceived and are responsible for creating, developing, and deploying analysis platforms and computational methods for analyzing genomic data.

Clinical Informatics:

Nilanjana Banerjee, Michael Cantor, Dadong Li, Deepika Sharma, Ashish Yadav

Contribution: All authors contributed to the clinical informatics of the project

Translational and Analytical Genetics:

Alessandro Di Gioia, Sahar Gelfman

Contribution: All authors contributed to the analysis of the project.

Research Program Management

Esteban Chen, Marcus B. Jones, Jason Mighty, Michelle G. LeBlanc and Lyndon J. Mitnaul

Contribution: All authors contributed to the management and coordination of all research activities, planning and execution. All authors contributed to the review process for the final version of the manuscript.

**GHS DiscovEHR banner authors**

Lance J. Adams, Jackie Blank, Dale Bodian, Derek Boris, Adam Buchanan, David J. Carey, Ryan D. Colonie, F. Daniel Davis, Dustin N. Hartzel, Melissa Kelly, H. Lester Kirchner, Joseph B. Leader, David H. Ledbetter, Ph.D., J. Neil Manus, Christa L. Martin, Raghu P. Metpally, Michelle Meyer, Tooraj Mirshahi, Matthew Oetjens, Thomas Nate Person, Christopher Still, Natasha Strande, Amy Sturm, Jen Wagner, Marc Williams

**Decibel-REGN Hearing Loss Collaboration Banner Authors**

All authors/contributors are listed in alphabetical order.

Collaboration Core Team

Joe Burns[1], Giovanni Coppola[2], Meghan Drummond-Samuelson[3], Aris Economides[2,3], David Frendewey[3], Scott Gallagher[1], John Lee[1], John Keilty[1], Christos Kyratsous[3], Lynn Macdonald[3], Adam T Palermo[1], Kavita Praveen[2], Leah Sabin[3], Jonathon Whitton[1], Brian Zambrowicz[3]

Contribution: Authors helped frame research questions and contributed to the discussion and review of data and results. Review and feedback on manuscript.

Program Management & Alliance Management

Sarah Deng[3], Geoff Horwitz[1], Alejandra K. King[3], Jung H Sung[3]

Contribution: Contributed to the management and coordination of discussions.

Affiliations:

1. Decibel Therapeutics, Boston, MA USA
2. Regeneron Genetics Center, Tarrytown, NY USA
3. Regeneron Pharmaceuticals, Tarrytown, NY USA

### SUPPLEMENTARY FIGURES

#### Supplementary Figure 1.

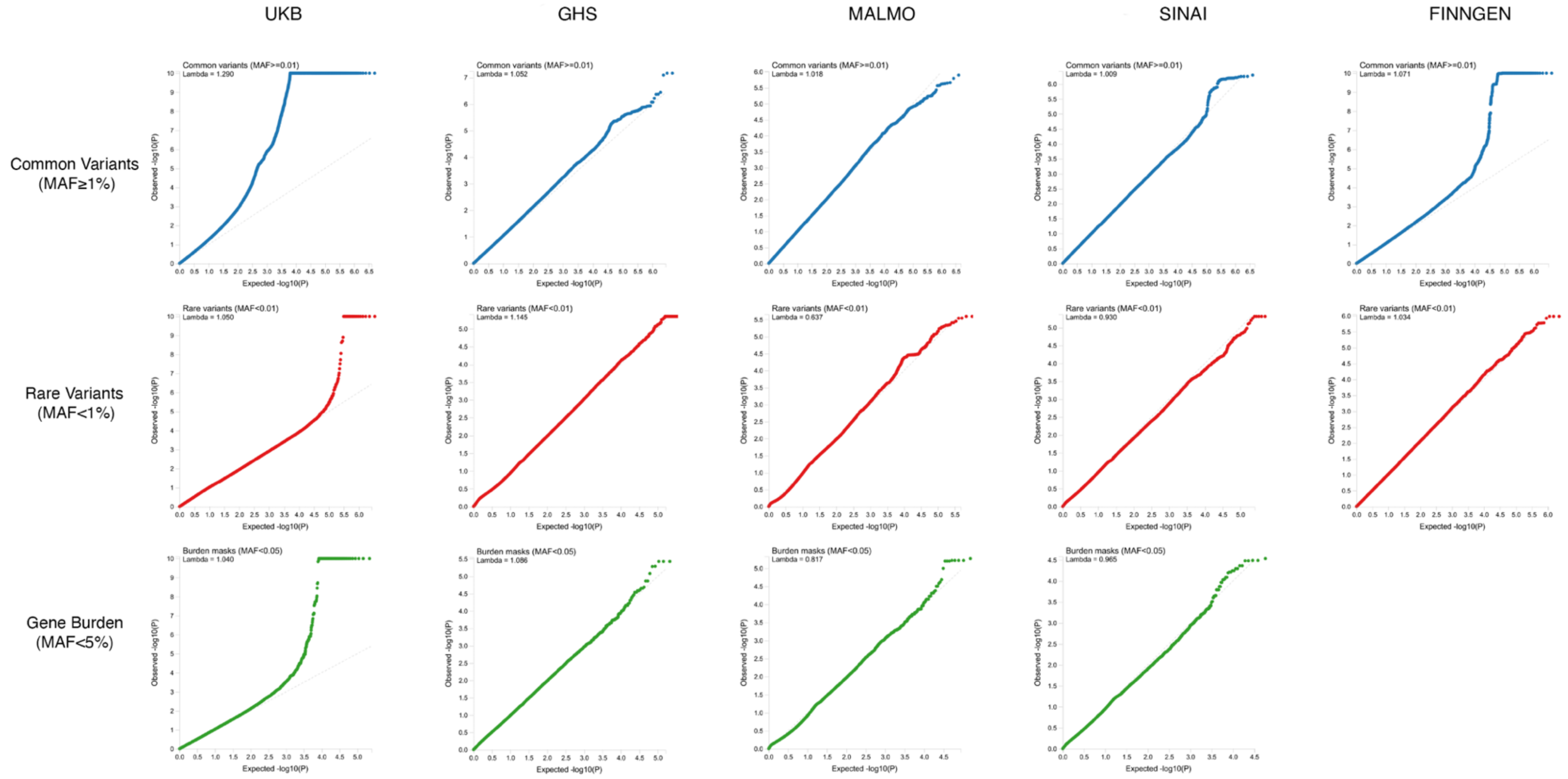

**Supplementary Figure 1: Q-Q plots for common and rare single variant and gene burden associations in UKB, GHS, MALMO, SINAI and FinnGen.**

#### Supplementary Figure 2.

**[See Supplemental File 2 for Supplementary Figure 2: Regional plots for novel common (MAF ≥ 1%) loci identified in hearing loss meta-analysis and forest plots corresponding to the index variant at each novel locus (panels A-R, 9 pages)]**

Supplementary Figure 3.

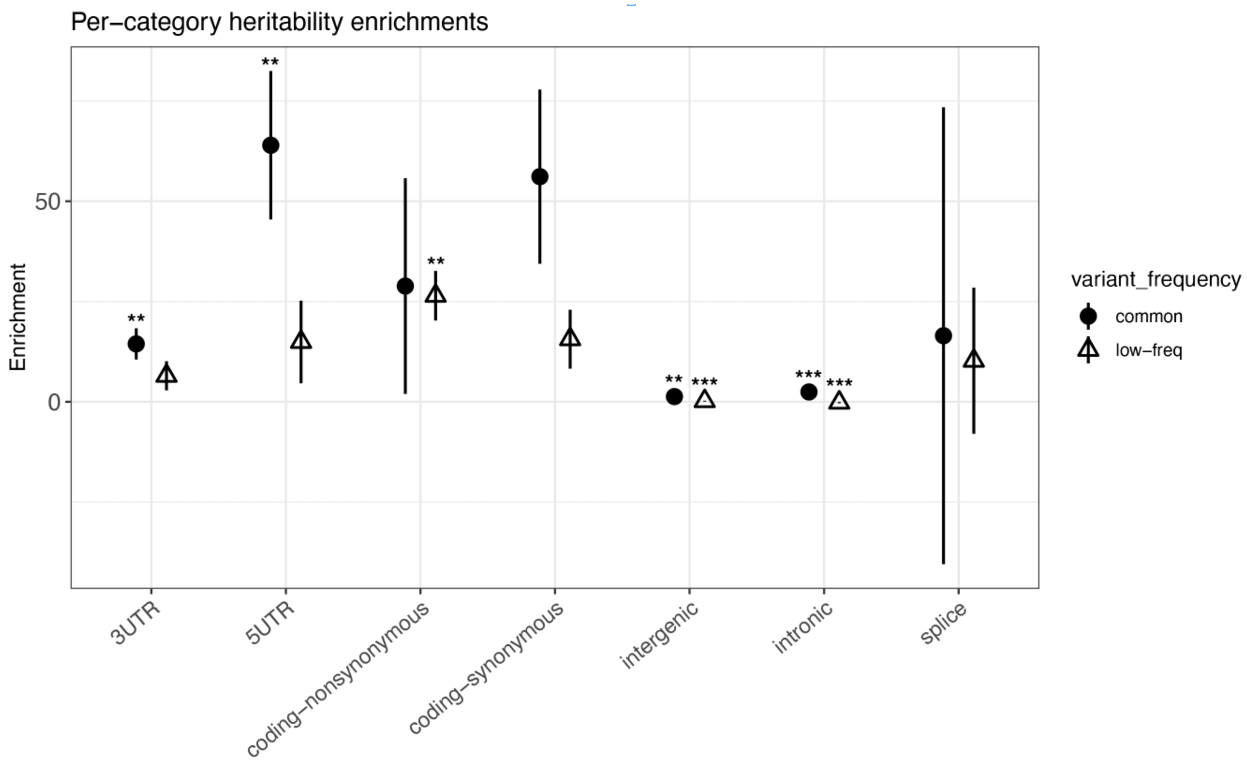

**Supplementary Figure 3: Heritability enrichments from stratified LD score regression analysis.** Total SNP heritability for seven functional categories, each further stratified by MAF into a common variant (CV, MAF ≥ 0.05) and low-frequency variant (LFV, 0.001 ≤ MAF<0.05) bin, was estimated and enrichments for these categories was calculated (proportion heritability / proportion variants). Plotted are the resulting enrichments (common variant bins shown as solid circles and low-frequency variant bins as open triangles) with standard errors. Significant enrichments and depletions are denoted by asterisks (Bonferroni-corrected p<0.05 = *; p<0.01 = **; p<0.001 = ***)

Supplementary Figure 4.

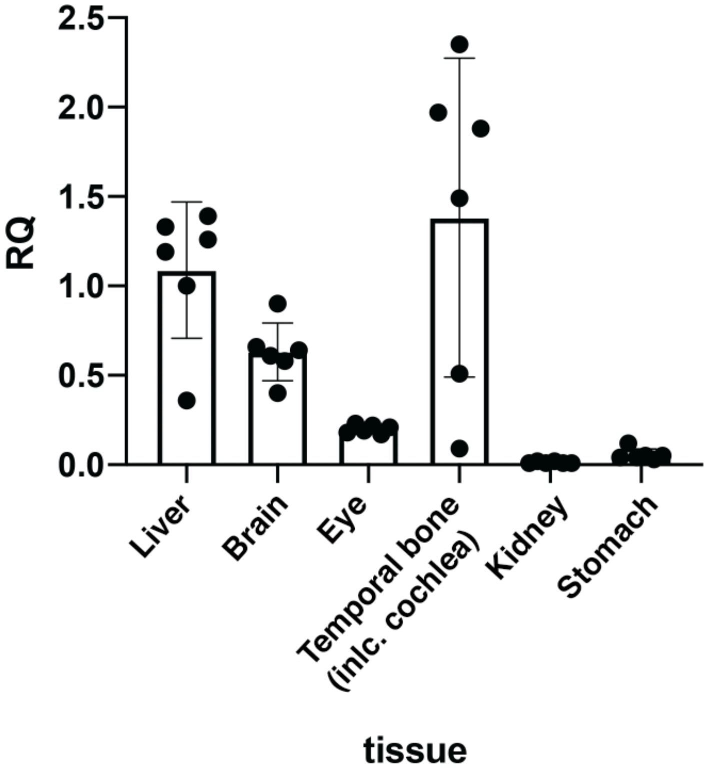

**Supplementary Figure 4: KLHDC7b is expressed in tissue within the temporal bone.** RNA was extracted from different organs, and qPCR was performed using a primer/probe combination for KLHDC7b, and one for Drosha as a housekeeping control. Data for each organ was normalized to the housekeeping control and then normalized to liver expression. Expression in cochlea and brain were not significantly different from expression in liver, which was relatively high. RQ = relative quantification compared to liver (see methods).

### SUPPLEMENTARY NOTE

#### Supplementary Methods

**Phenotype definition:**

Hearing loss in GHS, MALMO and SINAI was defined using ICD-10 codes: cases were individuals who had (1) a problem-list entry of the ICD-10 diagnosis code (H903-H908, H911, H919), (2) an inpatient hospitalization-discharge ICD-10 diagnosis code, or (3) an encounter ICD-10 diagnosis code entered for 2 separate outpatient visits on separate calendar days. Controls were individuals without any of the criteria for case definition. Individuals were excluded if they had the relevant ICD-10 code associated with only one outpatient encounter. We also excluded from controls any individuals who were cases for ICD-10 Q16 (congenital malformations of ear causing hearing impairment) or ICD-10 H931 (tinnitus).

In UKB, hearing loss was defined using ICD-10 codes and self-reports based on two questions: "Do you have any difficulty with your hearing?" (Field: 2247) and "Do you find it difficult to follow a conversation if there is background noise (such as TV, radio, children playing)?" (Field: 2257). Self-reported cases were individuals who (1) answered 'yes' to both questions or (2) were completely deaf or (3) were a case for any of the following ICD-10 codes: H903-H908, H911, H919. Phenotype definition of ICD10-based cases required one or more of the following: a) ≥ 1 diagnosis in inpatient Health Episode Statistics (HES) records, b) a cause-of-death diagnosis in death registry, c) ≥ 2 diagnoses in outpatient data (READ codes mapped to ICD10). ICD-based controls were individuals who did not meet the case critera, and were not cases for ICD-10 Q16 and ICD-10 H931. To obtain the overall cases in the analysis, self-reported and ICD-based cases were combined. Controls for the overall analysis were defined as individuals who (1) answered 'No' to both self-report hearing loss questions and, (2) did not report that they were deaf and (3) did not meet the criteria for ICD-based case definition and (4) did not have tinnitus based on ICD-10 (H931) or self-reported tinnitus (Field ID: 4803, 4814 and self-reported from verbal interview).

#### Genotyping

For SINAI and MALMO, DNA from participants was genotyped on the Global Screening Array (GSA), and for GHS genotyping was done on either the Illumina OmniExpress Exome (OMNI) or GSA. These cohorts were imputed to the TOPMed (GHS) or the HRC (MALMO, SINAI) reference panels (stratified by array for GHS) using the University of Michigan Imputation Server or the TOPMed Imputation Server (URLs). Prior to imputation, we retained variants that had a MAF ≥ 0.1%, missingness < 1% and HWE $P > 10^{-15}$. Following imputation for GHS, data from the OMNI and GSA datasets were merged for subsequent association analyses, which included an OMNI/GSA batch covariate in addition to other covariates described below. UKB DNA samples were genotyped as described previously[1] using the Applied Biosystems UK BiLEVE Axiom Array (N=49,950) or the closely related Applied Biosystems UK Biobank Axiom Array (N=438,427). Genotype data for variants not included in the arrays were imputed using three reference panels (Haplotype Reference Consortium, UK10K and 1000 Genomes Project phase 3) as described previously[1]. FinnGen data were derived from a custom Axiom array and imputed into the FinnGen SISu v3 reference panel (URLs).

#### Exome sequencing

High coverage whole exome sequencing was performed at the Regeneron Genetics Center as previously described[2,3]. NimbleGen probes (VCRome) or a modified version of the xGen design from Integrated DNA Technologies (IDT) were used for target sequence capture, and sequencing was performed using 75 bp paired-end reads on Illumina v4 HiSeq 2500 or NovaSeq instruments to a coverage depth greater than 20x at at least 85% of targeted bases in 96% of VCRome samples, and at least 90% of targeted bases in 99% of IDT samples. Sequence read alignment and variant calling was based on the GRCh38 Human Genome reference sequence. Ensembl v85 gene definitions were used to determine variants' functional impacts. Predicted LOF genetic variants included (a) insertions or deletions resulting in a frameshift, (b) insertions, deletions or single nucleotide variants resulting in the introduction of a premature stop codon or in the loss of the transcription start site or stop site, and (c) variants in donor or acceptor splice sites. Missense variants were classified for likely functional impact according to the number of *in silico* prediction algorithms that predicted deleteriousness using SIFT, Polyphen2_HDIV and Polyphen2_HVAR, LRT and MutationTaster. We aggregated rare variants for gene burden testing as previously described[4]. Briefly, rare variants were collapsed by gene region, such that individuals who are homozygous reference for all variants are considered homozygous reference, heterozygous carriers of any aggregated variant are considered heterozygous, and only minor allele homozygotes for an aggregated variant are considered as minor allele homozygotes. Genotypes were not phased to consider compound heterozygotes in burden testing. For each gene, we considered four categories of aggregates: a strict burden of rare pLOFs and three more permissive burden of rare pLOFs and missense variants. The missense variants in the burden aggregates were defined as 'strict deleterious missense' if predicted deleterious by 5/5 prediction algorithms (SIFT, Polyphen2_HDIV, Polyphen2_HVAR, LRT, MutationTaster), and 'deleterious missense' if predicted deleterious by at least 1/5[4]. For each of these groups, we considered five separate burden masks per gene, based on the frequency of the alternative allele of the variants that were screened in that group: MAF $\leq$ 1%, MAF $\leq$ 0.1%, MAF

≤ 0.01%, MAF ≤ 0.001%, and singletons only. For the purposes of gene burden testing, the singleton mask includes minor allele homozygotes if no other variant carriers are observed in the dataset. We conducted further QC of associated variants *post-hoc*, based on mappability statistics from read alignment.

**Genetic association analyses**

Association analyses in each study were performed using the Firth logistic mixed model regression test implemented in REGENIE[5]. We included in step 1 of REGENIE (i.e. prediction of individual trait values based on the genetic data) directly genotyped variants with a minor allele frequency (MAF) > 1%, < 10% missingness, Hardy-Weinberg equilibrium test $P > 10^{-15}$ and linkage-disequilibrium (LD) pruning (1000 variant windows, 100 variant sliding windows and $r^2 < 0.9$). The association model used in step 2 of REGENIE included as covariates (i) age, age$^2$, sex, age-by-sex and age$^2$-by-sex; (ii) 10 ancestry-informative principal components (PCs) derived from the analysis of a set of LD-pruned (50 variant windows, 5 variant sliding windows and $r^2 < 0.5$) common variants from the array (imputed for the GHS study) data generated separately for each ancestry; (iii) an indicator for exome sequencing batch (GHS: three batches; UKB: six batches); and (iv) 20 PCs derived from the analysis of exome variants with minor allele count ≤ 20 and MAF < 1% also generated separately for each ancestry.

We determined continental ancestries by projecting each sample onto reference principal components calculated from the HapMap3 reference panel. Briefly, we merged our samples with HapMap3 samples and kept only SNPs in common between the two datasets. We further excluded SNPs with MAF < 10%, genotype missingness > 5% or Hardy-Weinberg Equilibrium test $P < 10^{-5}$. We calculated PCs for the HapMap3 samples and projected each of our samples onto those PCs. To assign a continental ancestry group to each non-HapMap3 sample, we trained a kernel density estimator (KDE) using the HapMap3 PCs and used the KDEs to calculate the likelihood of a given sample belonging to each of the five continental ancestry groups. When the likelihood for a given ancestry group was > 0.3, the sample was assigned to that ancestry group. When two ancestry groups had a likelihood > 0.3, we arbitrarily assigned AFR over EUR, Admixed American (AMR) over EUR, AMR over East Asian (EAS), South Asian (SAS) over EUR, and AMR over AFR. Samples were excluded from analysis if no ancestry likelihoods were > 0.3, or if more than three ancestry likelihoods were > 0.3. Results were subsequently meta-analyzed across studies and ancestries using an inverse variance-weighted fixed-effects meta-analysis using an inverse variance-weighted model in METAL[6].

**Finemapping and follow-on genetic analyses**

LD score (LDSC) regression[7] was used to assess inflation (LDSC intercept) in our accounting for polygenic signal. We used LD scores calculated using genotyped or imputed variants INFO>0.3 and MAF>0.5% from 10,000 randomly chosen subjects from UKB, and restricted our analysis to HapMap3 variants.

We defined previously associated loci by their index variants reported in previous hearing loss GWAS, and excluded 1 Mb regions surrounding them in the identification of novel loci in our analysis. We defined genome-wide significant loci in our analysis by linkage disequilibrium ($r^2 > 0.1$) with lead variants.

Forward stepwise conditional analyses were carried out in every locus with GCTA-COJO using summary statistics and a UK Biobank subsample LD reference panel. Independent associations were determined using a joint P-value threshold of $1 \times 10^{-5}$ and $r^2$ cutoff of 0.9.

Bayesian causal variant inference was conducted in available individual level data using FINEMAP[8].

Rare variant association analyses conditional on the common variant signal were carried out for four loci with both common (MAF ≥ 0.01) and single rare variant (MAF < 0.01) genome-wide significant signals. For these loci, the dosages for variants representing the common variant signal were included as covariates in REGENIE logistic regression. The specific variants that best captured the common variant signal were ascertained through fine mapping analyses (FINEMAP 80% credible sets when available, or GCTA-COJO-identified independent [$r^2 < 0.9$] significant [joint P-value$<1 \times 10^{-4}$] variants). Burden analyses conditional on rare variants were carried out for five genes with significant single rare variants in addition to their burden associations. For these genes, conditional burden association statistics were obtained through inclusion of dosages for the top (most significant) single rare variant in each gene. Conditional analyses were performed for each cohort using REGENIE's Firth-corrected logistic regression and the resulting summary statistics were meta-analyzed as described above.

Assessment of heterozygous effects used association analyses excluding homozygotes as well as individuals potentially carrying compound heterozygous mutations (CHMs) called as follows. Available unphased genotype array data and genetically inferred pedigree structures determined by PRIMUS[9] were used to create a phased genetic scaffold with the program MakeScaffold. The scaffold and the unphased exome data where then provided to SHAPEIT4[9,10] to generate phased exome variant calls. We then identified pairs of exome sequenced variants (MAF < 2% and MAC > 1) within the same person and gene as potential CHMs (pCHM), and determined them to be in cis, trans or unknown based on the phased exome data. Trans or unknown-phase pCHMs were excluded.

Power curves were generated by specifying risk allele frequency (RAF), ranging from $1x10^{-6}$ to 1, and the numbers of cases and controls in our meta-analysis, and then determining which genotype relative risk (GRR) values provide 80 and 50 percent power given the risk allele frequency. *z*-score non-centrality parameters (NCP, i.e. expected values for Wald association test statistics) for a case-control study was obtained following Zaitlen et. al.[11], power was obtained using the `pnorm(u=NCP)` and `qnorm()` R functions, and GRRs for 50 and 80% power curves given RAFs were estimated numerically.

Heritability derived from variants in specific functional categories and minor allele frequency bins (in an approach similar to stratified LD score regression) was estimated using partitioned LD score regression (LDSC) of hearing loss association statistics on LD scores. In order to capture LD from both low-frequency and common variants, a reference panel (N = 10,000 samples) generated from the merging of UK Biobank (European-ancestry) imputed and exome data was used. Reference panel variants were annotated using an internal pipeline and LD scores with respect to seven functional categories (coding-synonymous, coding-nonsynonymous, 5-prime-UTR, 3-prime-UTR, splice site, intronic, and intergenic), each split into a common (MAF > 0.05) and low-frequency (0.001 < MAF ≤ 0.05) variant bin, were calculated. Variants used to calculate LD scores were filtered for false positives identified through support vector machine learning of QC metrics, and both reference panel variants and summary statistics were restricted to those with MAF > 0.001. Since our categories have a very small degree of overlap, with approximately 0.01 percent of variants falling into more than one category, reported per-category enrichment results were taken from the .results file (supplementary table 11) provided by LDSC when using the –overlap-annot flag. As LDSC does not provide per-category h2 estimates when using this flag, however, we used per-category heritability estimates taken from the .log file from non-overlap-annot runs to calculate $h^2_{CV}$ and $h^2_{LFV}$ each as the sum of the relevant common and low-frequency variant category heritabilities.

For genes overlapping genome-wide significant hearing loss loci, coloc2[12] was used to assess evidence for co-localization between our hearing loss GWAS and GTEx (release v8) eQTL derived from 48 tissues (URLs). GWAS and eQTL summary statistics for all common (MAF > 0.01) variants within each gene's cis-region were used as input to coloc2, which then estimates posterior probabilities for five hypotheses ($H_0$, no association; $H_1$, GWAS association only; $H_2$, eQTL association only; $H_3$, both but not co-localized; $H_4$, both and co-localized) given the association statistics and prior probabilities estimated from the observed data. Genes with posterior probability of $H_4 \geq 0.5$ were determined as having evidence for co-localization.

#### Single-Cell RNA Sequencing and Analysis

Cochlea and utricles from C57BL/6 mice at post-natal day 7 were micro-dissected and dissociated via incubation in 0.05% trypsin at 37°C for 20 - 30 min. Four volumes of 5% FBS in DMEM/F12 with a final concentration of DNAse (LK003170) greater than or equal to 100 Kunitz per mL was added to inactivate any remaining trypsin. The tissues were then triturated 20 times and passed through a 40-µm strainer to eliminate residual aggregates / clumps of cells. Cells were counted then resuspended in 0.04% BSA in PBS at a concentration of 200 cells / uL.

Single cells suspended in PBS with 0.04% BSA were loaded on a Chromium Single Cell Instrument (10xGenomics). RNAseq libraries were prepared using the Chromium Single Cell 3′Library, Gel Beads & Multiplex Kit (10x Genomics). Paired-end sequencing was performed on an Illumina NextSeq 500 (Read 1 26-bp for unique molecular identifier, UMI, and cell barcode, 8-bp i7 sample index, 0-bp i5, and Read 2 55-bp transcript read). Cell Ranger Single-Cell Software

Suite (10X Genomics, v2.0.0) was used to perform sample de-multiplexing, alignment, filtering, and UMI counting. The Mouse MM10 Genome assembly and UCSC gene models were used for alignment. CellRanger output was processed using Seurat V3.2 (URLs)[13]. Cells with number of genes detected of less than 500 or over 15000, or UMI ratio of mitochondria encoded genes vs. all genes over 0.15 were removed. Data normalization and scaling for each cell were achieved by using Seurat global-scaling "LogNormalize" method. To avoid potential sample-to-sample variation caused by technical variation at various experiment steps, we employed Seurat's data integration pipeline to merge all cochlea and utricle samples into a single Seurat object for downstream analysis. Statistically significant principal components identified by Seurat's "RunPCA" function were used to define the dimensions for the UMAP nonlinear dimensionality reduction analysis, which then visualized the cells on a 2D UMAP plot. Unsupervised clustering, via Seurat's "FindClusters" method (resolution = 0.6), identified groups of molecularly distinct cells on the plot. Clusters in the UMAP plot were annotated based on cluster-specific genes identified via Seurat's "FindAllMarkers" (min.pct = 0.25, thresh.use = 0.25) function. The expression of cluster marker genes as well as canonical cell type-specific genes were used to label the cell type for each cluster.

**Mouse cochlea dissection for KLHDC7B quantitation:**

There were six biological replicates and three technical replicates of each sample. Cochlea were dissected from B6.CAST-Cdh23Ahl$^{+/WT}$ mice ears. Both cochleae were pooled per mouse from five female mice and one male mouse aged 11-28 weeks. Animals were sacrificed by carbon dioxide inhalation followed by immediate removal of the organs in question, which were stored in RNA later. Cochleae were pierced at the apex and oval window, then gently flushed with ~100 $\mu$l of RNA later, frozen on dry ice and stored at -80 degrees Celsius, while other organs were placed in RNA later and frozen at -20 degrees Celsius until RNA extraction. The vestibular system was included with the cochlea.

*RNA extraction and analysis:*

Tissue/Cells were homogenized in TRIzol, and chloroform was used for phase separation. The aqueous phase, containing total RNA, was purified using MagMAX™-96 for Microarrays Total RNA Isolation Kit (Ambion by Life Technologies) according to manufacturer's specifications. Genomic DNA was removed using RNase-Free DNase Set (Qiagen).

mRNA was reverse-transcribed into cDNA using SuperScript® VILO™ Master Mix (Invitrogen by Life Technologies). cDNA was amplified with the SensiFAST Probe Lo-ROX (Meridian) using the 12K Flex System (Applied Biosystems). An endogenous control gene was used to normalize any cDNA input differences. The KLHDC7b primer/probe combination was forward: GGTGGCCCTGGATGGAATG, reverse: TCTGTGCGTGGGTCATAGC, probe: TTTATGCCATTGGTGGCGAGTGC. The Drosha housekeeping control spanned exons 34-35 and was acquired from Thermofisher (Mm01310009_m1, catalogue number 4331182). Data is reported as the comparative CT method using ΔΔCT. The ΔCt = Klhdc7b – Drosha (housekeeping transcript), ΔΔCt = ΔCt – ΔCt reference sample; Relative quantification (RQ) = 2^ - ΔΔCt.
