## Supplementary Figure 2 for "Population-scale analysis of common and rare genetic variation associated with hearing loss in adults"

**SUPPLEMENTAL FILE 2.**
**Praveen et al.**

**Supplementary Figure 2: Regional plots for novel common (MAF ≥ 1%) loci identified in hearing loss meta-analysis and forest plots corresponding to the index variant at each novel locus. (Panels A through R, pages 1-9.)**

A

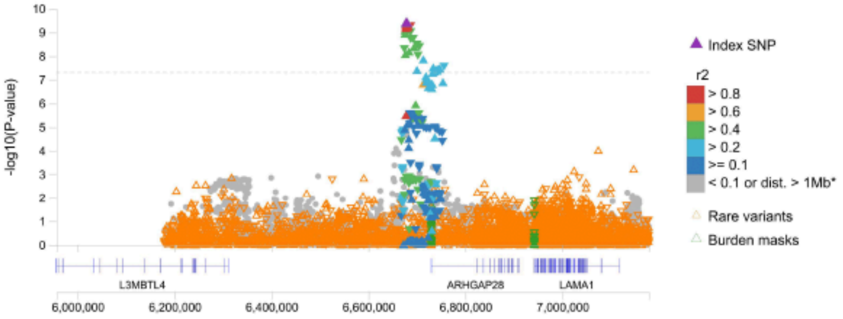


**Association with 18:6678716:T:A**

| STUDY | N CASES RR\|RA\|AA | N CONTROLS RR\|RA\|AA | OR [95% CI] | PVALUE | AAF |
|---|---|---|---|---|---|
| UKB | 27707\|52818\|25309 | 56254\|105268\|49118 | 1.025 [1.014,1.036] | 6.6e-06 | 0.485 |
| GHS | 2313\|4796\|2384 | 24697\|49910\|24787 | 1.019 [0.986,1.052] | 0.26 | 0.49 |
| MALMO | 102\|259\|139 | 6266\|13459\|7387 | 1.082 [0.949,1.233] | 0.24 | 0.512 |
| SINAI | 72\|155\|87 | 2589\|4998\|2437 | 1.162 [0.98,1.378] | 0.083 | 0.484 |
| FinnGen | 2340\|4803\|2464 | 32356\|61113\|28856 | 1.086 [1.054,1.12] | 1.1e-07 | 0.565 |
| **META** | **32534\|62831\|30383** | **122162\|234748\|112585** | **1.031 [1.021,1.042]** | **4.3e-10** | **0.49** |

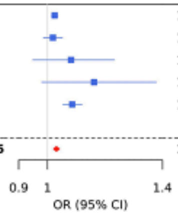


B

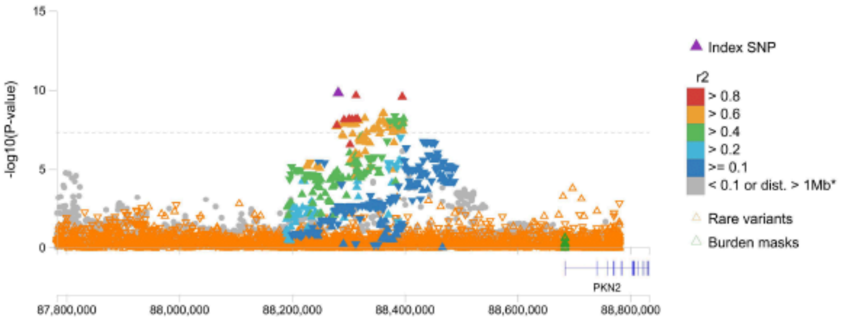


**Association with 1:88282464:G:A**

| STUDY | N CASES RR\|RA\|AA | N CONTROLS RR\|RA\|AA | OR [95% CI] | PVALUE | AAF |
|---|---|---|---|---|---|
| UKB | 8159\|42859\|54816 | 17370\|86039\|107231 | 1.036 [1.024,1.049] | 6.1e-09 | 0.716 |
| GHS | 706\|3759\|5028 | 7909\|39801\|51684 | 1.04 [1.004,1.077] | 0.031 | 0.717 |
| MALMO | 48\|217\|235 | 2597\|11506\|13009 | 0.986 [0.859,1.133] | 0.85 | 0.684 |
| SINAI | 17\|107\|190 | 577\|3560\|5887 | 1.049 [0.851,1.294] | 0.65 | 0.738 |
| FinnGen | 794\|3936\|4877 | 10524\|50713\|61088 | 1.028 [0.996,1.062] | 0.084 | 0.656 |
| **META** | **9724\|50878\|65146** | **38977\|191619\|238899** | **1.036 [1.024,1.047]** | **1.6e-10** | **0.714** |

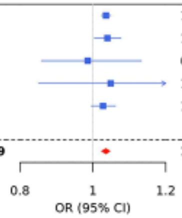

C

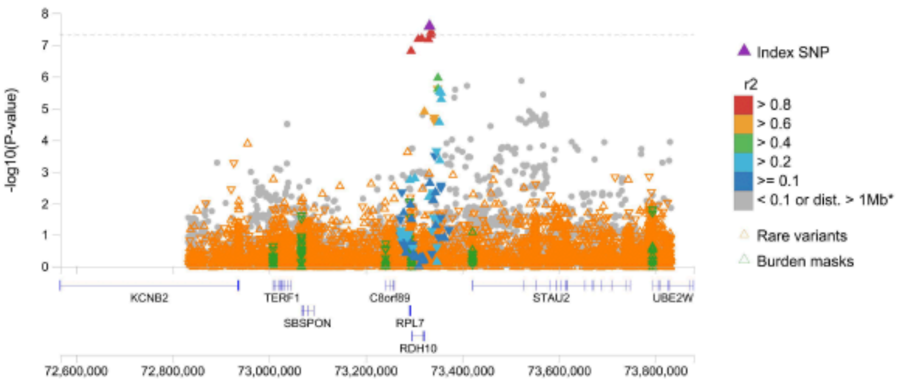


**Association with 8:73332501:T:C**

| STUDY | N CASES RR\|RA\|AA | N CONTROLS RR\|RA\|AA | OR [95% CI] | PVALUE | AAF |
|---|---|---|---|---|---|
| UKB | 67351\|34139\|4344 | 135545\|66806\|8289 | 1.033 [1.019,1.047] | 3.6e-06 | 0.199 |
| GHS | 6115\|3040\|338 | 65045\|30692\|3657 | 1.031 [0.99,1.074] | 0.13 | 0.191 |
| MALMO | 337\|145\|18 | 17942\|8227\|943 | 0.954 [0.809,1.124] | 0.57 | 0.186 |
| SINAI | 184\|110\|20 | 6087\|3449\|488 | 1.056 [0.866,1.286] | 0.59 | 0.221 |
| FinnGen | 6052\|3146\|409 | 79037\|38580\|4708 | 1.065 [1.024,1.108] | 0.0017 | 0.179 |
| **META** | **80039\|40580\|5129** | **303656\|147754\|18085** | **1.035 [1.023,1.048]** | **2.6e-08** | **0.197** |

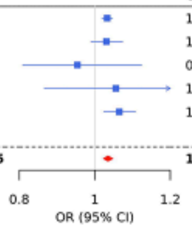


D

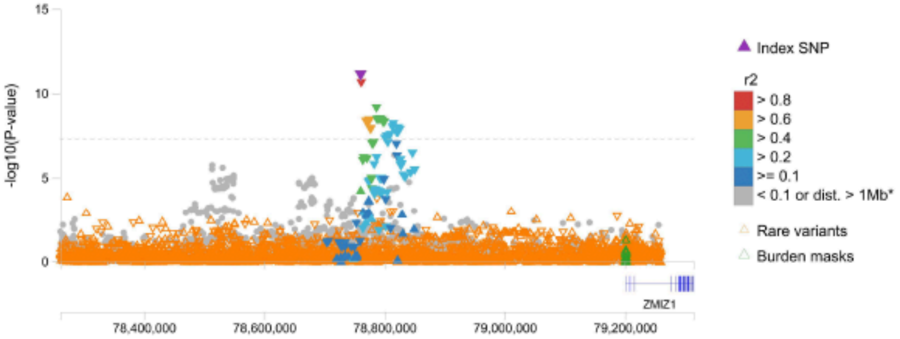


**Association with 10:78760556:T:C**

| STUDY | N CASES RR\|RA\|AA | N CONTROLS RR\|RA\|AA | OR [95% CI] | PVALUE | AAF |
|---|---|---|---|---|---|
| UKB | 65454\|35583\|4797 | 127824\|72736\|10080 | 0.959 [0.946,0.972] | 6.9e-10 | 0.219 |
| GHS | 5849\|3189\|455 | 60842\|33661\|4891 | 0.977 [0.94,1.015] | 0.23 | 0.218 |
| MALMO | 306\|169\|25 | 16609\|9200\|1303 | 1.007 [0.865,1.173] | 0.93 | 0.218 |
| SINAI | 218\|85\|11 | 6407\|3226\|391 | 0.853 [0.692,1.05] | 0.13 | 0.199 |
| FinnGen | 6126\|3091\|390 | 76053\|40800\|5472 | 0.941 [0.903,0.98] | 0.0033 | 0.166 |
| **META** | **77953\|42117\|5678** | **287735\|159623\|22137** | **0.959 [0.947,0.97]** | **7.1e-12** | **0.216** |

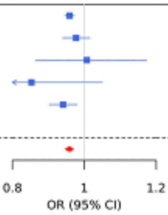

E

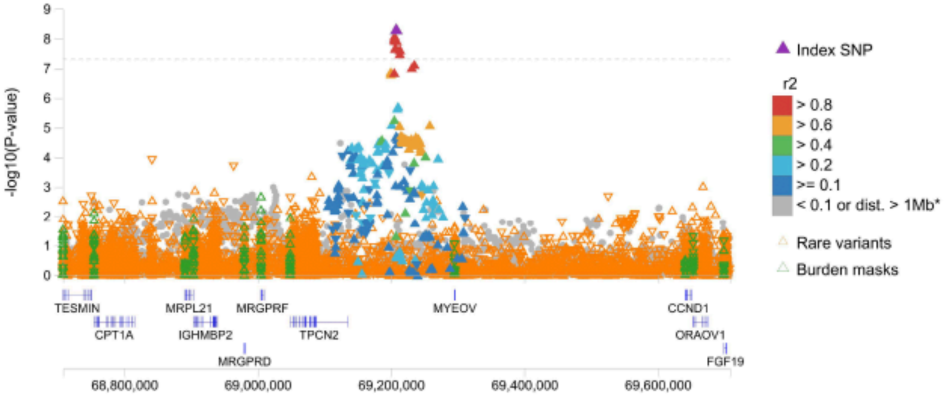


**Association with 11:69208057:T:C**

<table>
<tr><th>STUDY</th><th>N CASES<br>RR|RA|AA</th><th>N CONTROLS<br>RR|RA|AA</th><th>OR [95% CI]</th><th>PVALUE</th><th>AAF</th></tr>
<tr><td>UKB</td><td>15357|49729|40748</td><td>31512|99753|79375</td><td>1.031 [1.02,1.043]</td><td>7.1e-08</td><td>0.616</td></tr>
<tr><td>GHS</td><td>1285|4431|3777</td><td>14133|46571|38690</td><td>1.041 [1.008,1.076]</td><td>0.015</td><td>0.622</td></tr>
<tr><td>MALMO</td><td>74|257|169</td><td>4061|12819|10232</td><td>0.925 [0.813,1.053]</td><td>0.24</td><td>0.611</td></tr>
<tr><td>SINAI</td><td>35|143|136</td><td>1246|4417|4361</td><td>0.978 [0.822,1.164]</td><td>0.81</td><td>0.648</td></tr>
<tr><td>FinnGen</td><td>1375|4519|3713</td><td>17939|57811|46575</td><td>1.02 [0.988,1.053]</td><td>0.23</td><td>0.658</td></tr>
<tr><td>META</td><td>18126|59079|48543</td><td>68891|221371|179233</td><td>1.03 [1.02,1.041]</td><td>5.4e-09</td><td>0.618</td></tr>
</table>

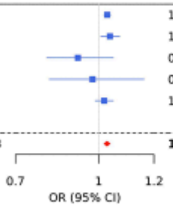


F

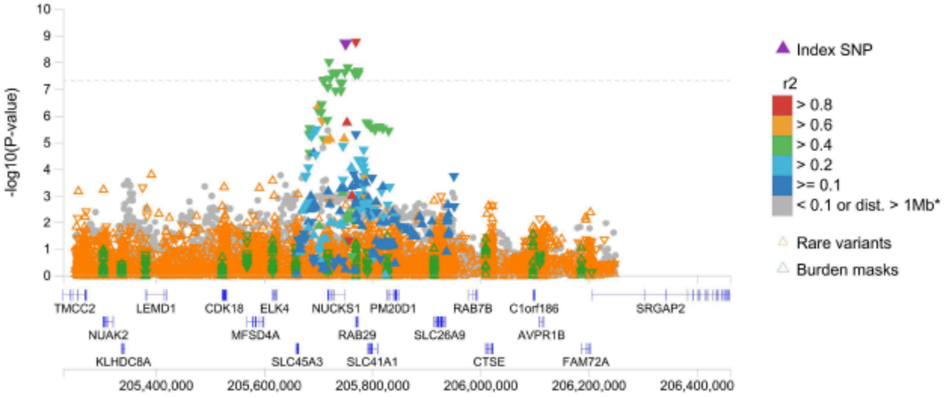


**Association with 1:205751355:G:A**

<table>
<tr><th>STUDY</th><th>N CASES<br>RR|RA|AA</th><th>N CONTROLS<br>RR|RA|AA</th><th>OR [95% CI]</th><th>PVALUE</th><th>AAF</th></tr>
<tr><td>UKB</td><td>22164|52368|31302</td><td>42556|104212|63872</td><td>0.969 [0.959,0.98]</td><td>2.1e-08</td><td>0.548</td></tr>
<tr><td>GHS</td><td>1893|4664|2936</td><td>19127|48859|31408</td><td>0.965 [0.934,0.996]</td><td>0.026</td><td>0.558</td></tr>
<tr><td>MALMO</td><td>89|240|171</td><td>5396|13374|8342</td><td>1.12 [0.985,1.273]</td><td>0.083</td><td>0.548</td></tr>
<tr><td>SINAI</td><td>62|147|105</td><td>1739|4965|3320</td><td>0.983 [0.825,1.171]</td><td>0.85</td><td>0.566</td></tr>
<tr><td>FinnGen</td><td>2086|4781|2739</td><td>25957|60784|35585</td><td>0.979 [0.949,1.009]</td><td>0.17</td><td>0.582</td></tr>
<tr><td>META</td><td>26294|62200|37253</td><td>94775|232194|142527</td><td>0.971 [0.961,0.98]</td><td>2e-09</td><td>0.549</td></tr>
</table>

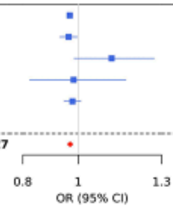

G

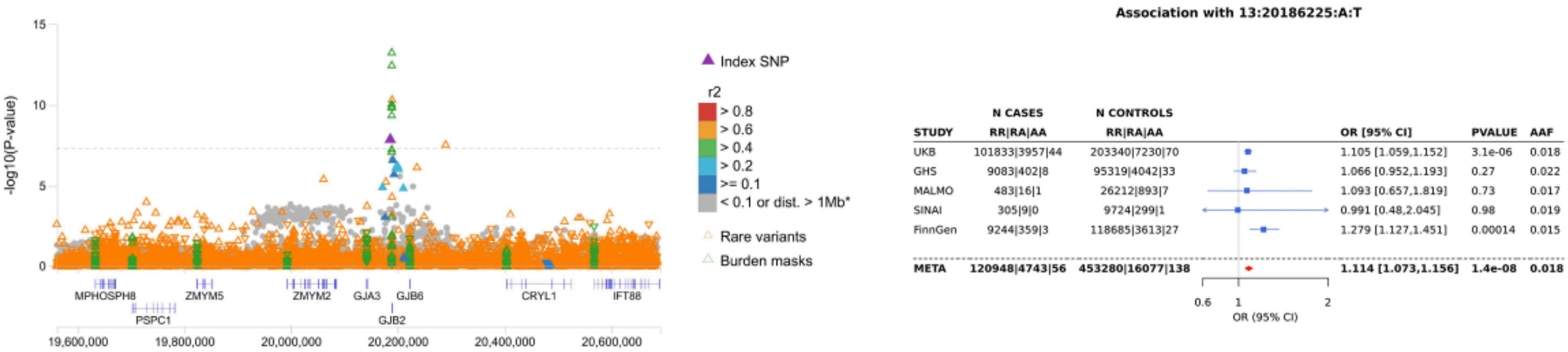
-log10(P-value)
15
10
5
0
MPHOSPH8
PSPC1
ZMYM5
ZMYM2
GJA3
GJB6
GJB2
CRYL1
IFT88
19,600,000
19,800,000
20,000,000
20,200,000
20,400,000
20,600,000
Index SNP
r2
> 0.8
> 0.6
> 0.4
> 0.2
>= 0.1
< 0.1 or dist. > 1Mb*
Rare variants
Burden masks
Association with 13:20186225:A:T
STUDY | N CASES RR|RA|AA | N CONTROLS RR|RA|AA | OR [95% CI] | PVALUE | AAF
UKB | 101833|3957|44 | 203340|7230|70 | 1.105 [1.059,1.152] | 3.1e-06 | 0.018
GHS | 9083|402|8 | 95319|4042|33 | 1.066 [0.952,1.193] | 0.27 | 0.022
MALMO | 483|16|1 | 26212|893|7 | 1.093 [0.657,1.819] | 0.73 | 0.017
SINAI | 305|9|0 | 9724|299|1 | 0.991 [0.48,2.045] | 0.98 | 0.019
FinnGen | 9244|359|3 | 118685|3613|27 | 1.279 [1.127,1.451] | 0.00014 | 0.015
META | 120948|4743|56 | 453280|16077|138 | 1.114 [1.073,1.156] | 1.4e-08 | 0.018
0.6
1
2
OR (95% CI)

H

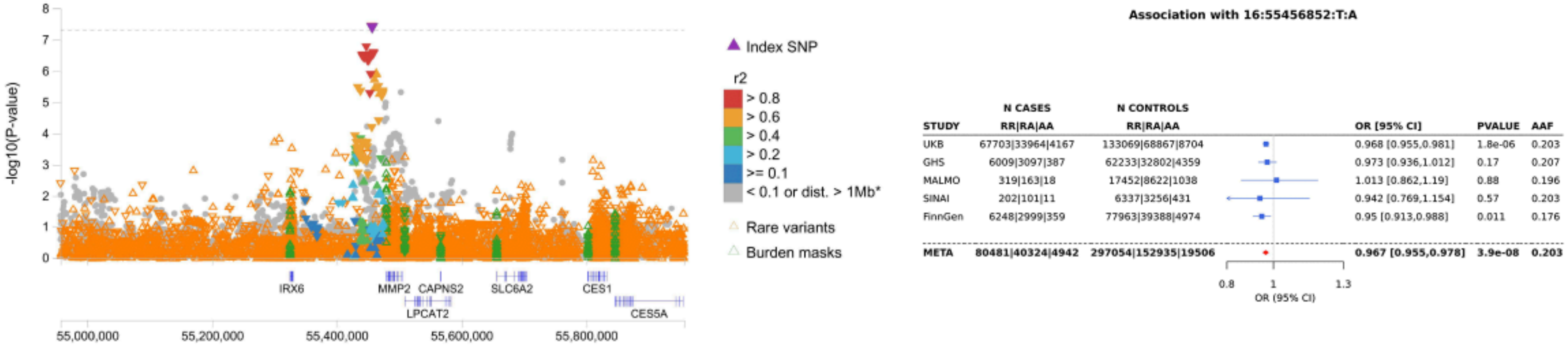
-log10(P-value)
8
7
6
5
4
3
2
1
0
IRX6
MMP2
CAPNS2
LPCAT2
SLC6A2
CES1
CES5A
55,000,000
55,200,000
55,400,000
55,600,000
55,800,000
Index SNP
r2
> 0.8
> 0.6
> 0.4
> 0.2
>= 0.1
< 0.1 or dist. > 1Mb*
Rare variants
Burden masks
Association with 16:55456852:T:A
STUDY | N CASES RR|RA|AA | N CONTROLS RR|RA|AA | OR [95% CI] | PVALUE | AAF
UKB | 67703|33964|4167 | 133069|68867|8704 | 0.968 [0.955,0.981] | 1.8e-06 | 0.203
GHS | 6009|3097|387 | 62233|32802|4359 | 0.973 [0.936,1.012] | 0.17 | 0.207
MALMO | 319|163|18 | 17452|8622|1038 | 1.013 [0.862,1.19] | 0.88 | 0.196
SINAI | 202|101|11 | 6337|3256|431 | 0.942 [0.769,1.154] | 0.57 | 0.203
FinnGen | 6248|2999|359 | 77963|39388|4974 | 0.95 [0.913,0.988] | 0.011 | 0.176
META | 80481|40324|4942 | 297054|152935|19506 | 0.967 [0.955,0.978] | 3.9e-08 | 0.203
0.8
1
1.3
OR (95% CI)

I

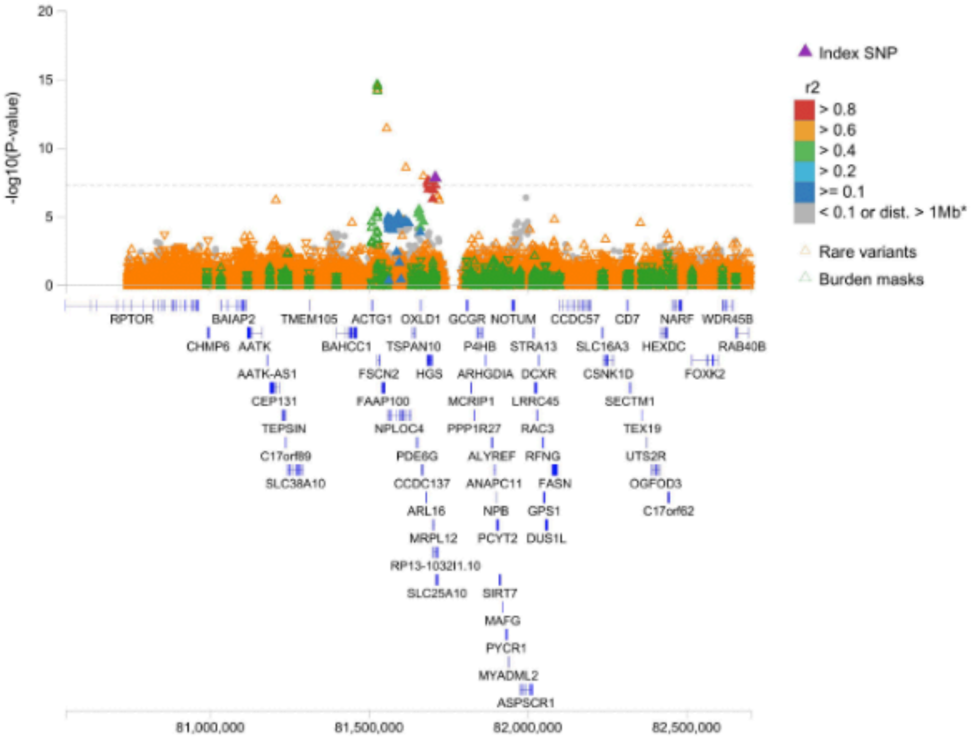

20
15
10
5
0
-log10(P-value)
Index SNP
r2
> 0.8
> 0.6
> 0.4
> 0.2
>= 0.1
< 0.1 or dist. > 1Mb*
Rare variants
Burden masks
RPTOR
BAIAP2
TMEM105
ACTG1
OXLD1
GCGR
NOTUM
CCDC57
CD7
NARF
WDR45B
CHMP6
AATK
BAHCC1
TSPAN10
P4HB
STRA13
SLC16A3
HEXDC
RAB40B
AATK-AS1
FSCN2
HGS
ARHGDIA
DCXR
CSNK1D
FOXK2
CEP131
FAAP100
MCRIP1
LRRC45
SECTM1
TEPSIN
NPLOC4
PPP1R27
RAC3
TEX19
C17orf89
PDE6G
ALYREF
RFNG
UTS2R
SLC38A10
CCDC137
ANAPC11
FASN
OGFOD3
ARL16
NPB
GPS1
C17orf62
MRPL12
PCYT2
DUS1L
RP13-1032I1.10
SLC25A10
SIRT7
MAFG
PYCR1
MYADML2
ASPSCR1
81,000,000
81,500,000
82,000,000
82,500,000


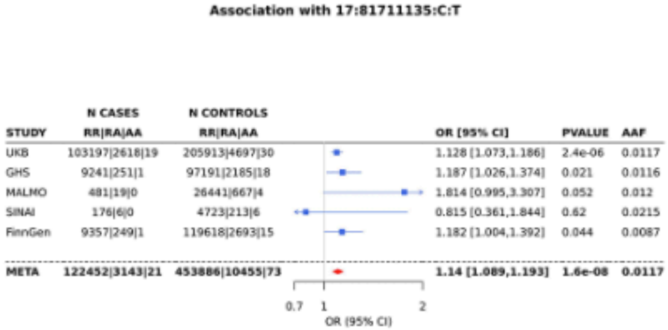

Association with 17:81711135:C:T
N CASES
N CONTROLS
STUDY
RR|RA|AA
RR|RA|AA
OR [95% CI]
PVALUE
AAF
UKB 103197|2618|19 205913|4697|30 1.128 [1.073,1.186] 2.4e-06 0.0117
GHS 9241|251|1 97191|2185|18 1.187 [1.026,1.374] 0.021 0.0116
MALMO 481|19|0 26441|667|4 1.814 [0.995,3.307] 0.052 0.012
SINAI 176|6|0 4723|213|6 0.815 [0.361,1.844] 0.62 0.0215
FinnGen 9357|249|1 119618|2693|15 1.182 [1.004,1.392] 0.044 0.0087
META 122452|3143|21 453886|10455|73 1.14 [1.089,1.193] 1.6e-08 0.0117
0.7
1
2
OR (95% CI)

J

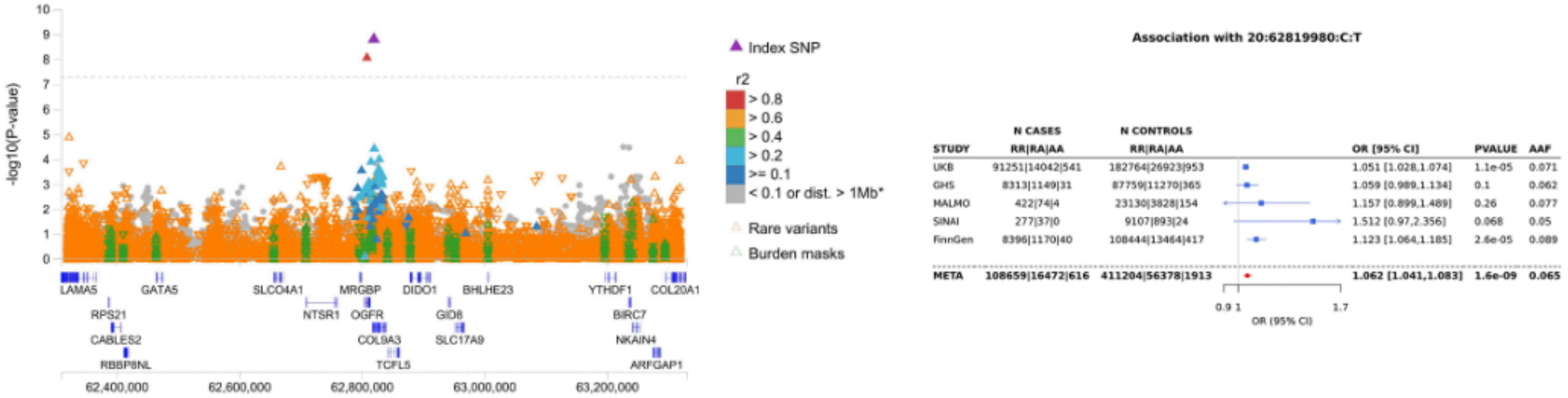

-log10(P-value)
10
9
8
7
6
5
4
3
2
1
0
LAMA5
GATA5
SLCO4A1
MRGBP
DIDO1
BHLHE23
YTHDF1
COL20A1
RPS21
NTSR1
OGFR
GID8
BIRC7
CABLES2
COL9A3
SLC17A9
NKAIN4
RBBP8NL
TCFL5
ARFGAP1
62,400,000
62,600,000
62,800,000
63,000,000
63,200,000
Index SNP
r2
> 0.8
> 0.6
> 0.4
> 0.2
>= 0.1
< 0.1 or dist. > 1Mb*
Rare variants
Burden masks
Association with 20:62819980:C:T
STUDY
N CASES RR|RA|AA
N CONTROLS RR|RA|AA
OR [95% CI]
PVALUE
AAF
UKB 91251|14042|541 182764|26923|953 1.051 [1.028,1.074] 1.1e-05 0.071
GHS 8313|1149|31 87759|11270|365 1.059 [0.989,1.134] 0.1 0.062
MALMO 422|74|4 23130|3828|154 1.157 [0.899,1.489] 0.26 0.077
SINAI 277|37|0 9107|893|24 1.512 [0.97,2.356] 0.068 0.05
FinnGen 8396|1170|40 108444|13464|417 1.123 [1.064,1.185] 2.6e-05 0.089
META 108659|16472|616 411204|56378|1913 1.062 [1.041,1.083] 1.6e-09 0.065
0.9 1
1.7
OR (95% CI)


K

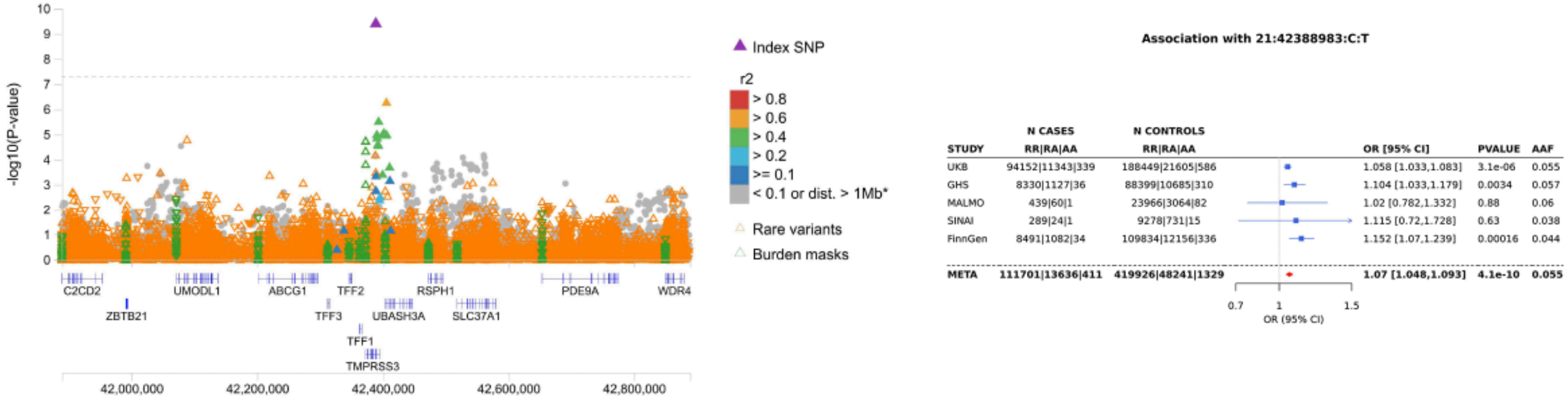

-log10(P-value)
10
9
8
7
6
5
4
3
2
1
0
C2CD2
UMODL1
ABCG1
TFF2
RSPH1
PDE9A
WDR4
ZBTB21
TFF3
UBASH3A
SLC37A1
TFF1
TMPRSS3
42,000,000
42,200,000
42,400,000
42,600,000
42,800,000
Index SNP
r2
> 0.8
> 0.6
> 0.4
> 0.2
>= 0.1
< 0.1 or dist. > 1Mb*
Rare variants
Burden masks
Association with 21:42388983:C:T
STUDY
N CASES RR|RA|AA
N CONTROLS RR|RA|AA
OR [95% CI]
PVALUE
AAF
UKB 94152|11343|339 188449|21605|586 1.058 [1.033,1.083] 3.1e-06 0.055
GHS 8330|1127|36 88399|10685|310 1.104 [1.033,1.179] 0.0034 0.057
MALMO 439|60|1 23966|3064|82 1.02 [0.782,1.332] 0.88 0.06
SINAI 289|24|1 9278|731|15 1.115 [0.72,1.728] 0.63 0.038
FinnGen 8491|1082|34 109834|12156|336 1.152 [1.07,1.239] 0.00016 0.044
META 111701|13636|411 419926|48241|1329 1.07 [1.048,1.093] 4.1e-10 0.055
0.7
1
1.5
OR (95% CI)

L

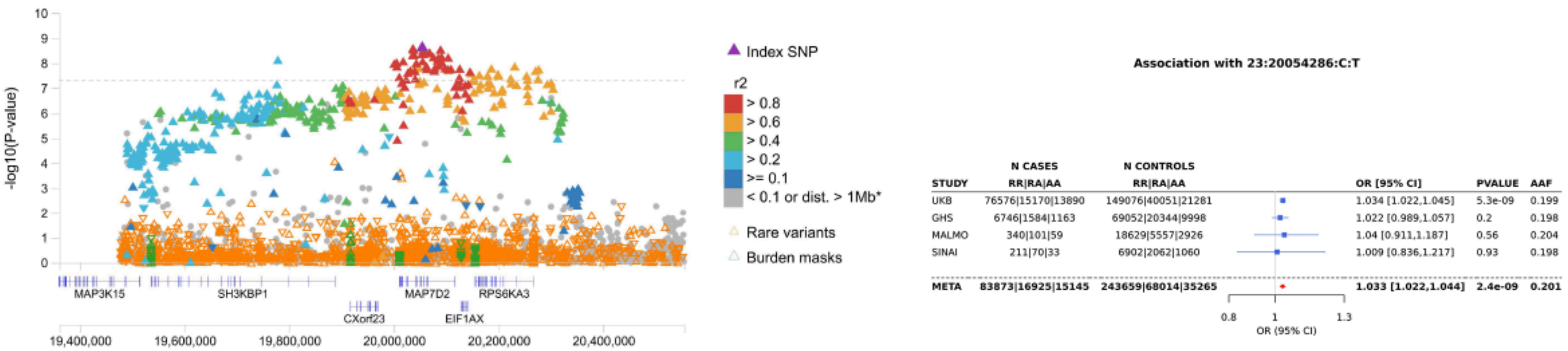

-log10(P-value)
10
9
8
7
6
5
4
3
2
1
0
MAP3K15
SH3KBP1
MAP7D2
RPS6KA3
CXorf23
EIF1AX
19,400,000
19,600,000
19,800,000
20,000,000
20,200,000
20,400,000
Index SNP
r2
> 0.8
> 0.6
> 0.4
> 0.2
>= 0.1
< 0.1 or dist. > 1Mb*
Rare variants
Burden masks
Association with 23:20054286:C:T
STUDY | N CASES RR|RA|AA | N CONTROLS RR|RA|AA | OR [95% CI] | PVALUE | AAF
UKB | 76576|15170|13890 | 149076|40051|21281 | 1.034 [1.022,1.045] | 5.3e-09 | 0.199
GHS | 6746|1584|1163 | 69052|20344|9998 | 1.022 [0.989,1.057] | 0.2 | 0.198
MALMO | 340|101|59 | 18629|5557|2926 | 1.04 [0.911,1.187] | 0.56 | 0.204
SINAI | 211|70|33 | 6902|2062|1060 | 1.009 [0.836,1.217] | 0.93 | 0.198
META | 83873|16925|15145 | 243659|68014|35265 | 1.033 [1.022,1.044] | 2.4e-09 | 0.201
0.8
1
1.3
OR (95% CI)


M

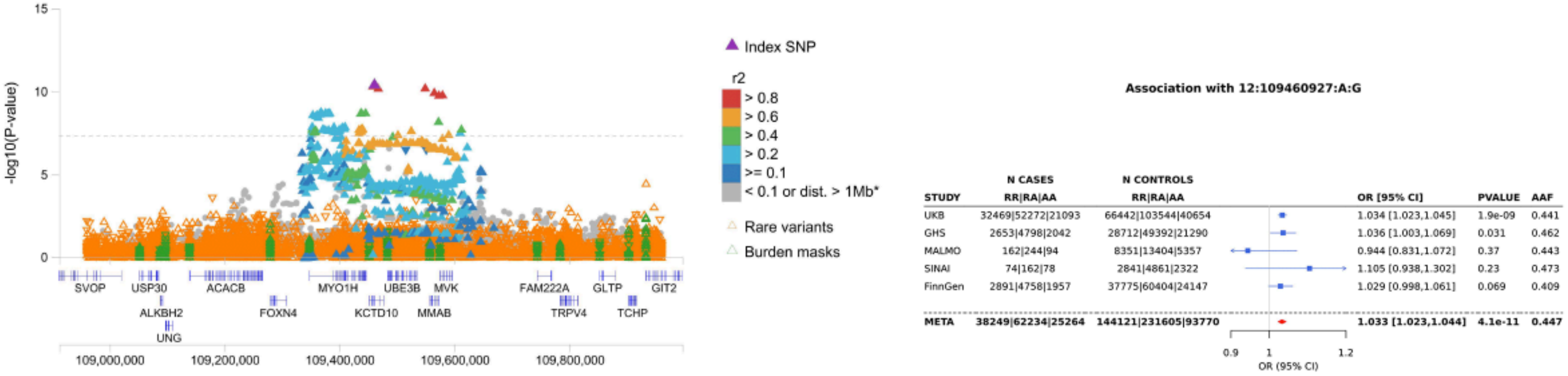

-log10(P-value)
15
10
5
0
SVOP
USP30
ACACB
MYO1H
UBE3B
MVK
FAM222A
GLTP
GIT2
ALKBH2
FOXN4
KCTD10
MMAB
TRPV4
TCHP
UNG
109,000,000
109,200,000
109,400,000
109,600,000
109,800,000
Index SNP
r2
> 0.8
> 0.6
> 0.4
> 0.2
>= 0.1
< 0.1 or dist. > 1Mb*
Rare variants
Burden masks
Association with 12:109460927:A:G
STUDY | N CASES RR|RA|AA | N CONTROLS RR|RA|AA | OR [95% CI] | PVALUE | AAF
UKB | 32469|52272|21093 | 66442|103544|40654 | 1.034 [1.023,1.045] | 1.9e-09 | 0.441
GHS | 2653|4798|2042 | 28712|49392|21290 | 1.036 [1.003,1.069] | 0.031 | 0.462
MALMO | 162|244|94 | 8351|13404|5357 | 0.944 [0.831,1.072] | 0.37 | 0.443
SINAI | 74|162|78 | 2841|4861|2322 | 1.105 [0.938,1.302] | 0.23 | 0.473
FinnGen | 2891|4758|1957 | 37775|60404|24147 | 1.029 [0.998,1.061] | 0.069 | 0.409
META | 38249|62234|25264 | 144121|231605|93770 | 1.033 [1.023,1.044] | 4.1e-11 | 0.447
0.9
1
1.2
OR (95% CI)

N

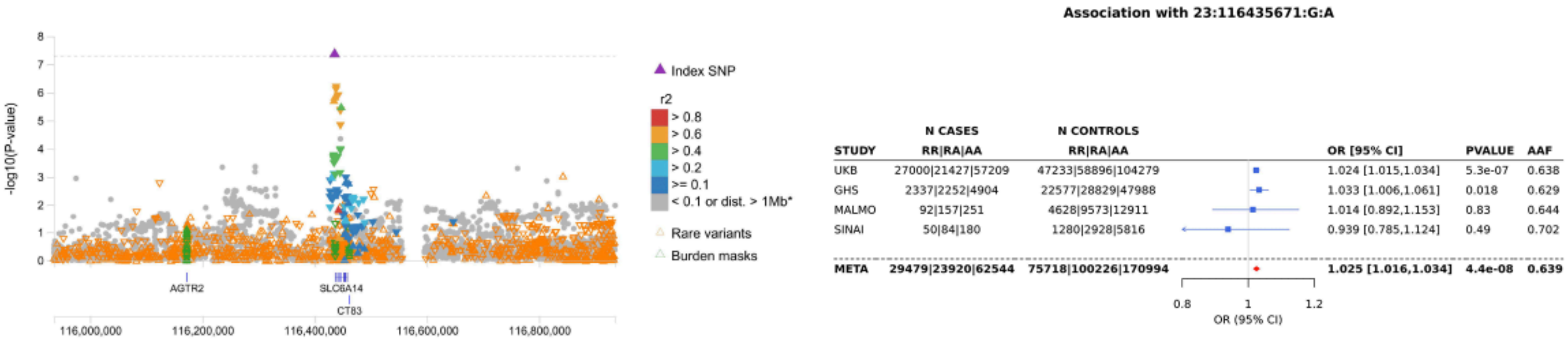

-log10(P-value)
Index SNP
r2
> 0.8
> 0.6
> 0.4
> 0.2
>= 0.1
< 0.1 or dist. > 1Mb*
Rare variants
Burden masks
AGTR2
SLC6A14
CT83
116,000,000
116,200,000
116,400,000
116,600,000
116,800,000
Association with 23:116435671:G:A
STUDY | N CASES RR|RA|AA | N CONTROLS RR|RA|AA | OR [95% CI] | PVALUE | AAF
UKB | 27000|21427|57209 | 47233|58896|104279 | 1.024 [1.015,1.034] | 5.3e-07 | 0.638
GHS | 2337|2252|4904 | 22577|28829|47988 | 1.033 [1.006,1.061] | 0.018 | 0.629
MALMO | 92|157|251 | 4628|9573|12911 | 1.014 [0.892,1.153] | 0.83 | 0.644
SINAI | 50|84|180 | 1280|2928|5816 | 0.939 [0.785,1.124] | 0.49 | 0.702
META | 29479|23920|62544 | 75718|100226|170994 | 1.025 [1.016,1.034] | 4.4e-08 | 0.639
0.8
1
1.2
OR (95% CI)


O

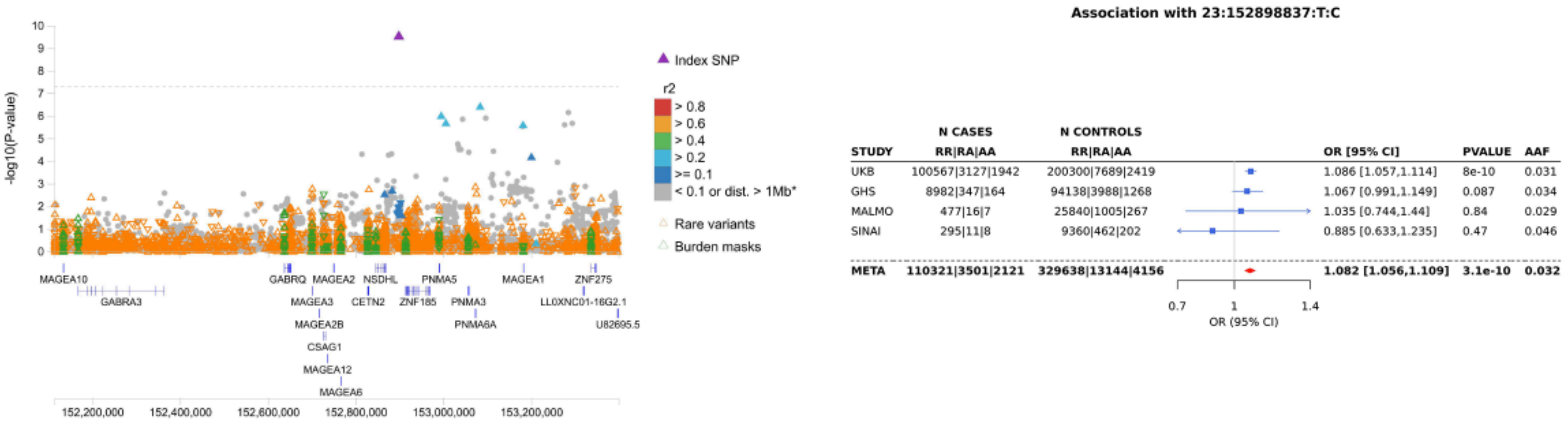

-log10(P-value)
Index SNP
r2
> 0.8
> 0.6
> 0.4
> 0.2
>= 0.1
< 0.1 or dist. > 1Mb*
Rare variants
Burden masks
MAGEA10
GABRA3
GABRQ
MAGEA2
MAGEA3
MAGEA2B
CSAG1
MAGEA12
MAGEA6
NSDHL
CETN2
ZNF185
PNMA5
PNMA3
PNMA6A
MAGEA1
ZNF275
LLOXNC01-16G2.1
U82695.5
152,200,000
152,400,000
152,600,000
152,800,000
153,000,000
153,200,000
Association with 23:152898837:T:C
STUDY | N CASES RR|RA|AA | N CONTROLS RR|RA|AA | OR [95% CI] | PVALUE | AAF
UKB | 100567|3127|1942 | 200300|7689|2419 | 1.086 [1.057,1.114] | 8e-10 | 0.031
GHS | 8982|347|164 | 94138|3988|1268 | 1.067 [0.991,1.149] | 0.087 | 0.034
MALMO | 477|16|7 | 25840|1005|267 | 1.035 [0.744,1.44] | 0.84 | 0.029
SINAI | 295|11|8 | 9360|462|202 | 0.885 [0.633,1.235] | 0.47 | 0.046
META | 110321|3501|2121 | 329638|13144|4156 | 1.082 [1.056,1.109] | 3.1e-10 | 0.032
0.7
1
1.4
OR (95% CI)
