## Supplementary Figure 5 for "Population-scale analysis of common and rare genetic variation associated with hearing loss in adults"

Adipose_Subcutaneous: CCDC68 GWAS P−values

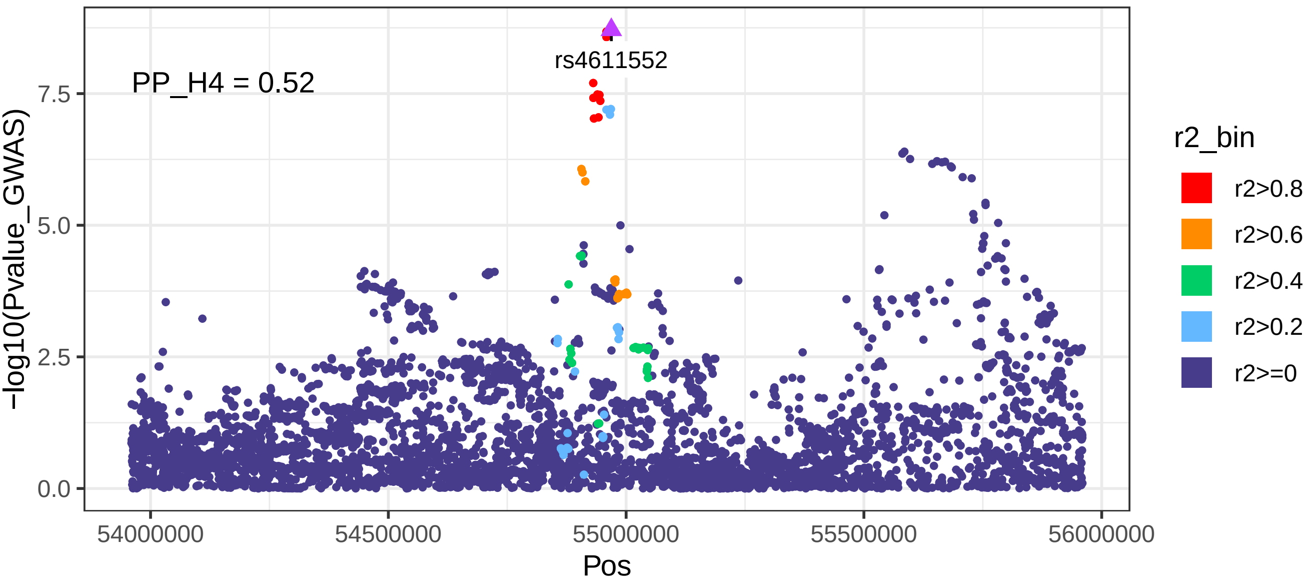


Adipose_Subcutaneous: CCDC68 eQTL P−values

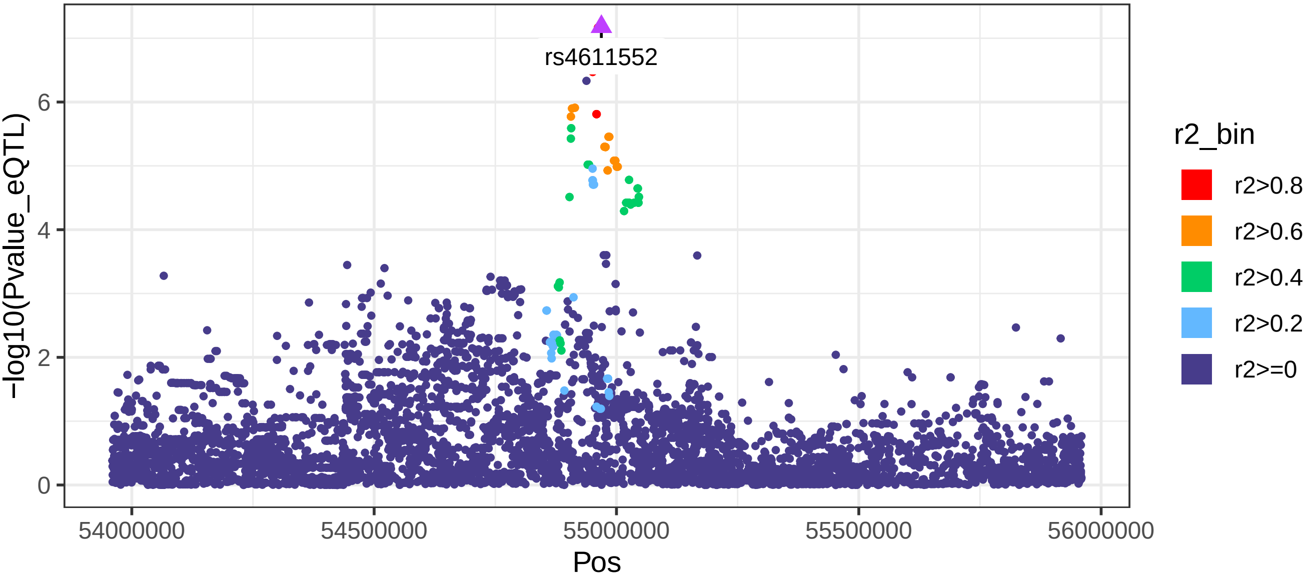

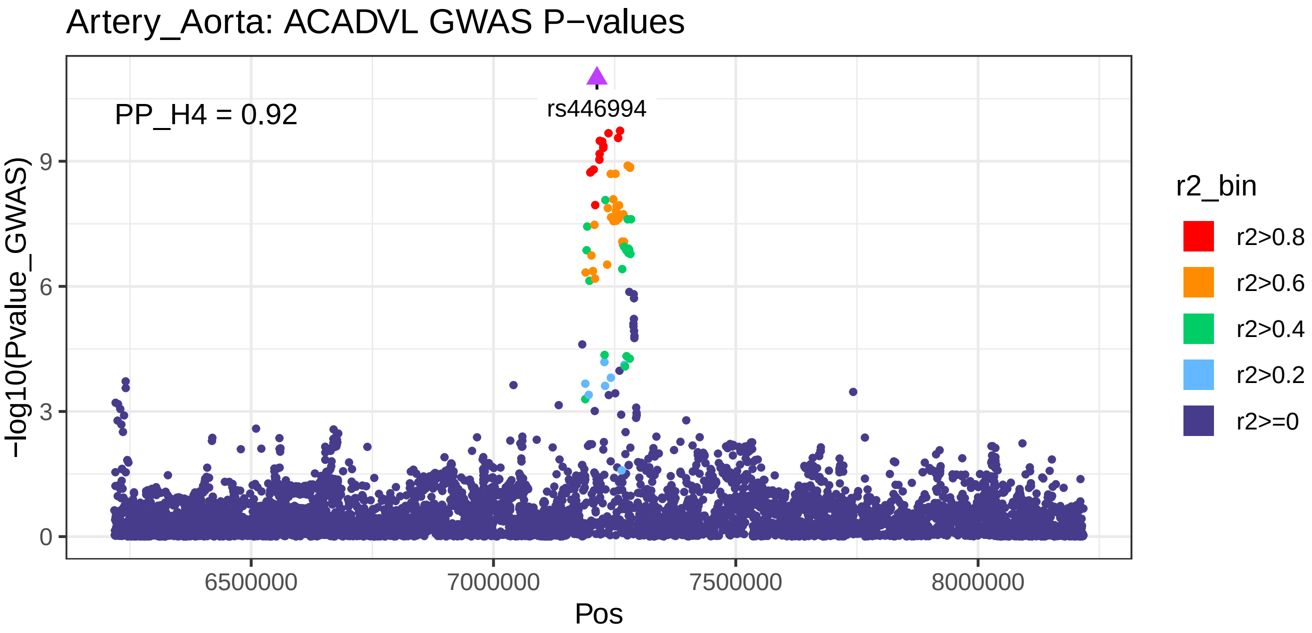
Artery_Aorta: ACADVL GWAS P−values
PP_H4 = 0.92
rs446994
−log10(Pvalue_GWAS)
0
3
6
9
6500000
7000000
7500000
8000000
Pos
r2_bin
r2>0.8
r2>0.6
r2>0.4
r2>0.2
r2>=0

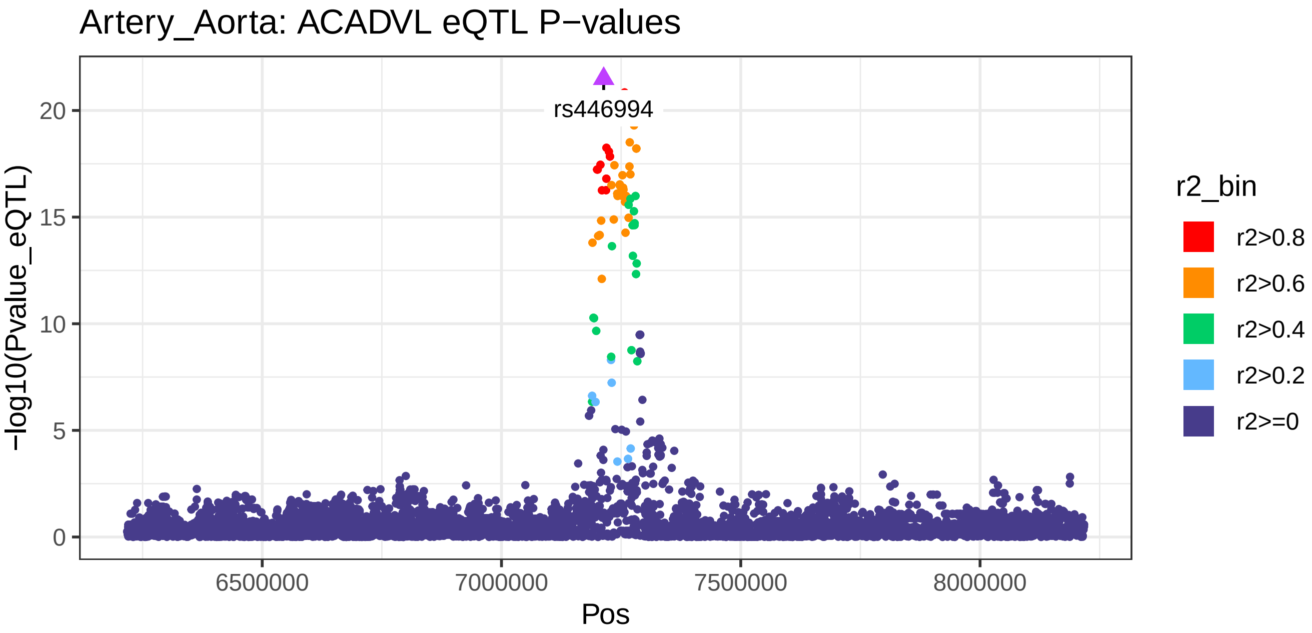
Artery_Aorta: ACADVL eQTL P−values
rs446994
−log10(Pvalue_eQTL)
0
5
10
15
20
6500000
7000000
7500000
8000000
Pos
r2_bin
r2>0.8
r2>0.6
r2>0.4
r2>0.2
r2>=0

Artery_Aorta: RAB29 GWAS P−values

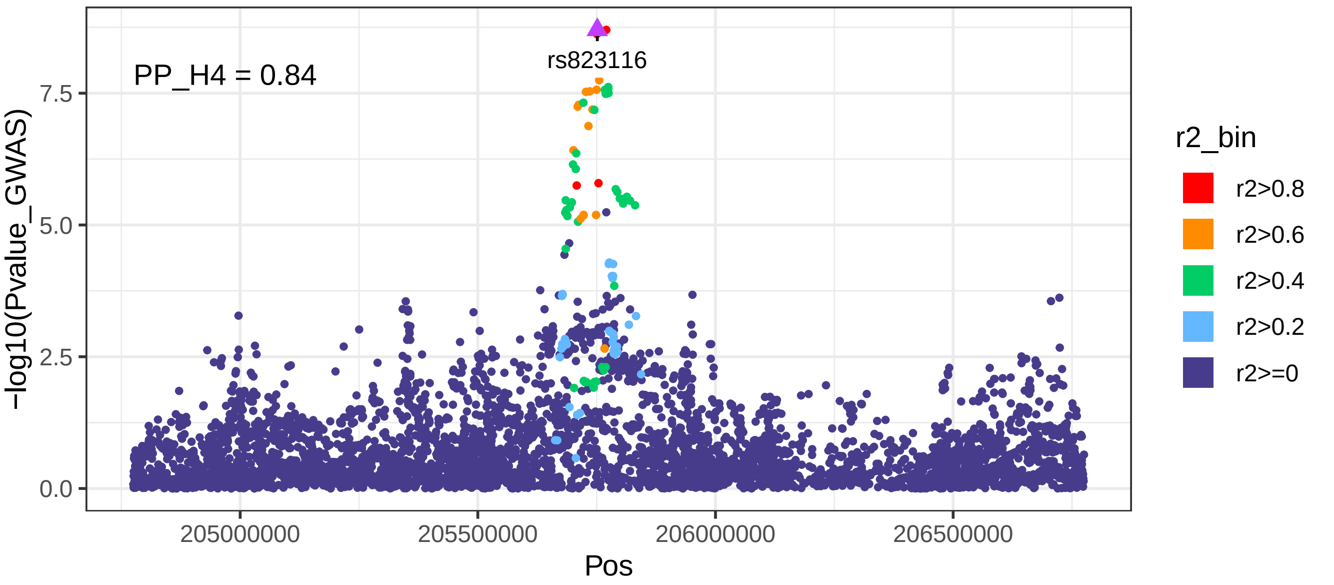


Artery_Aorta: RAB29 eQTL P−values

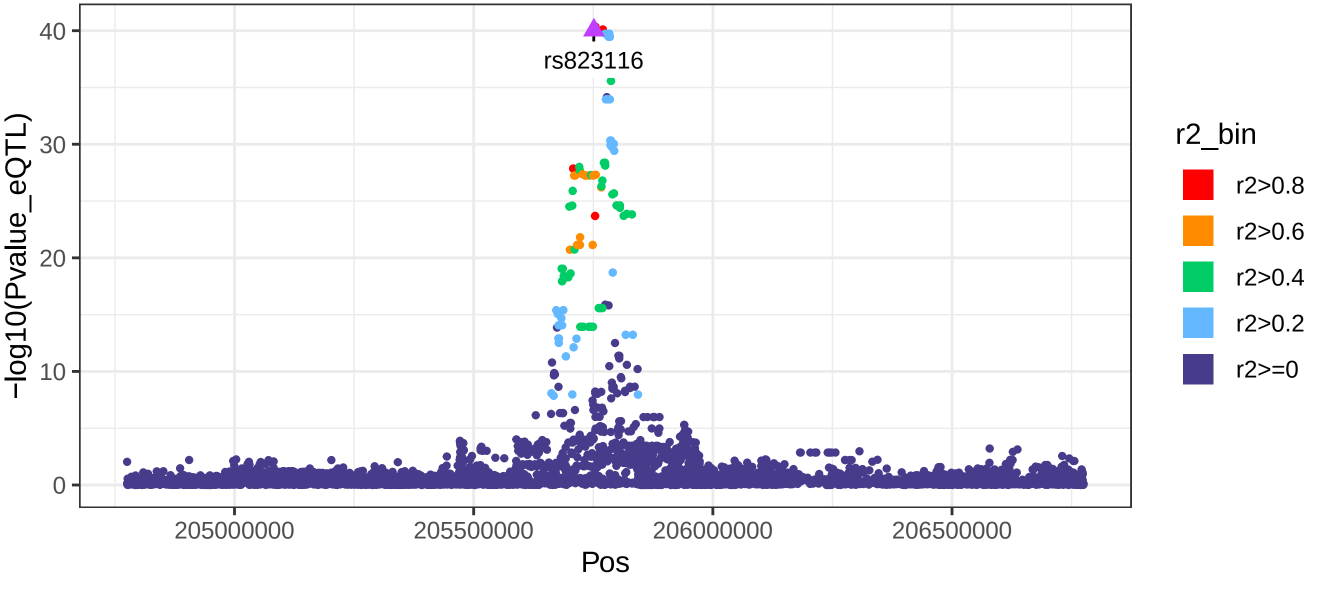

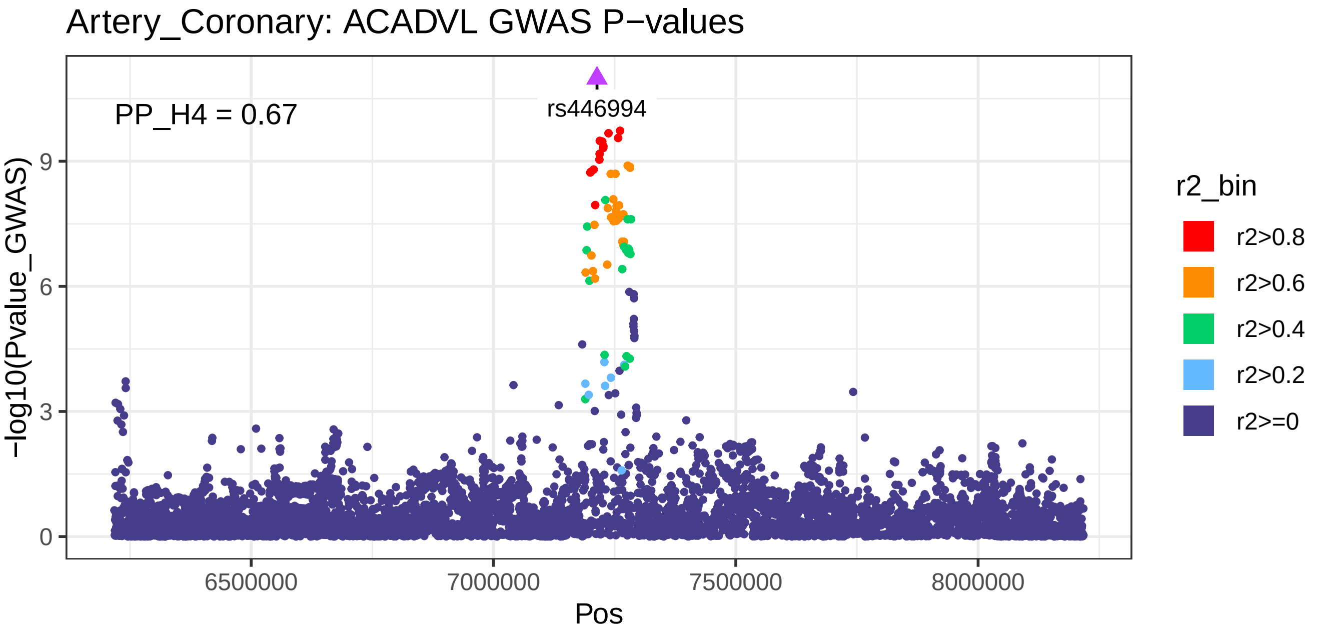
Artery_Coronary: ACADVL GWAS P−values
PP_H4 = 0.67
rs446994
−log10(Pvalue_GWAS)
0
3
6
9
6500000
7000000
7500000
8000000
Pos
r2_bin
r2>0.8
r2>0.6
r2>0.4
r2>0.2
r2>=0

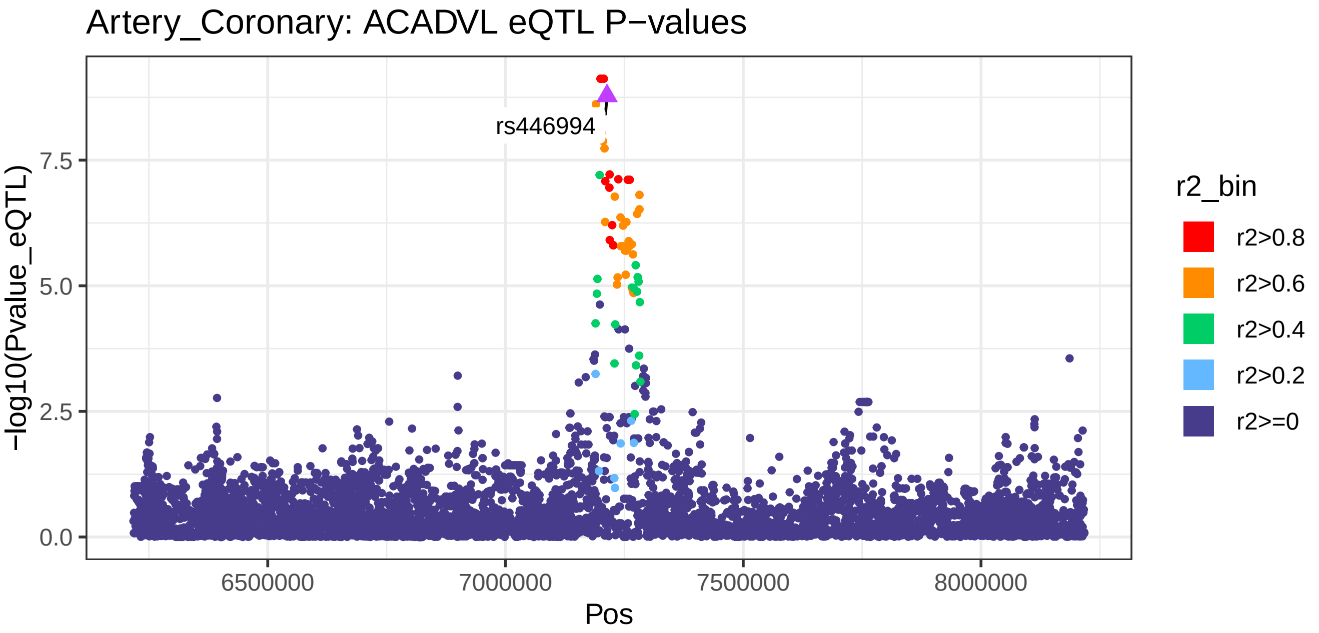
Artery_Coronary: ACADVL eQTL P−values
rs446994
−log10(Pvalue_eQTL)
0.0
2.5
5.0
7.5
6500000
7000000
7500000
8000000
Pos
r2_bin
r2>0.8
r2>0.6
r2>0.4
r2>0.2
r2>=0

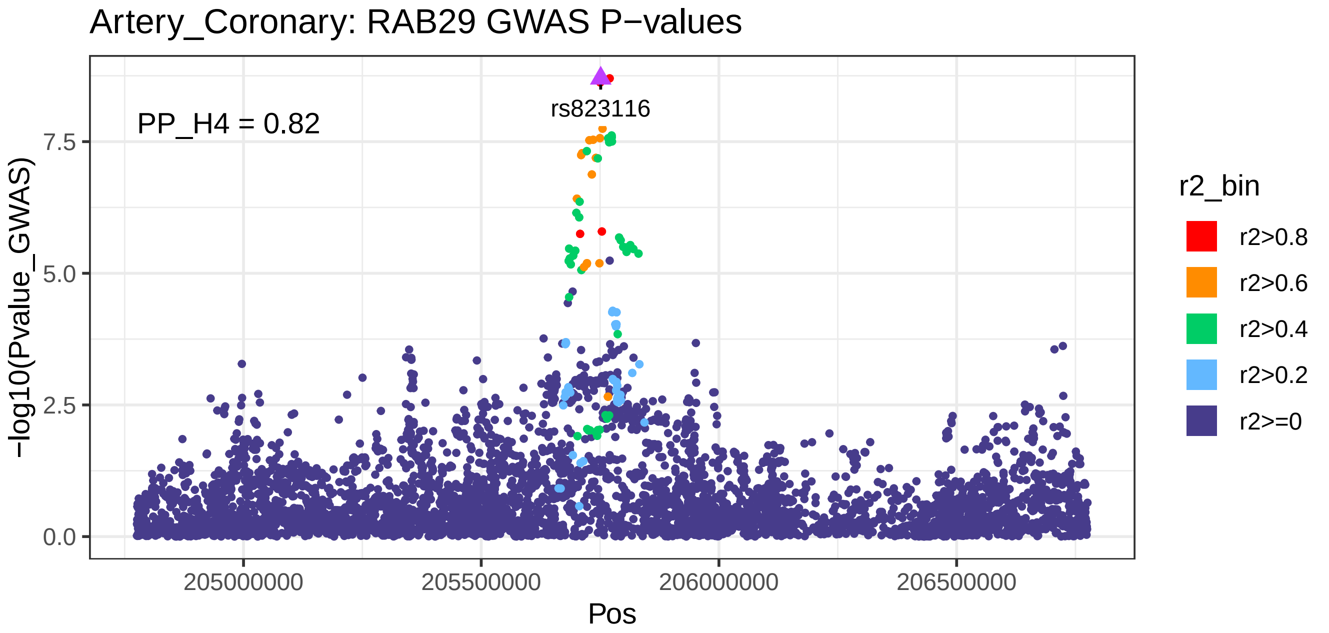
Artery_Coronary: RAB29 GWAS P−values
PP_H4 = 0.82
rs823116
−log10(Pvalue_GWAS)
0.0
2.5
5.0
7.5
205000000
205500000
206000000
206500000
Pos
r2_bin
r2>0.8
r2>0.6
r2>0.4
r2>0.2
r2>=0

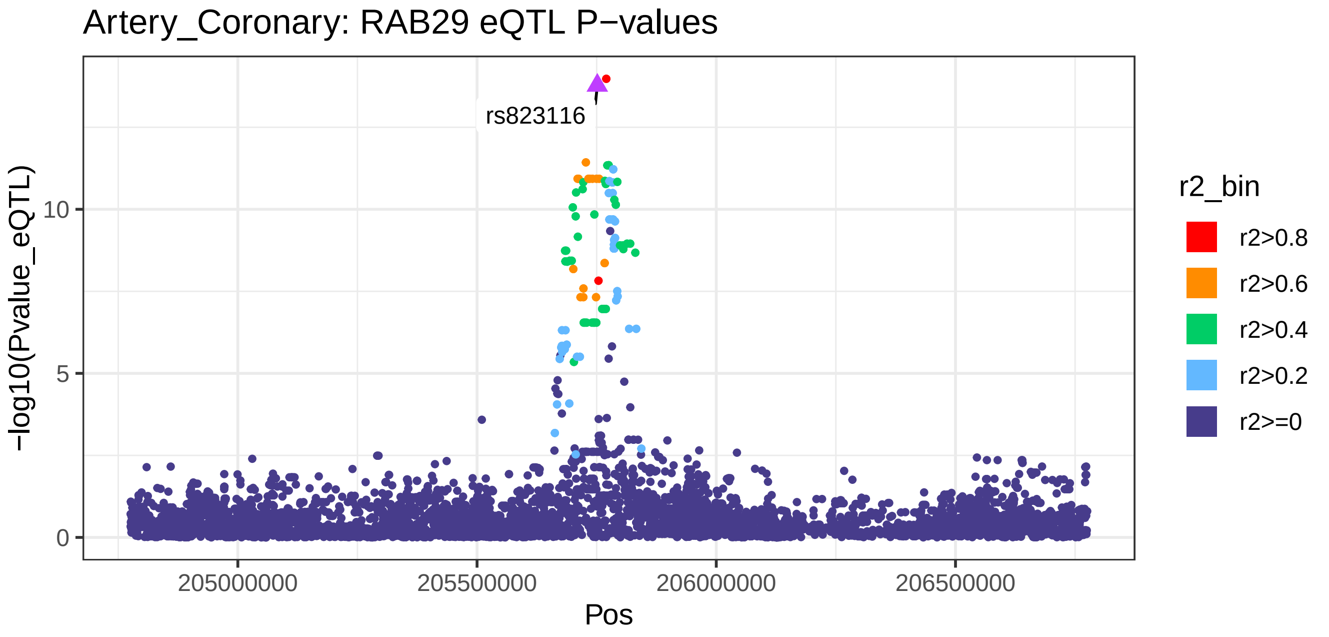
Artery_Coronary: RAB29 eQTL P−values
rs823116
−log10(Pvalue_eQTL)
0
5
10
205000000
205500000
206000000
206500000
Pos
r2_bin
r2>0.8
r2>0.6
r2>0.4
r2>0.2
r2>=0

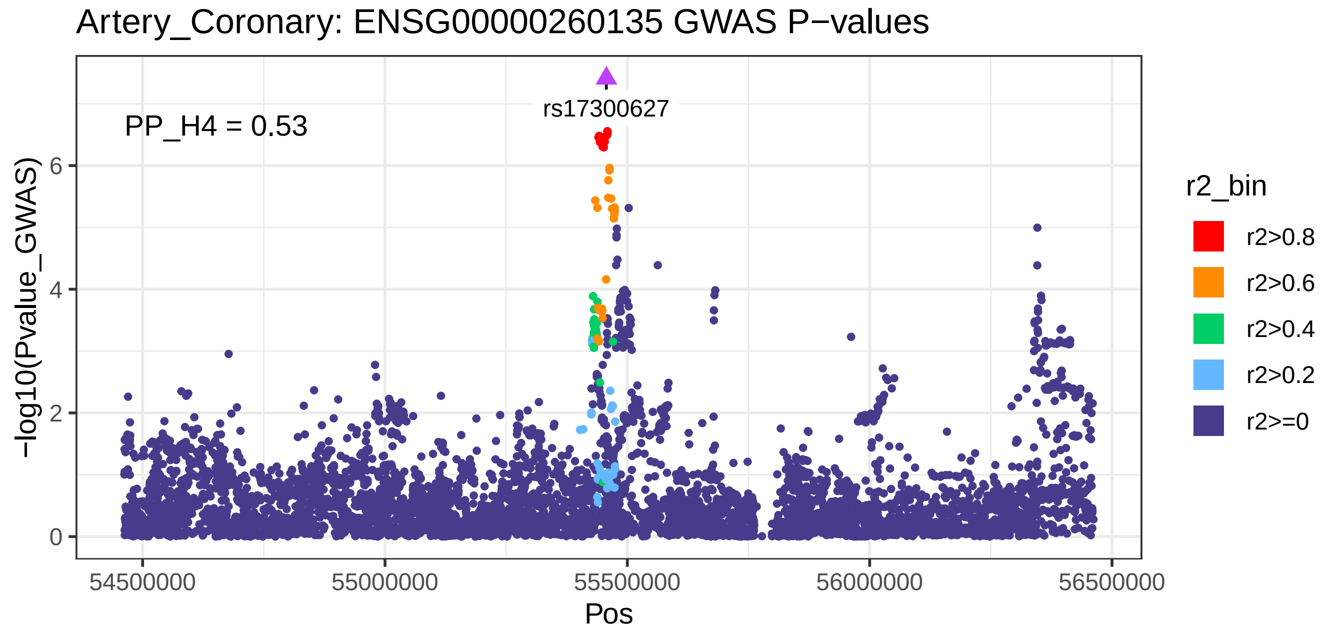
Artery_Coronary: ENSG00000260135 GWAS P−values
PP_H4 = 0.53
rs17300627
−log10(Pvalue_GWAS)
Pos
54500000
55000000
55500000
56000000
56500000
0
2
4
6
r2_bin
r2>0.8
r2>0.6
r2>0.4
r2>0.2
r2>=0

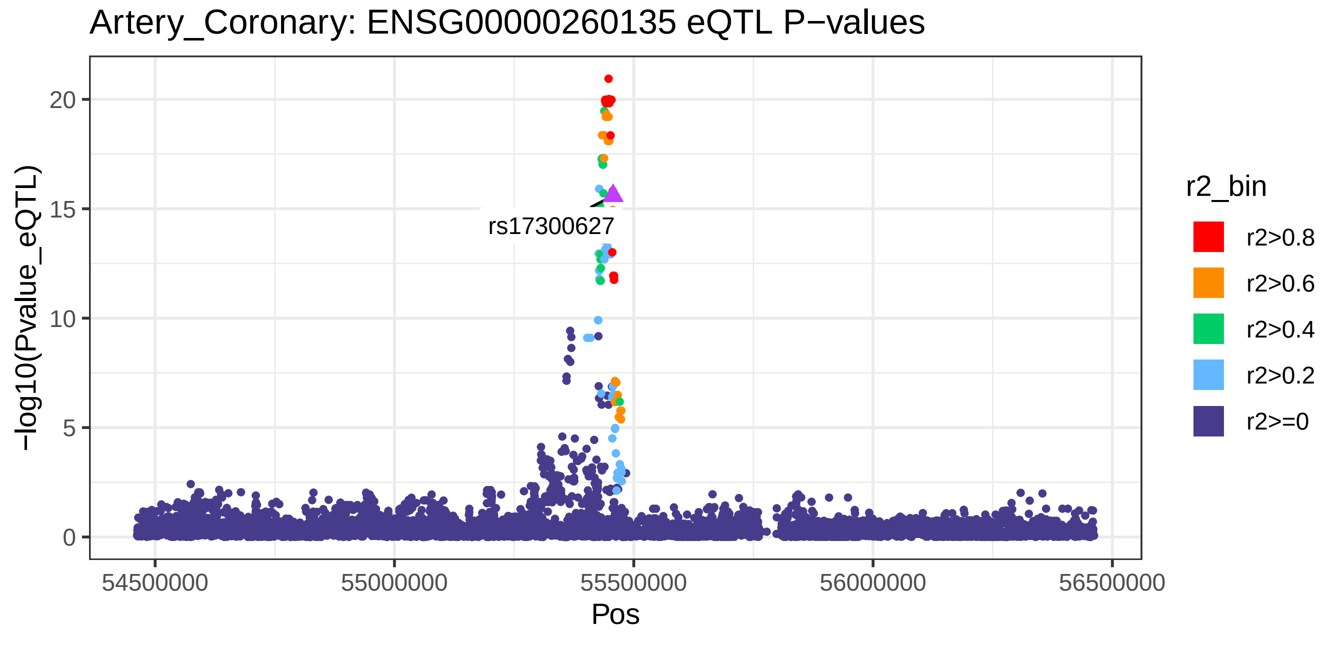
Artery_Coronary: ENSG00000260135 eQTL P−values
rs17300627
−log10(Pvalue_eQTL)
Pos
54500000
55000000
55500000
56000000
56500000
0
5
10
15
20
r2_bin
r2>0.8
r2>0.6
r2>0.4
r2>0.2
r2>=0

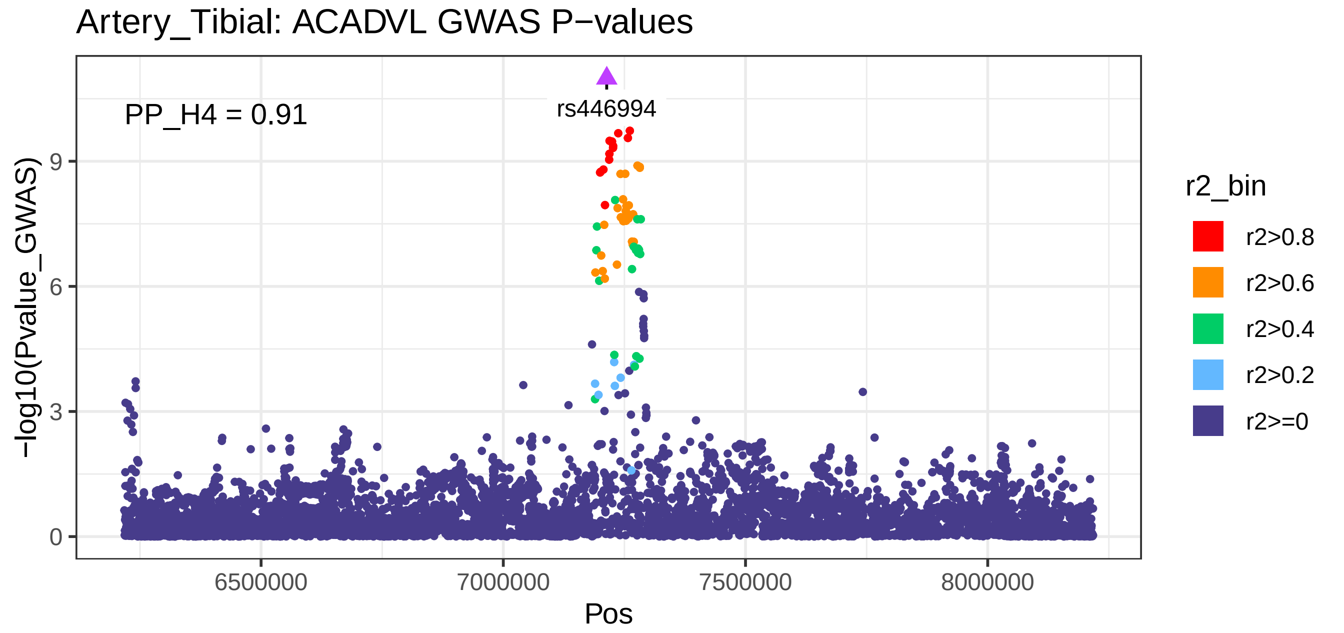
Artery_Tibial: ACADVL GWAS P−values
PP_H4 = 0.91
rs446994
−log10(Pvalue_GWAS)
0
3
6
9
6500000
7000000
7500000
8000000
Pos
r2_bin
r2>0.8
r2>0.6
r2>0.4
r2>0.2
r2>=0

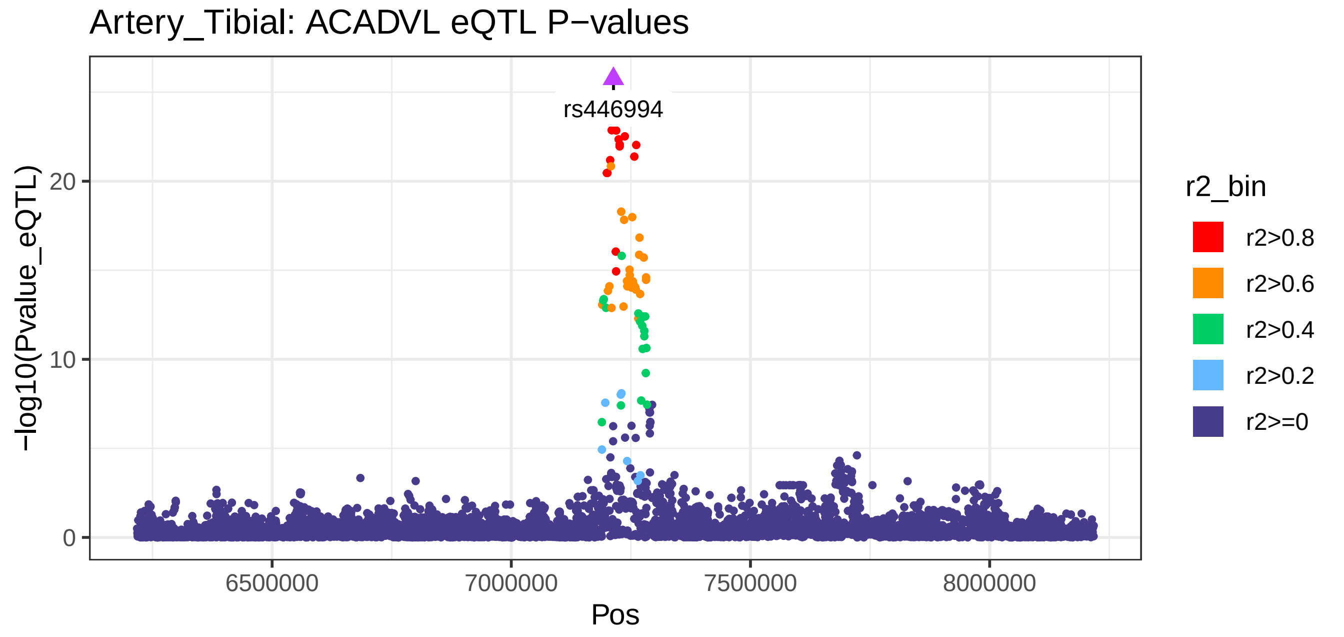
Artery_Tibial: ACADVL eQTL P−values
rs446994
−log10(Pvalue_eQTL)
0
10
20
6500000
7000000
7500000
8000000
Pos
r2_bin
r2>0.8
r2>0.6
r2>0.4
r2>0.2
r2>=0

Artery_Tibial: RAB29 GWAS P−values

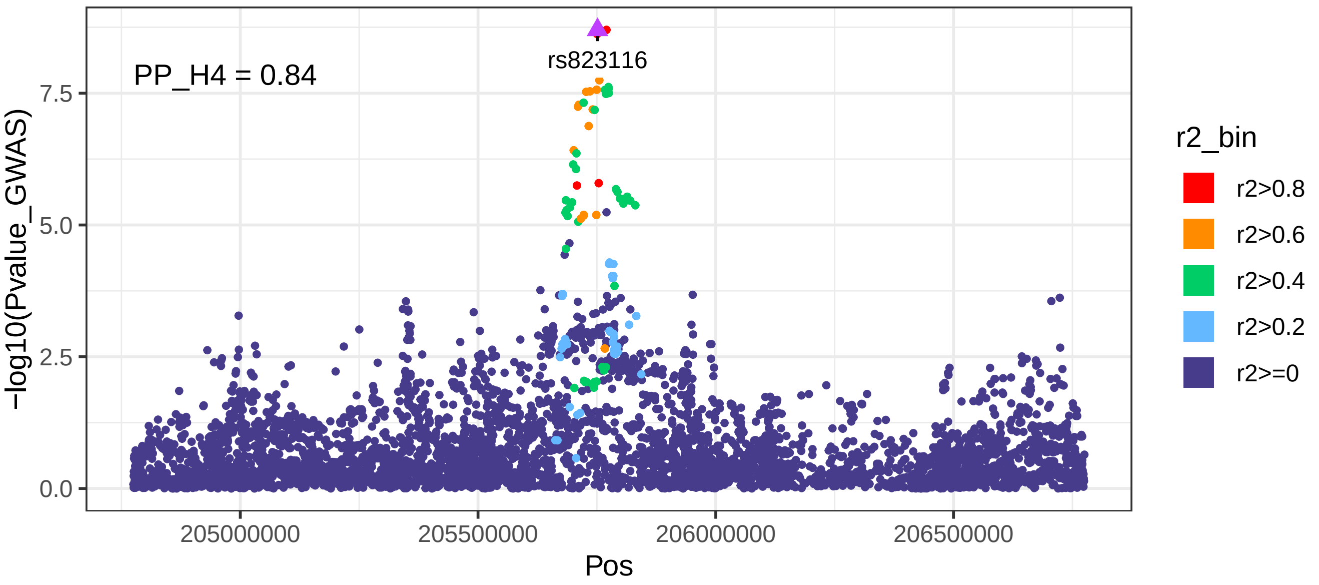


Artery_Tibial: RAB29 eQTL P−values

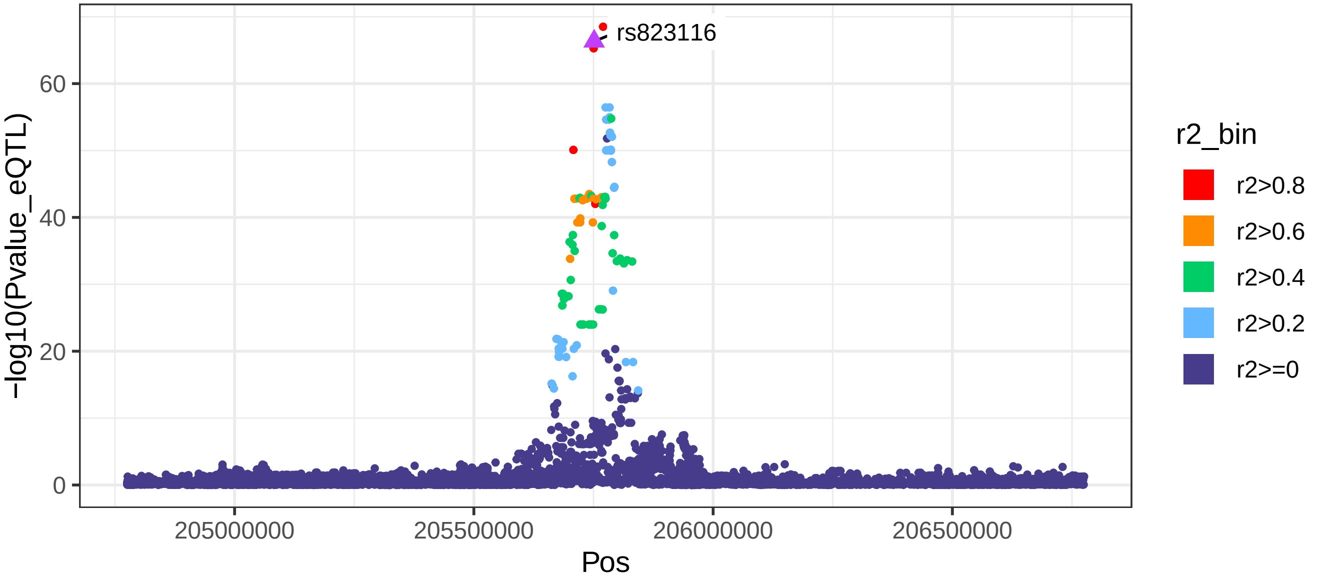

Brain_Cerebellum: RAB29 GWAS P−values

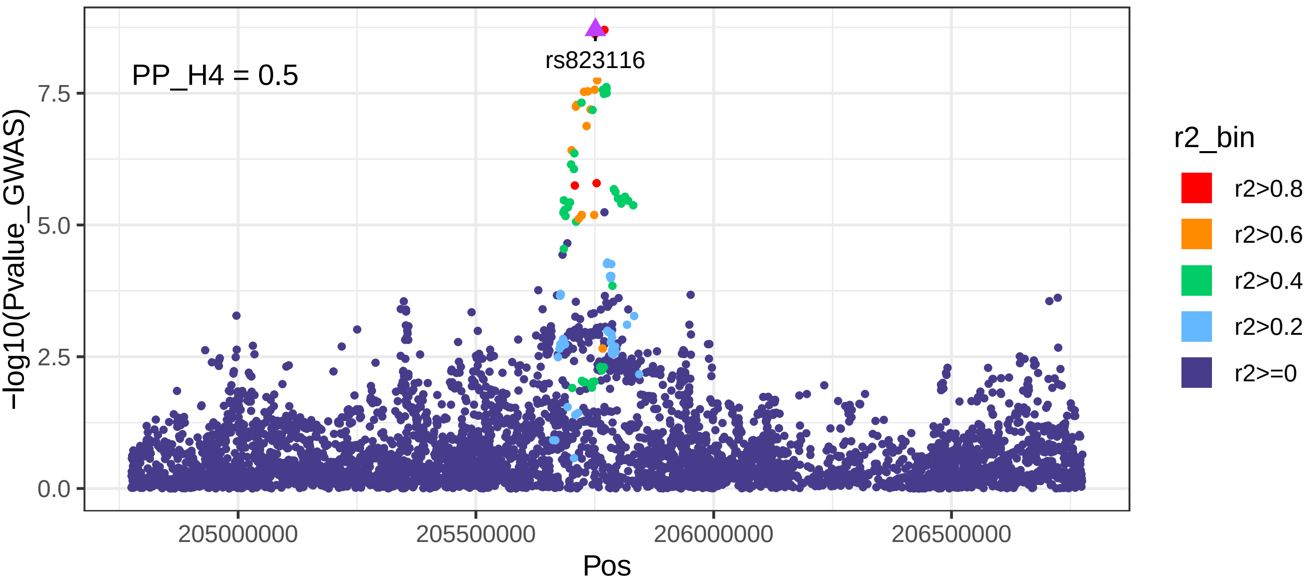


Brain_Cerebellum: RAB29 eQTL P−values

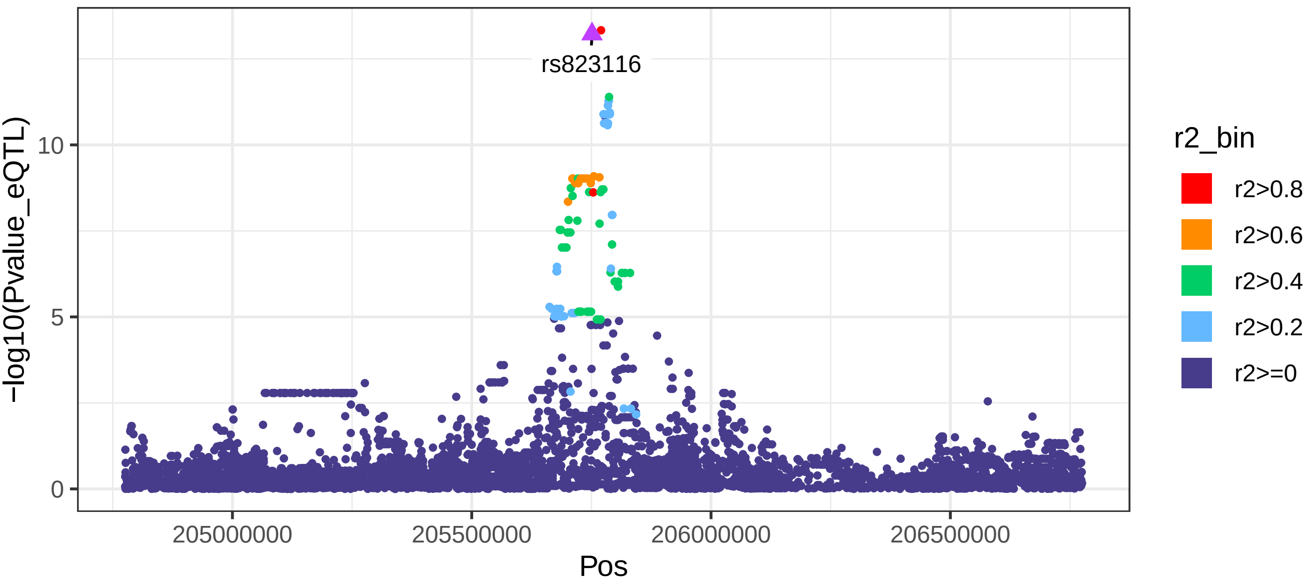

Brain_Cortex: RAB29 GWAS P−values

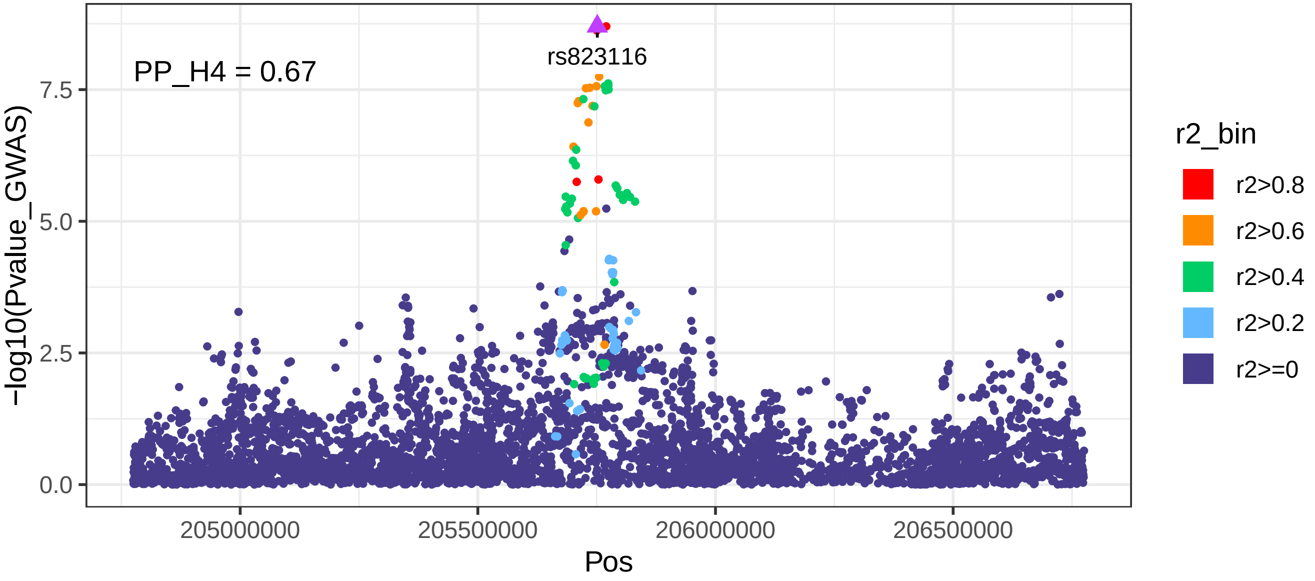


Brain_Cortex: RAB29 eQTL P−values

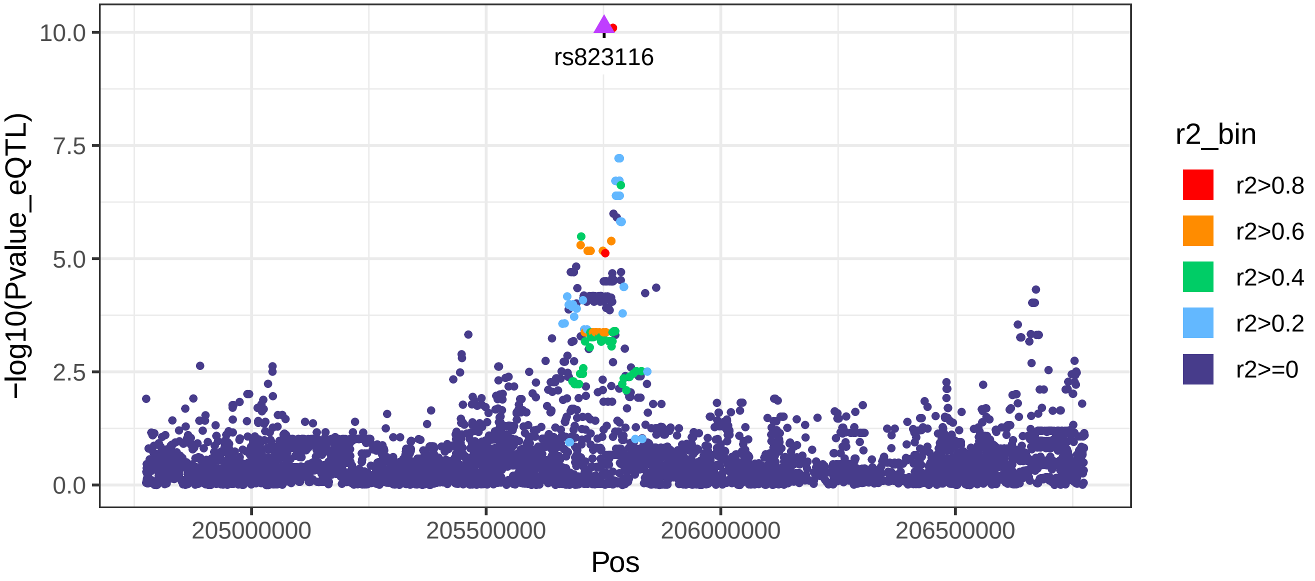

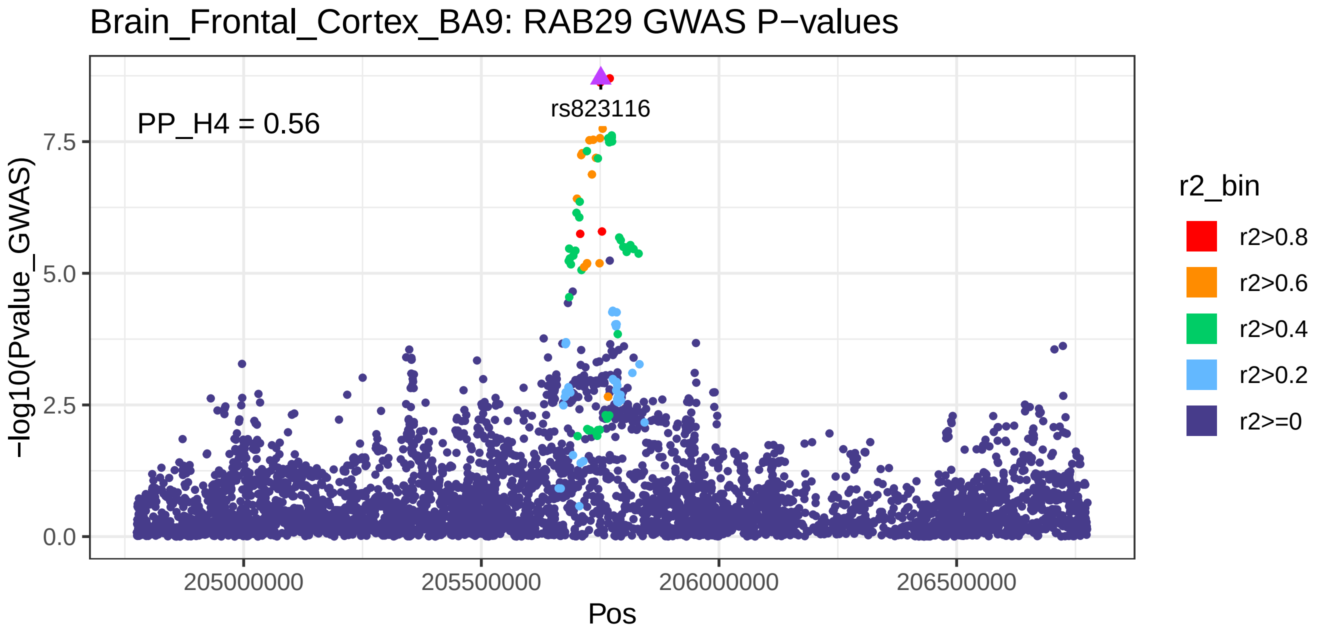
Brain_Frontal_Cortex_BA9: RAB29 GWAS P−values
PP_H4 = 0.56
rs823116
−log10(Pvalue_GWAS)
7.5
5.0
2.5
0.0
205000000
205500000
206000000
206500000
Pos
r2_bin
r2>0.8
r2>0.6
r2>0.4
r2>0.2
r2>=0

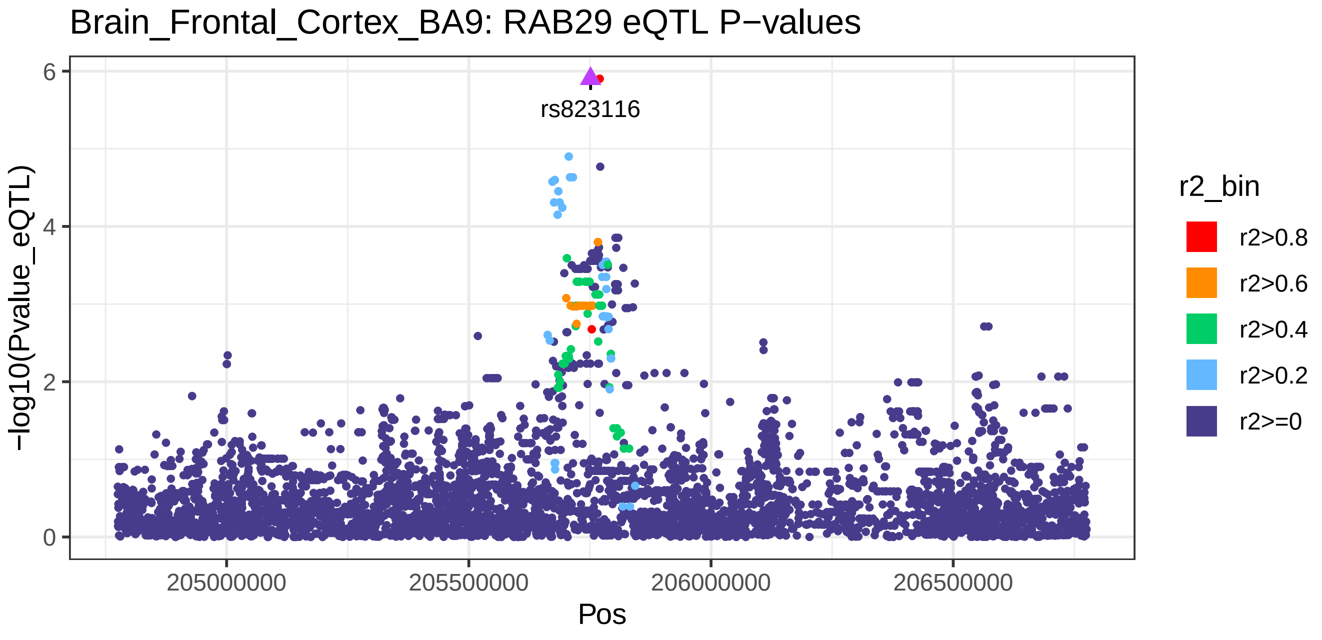
Brain_Frontal_Cortex_BA9: RAB29 eQTL P−values
rs823116
−log10(Pvalue_eQTL)
6
4
2
0
205000000
205500000
206000000
206500000
Pos
r2_bin
r2>0.8
r2>0.6
r2>0.4
r2>0.2
r2>=0

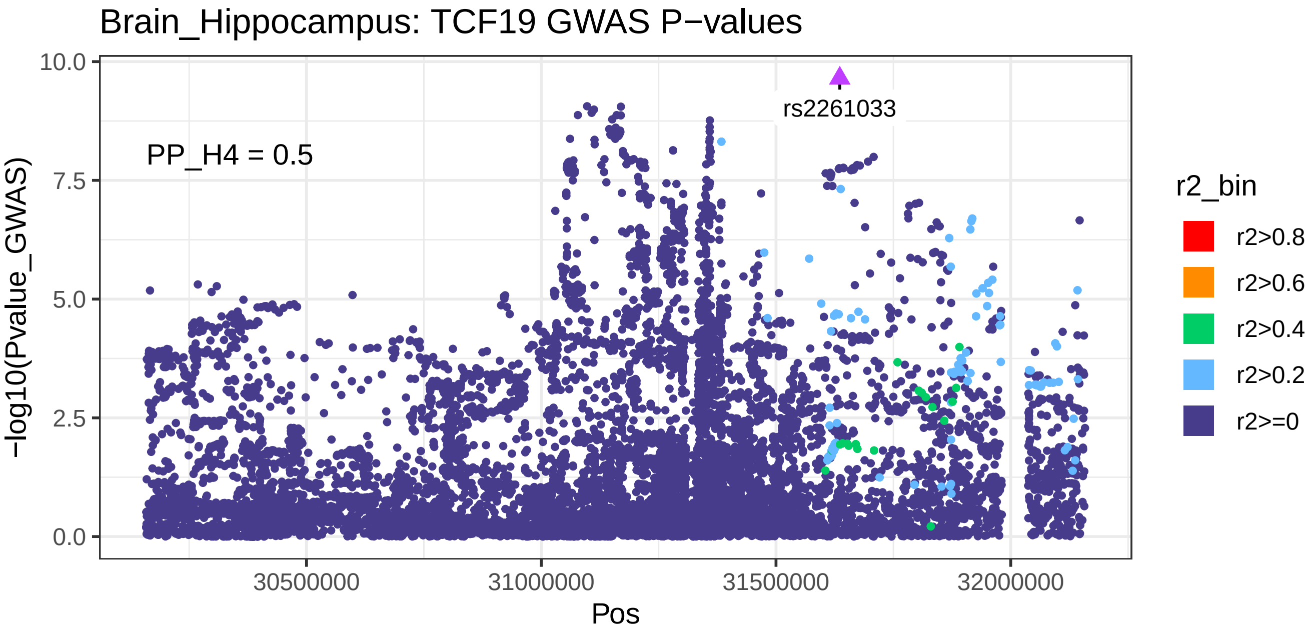

Brain_Hippocampus: TCF19 GWAS P−values
PP_H4 = 0.5
rs2261033
−log10(Pvalue_GWAS)
10.0
7.5
5.0
2.5
0.0
30500000
31000000
31500000
32000000
Pos
r2_bin
r2>0.8
r2>0.6
r2>0.4
r2>0.2
r2>=0


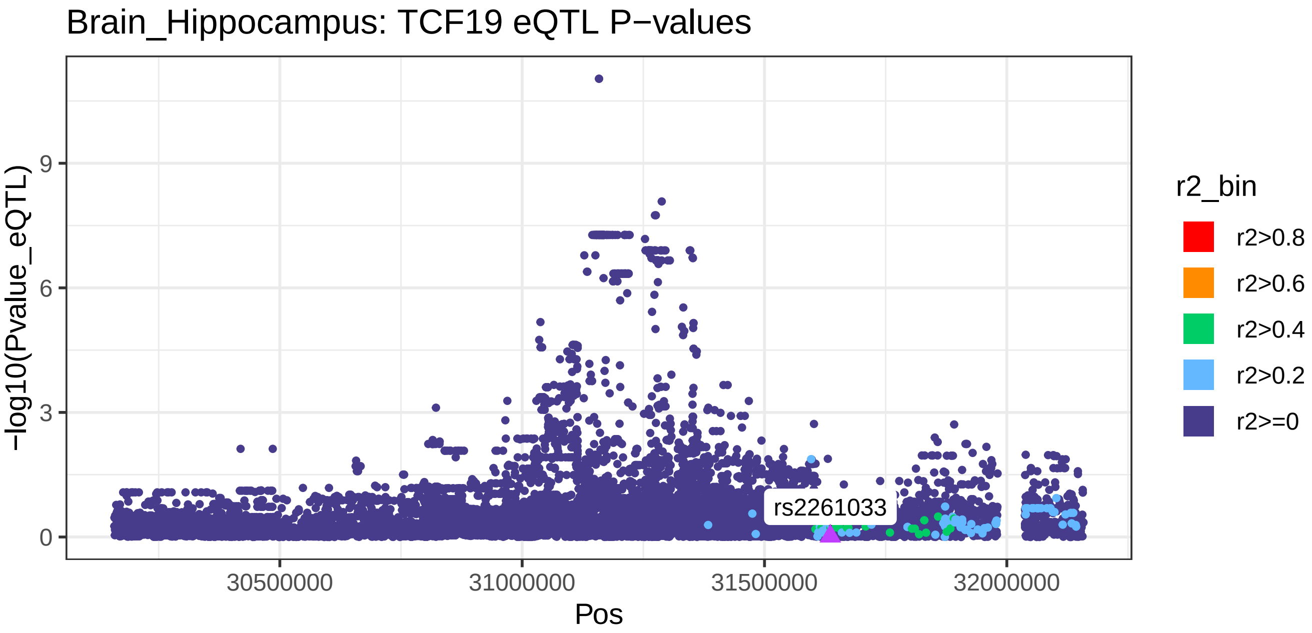

Brain_Hippocampus: TCF19 eQTL P−values
rs2261033
−log10(Pvalue_eQTL)
9
6
3
0
30500000
31000000
31500000
32000000
Pos
r2_bin
r2>0.8
r2>0.6
r2>0.4
r2>0.2
r2>=0

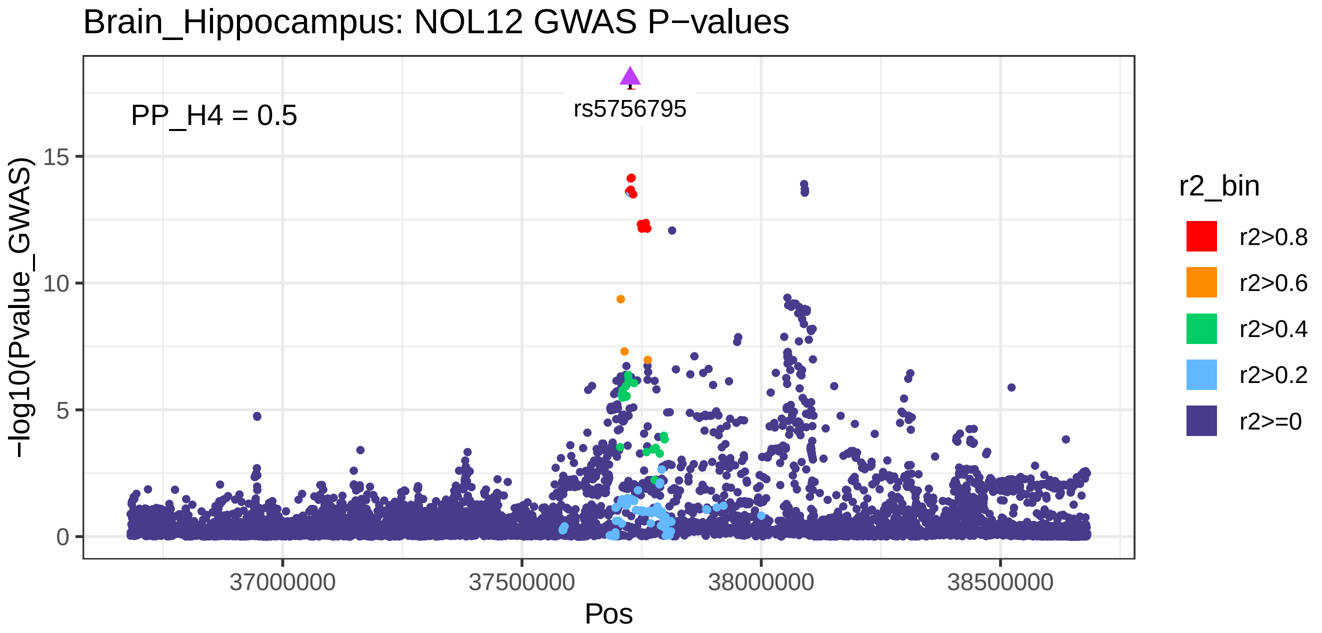
Brain_Hippocampus: NOL12 GWAS P−values
PP_H4 = 0.5
rs5756795
−log10(Pvalue_GWAS)
0
5
10
15
37000000
37500000
38000000
38500000
Pos
r2_bin
r2>0.8
r2>0.6
r2>0.4
r2>0.2
r2>=0

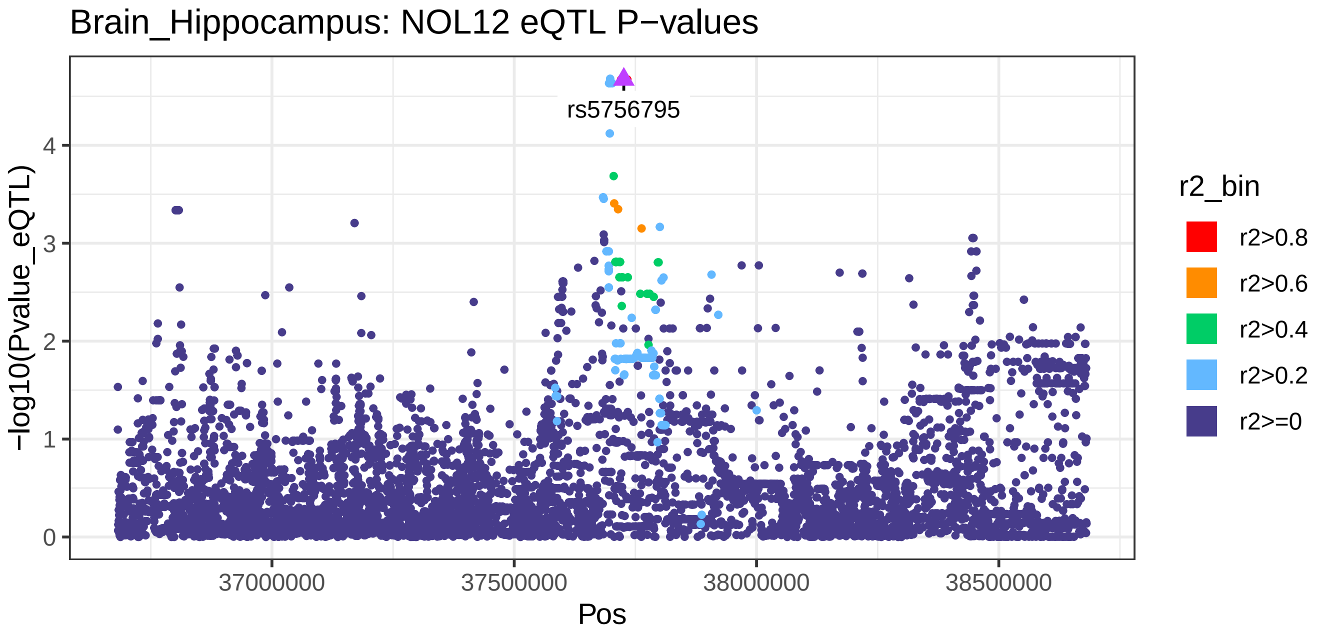
Brain_Hippocampus: NOL12 eQTL P−values
rs5756795
−log10(Pvalue_eQTL)
0
1
2
3
4
37000000
37500000
38000000
38500000
Pos
r2_bin
r2>0.8
r2>0.6
r2>0.4
r2>0.2
r2>=0

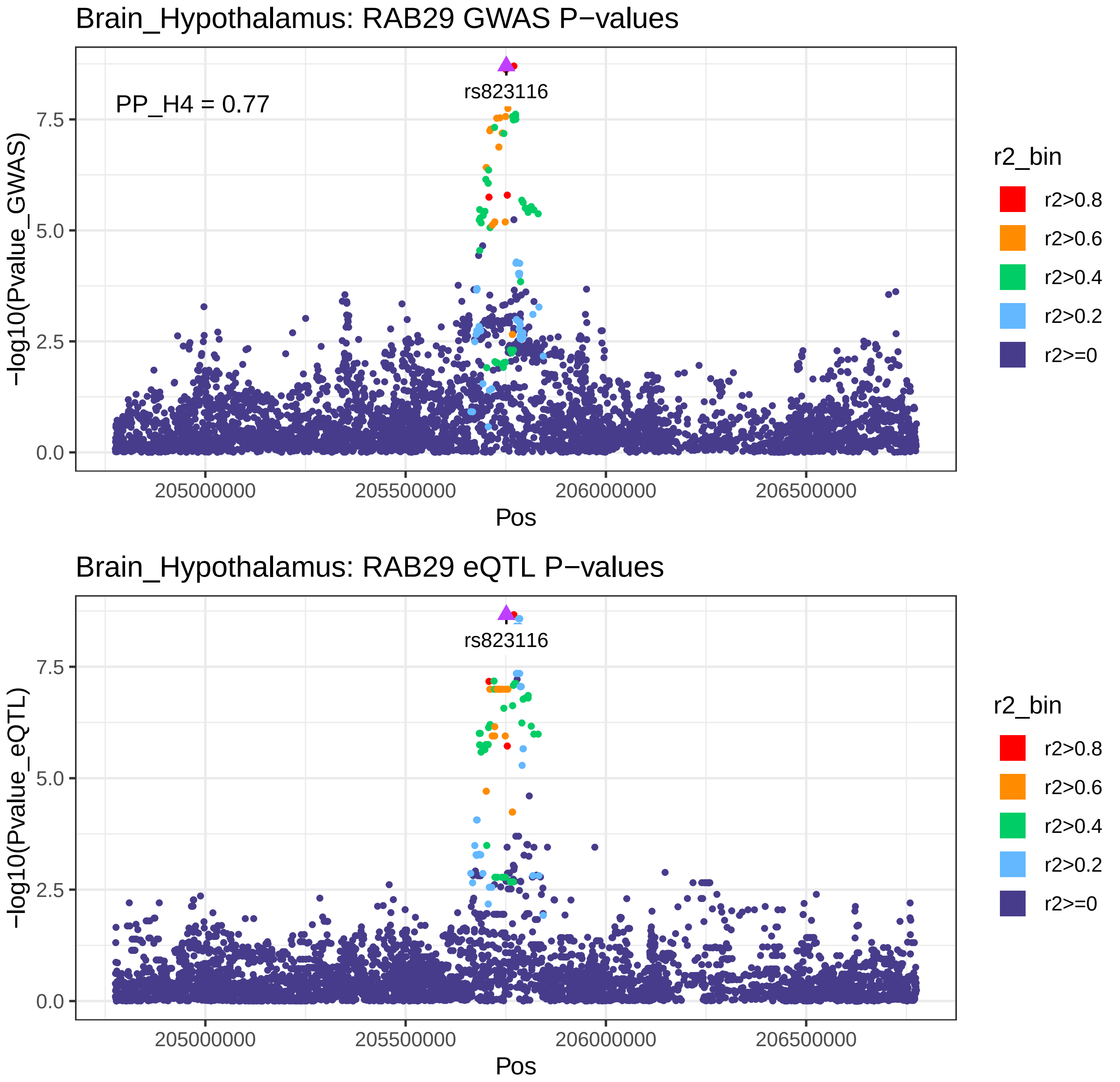
Brain_Hypothalamus: RAB29 GWAS P−values
PP_H4 = 0.77
rs823116
−log10(Pvalue_GWAS)
0.0
2.5
5.0
7.5
205000000
205500000
206000000
206500000
Pos
r2_bin
r2>0.8
r2>0.6
r2>0.4
r2>0.2
r2>=0
Brain_Hypothalamus: RAB29 eQTL P−values
rs823116
−log10(Pvalue_eQTL)
0.0
2.5
5.0
7.5
205000000
205500000
206000000
206500000
Pos
r2_bin
r2>0.8
r2>0.6
r2>0.4
r2>0.2
r2>=0

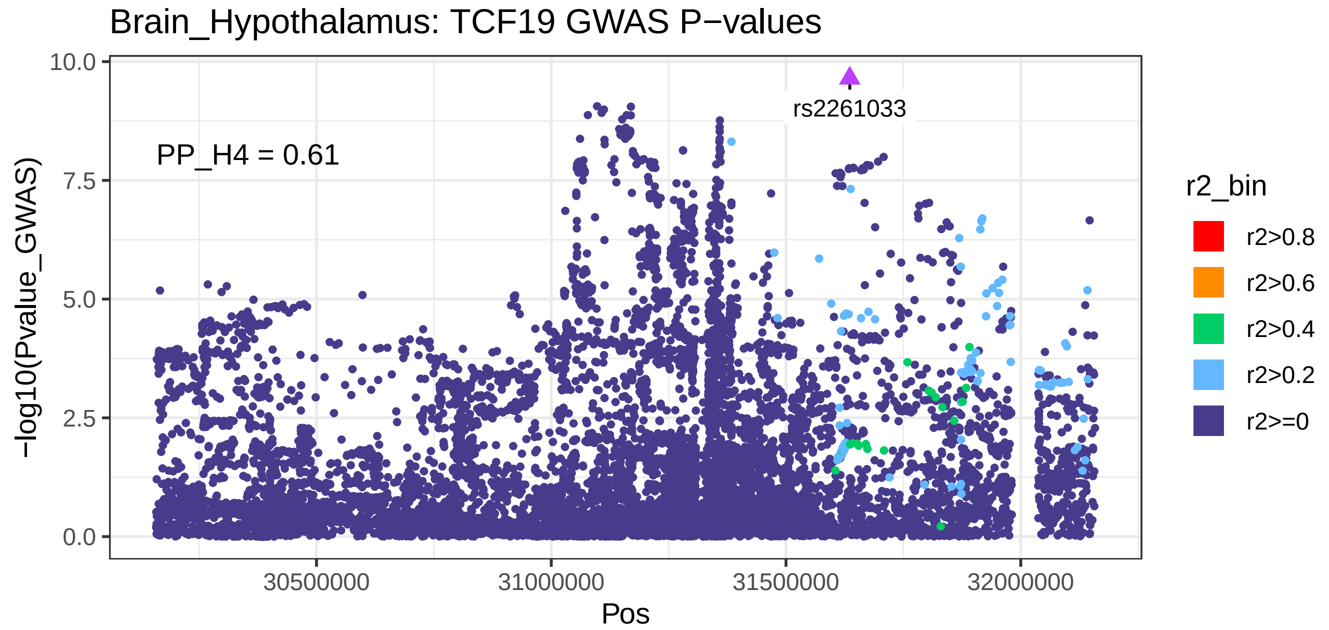
Brain_Hypothalamus: TCF19 GWAS P−values
PP_H4 = 0.61
rs2261033
−log10(Pvalue_GWAS)
10.0
7.5
5.0
2.5
0.0
30500000
31000000
31500000
32000000
Pos
r2_bin
r2>0.8
r2>0.6
r2>0.4
r2>0.2
r2>=0

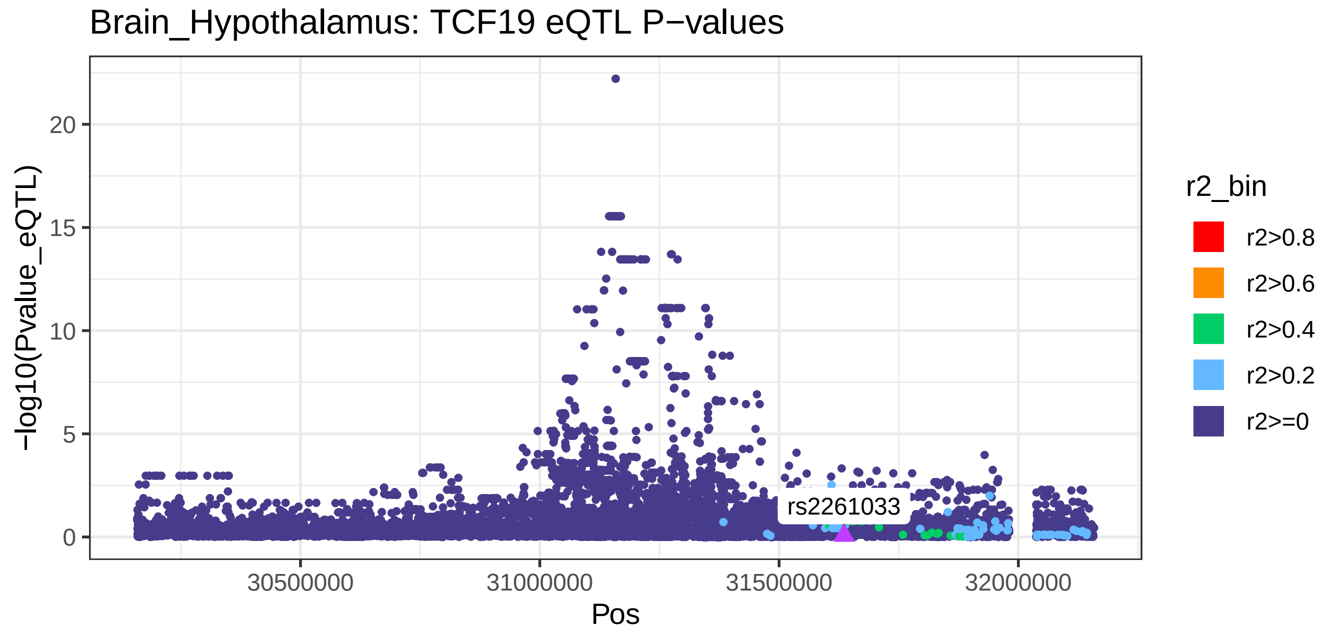
Brain_Hypothalamus: TCF19 eQTL P−values
rs2261033
−log10(Pvalue_eQTL)
20
15
10
5
0
30500000
31000000
31500000
32000000
Pos
r2_bin
r2>0.8
r2>0.6
r2>0.4
r2>0.2
r2>=0

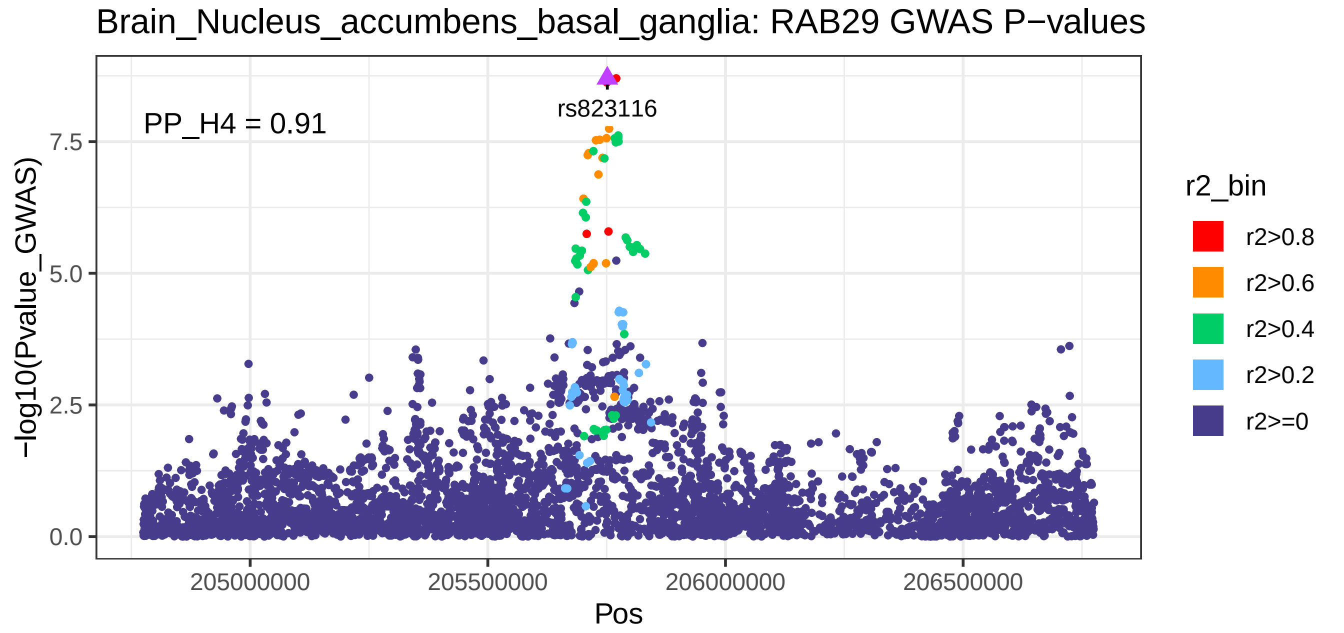

Brain_Nucleus_accumbens_basal_ganglia: RAB29 GWAS P−values
PP_H4 = 0.91
rs823116
−log10(Pvalue_GWAS)
0.0
2.5
5.0
7.5
205000000
205500000
206000000
206500000
Pos
r2_bin
r2>0.8
r2>0.6
r2>0.4
r2>0.2
r2>=0


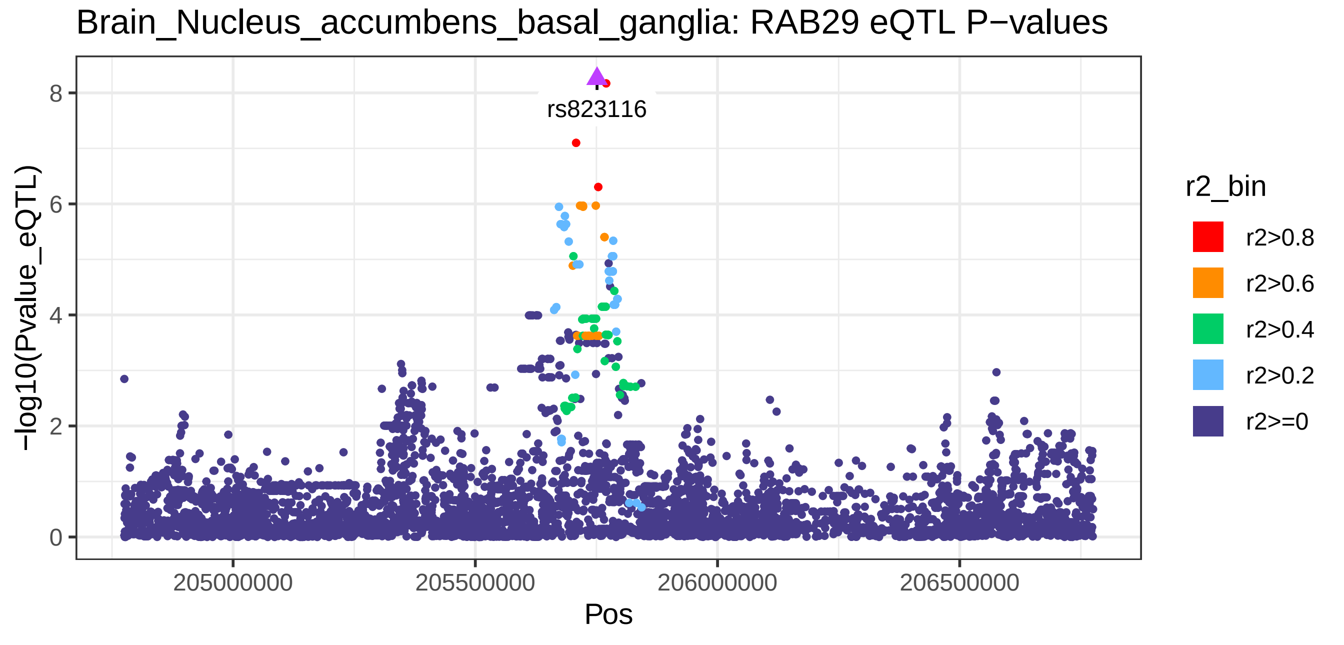

Brain_Nucleus_accumbens_basal_ganglia: RAB29 eQTL P−values
rs823116
−log10(Pvalue_eQTL)
0
2
4
6
8
205000000
205500000
206000000
206500000
Pos
r2_bin
r2>0.8
r2>0.6
r2>0.4
r2>0.2
r2>=0

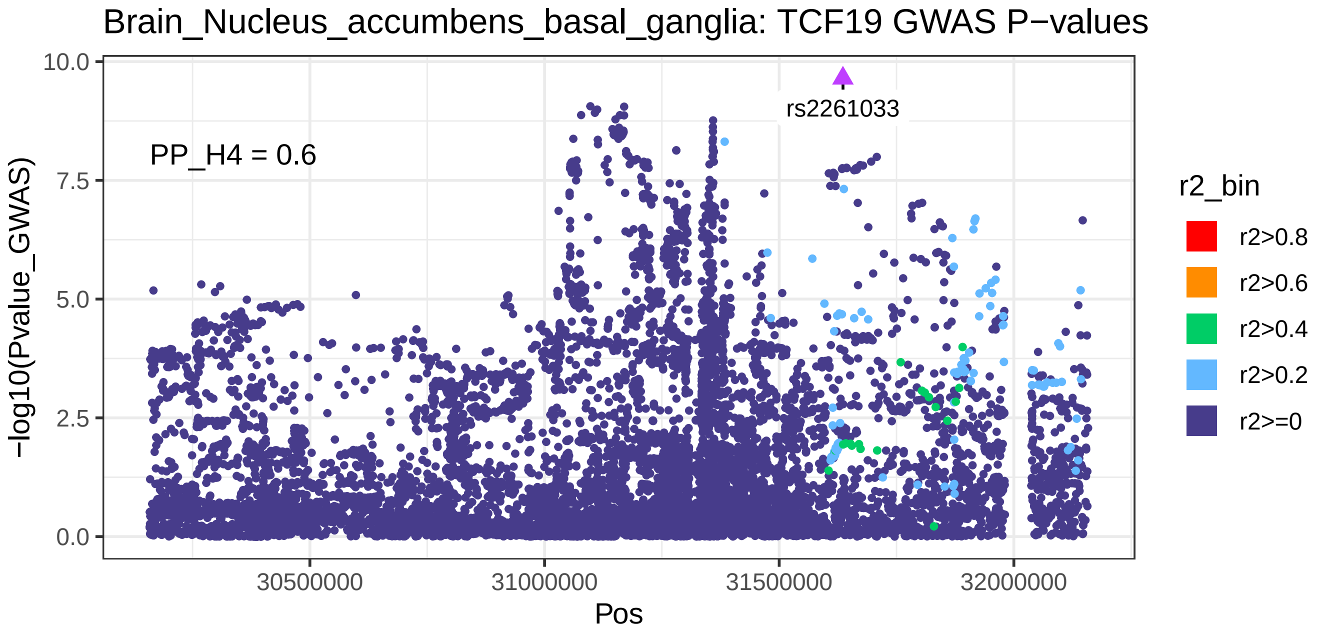
Brain_Nucleus_accumbens_basal_ganglia: TCF19 GWAS P−values
PP_H4 = 0.6
rs2261033
−log10(Pvalue_GWAS)
Pos
r2_bin
r2>0.8
r2>0.6
r2>0.4
r2>0.2
r2>=0

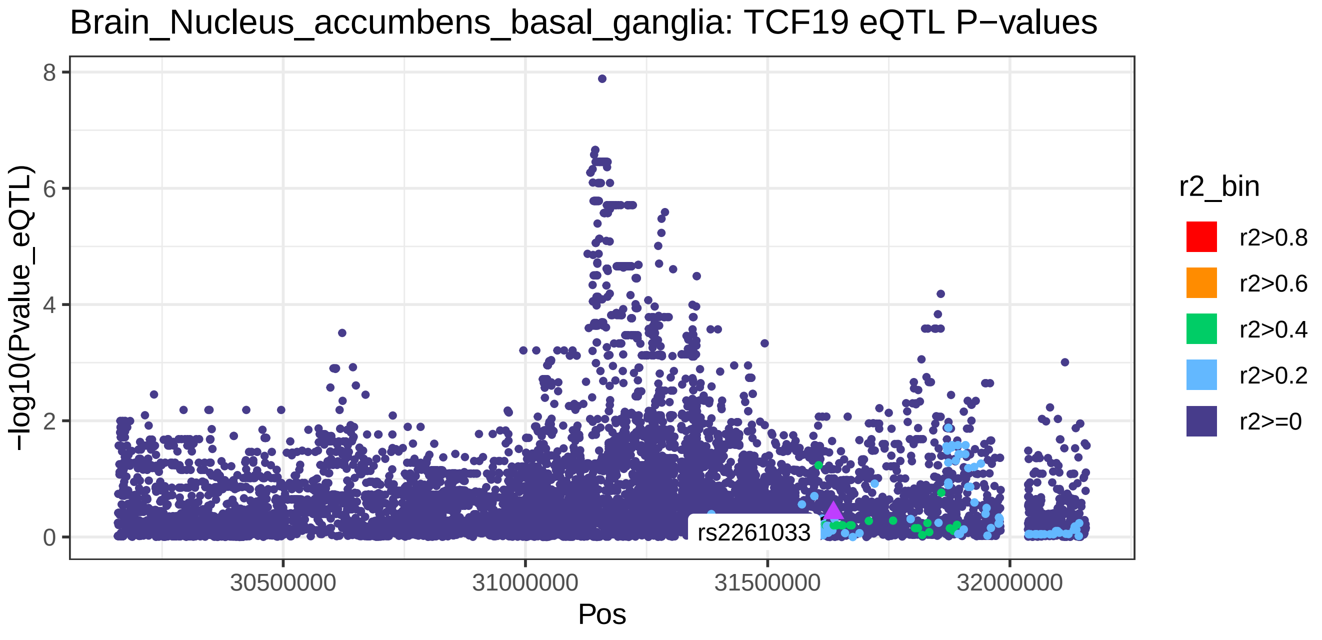
Brain_Nucleus_accumbens_basal_ganglia: TCF19 eQTL P−values
rs2261033
−log10(Pvalue_eQTL)
Pos
r2_bin
r2>0.8
r2>0.6
r2>0.4
r2>0.2
r2>=0

Brain_Nucleus_accumbens_basal_ganglia: CLDN7 GWAS P−values

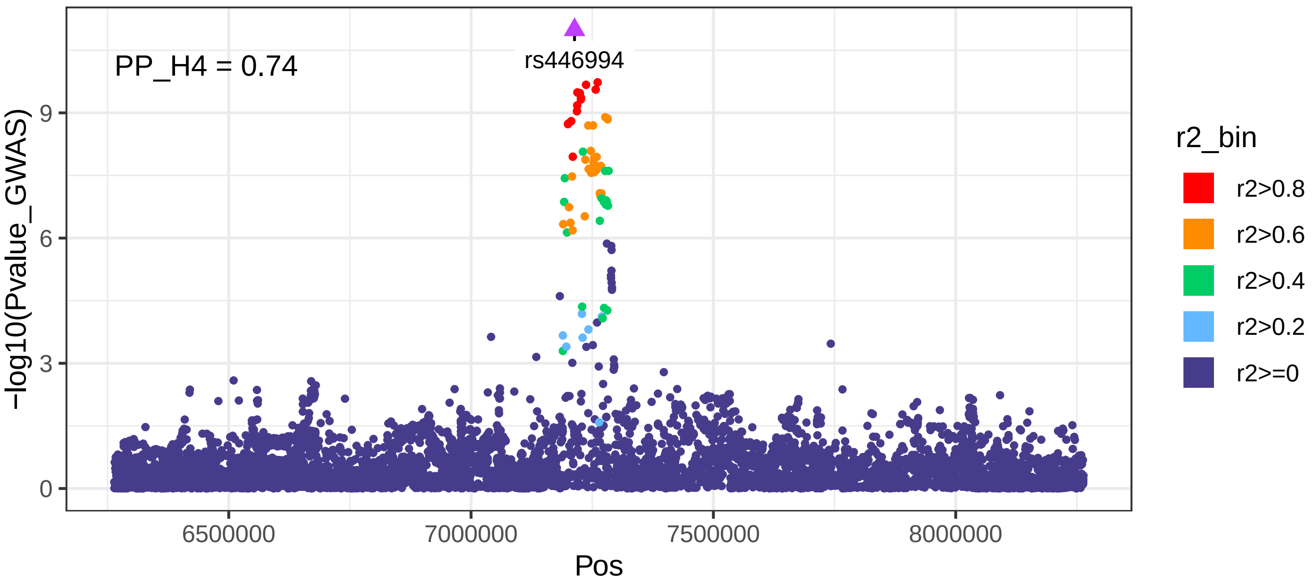


Brain_Nucleus_accumbens_basal_ganglia: CLDN7 eQTL P−values

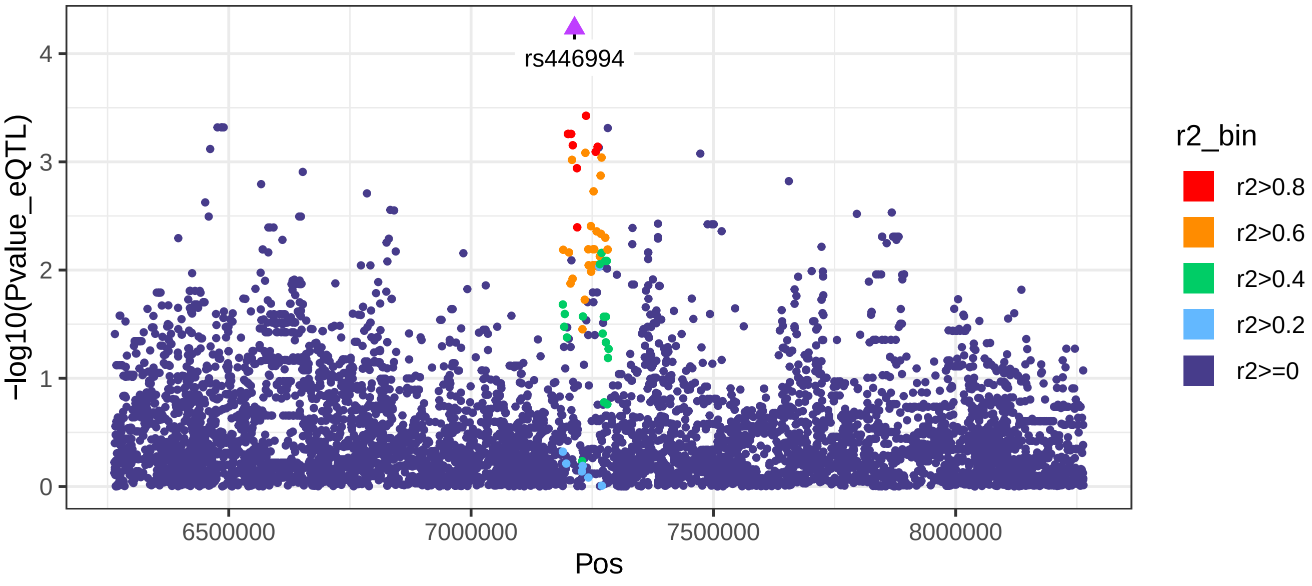

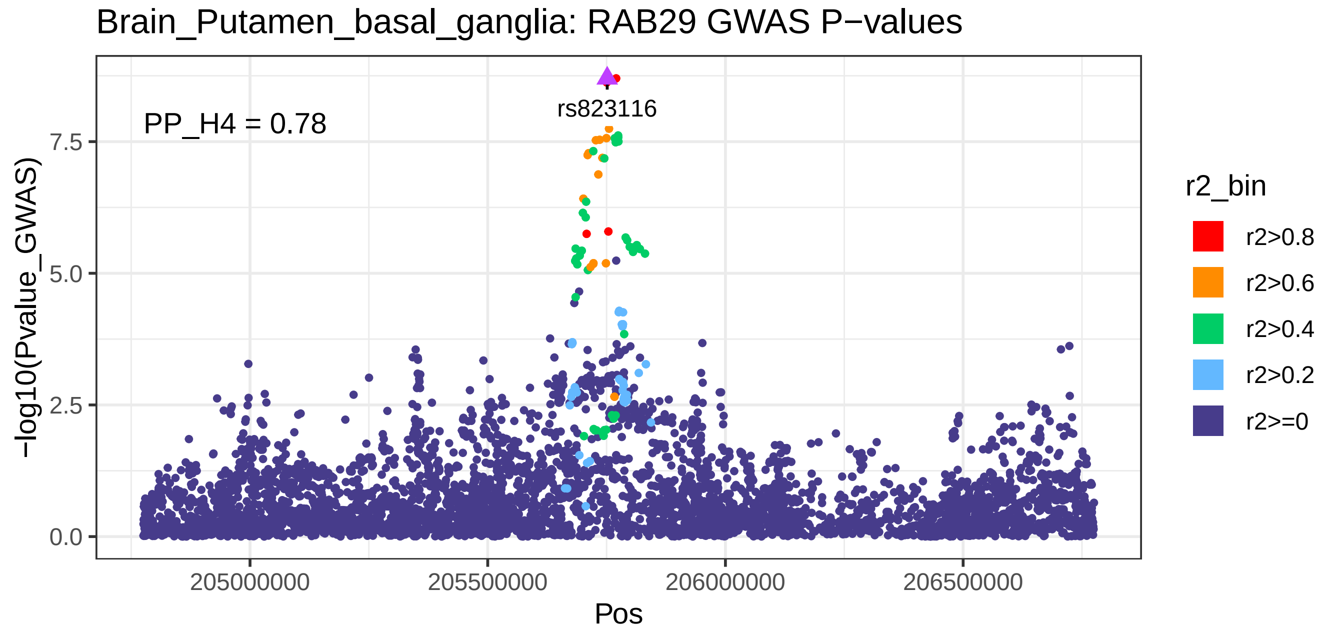

Brain_Putamen_basal_ganglia: RAB29 GWAS P−values
PP_H4 = 0.78
rs823116
−log10(Pvalue_GWAS)
0.0
2.5
5.0
7.5
205000000
205500000
206000000
206500000
Pos
r2_bin
r2>0.8
r2>0.6
r2>0.4
r2>0.2
r2>=0


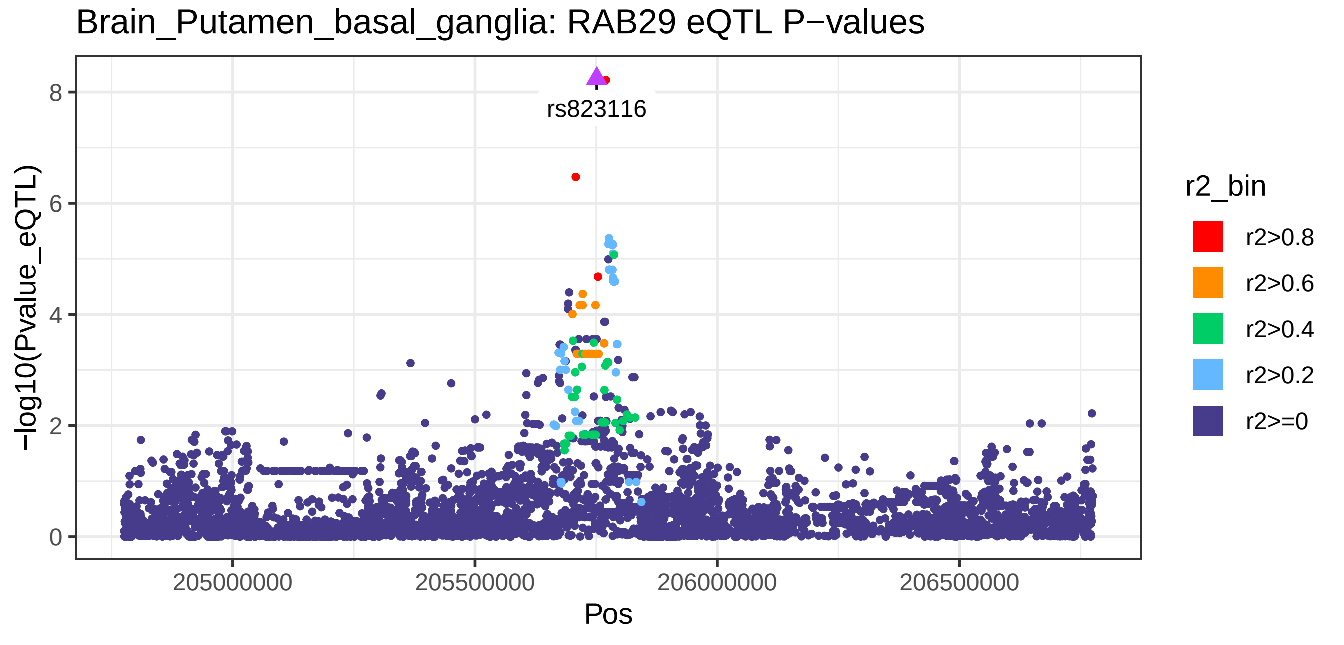

Brain_Putamen_basal_ganglia: RAB29 eQTL P−values
rs823116
−log10(Pvalue_eQTL)
0
2
4
6
8
205000000
205500000
206000000
206500000
Pos
r2_bin
r2>0.8
r2>0.6
r2>0.4
r2>0.2
r2>=0

Brain_Spinal_cord_cervical_c−1: RAB29 GWAS P−values

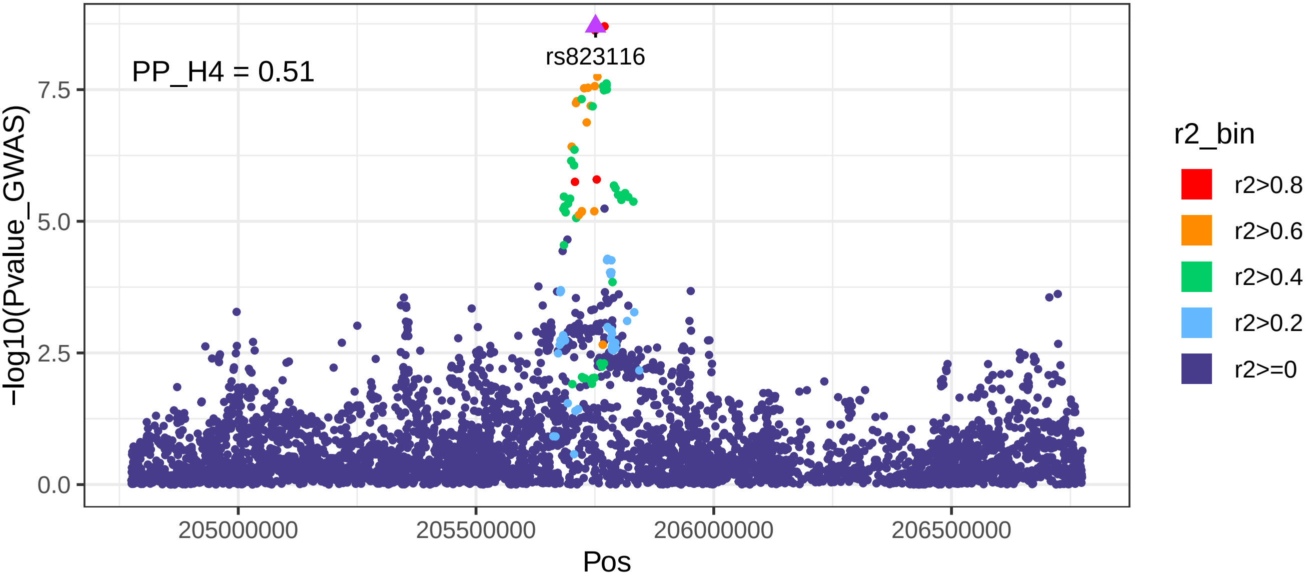


Brain_Spinal_cord_cervical_c−1: RAB29 eQTL P−values

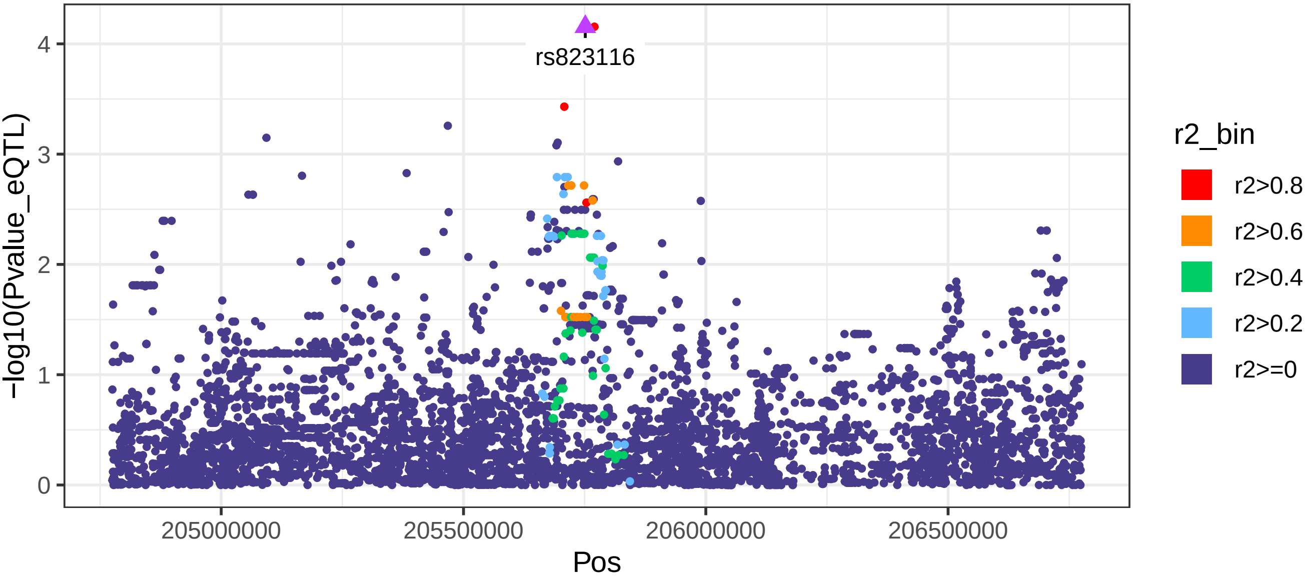

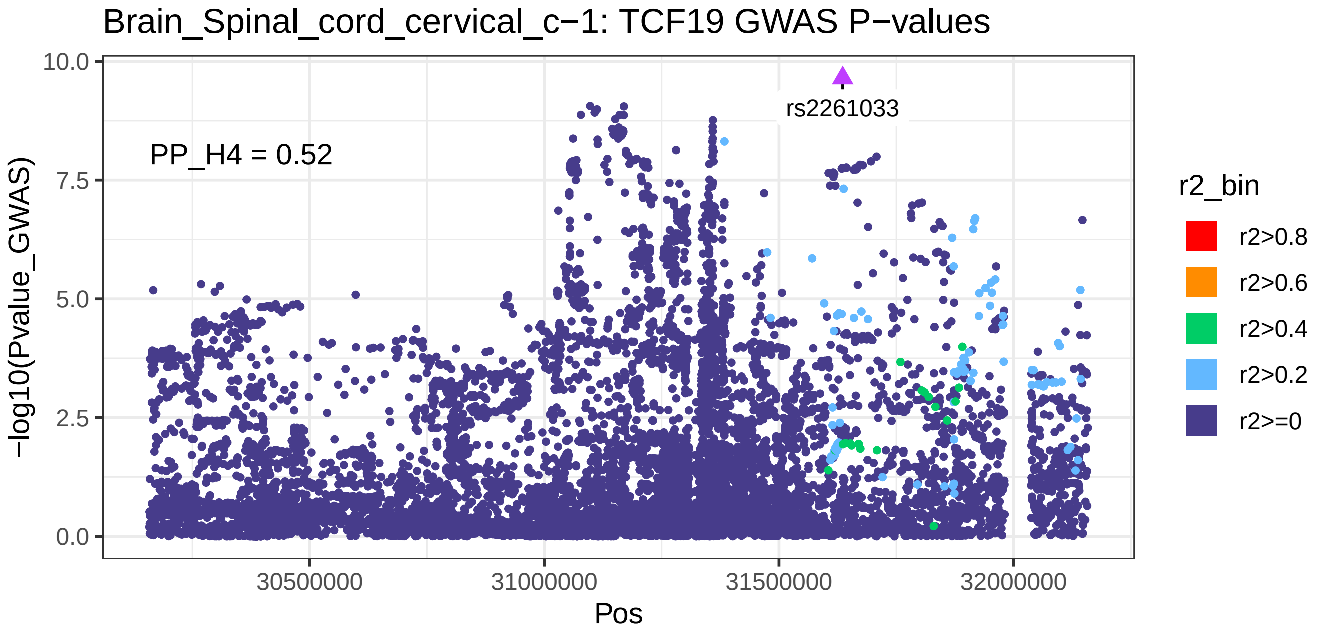

Brain_Spinal_cord_cervical_c−1: TCF19 GWAS P−values
PP_H4 = 0.52
rs2261033
−log10(Pvalue_GWAS)
10.0
7.5
5.0
2.5
0.0
30500000
31000000
31500000
32000000
Pos
r2_bin
r2>0.8
r2>0.6
r2>0.4
r2>0.2
r2>=0


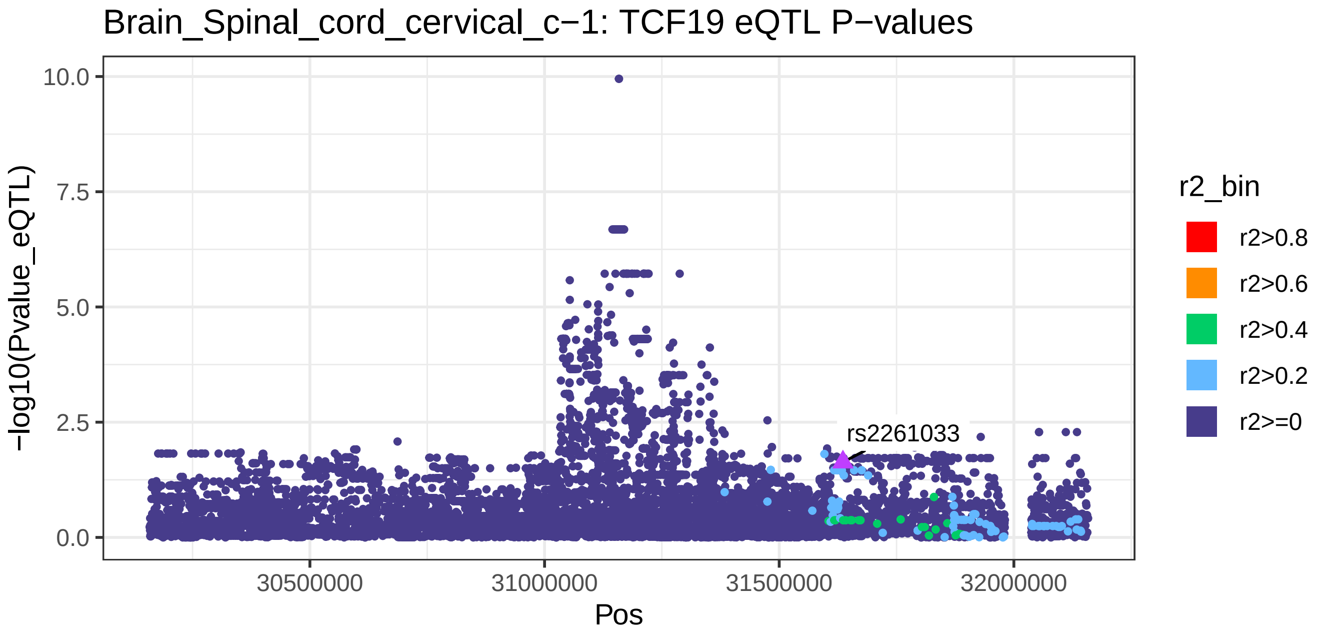

Brain_Spinal_cord_cervical_c−1: TCF19 eQTL P−values
rs2261033
−log10(Pvalue_eQTL)
10.0
7.5
5.0
2.5
0.0
30500000
31000000
31500000
32000000
Pos
r2_bin
r2>0.8
r2>0.6
r2>0.4
r2>0.2
r2>=0

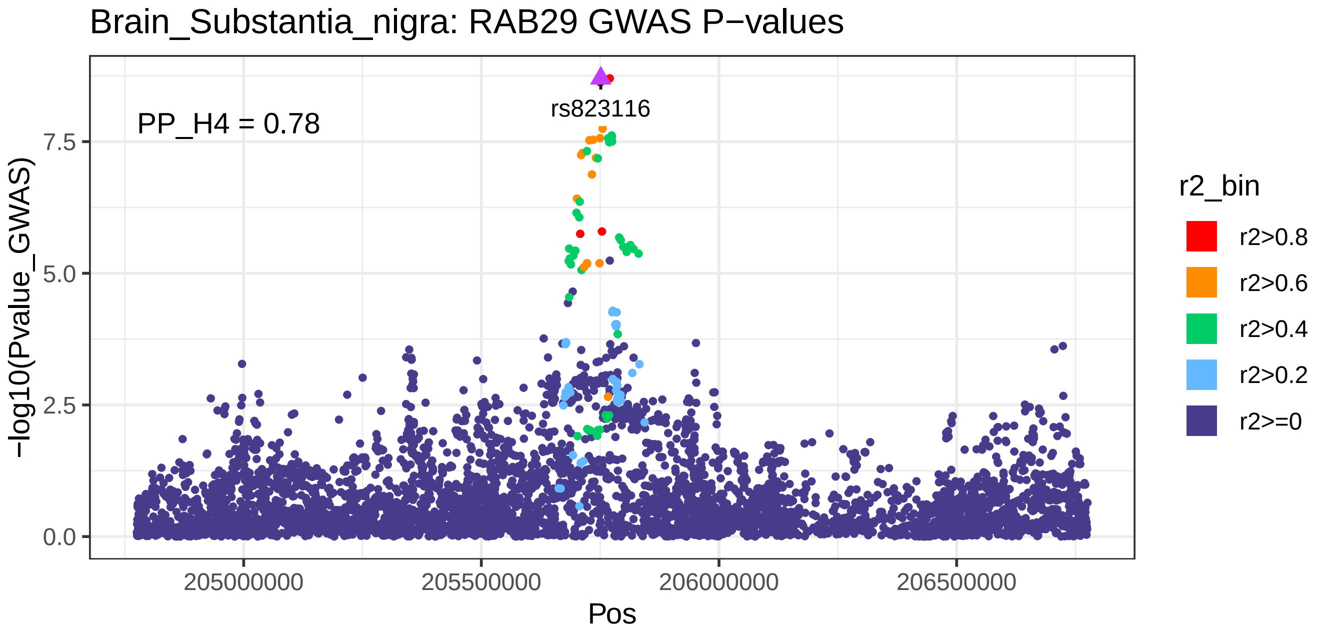
Brain_Substantia_nigra: RAB29 GWAS P−values
PP_H4 = 0.78
rs823116
−log10(Pvalue_GWAS)
0.0
2.5
5.0
7.5
205000000
205500000
206000000
206500000
Pos
r2_bin
r2>0.8
r2>0.6
r2>0.4
r2>0.2
r2>=0

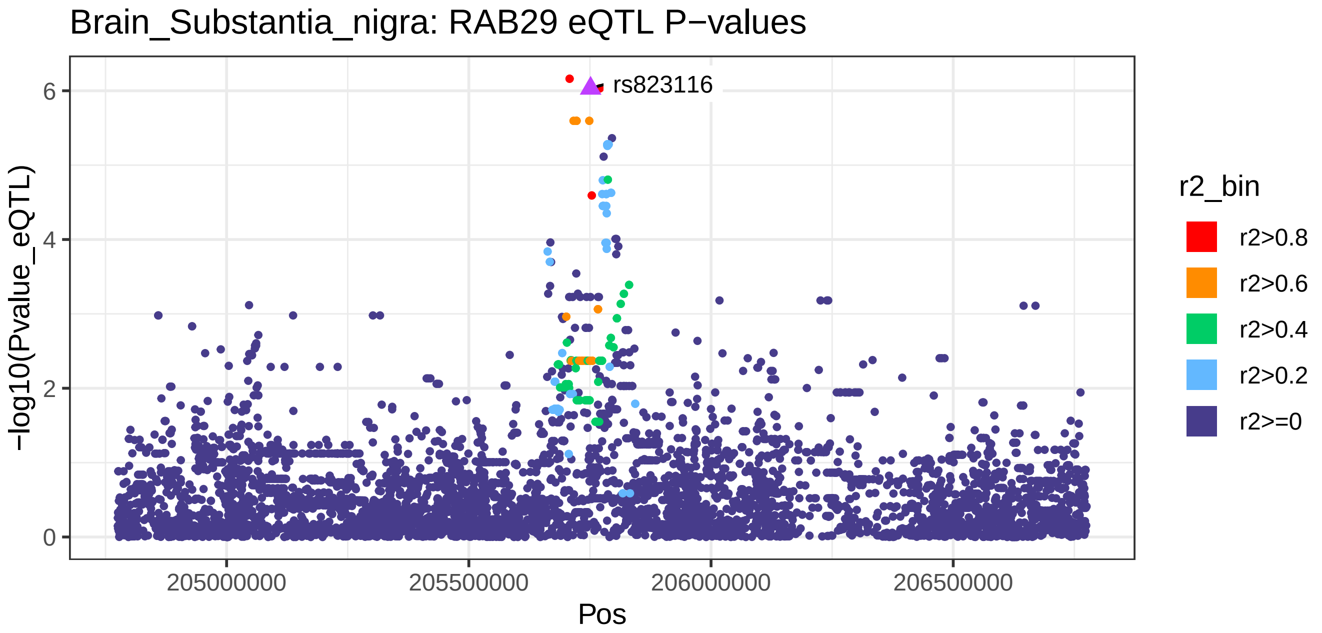
Brain_Substantia_nigra: RAB29 eQTL P−values
rs823116
−log10(Pvalue_eQTL)
0
2
4
6
205000000
205500000
206000000
206500000
Pos
r2_bin
r2>0.8
r2>0.6
r2>0.4
r2>0.2
r2>=0

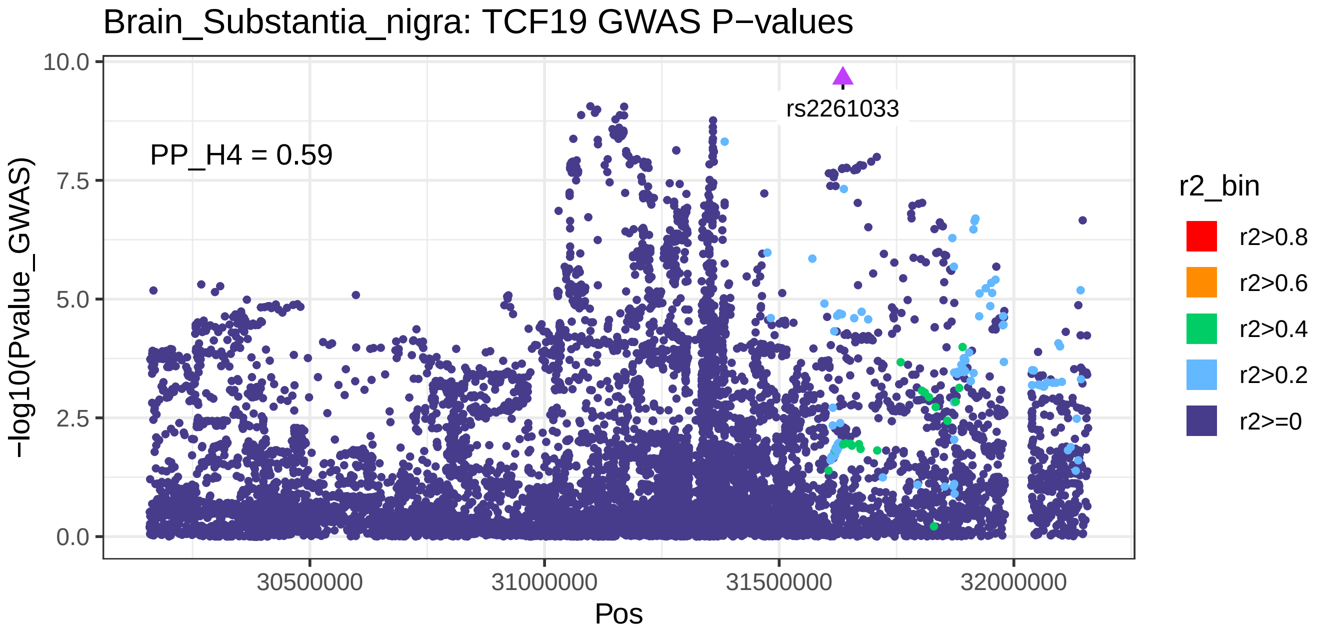
Brain_Substantia_nigra: TCF19 GWAS P−values
PP_H4 = 0.59
rs2261033
−log10(Pvalue_GWAS)
10.0
7.5
5.0
2.5
0.0
30500000
31000000
31500000
32000000
Pos
r2_bin
r2>0.8
r2>0.6
r2>0.4
r2>0.2
r2>=0

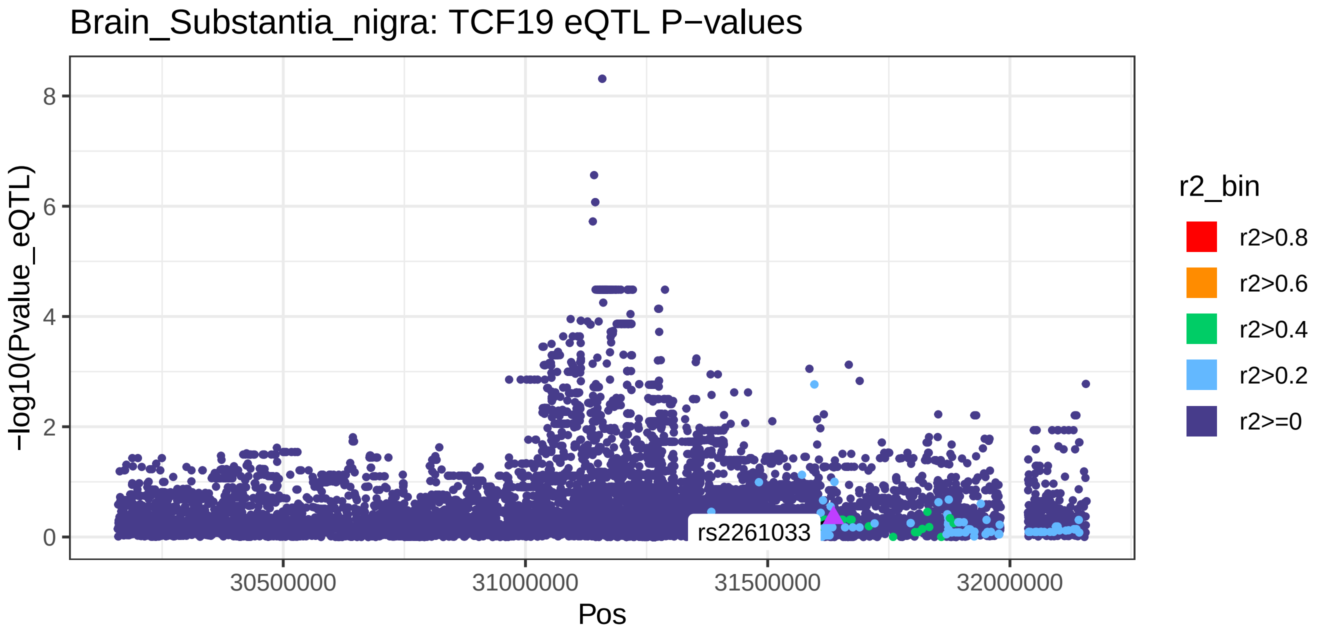
Brain_Substantia_nigra: TCF19 eQTL P−values
rs2261033
−log10(Pvalue_eQTL)
8
6
4
2
0
30500000
31000000
31500000
32000000
Pos
r2_bin
r2>0.8
r2>0.6
r2>0.4
r2>0.2
r2>=0

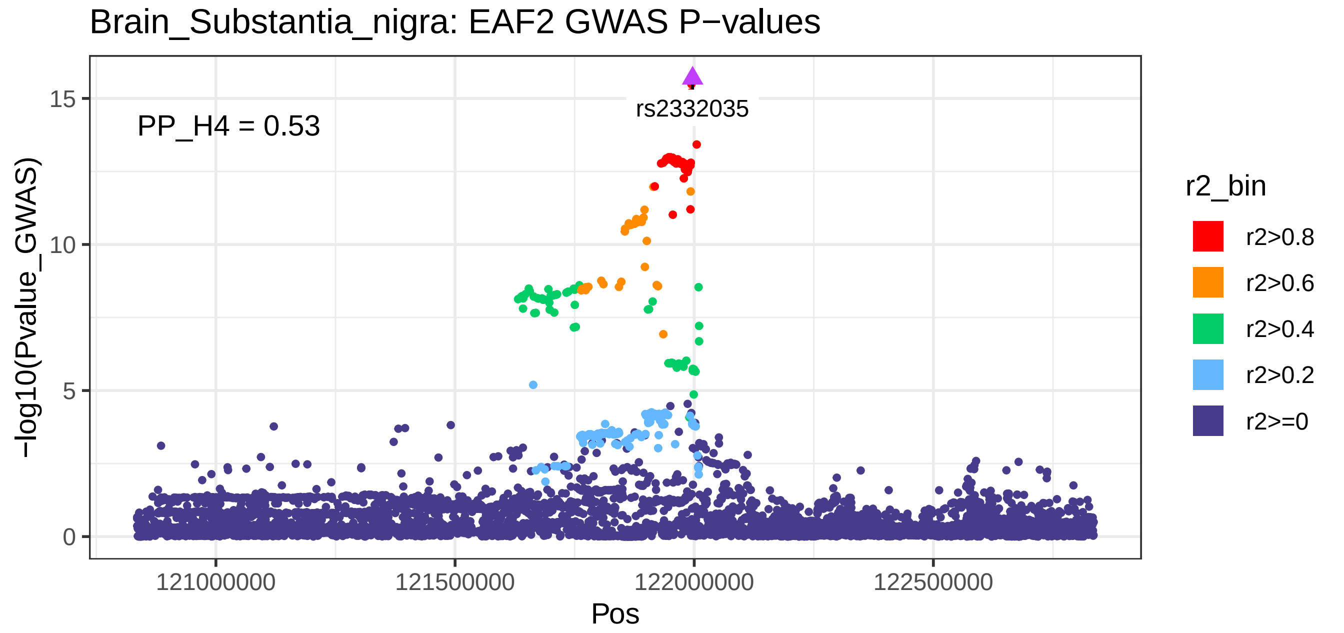
Brain_Substantia_nigra: EAF2 GWAS P−values
PP_H4 = 0.53
rs2332035
−log10(Pvalue_GWAS)
15
10
5
0
121000000
121500000
122000000
122500000
Pos
r2_bin
r2>0.8
r2>0.6
r2>0.4
r2>0.2
r2>=0

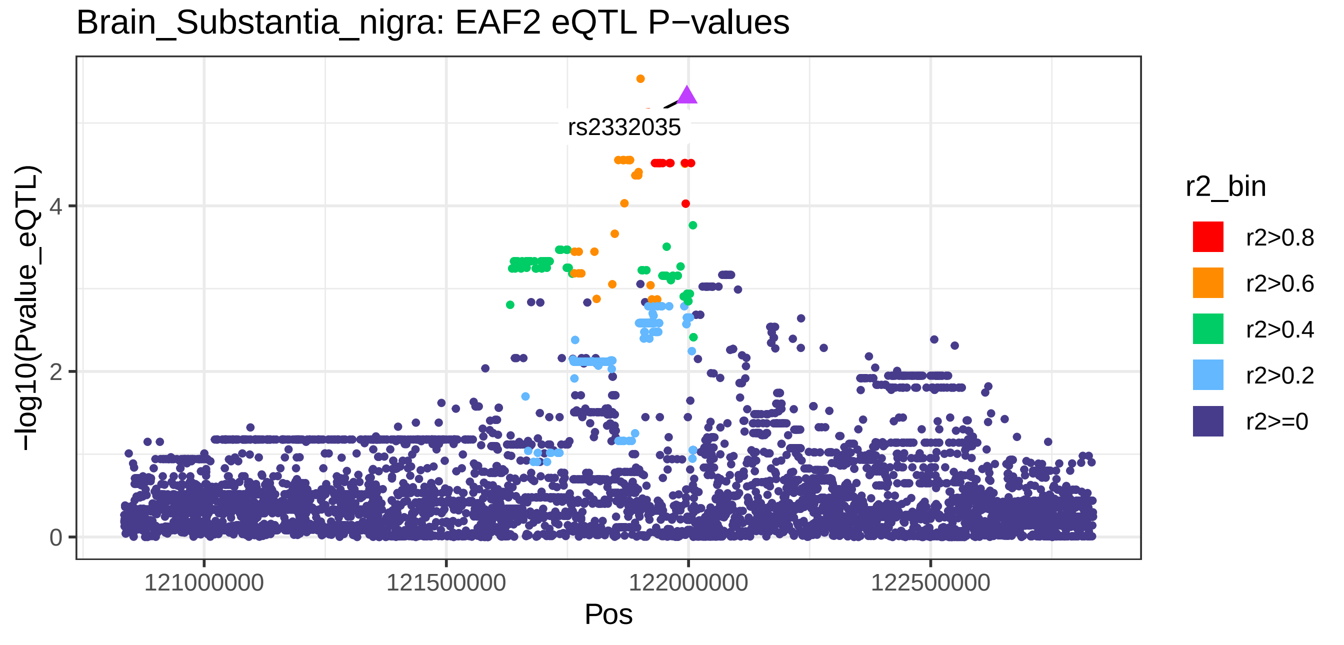
Brain_Substantia_nigra: EAF2 eQTL P−values
rs2332035
−log10(Pvalue_eQTL)
4
2
0
121000000
121500000
122000000
122500000
Pos
r2_bin
r2>0.8
r2>0.6
r2>0.4
r2>0.2
r2>=0

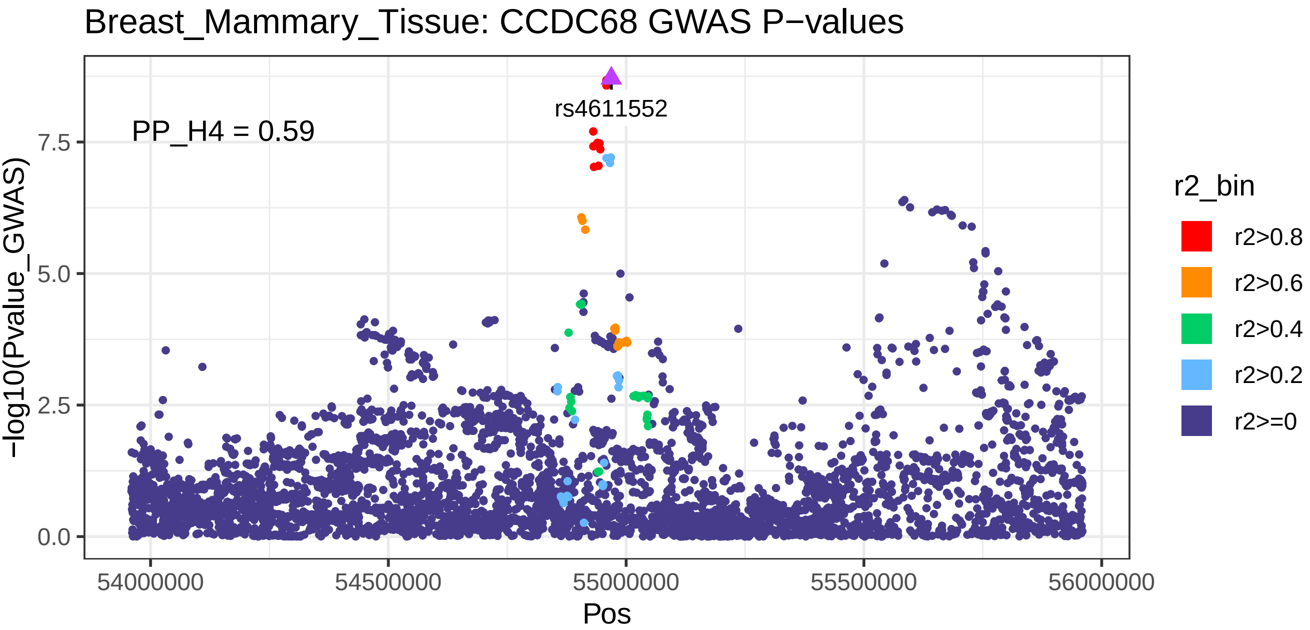
Breast_Mammary_Tissue: CCDC68 GWAS P−values
PP_H4 = 0.59
rs4611552
−log10(Pvalue_GWAS)
0.0
2.5
5.0
7.5
54000000
54500000
55000000
55500000
56000000
Pos
r2_bin
r2>0.8
r2>0.6
r2>0.4
r2>0.2
r2>=0

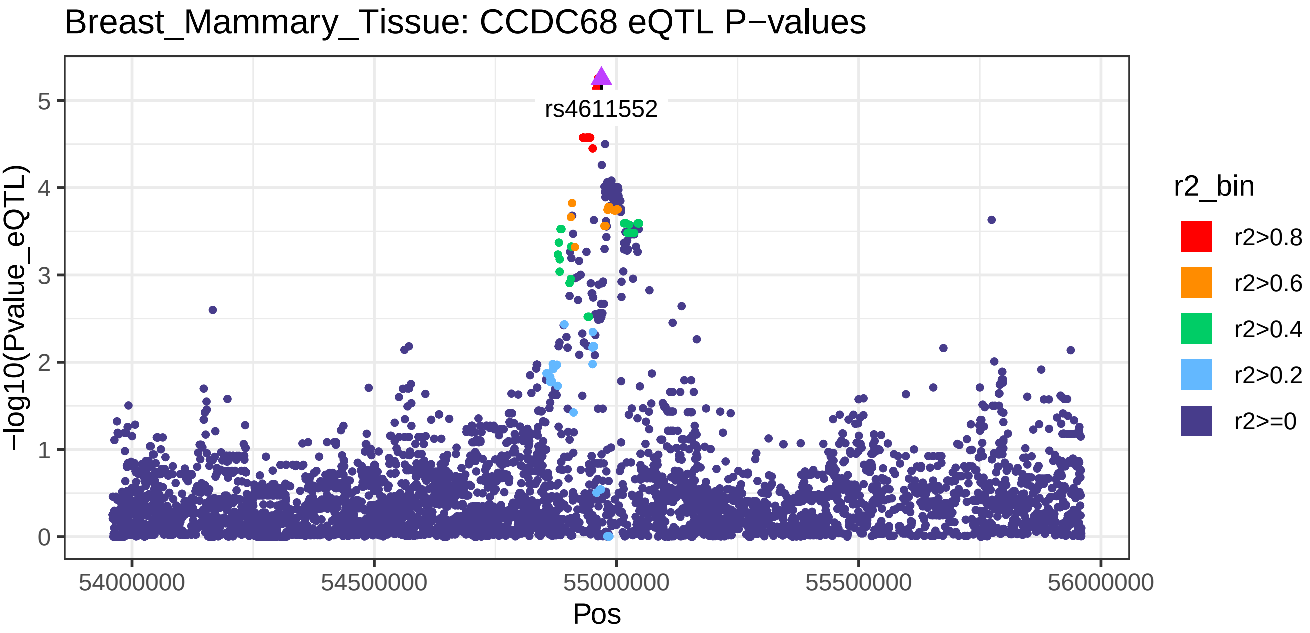
Breast_Mammary_Tissue: CCDC68 eQTL P−values
rs4611552
−log10(Pvalue_eQTL)
0
1
2
3
4
5
54000000
54500000
55000000
55500000
56000000
Pos
r2_bin
r2>0.8
r2>0.6
r2>0.4
r2>0.2
r2>=0

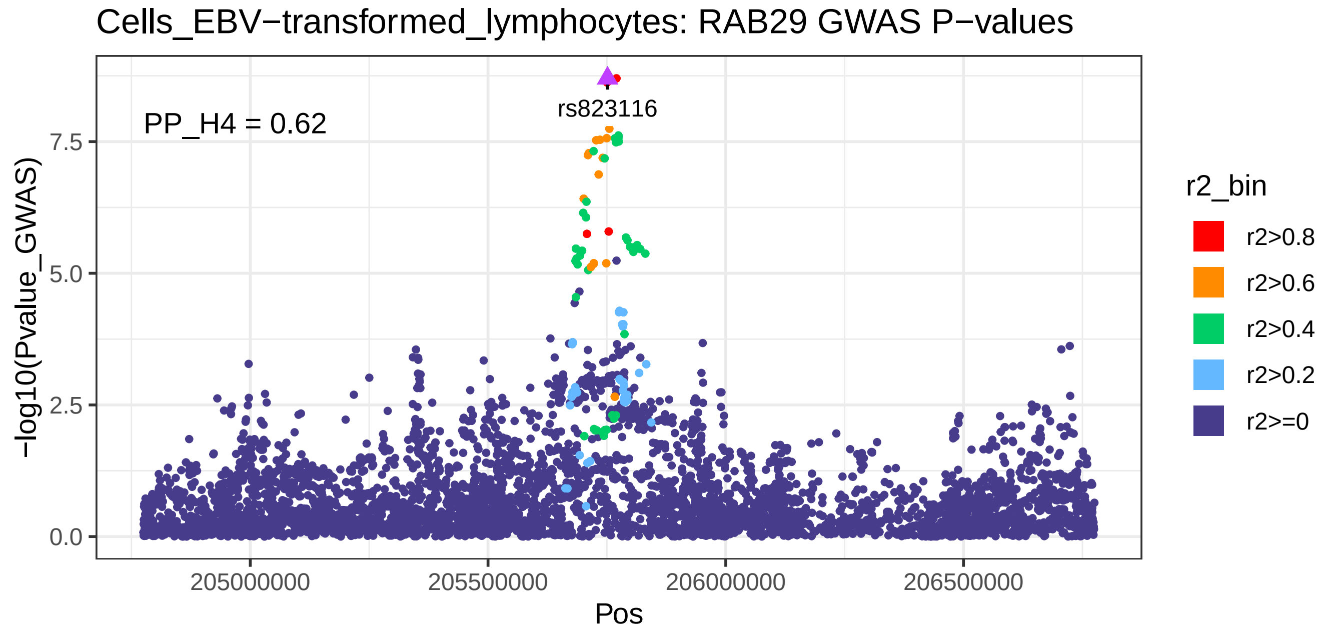
Cells_EBV−transformed_lymphocytes: RAB29 GWAS P−values
PP_H4 = 0.62
rs823116
−log10(Pvalue_GWAS)
0.0
2.5
5.0
7.5
205000000
205500000
206000000
206500000
Pos
r2_bin
r2>0.8
r2>0.6
r2>0.4
r2>0.2
r2>=0

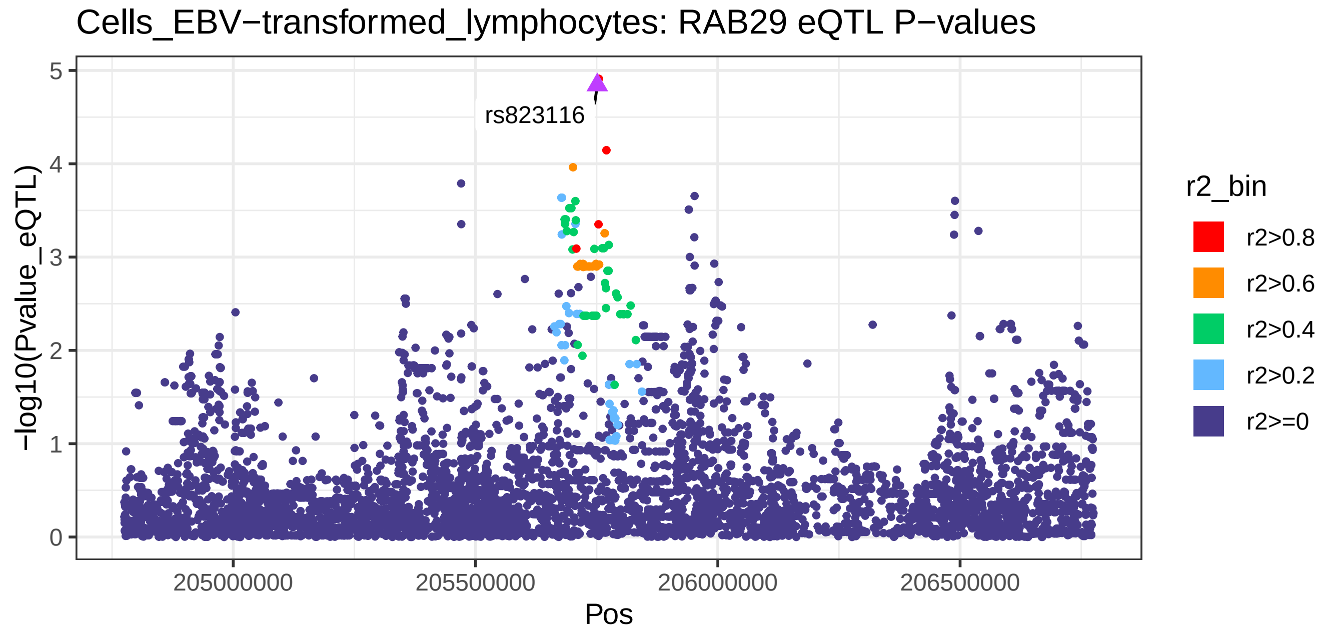
Cells_EBV−transformed_lymphocytes: RAB29 eQTL P−values
rs823116
−log10(Pvalue_eQTL)
0
1
2
3
4
5
205000000
205500000
206000000
206500000
Pos
r2_bin
r2>0.8
r2>0.6
r2>0.4
r2>0.2
r2>=0

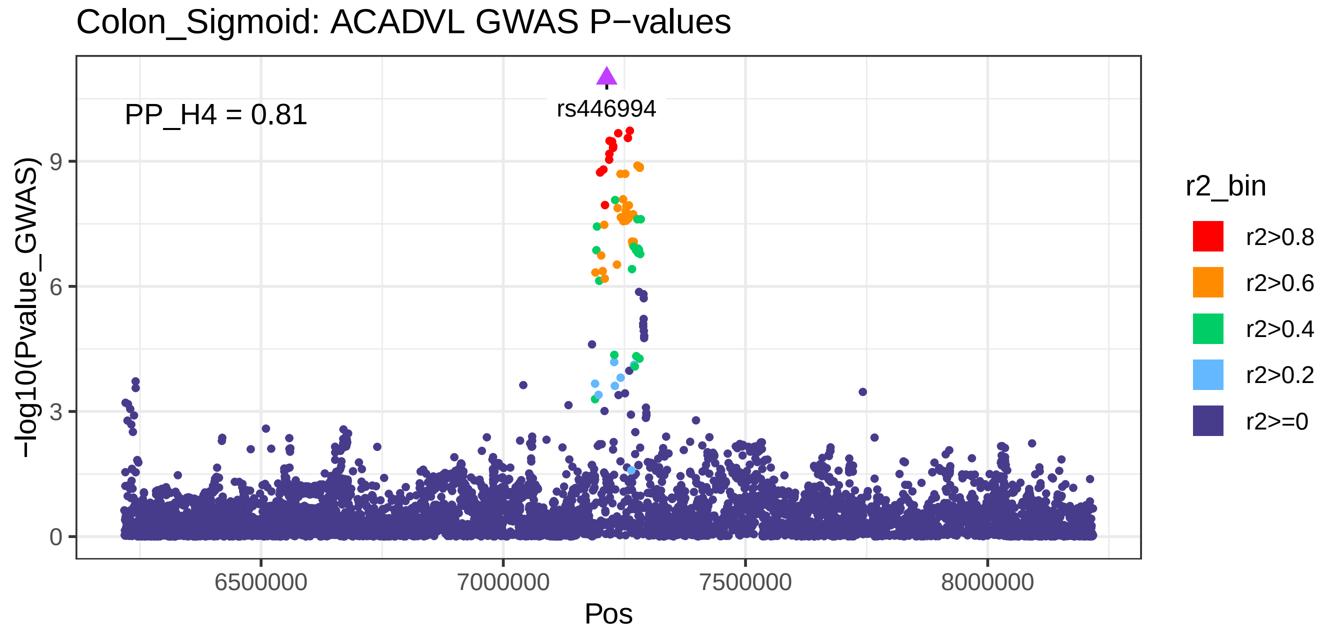
Colon_Sigmoid: ACADVL GWAS P−values
PP_H4 = 0.81
rs446994
−log10(Pvalue_GWAS)
0
3
6
9
6500000
7000000
7500000
8000000
Pos
r2_bin
r2>0.8
r2>0.6
r2>0.4
r2>0.2
r2>=0

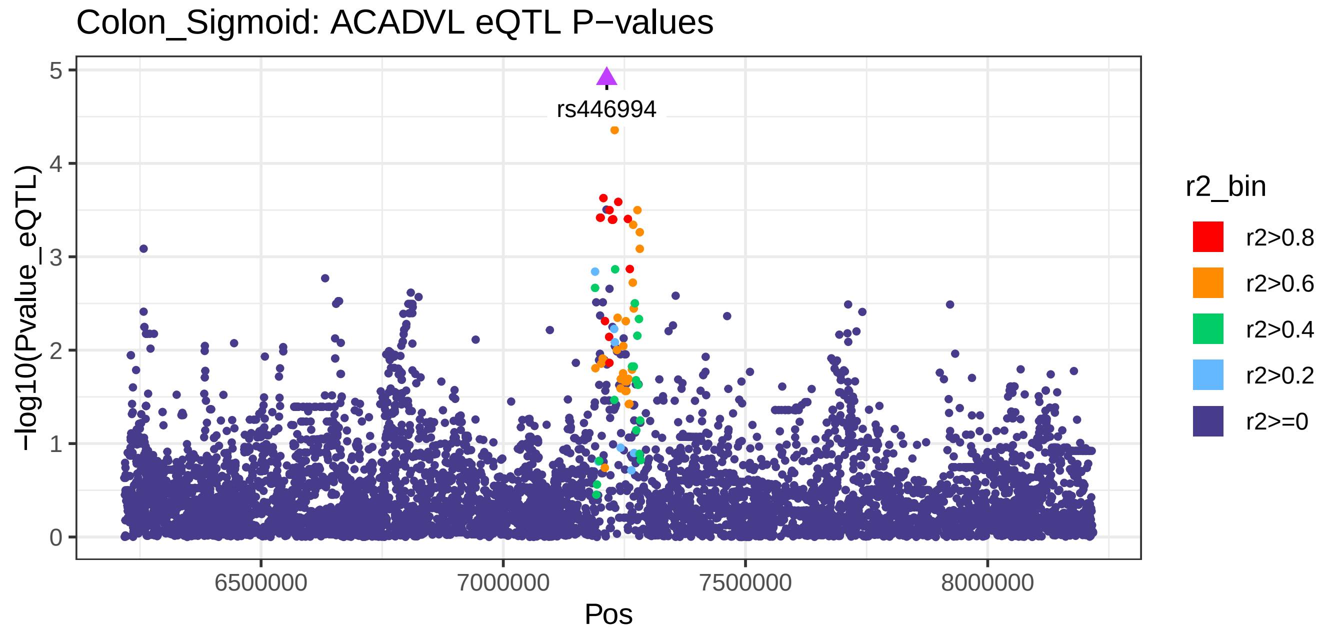
Colon_Sigmoid: ACADVL eQTL P−values
rs446994
−log10(Pvalue_eQTL)
0
1
2
3
4
5
6500000
7000000
7500000
8000000
Pos
r2_bin
r2>0.8
r2>0.6
r2>0.4
r2>0.2
r2>=0

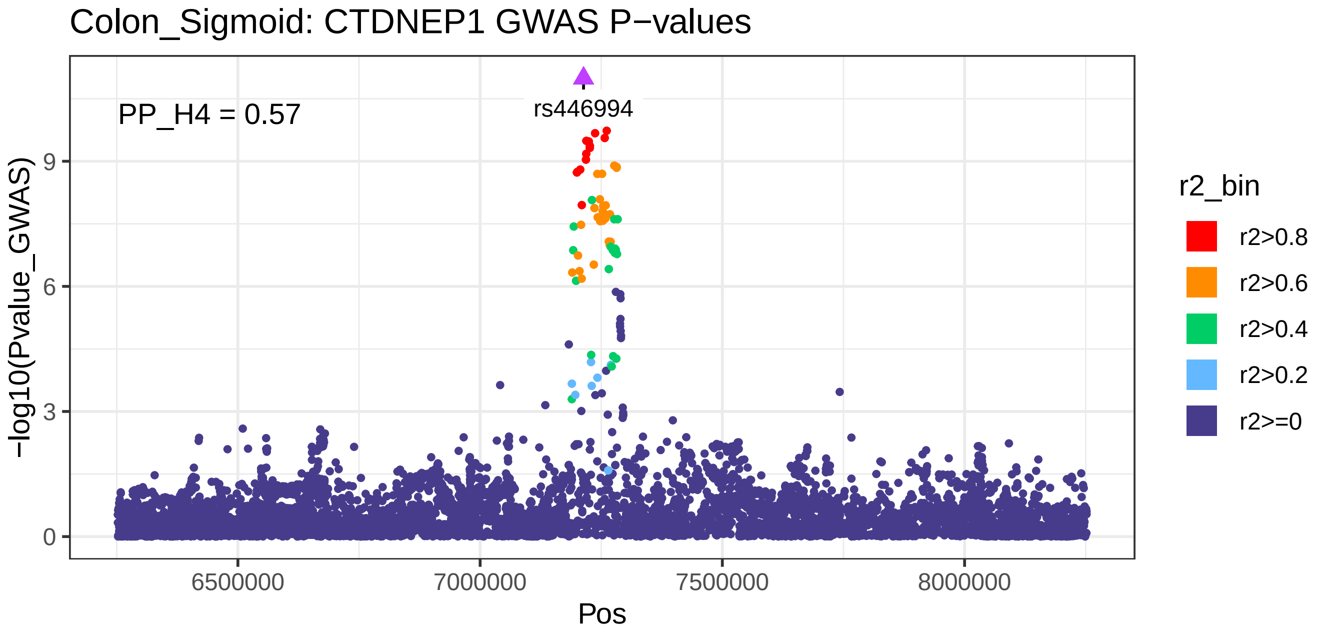
Colon_Sigmoid: CTDNEP1 GWAS P−values
PP_H4 = 0.57
rs446994
−log10(Pvalue_GWAS)
0
3
6
9
6500000
7000000
7500000
8000000
Pos
r2_bin
r2>0.8
r2>0.6
r2>0.4
r2>0.2
r2>=0

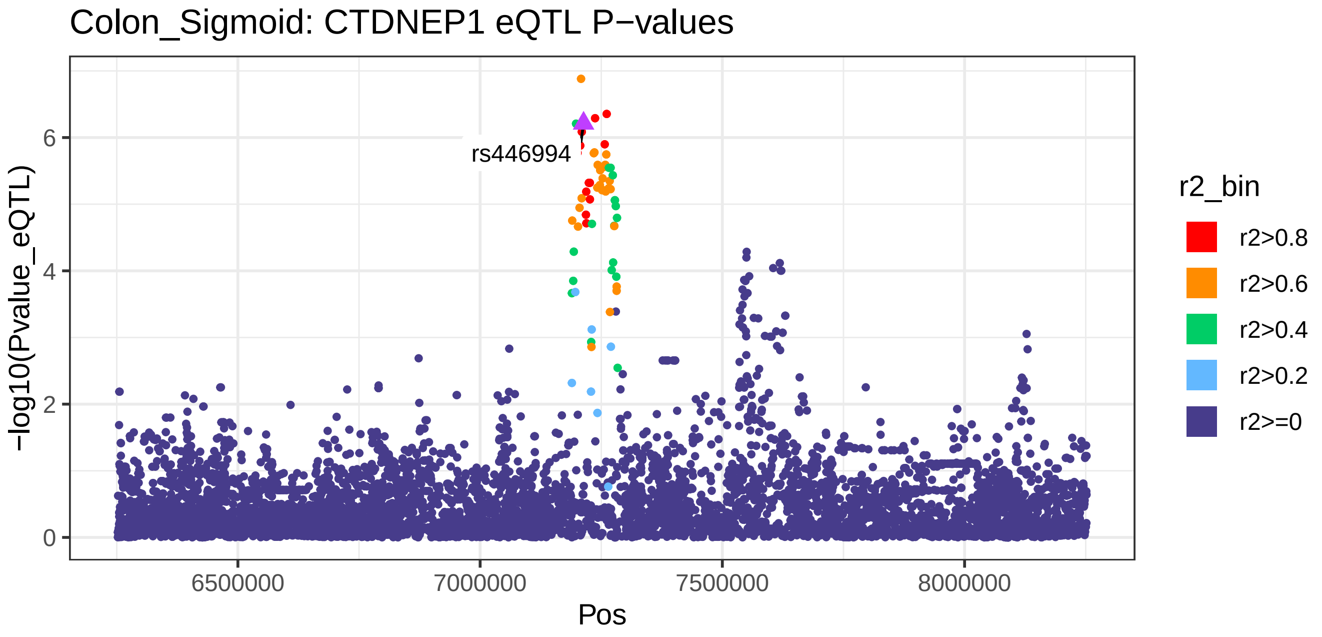
Colon_Sigmoid: CTDNEP1 eQTL P−values
rs446994
−log10(Pvalue_eQTL)
0
2
4
6
6500000
7000000
7500000
8000000
Pos
r2_bin
r2>0.8
r2>0.6
r2>0.4
r2>0.2
r2>=0

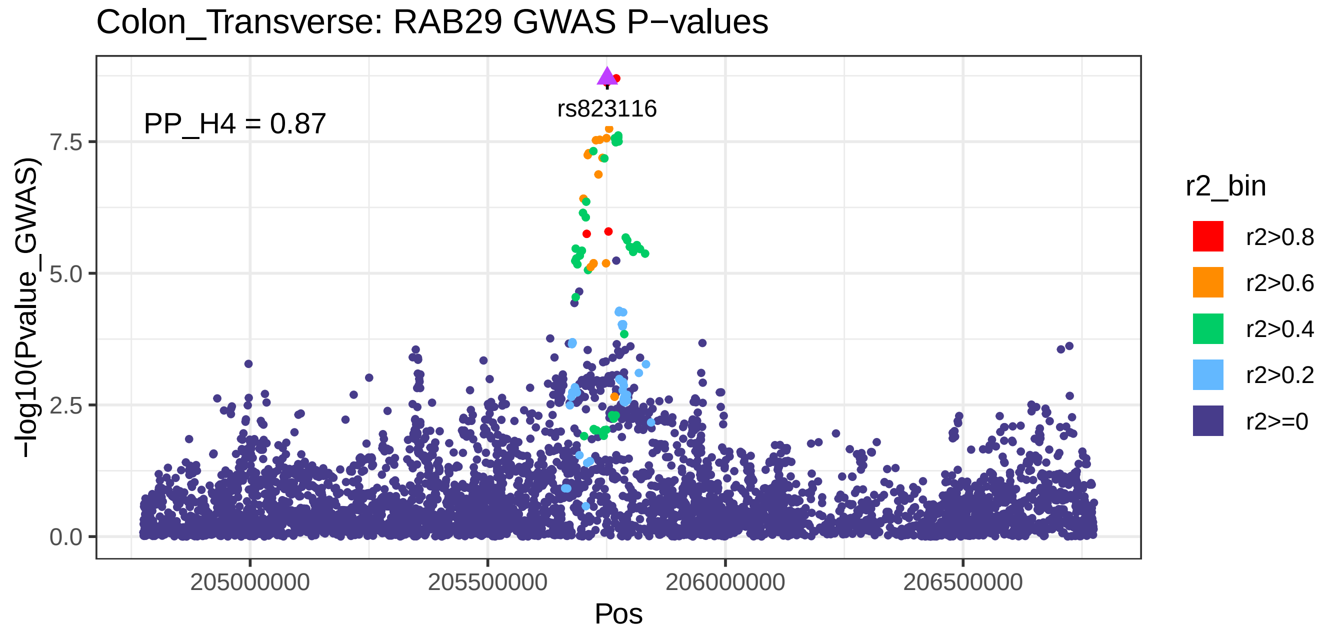
Colon_Transverse: RAB29 GWAS P−values
PP_H4 = 0.87
rs823116
−log10(Pvalue_GWAS)
0.0
2.5
5.0
7.5
205000000
205500000
206000000
206500000
Pos
r2_bin
r2>0.8
r2>0.6
r2>0.4
r2>0.2
r2>=0

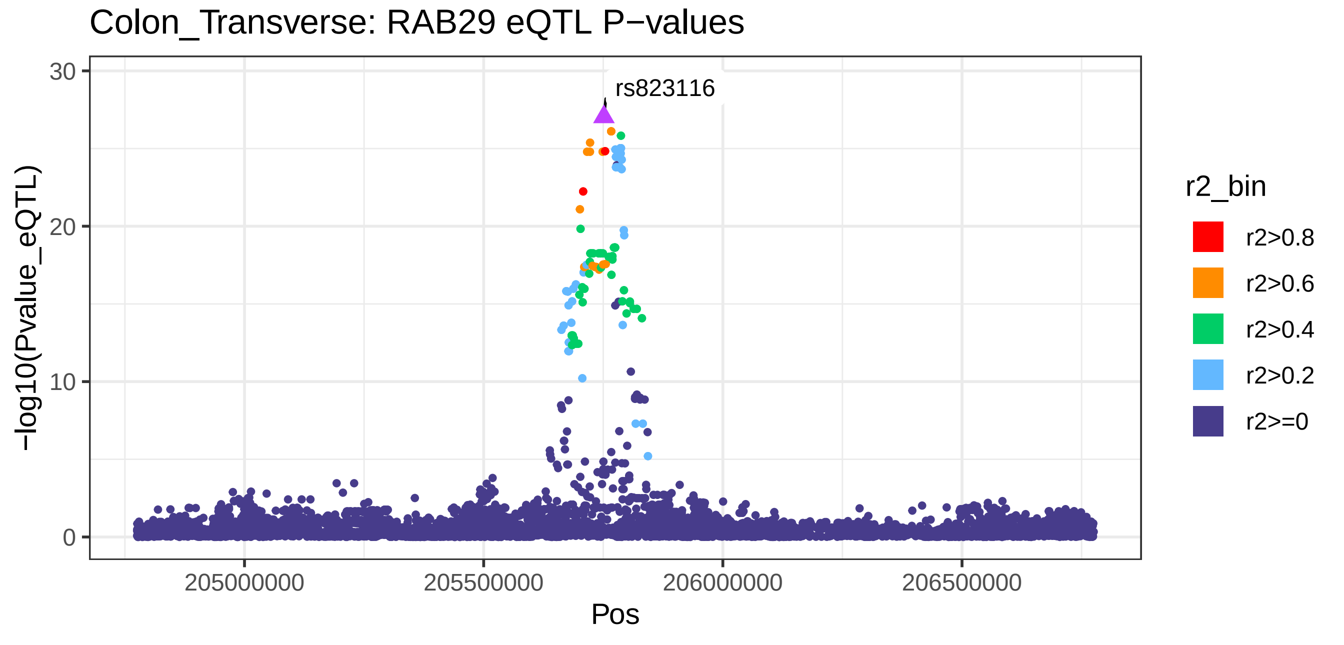
Colon_Transverse: RAB29 eQTL P−values
rs823116
−log10(Pvalue_eQTL)
0
10
20
30
205000000
205500000
206000000
206500000
Pos
r2_bin
r2>0.8
r2>0.6
r2>0.4
r2>0.2
r2>=0

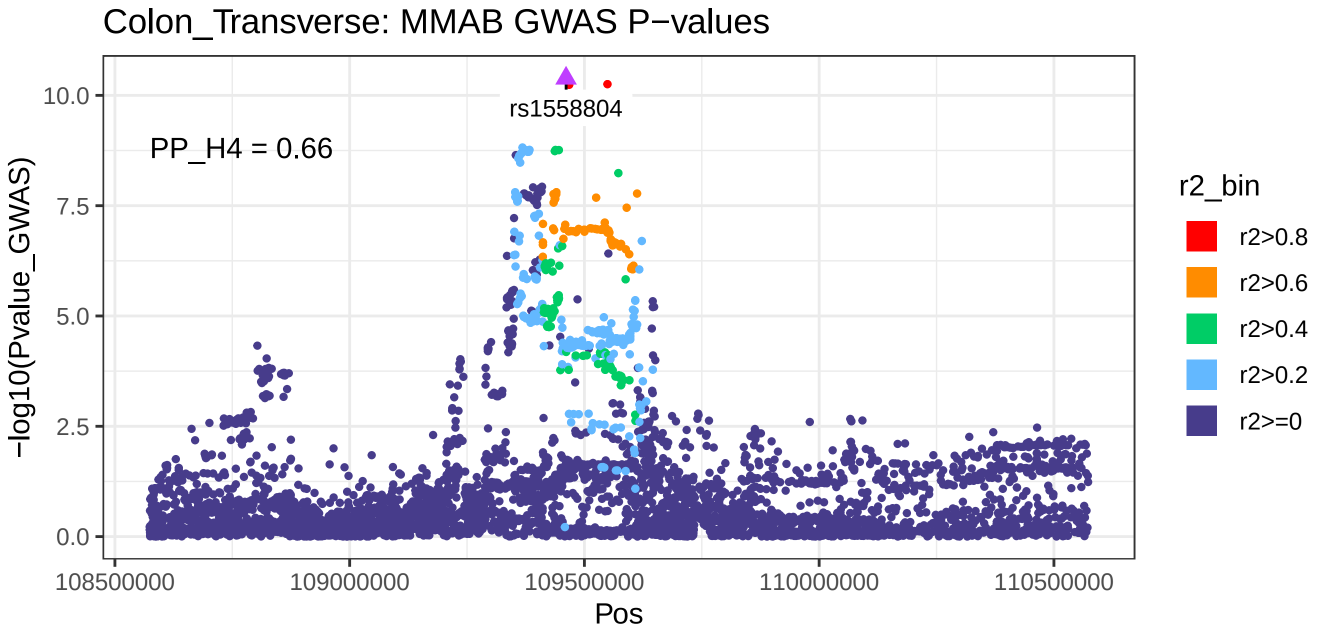

Colon_Transverse: MMAB GWAS P−values
rs1558804
PP_H4 = 0.66
−log10(Pvalue_GWAS)
10.0
7.5
5.0
2.5
0.0
108500000
109000000
109500000
110000000
110500000
Pos
r2_bin
r2>0.8
r2>0.6
r2>0.4
r2>0.2
r2>=0


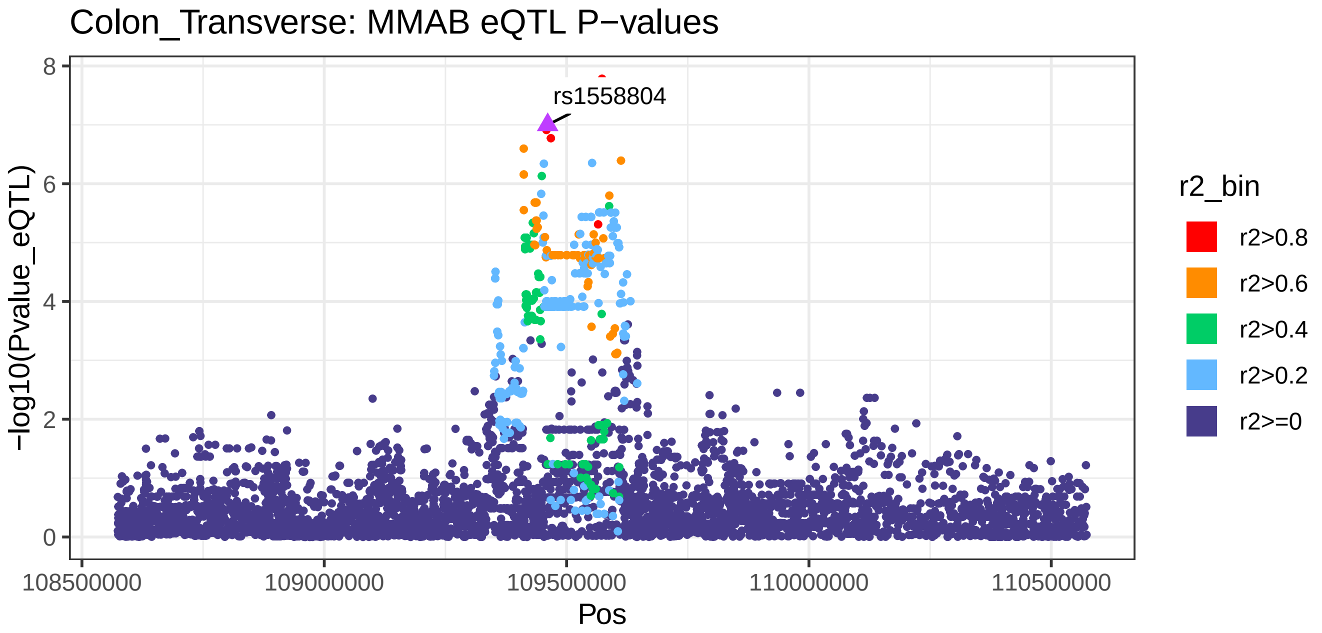

Colon_Transverse: MMAB eQTL P−values
rs1558804
−log10(Pvalue_eQTL)
8
6
4
2
0
108500000
109000000
109500000
110000000
110500000
Pos
r2_bin
r2>0.8
r2>0.6
r2>0.4
r2>0.2
r2>=0

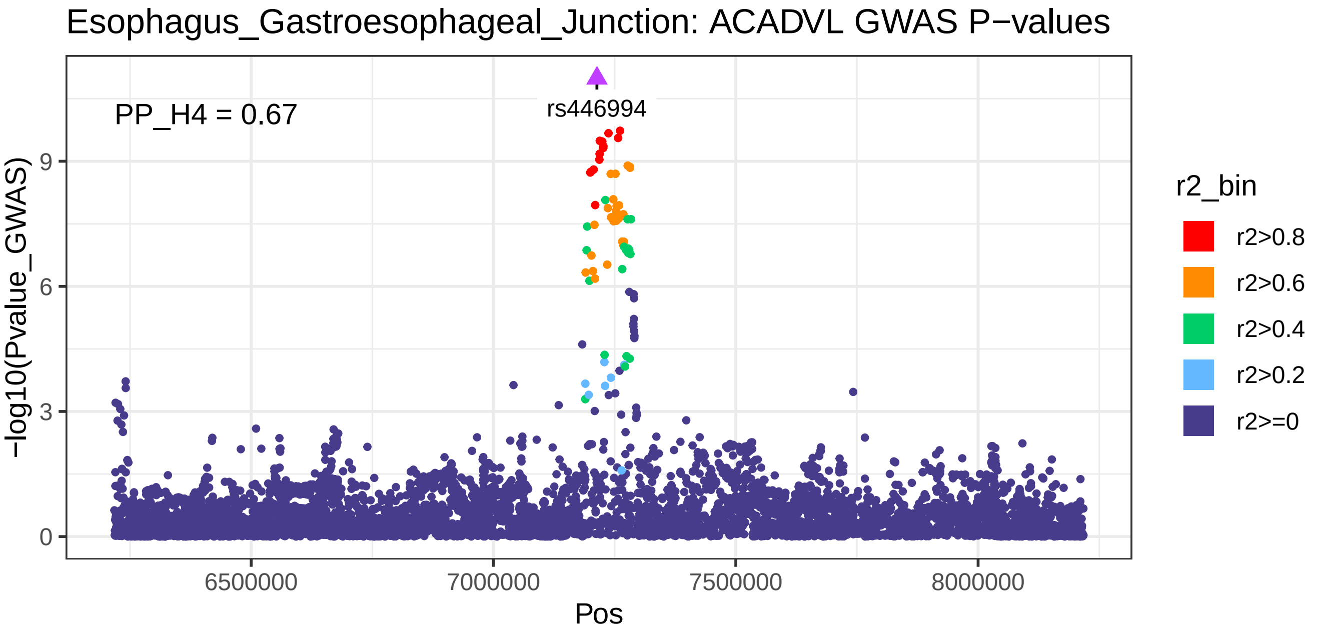
Esophagus_Gastroesophageal_Junction: ACADVL GWAS P−values
PP_H4 = 0.67
rs446994
−log10(Pvalue_GWAS)
0
3
6
9
6500000
7000000
7500000
8000000
Pos
r2_bin
r2>0.8
r2>0.6
r2>0.4
r2>0.2
r2>=0

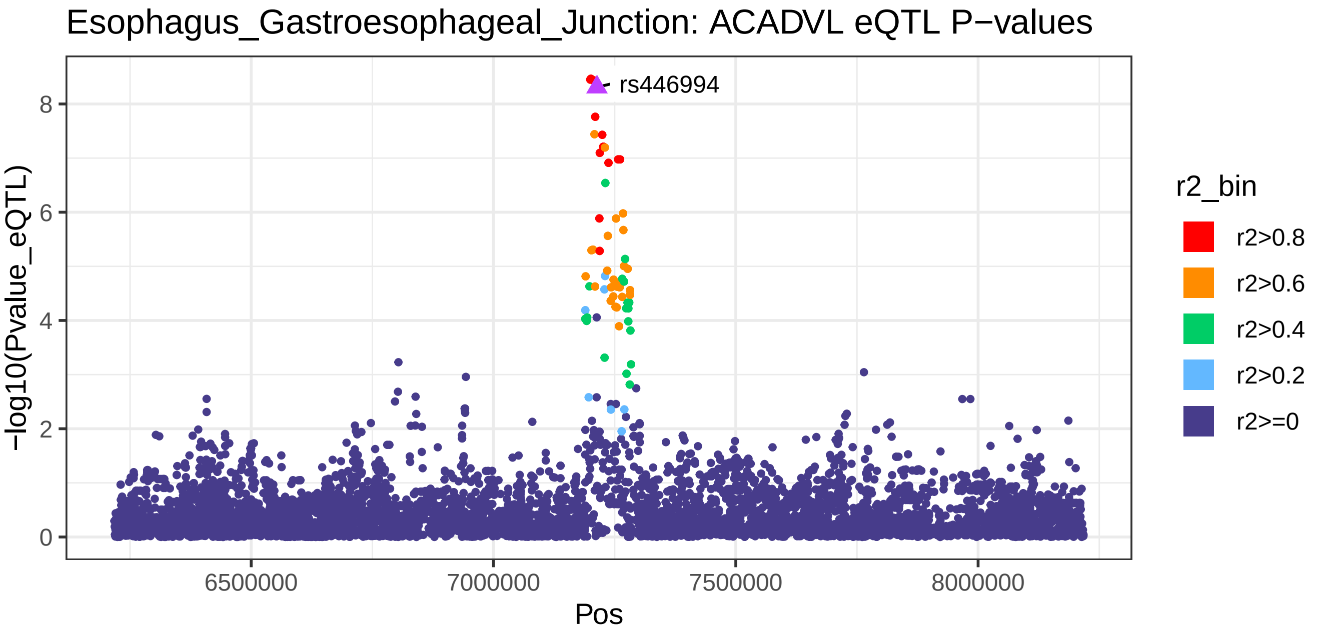
Esophagus_Gastroesophageal_Junction: ACADVL eQTL P−values
rs446994
−log10(Pvalue_eQTL)
0
2
4
6
8
6500000
7000000
7500000
8000000
Pos
r2_bin
r2>0.8
r2>0.6
r2>0.4
r2>0.2
r2>=0

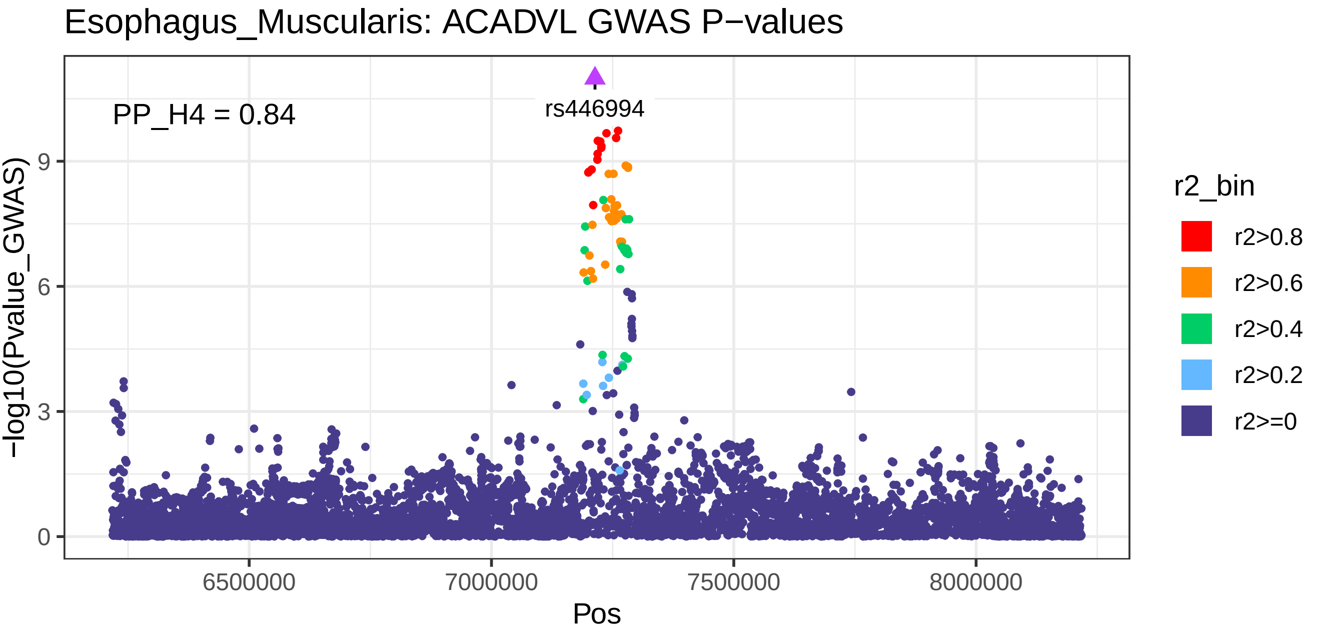

Esophagus_Muscularis: ACADVL GWAS P−values
PP_H4 = 0.84
rs446994
−log10(Pvalue_GWAS)
0
3
6
9
6500000
7000000
7500000
8000000
Pos
r2_bin
r2>0.8
r2>0.6
r2>0.4
r2>0.2
r2>=0


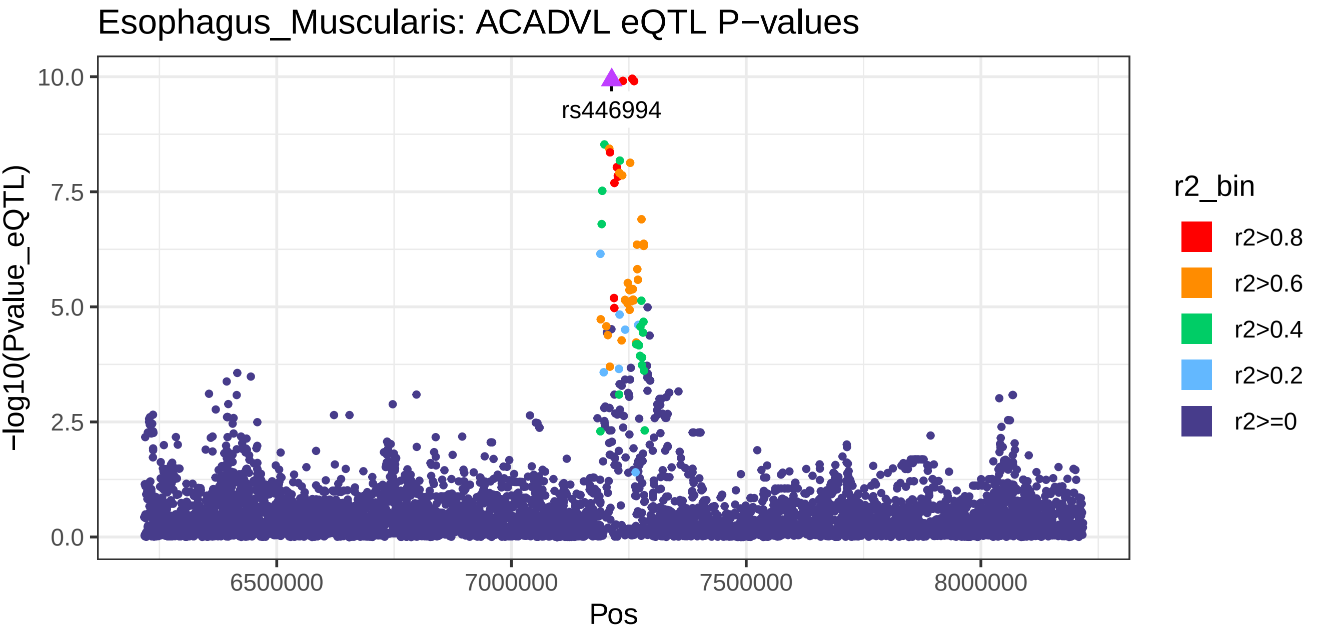

Esophagus_Muscularis: ACADVL eQTL P−values
rs446994
−log10(Pvalue_eQTL)
0.0
2.5
5.0
7.5
10.0
6500000
7000000
7500000
8000000
Pos
r2_bin
r2>0.8
r2>0.6
r2>0.4
r2>0.2
r2>=0

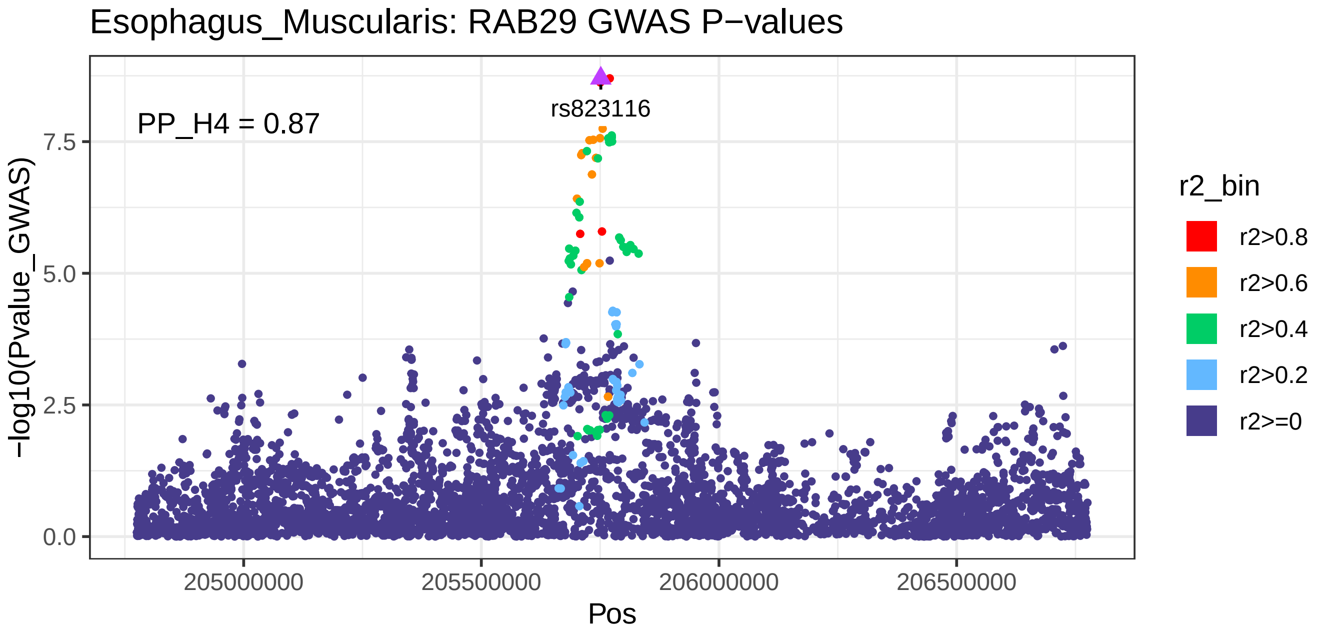
Esophagus_Muscularis: RAB29 GWAS P−values
PP_H4 = 0.87
rs823116
−log10(Pvalue_GWAS)
0.0
2.5
5.0
7.5
205000000
205500000
206000000
206500000
Pos
r2_bin
r2>0.8
r2>0.6
r2>0.4
r2>0.2
r2>=0

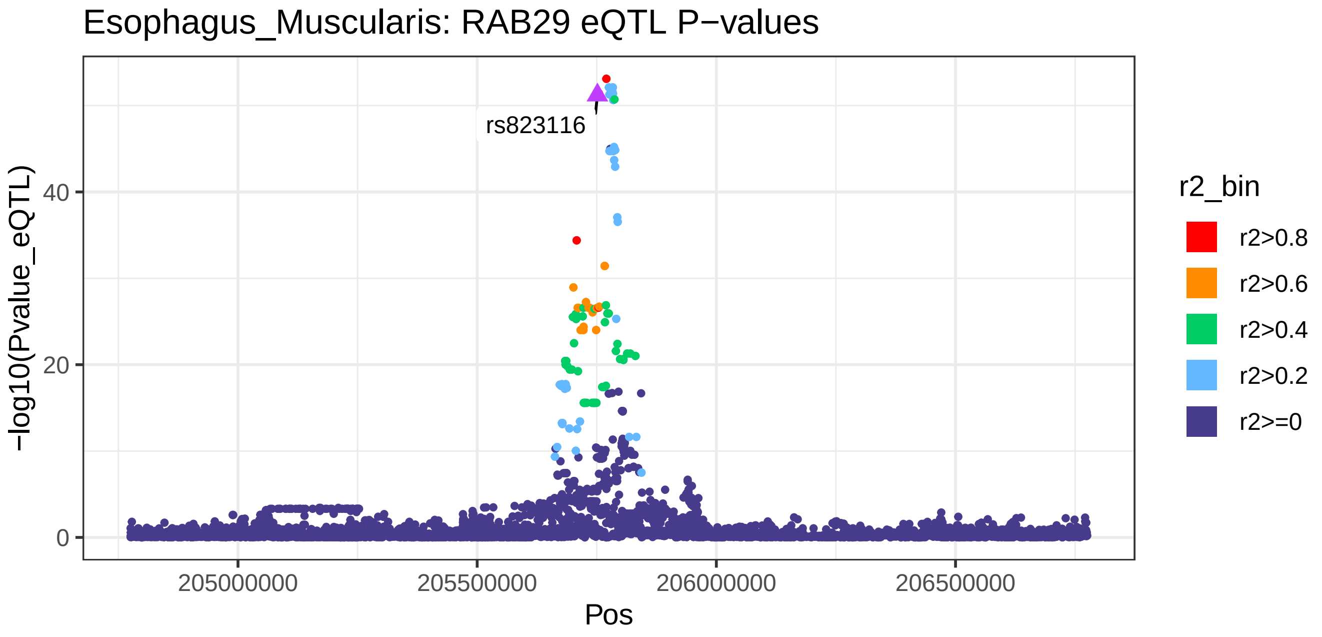
Esophagus_Muscularis: RAB29 eQTL P−values
rs823116
−log10(Pvalue_eQTL)
0
20
40
205000000
205500000
206000000
206500000
Pos
r2_bin
r2>0.8
r2>0.6
r2>0.4
r2>0.2
r2>=0

Esophagus_Muscularis: LMO7 GWAS P−values

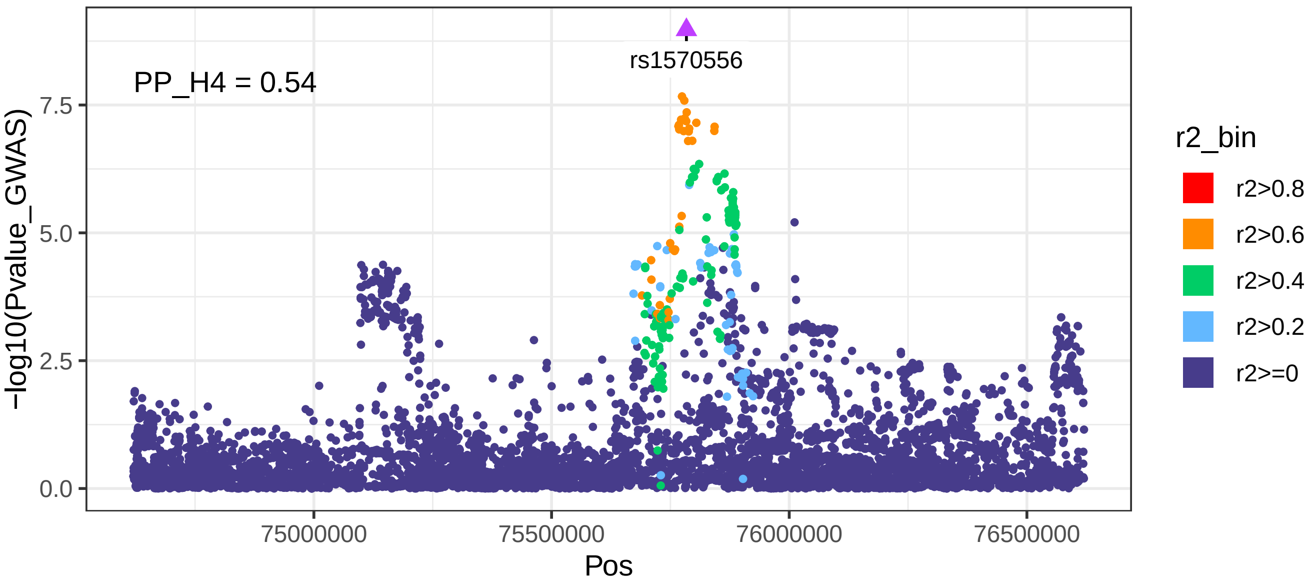


Esophagus_Muscularis: LMO7 eQTL P−values

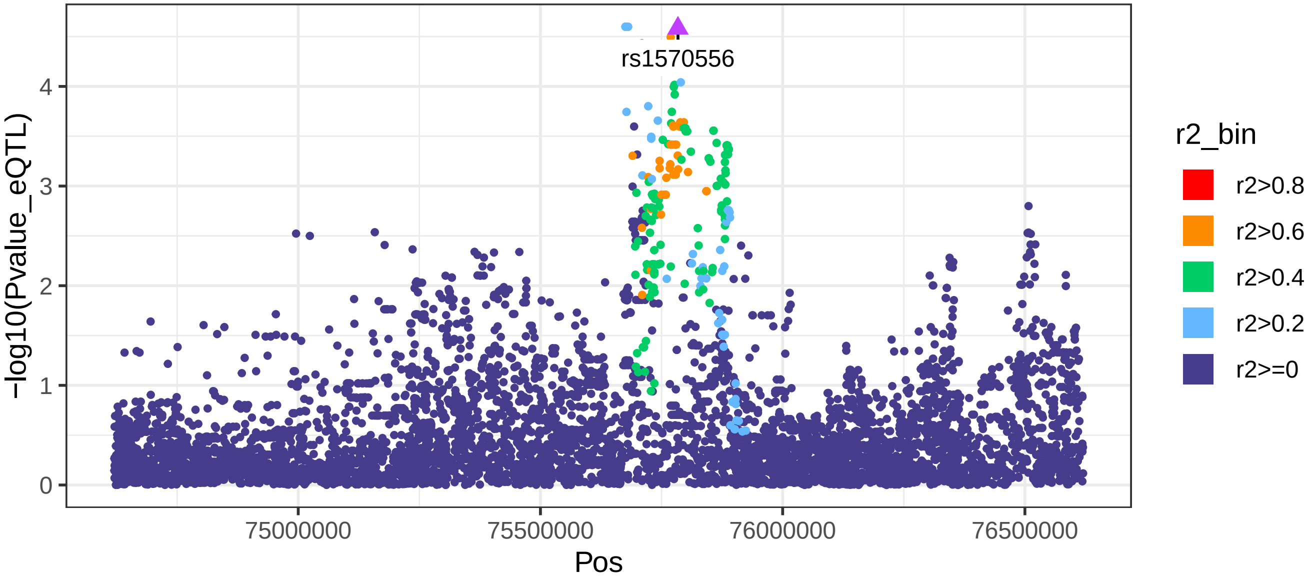

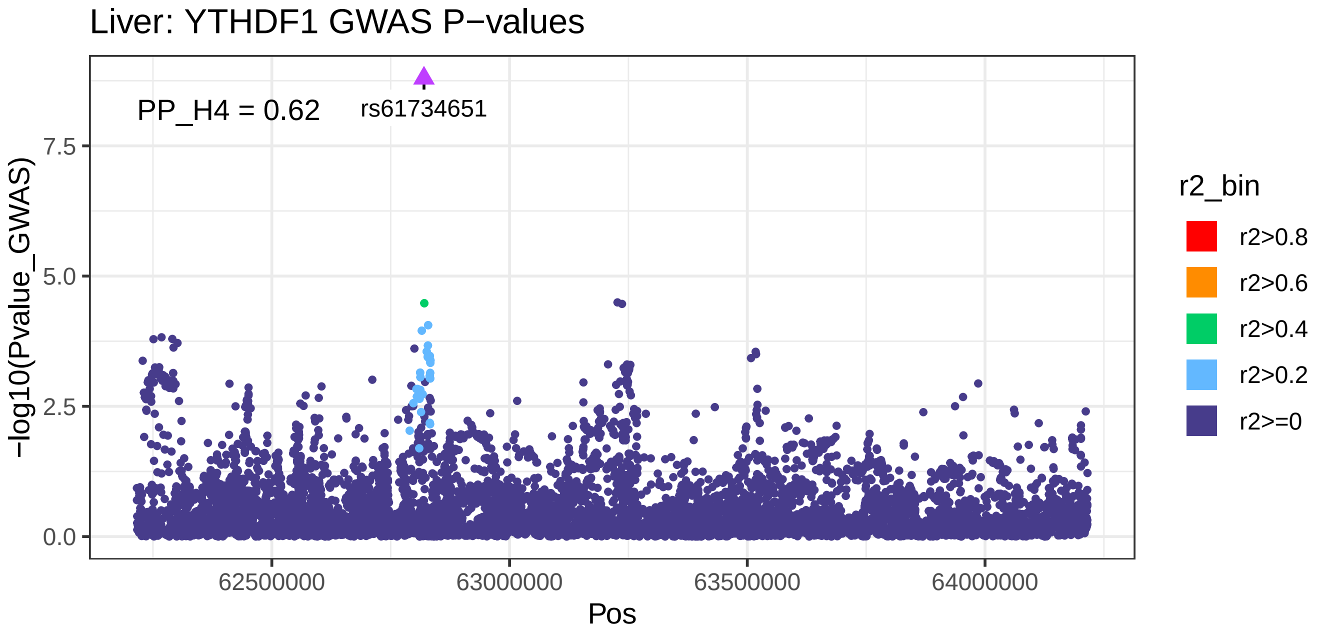
Liver: YTHDF1 GWAS P−values
PP_H4 = 0.62
rs61734651
7.5
5.0
2.5
0.0
−log10(Pvalue_GWAS)
62500000
63000000
63500000
64000000
Pos
r2_bin
r2>0.8
r2>0.6
r2>0.4
r2>0.2
r2>=0

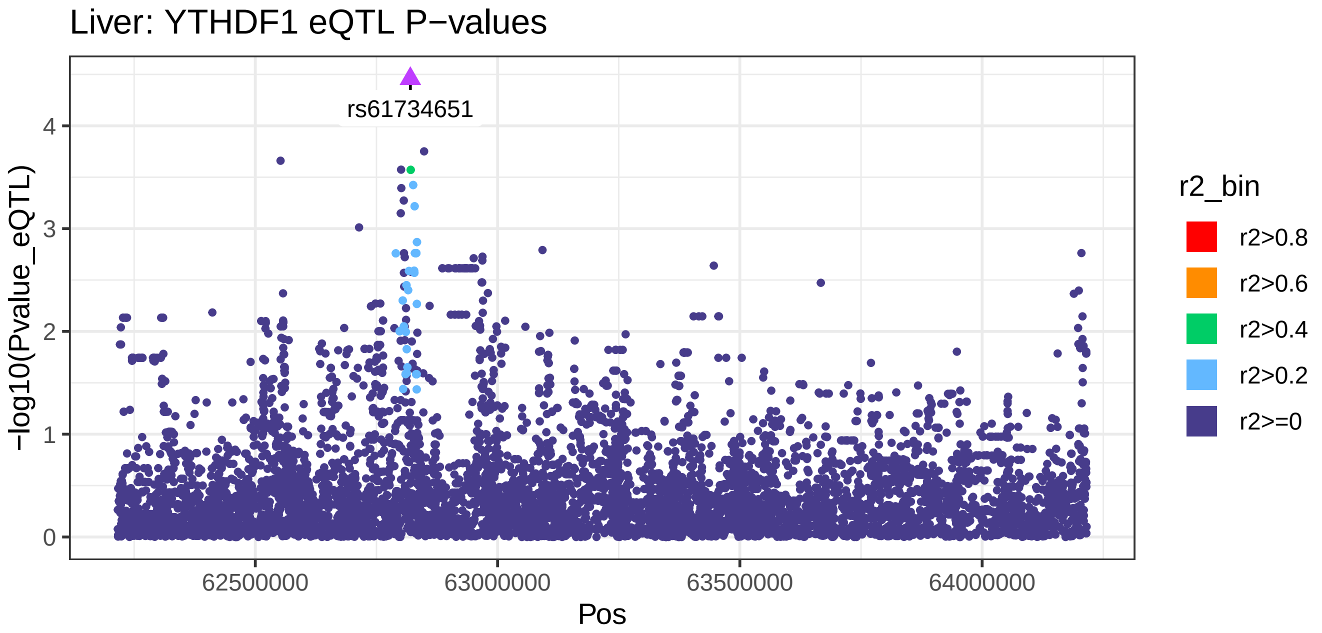
Liver: YTHDF1 eQTL P−values
rs61734651
4
3
2
1
0
−log10(Pvalue_eQTL)
62500000
63000000
63500000
64000000
Pos
r2_bin
r2>0.8
r2>0.6
r2>0.4
r2>0.2
r2>=0

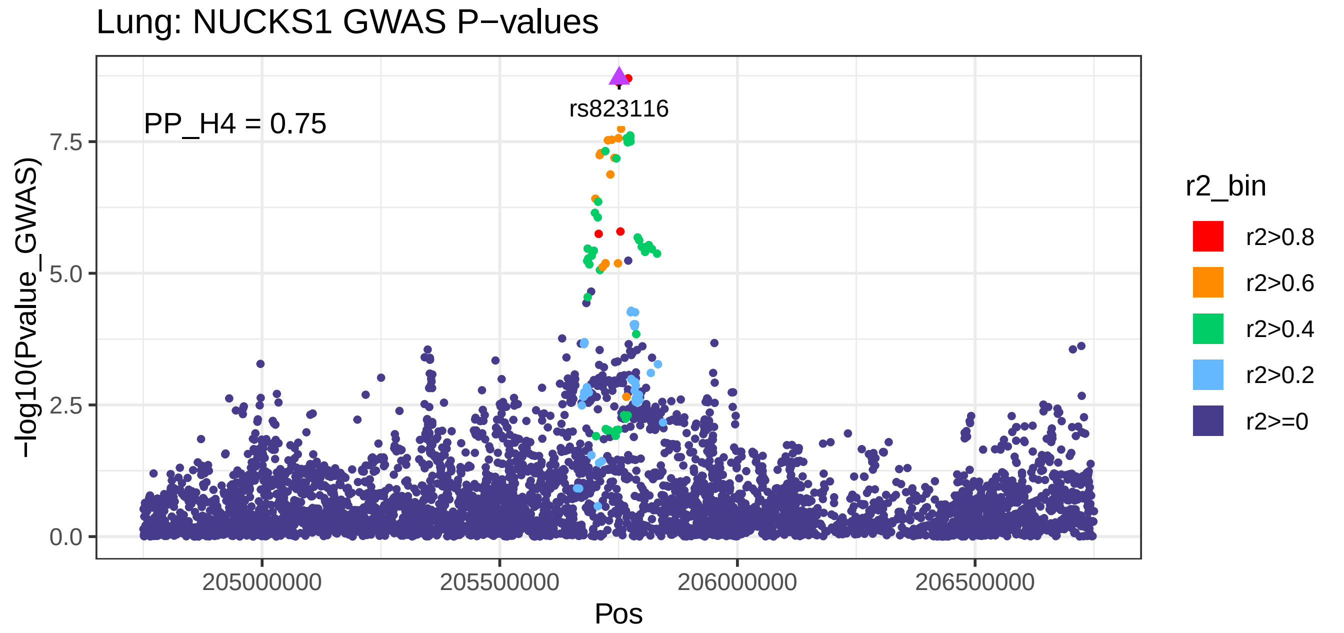

Lung: NUCKS1 GWAS P−values
PP_H4 = 0.75
rs823116
−log10(Pvalue_GWAS)
0.0
2.5
5.0
7.5
205000000
205500000
206000000
206500000
Pos
r2_bin
r2>0.8
r2>0.6
r2>0.4
r2>0.2
r2>=0


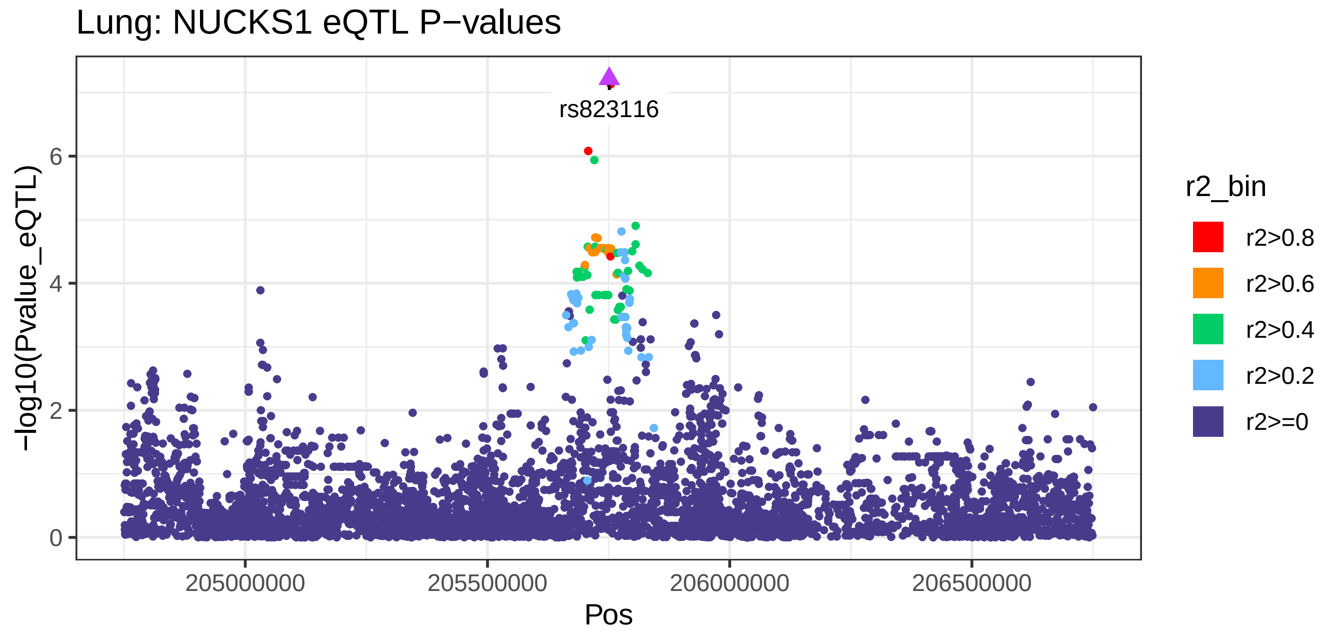

Lung: NUCKS1 eQTL P−values
rs823116
−log10(Pvalue_eQTL)
0
2
4
6
205000000
205500000
206000000
206500000
Pos
r2_bin
r2>0.8
r2>0.6
r2>0.4
r2>0.2
r2>=0

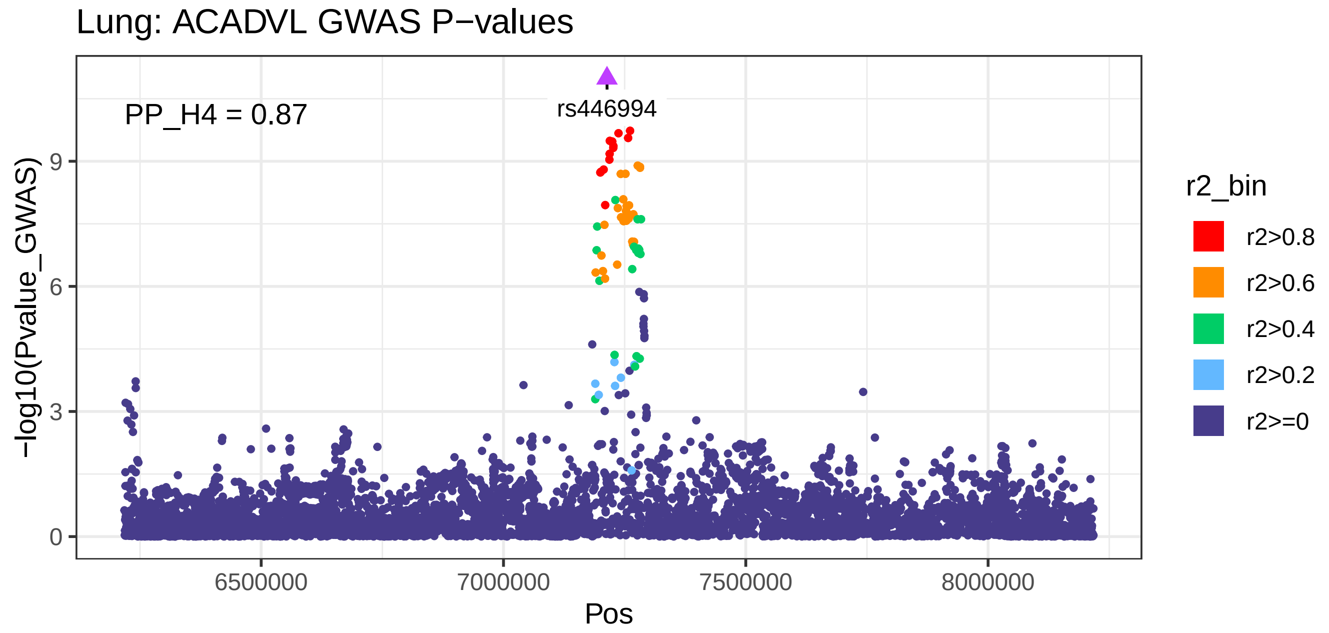
Lung: ACADVL GWAS P−values
PP_H4 = 0.87
rs446994
−log10(Pvalue_GWAS)
0
3
6
9
6500000
7000000
7500000
8000000
Pos
r2_bin
r2>0.8
r2>0.6
r2>0.4
r2>0.2
r2>=0

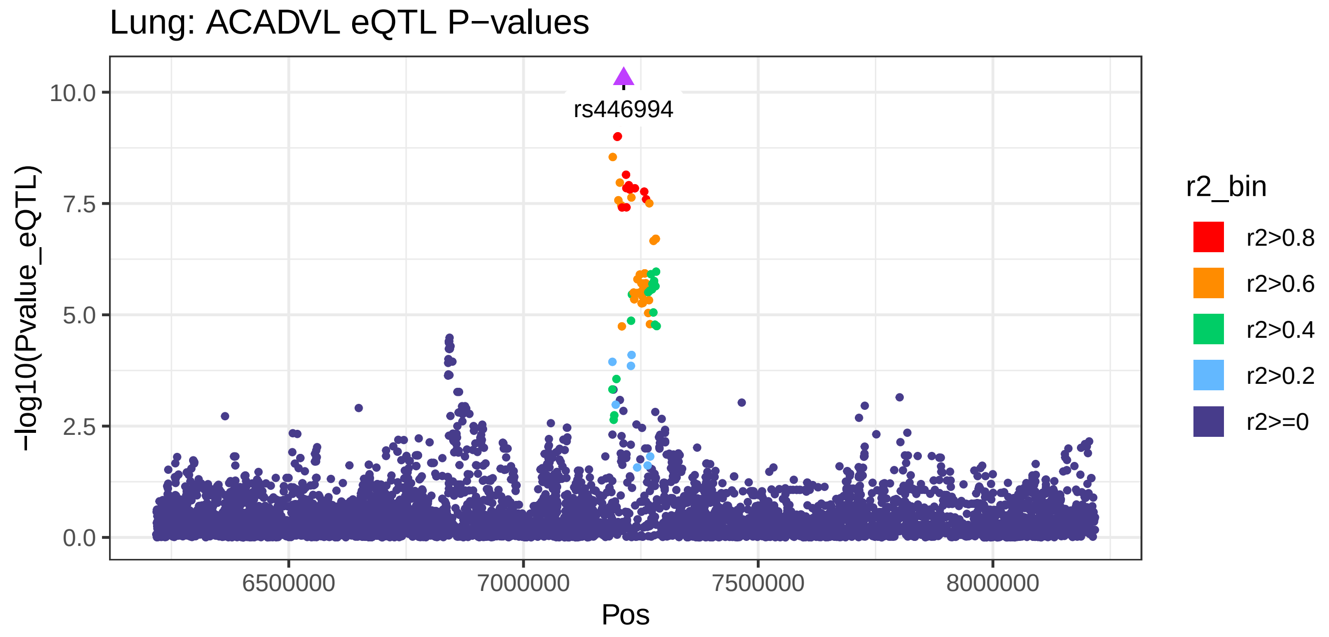
Lung: ACADVL eQTL P−values
rs446994
−log10(Pvalue_eQTL)
0.0
2.5
5.0
7.5
10.0
6500000
7000000
7500000
8000000
Pos
r2_bin
r2>0.8
r2>0.6
r2>0.4
r2>0.2
r2>=0

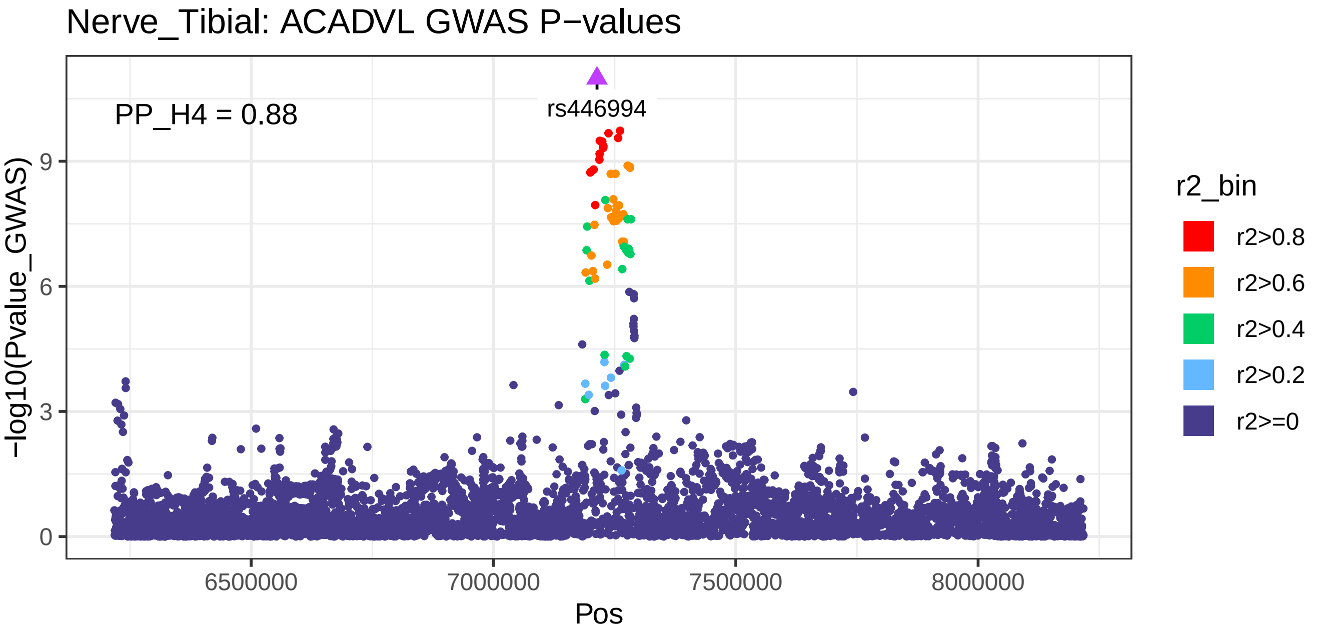
Nerve_Tibial: ACADVL GWAS P−values
PP_H4 = 0.88
rs446994
−log10(Pvalue_GWAS)
9
6
3
0
6500000
7000000
7500000
8000000
Pos
r2_bin
r2>0.8
r2>0.6
r2>0.4
r2>0.2
r2>=0

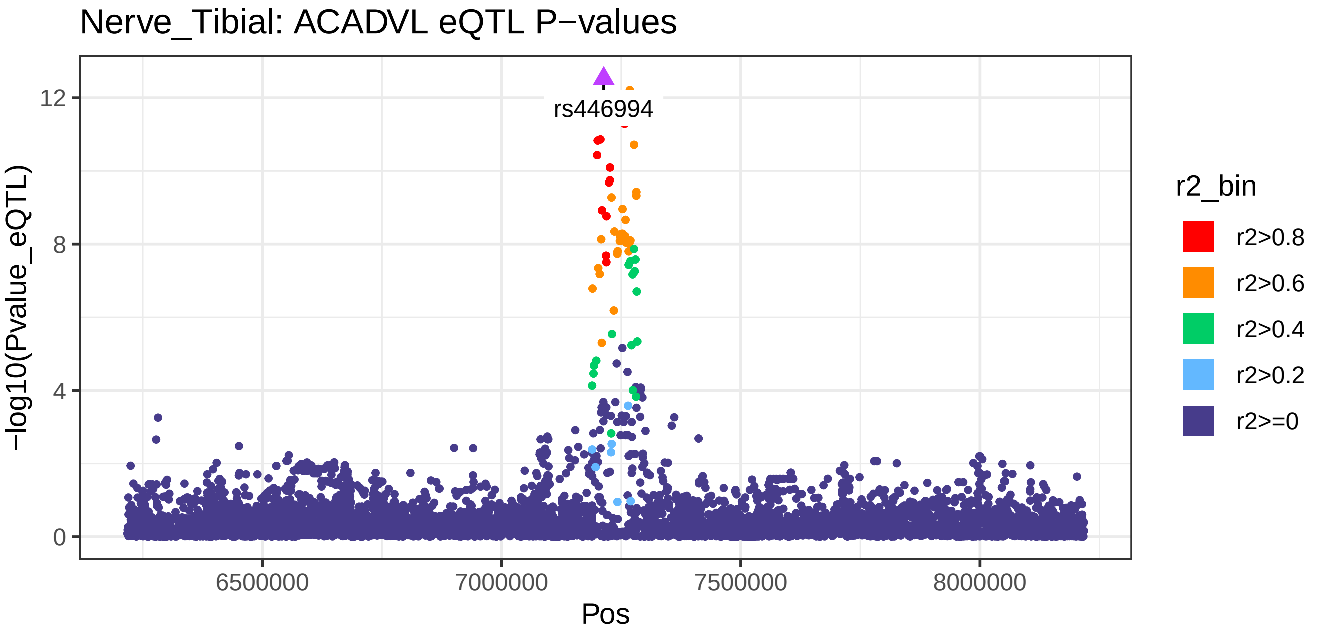
Nerve_Tibial: ACADVL eQTL P−values
rs446994
−log10(Pvalue_eQTL)
12
8
4
0
6500000
7000000
7500000
8000000
Pos
r2_bin
r2>0.8
r2>0.6
r2>0.4
r2>0.2
r2>=0

Nerve_Tibial: ENSG00000255176 GWAS P−values

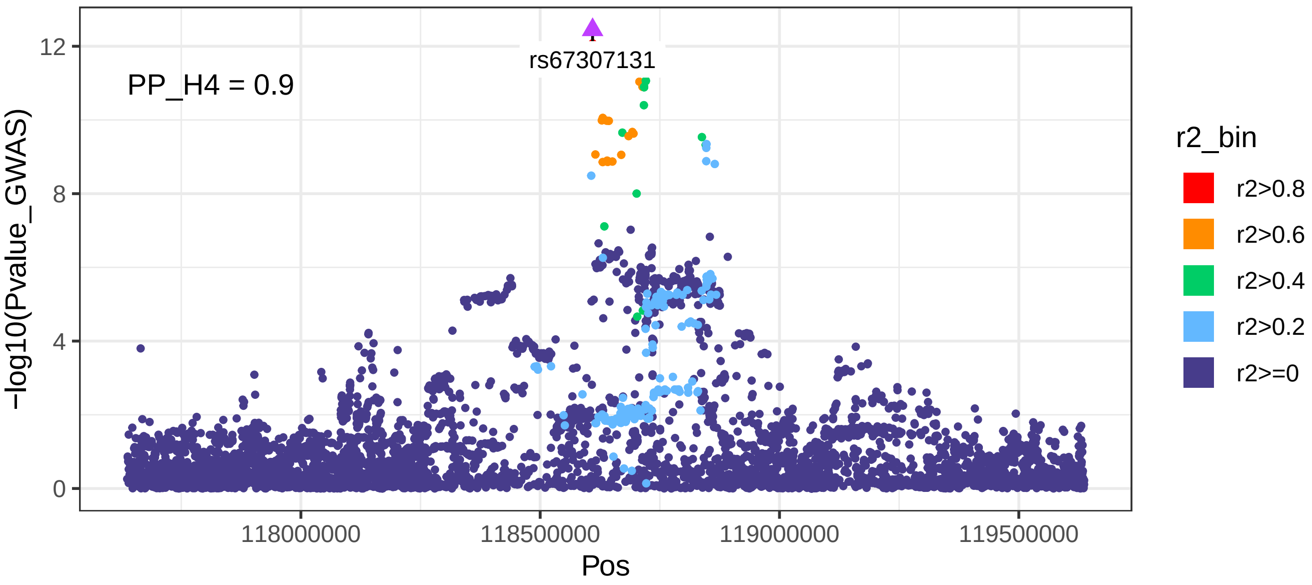


Nerve_Tibial: ENSG00000255176 eQTL P−values

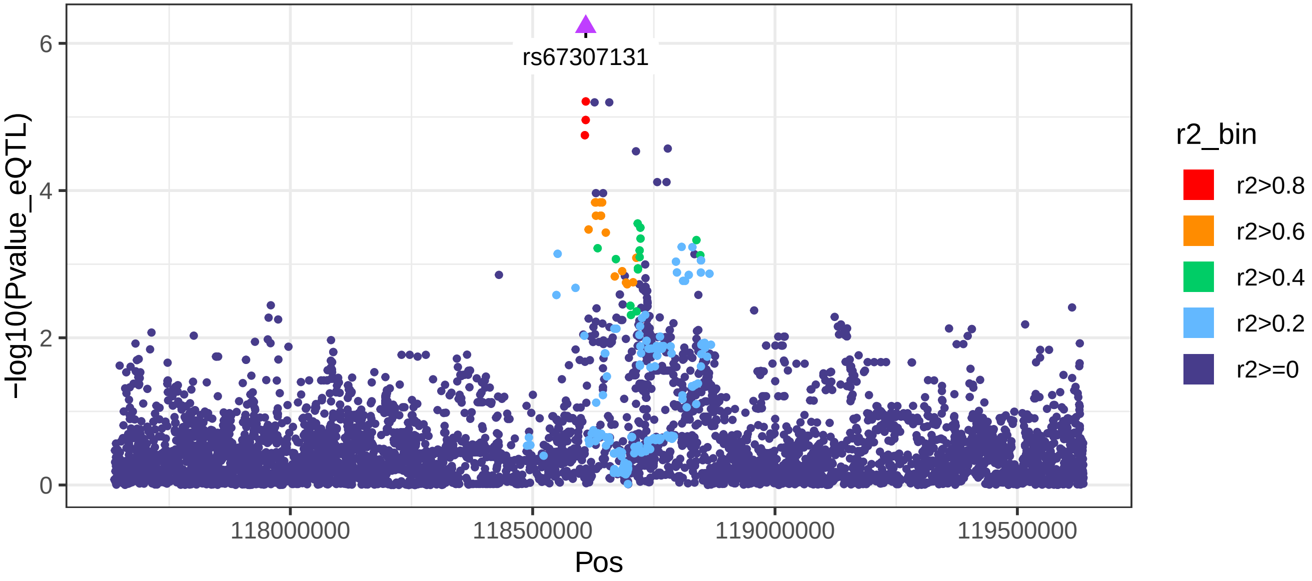

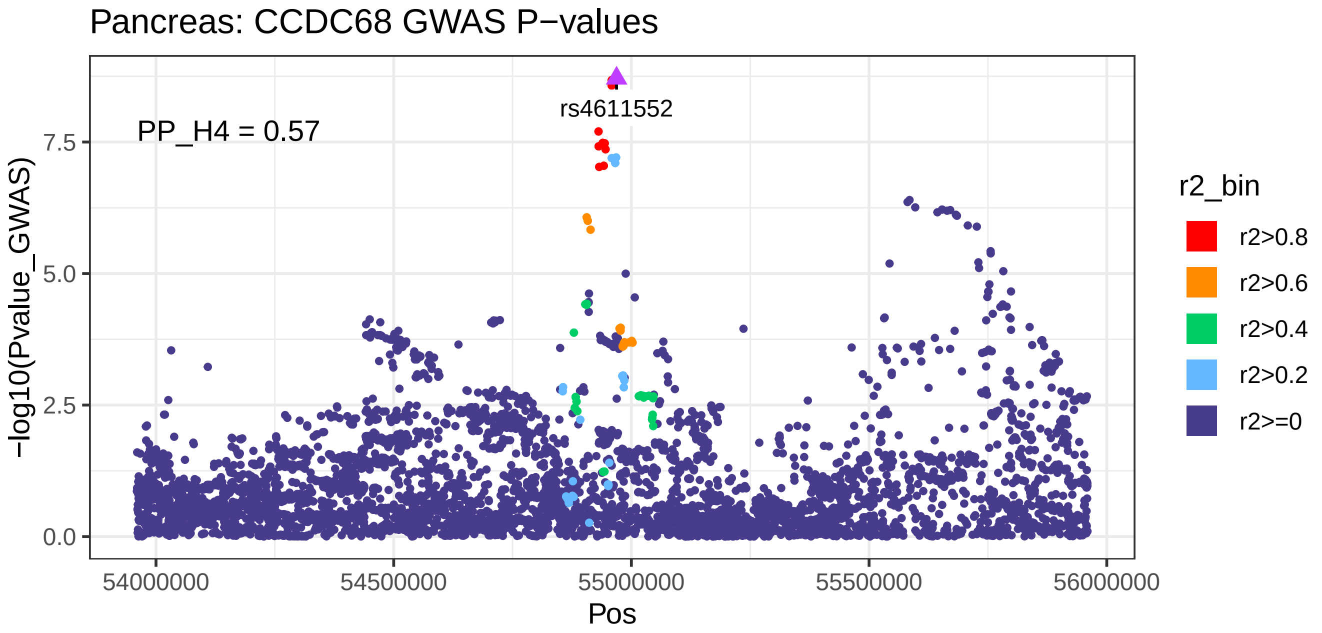
Pancreas: CCDC68 GWAS P−values
PP_H4 = 0.57
rs4611552
−log10(Pvalue_GWAS)
0.0
2.5
5.0
7.5
54000000
54500000
55000000
55500000
56000000
Pos
r2_bin
r2>0.8
r2>0.6
r2>0.4
r2>0.2
r2>=0

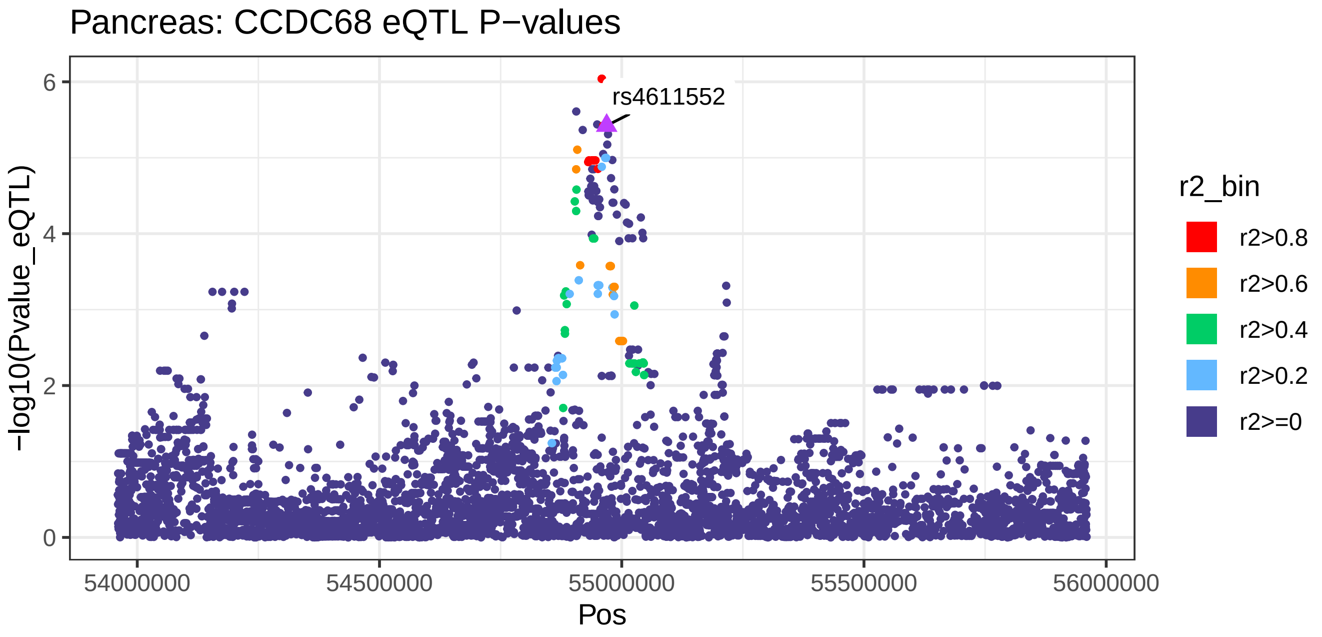
Pancreas: CCDC68 eQTL P−values
rs4611552
−log10(Pvalue_eQTL)
0
2
4
6
54000000
54500000
55000000
55500000
56000000
Pos
r2_bin
r2>0.8
r2>0.6
r2>0.4
r2>0.2
r2>=0

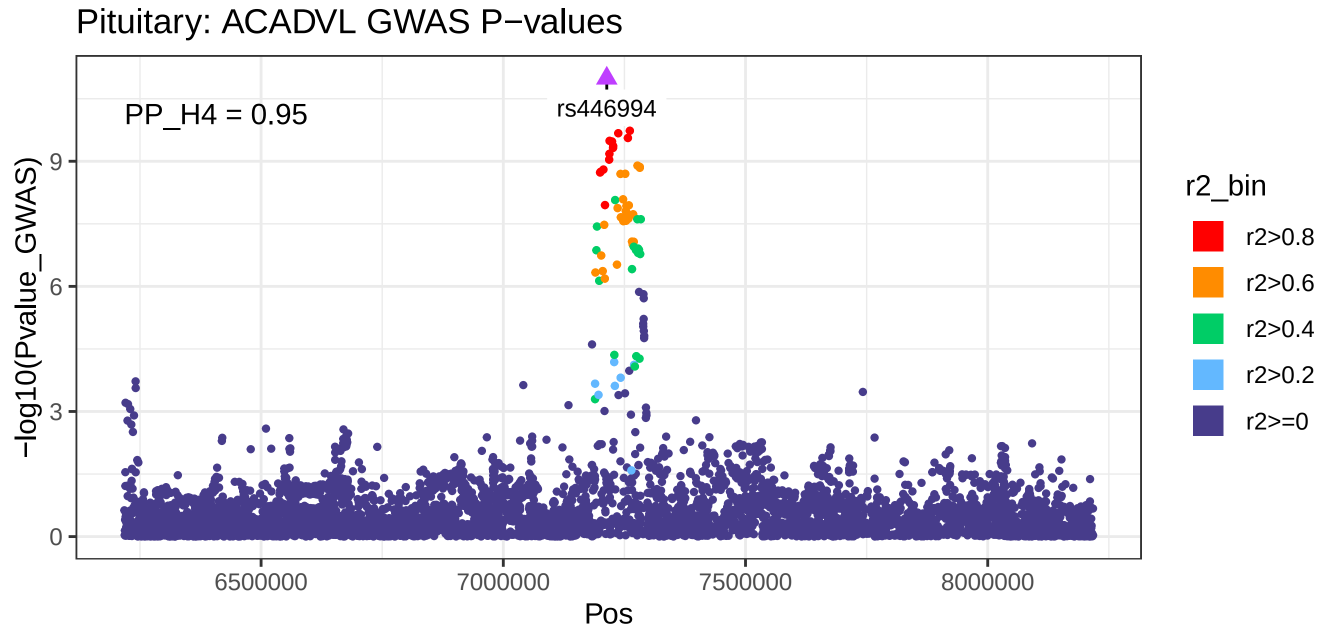
Pituitary: ACADVL GWAS P−values
PP_H4 = 0.95
rs446994
−log10(Pvalue_GWAS)
0
3
6
9
6500000
7000000
7500000
8000000
Pos
r2_bin
r2>0.8
r2>0.6
r2>0.4
r2>0.2
r2>=0

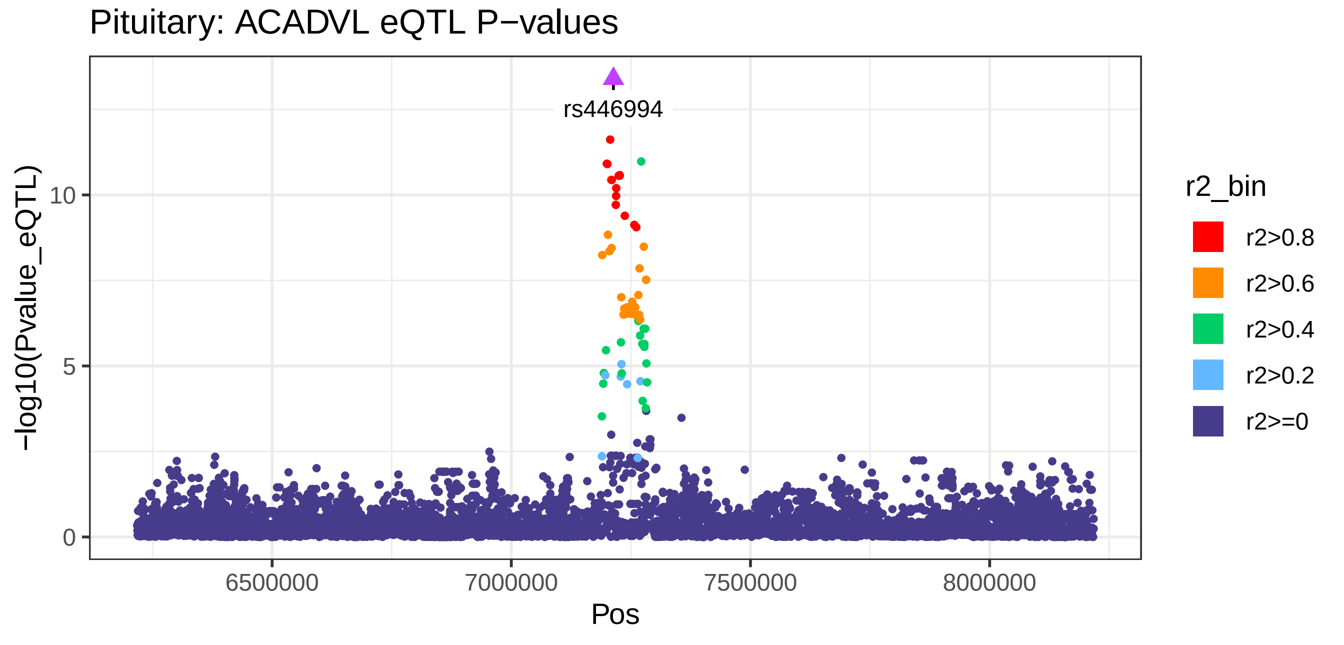
Pituitary: ACADVL eQTL P−values
rs446994
−log10(Pvalue_eQTL)
0
5
10
6500000
7000000
7500000
8000000
Pos
r2_bin
r2>0.8
r2>0.6
r2>0.4
r2>0.2
r2>=0

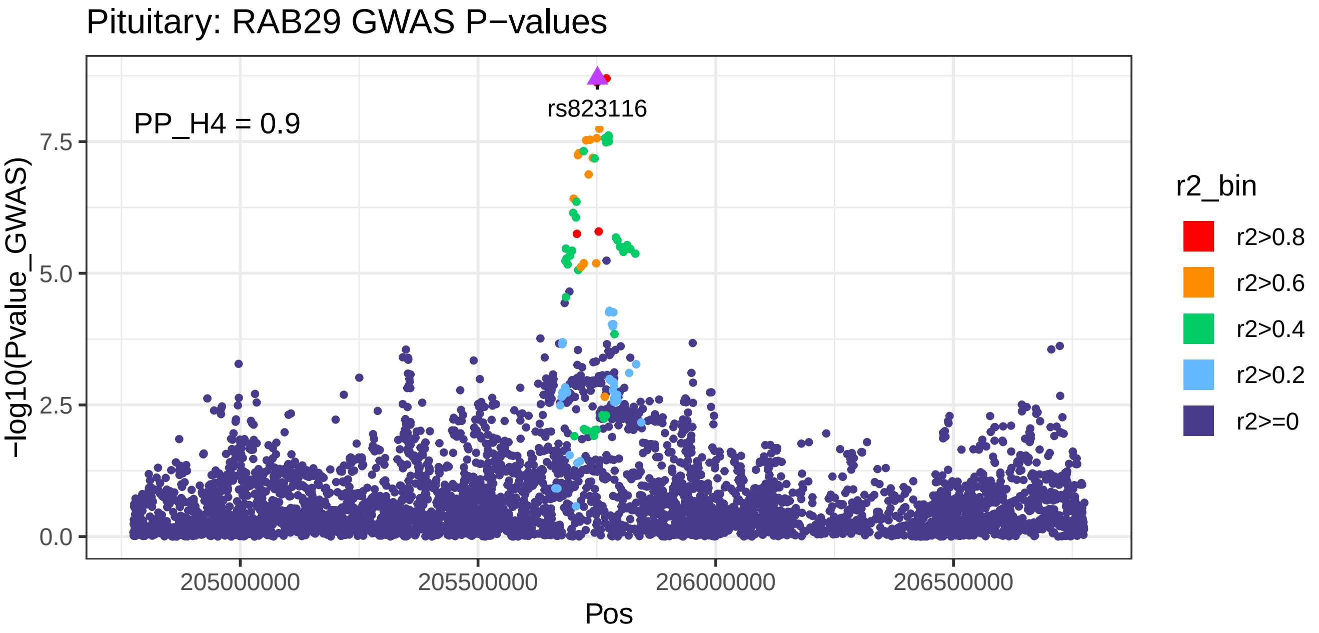
Pituitary: RAB29 GWAS P−values
PP_H4 = 0.9
rs823116
−log10(Pvalue_GWAS)
0.0
2.5
5.0
7.5
205000000
205500000
206000000
206500000
Pos
r2_bin
r2>0.8
r2>0.6
r2>0.4
r2>0.2
r2>=0

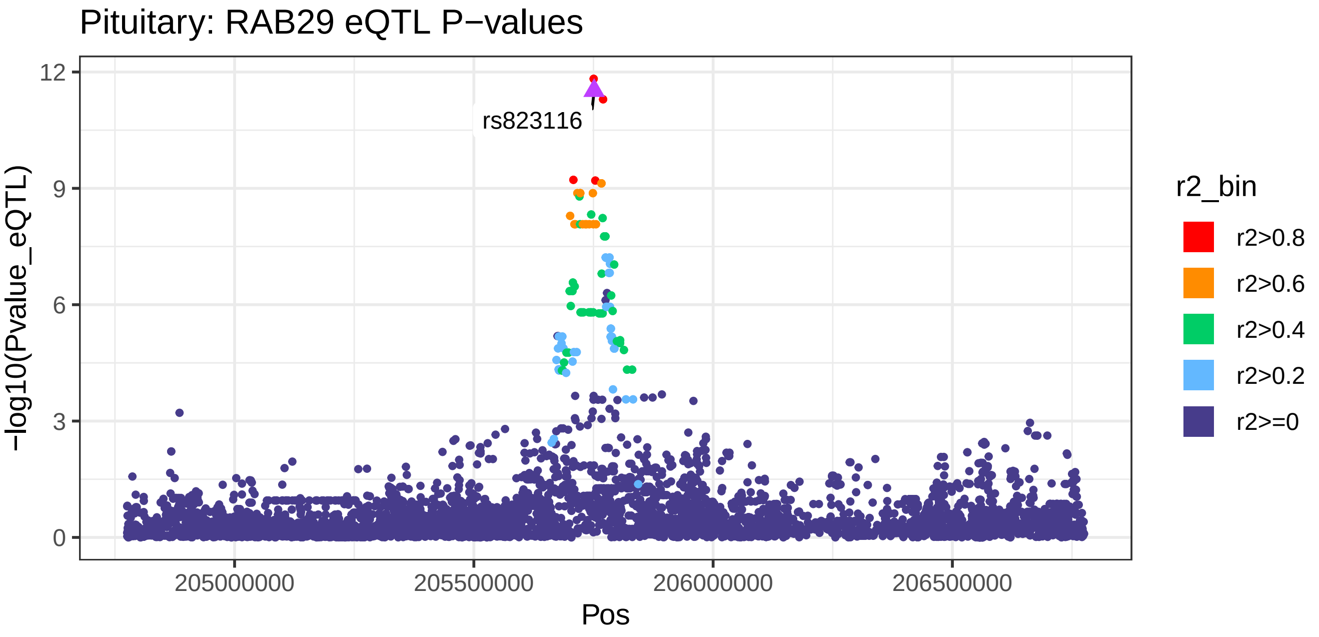
Pituitary: RAB29 eQTL P−values
rs823116
−log10(Pvalue_eQTL)
0
3
6
9
12
205000000
205500000
206000000
206500000
Pos
r2_bin
r2>0.8
r2>0.6
r2>0.4
r2>0.2
r2>=0

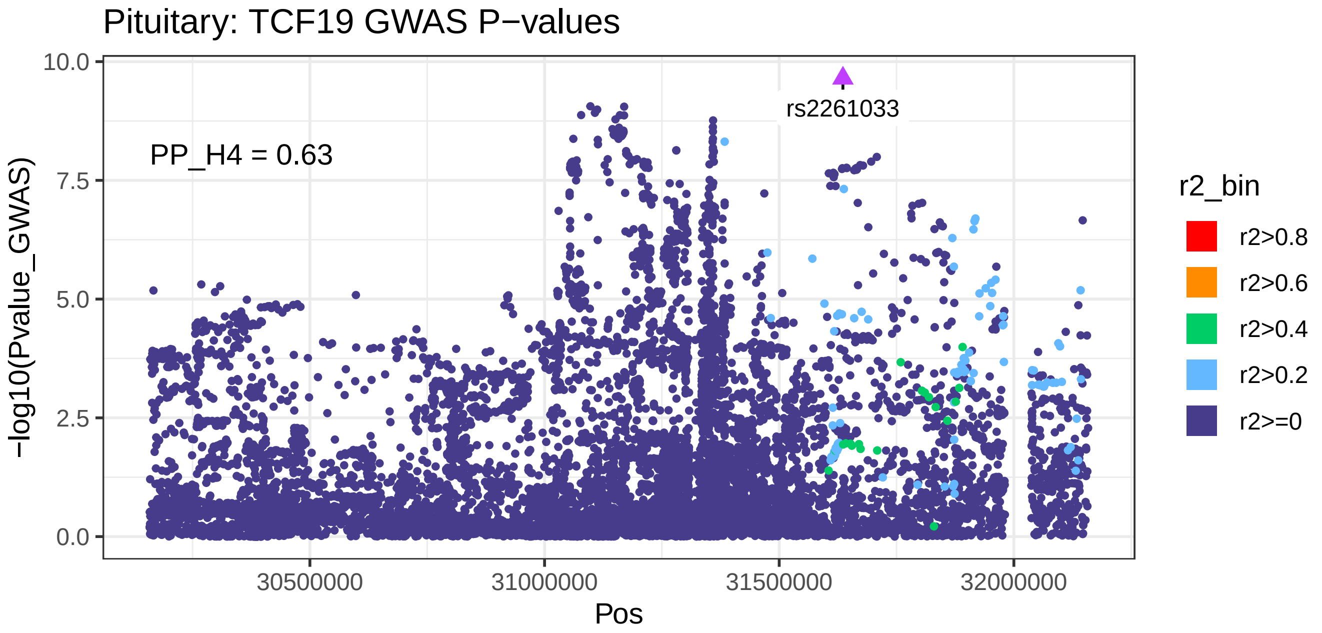
Pituitary: TCF19 GWAS P−values
rs2261033
PP_H4 = 0.63
10.0
7.5
5.0
2.5
0.0
−log10(Pvalue_GWAS)
30500000
31000000
31500000
32000000
Pos
r2_bin
r2>0.8
r2>0.6
r2>0.4
r2>0.2
r2>=0

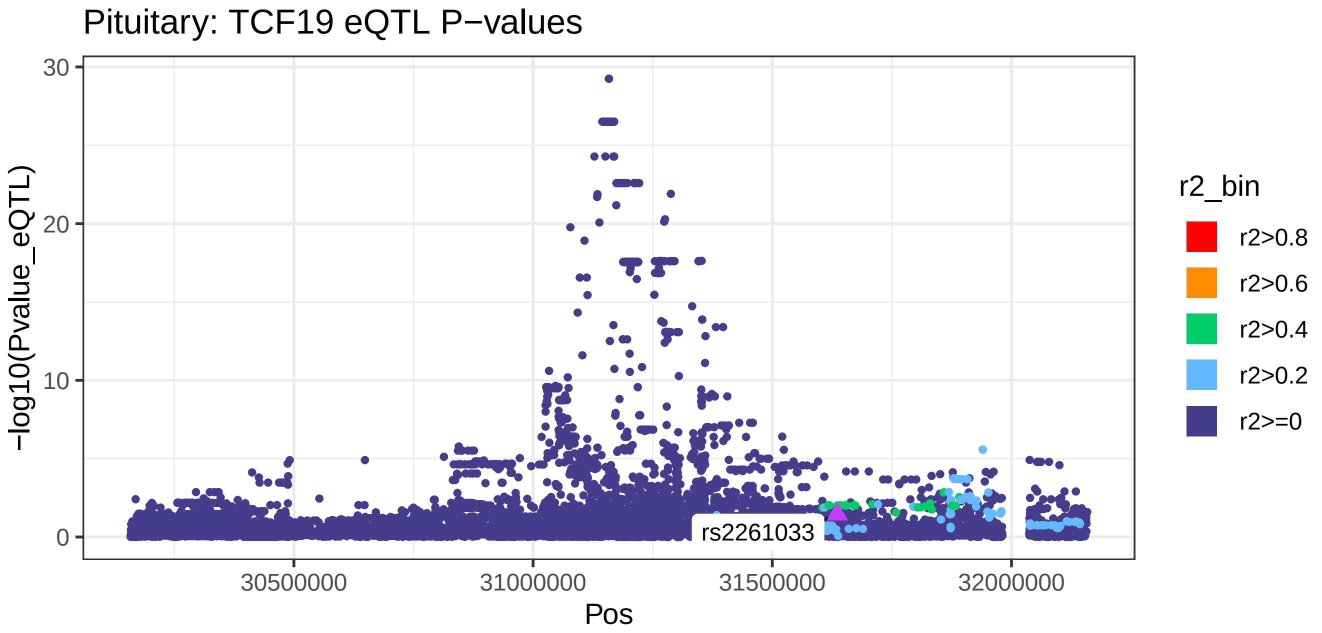
Pituitary: TCF19 eQTL P−values
30
20
10
0
−log10(Pvalue_eQTL)
rs2261033
30500000
31000000
31500000
32000000
Pos
r2_bin
r2>0.8
r2>0.6
r2>0.4
r2>0.2
r2>=0

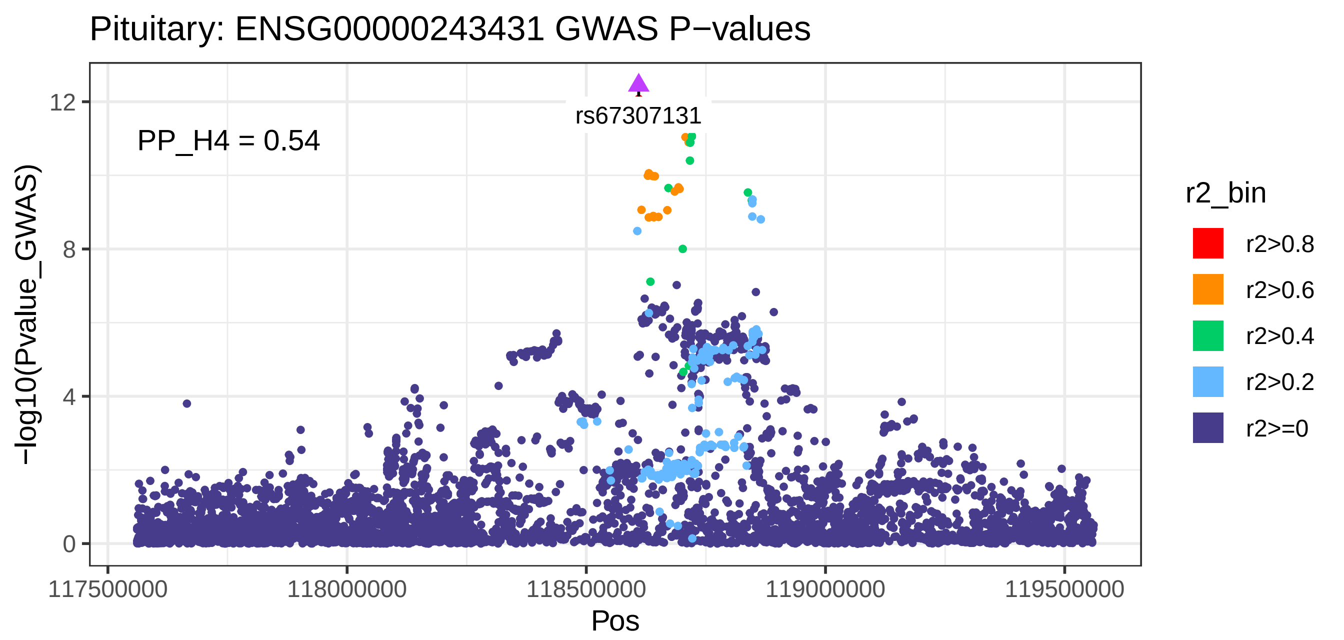
Pituitary: ENSG00000243431 GWAS P−values
PP_H4 = 0.54
rs67307131
−log10(Pvalue_GWAS)
12
8
4
0
117500000
118000000
118500000
119000000
119500000
Pos
r2_bin
r2>0.8
r2>0.6
r2>0.4
r2>0.2
r2>=0

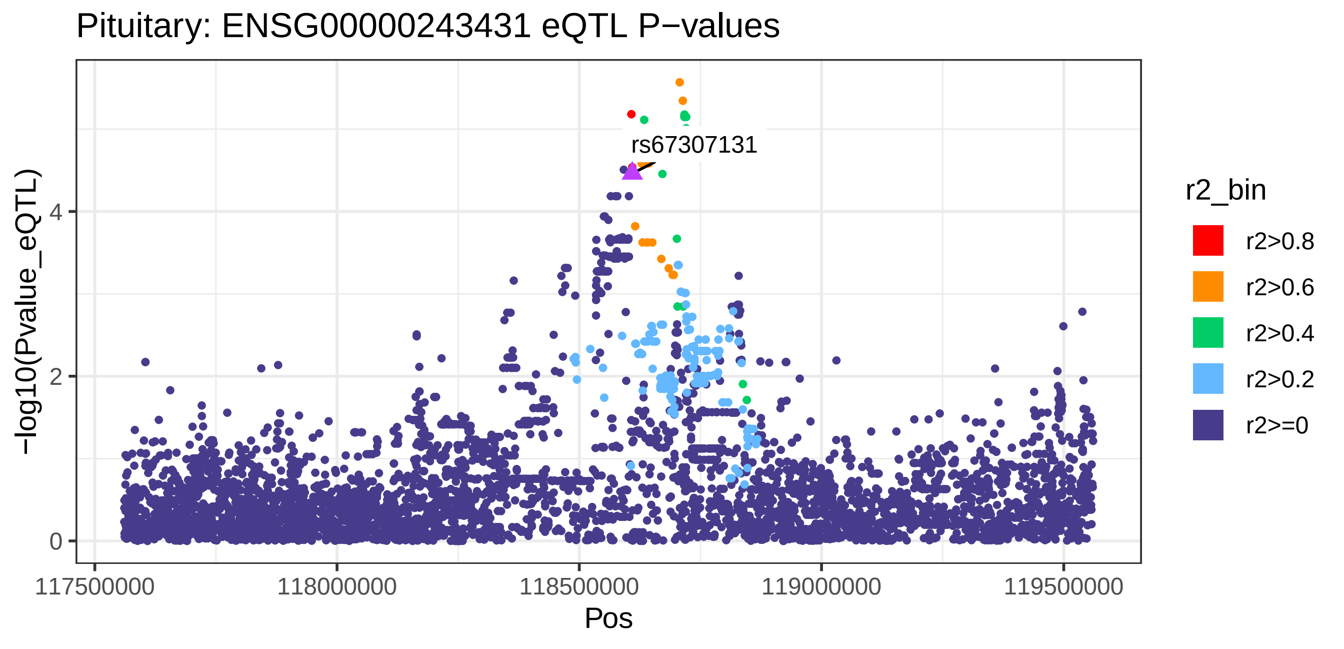
Pituitary: ENSG00000243431 eQTL P−values
rs67307131
−log10(Pvalue_eQTL)
4
2
0
117500000
118000000
118500000
119000000
119500000
Pos
r2_bin
r2>0.8
r2>0.6
r2>0.4
r2>0.2
r2>=0

Prostate: RAB29 GWAS P−values

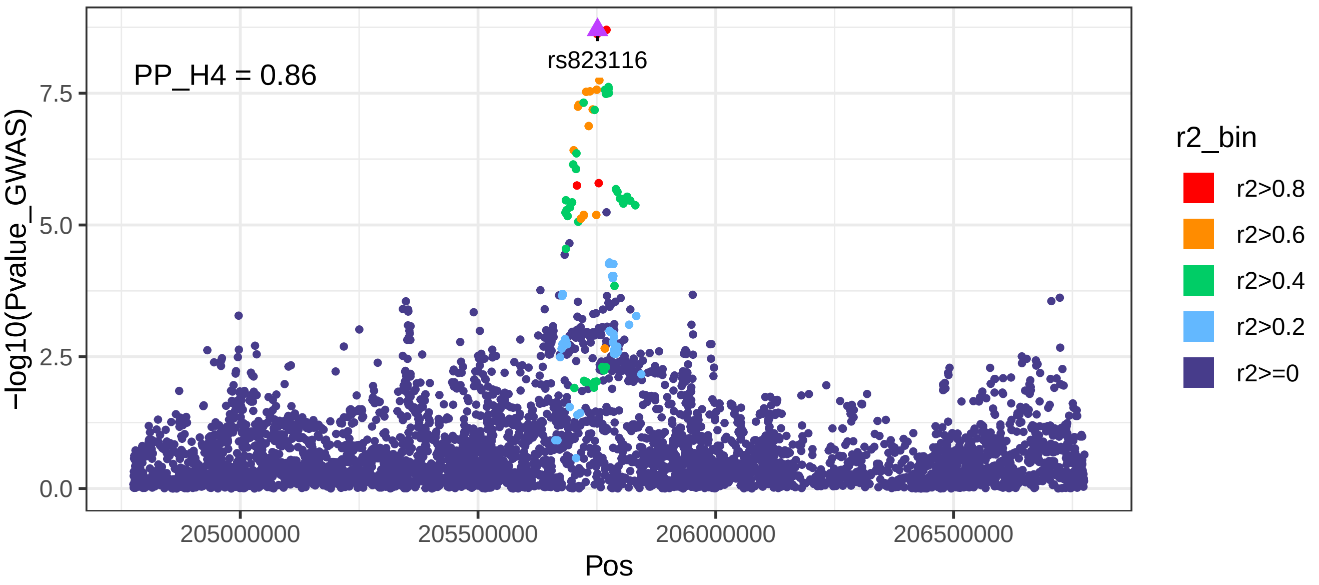


Prostate: RAB29 eQTL P−values

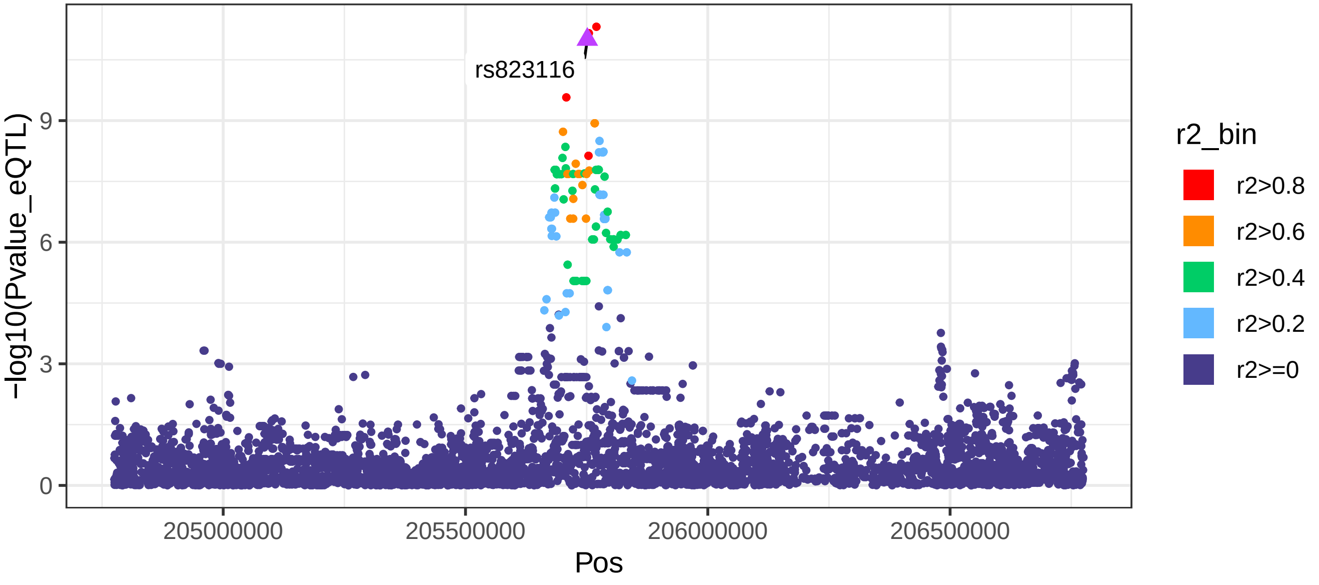

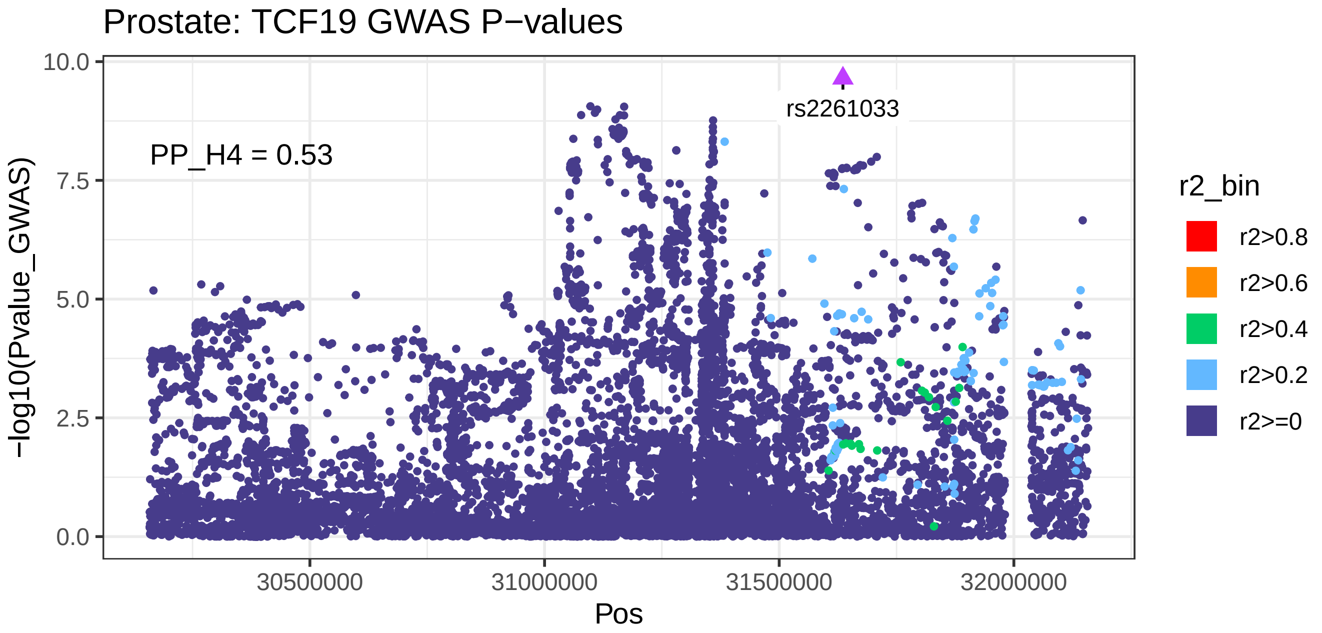
Prostate: TCF19 GWAS P−values
PP_H4 = 0.53
rs2261033
−log10(Pvalue_GWAS)
0.0
2.5
5.0
7.5
10.0
30500000
31000000
31500000
32000000
Pos
r2_bin
r2>0.8
r2>0.6
r2>0.4
r2>0.2
r2>=0

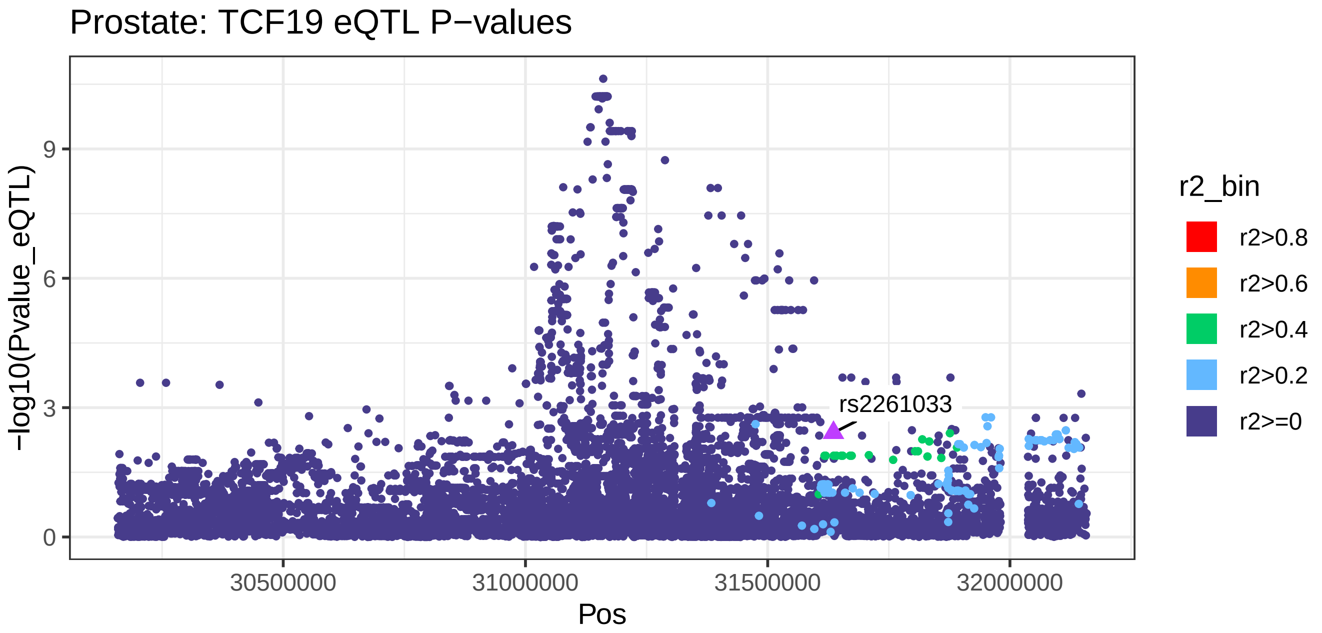
Prostate: TCF19 eQTL P−values
rs2261033
−log10(Pvalue_eQTL)
0
3
6
9
30500000
31000000
31500000
32000000
Pos
r2_bin
r2>0.8
r2>0.6
r2>0.4
r2>0.2
r2>=0

Small_Intestine_Terminal_Ileum: RAB29 GWAS P−values

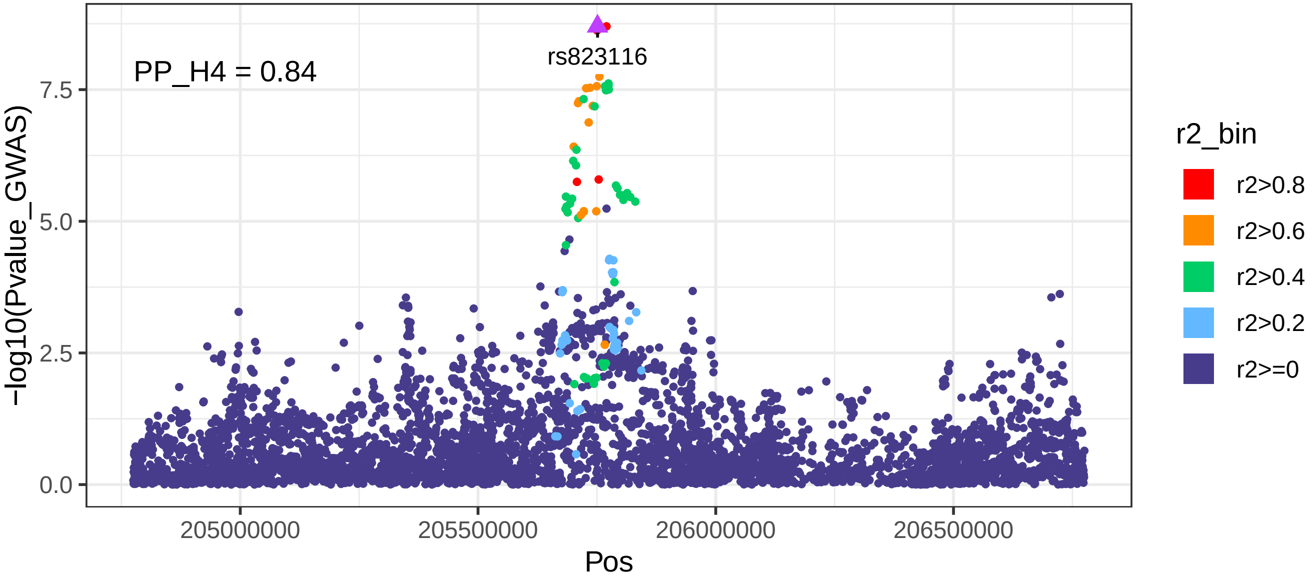


Small_Intestine_Terminal_Ileum: RAB29 eQTL P−values

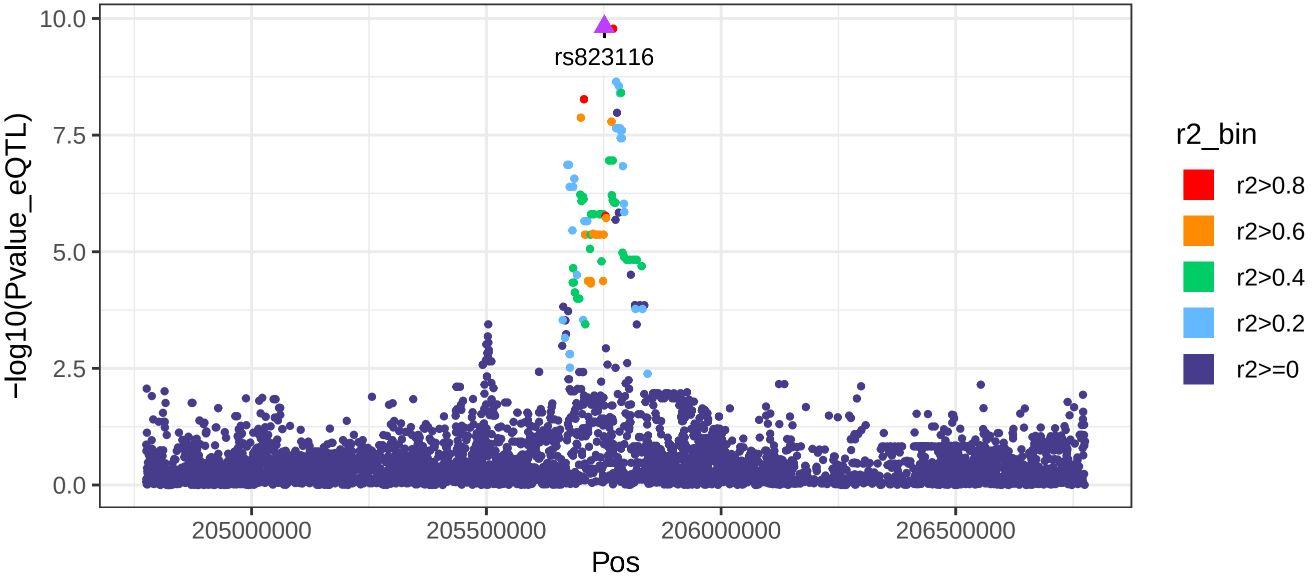

Testis: NUCKS1 GWAS P−values

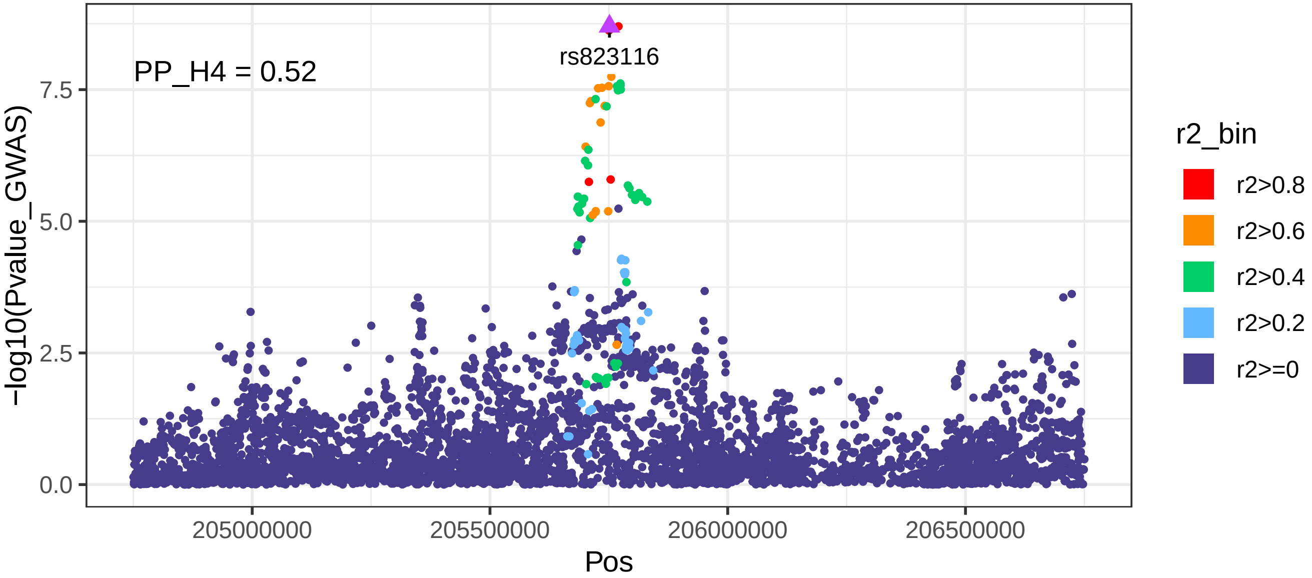


Testis: NUCKS1 eQTL P−values

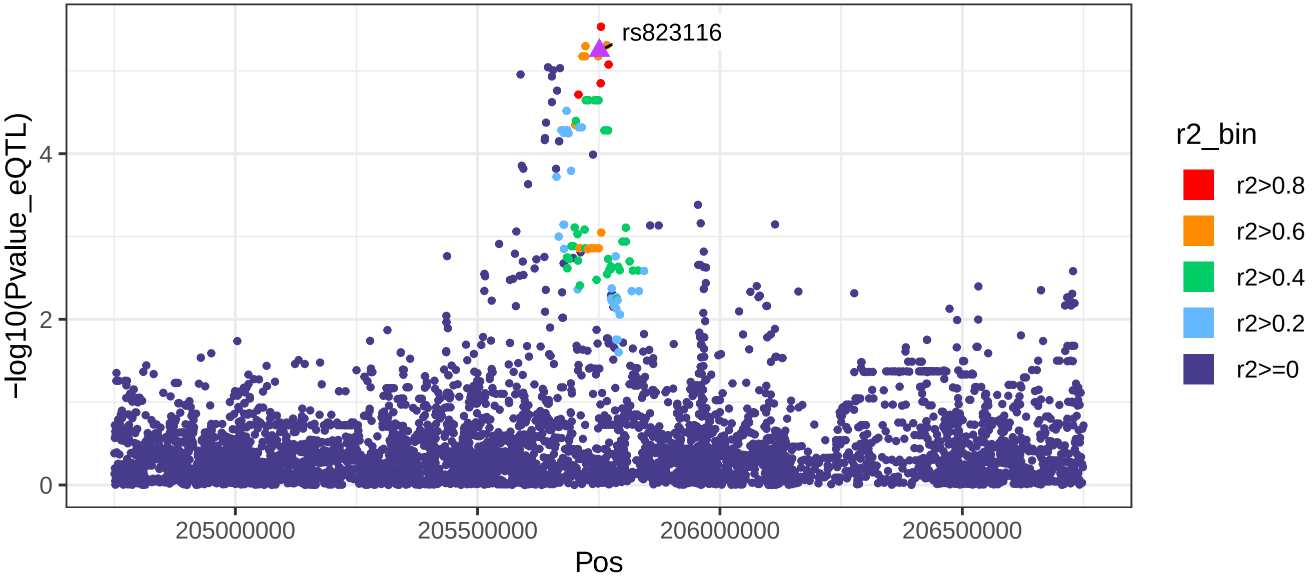

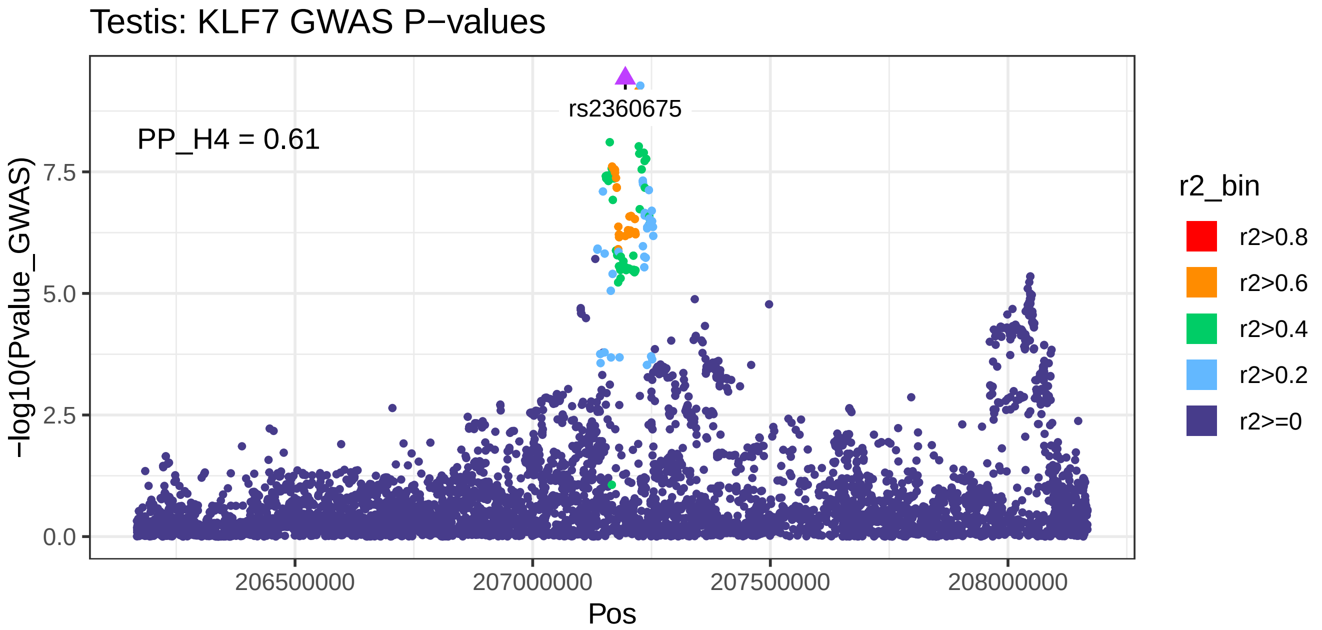
Testis: KLF7 GWAS P−values
rs2360675
PP_H4 = 0.61
−log10(Pvalue_GWAS)
0.0
2.5
5.0
7.5
206500000
207000000
207500000
208000000
Pos
r2_bin
r2>0.8
r2>0.6
r2>0.4
r2>0.2
r2>=0

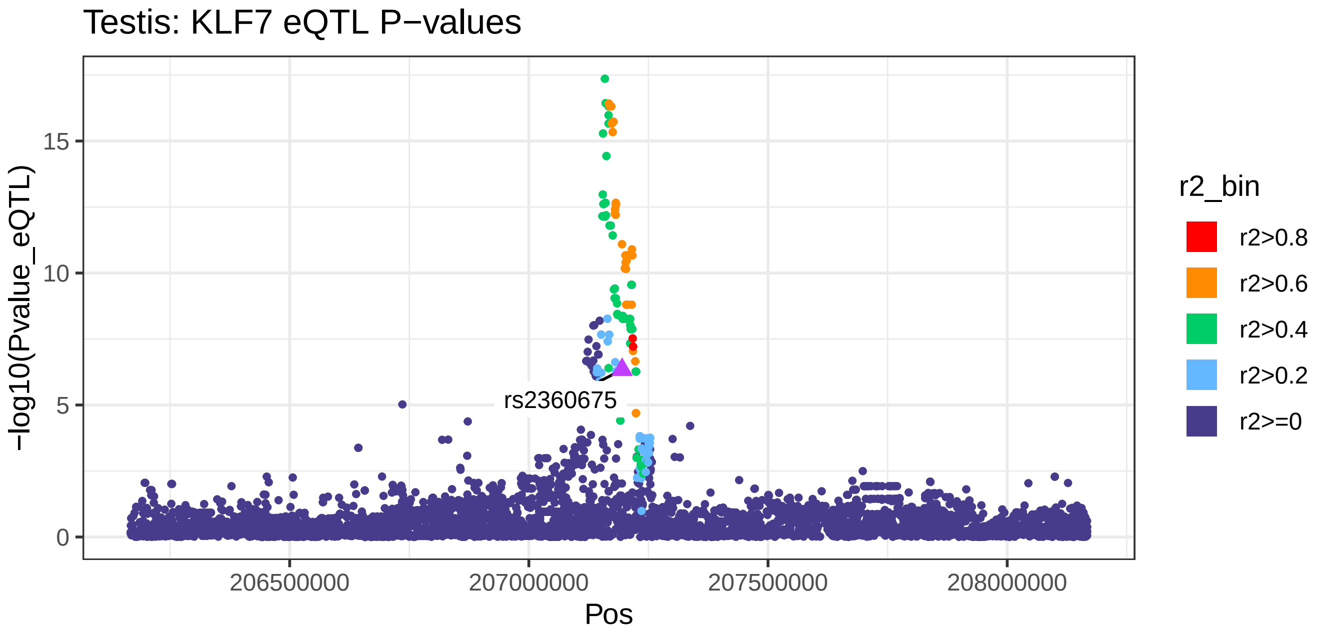
Testis: KLF7 eQTL P−values
rs2360675
−log10(Pvalue_eQTL)
0
5
10
15
206500000
207000000
207500000
208000000
Pos
r2_bin
r2>0.8
r2>0.6
r2>0.4
r2>0.2
r2>=0

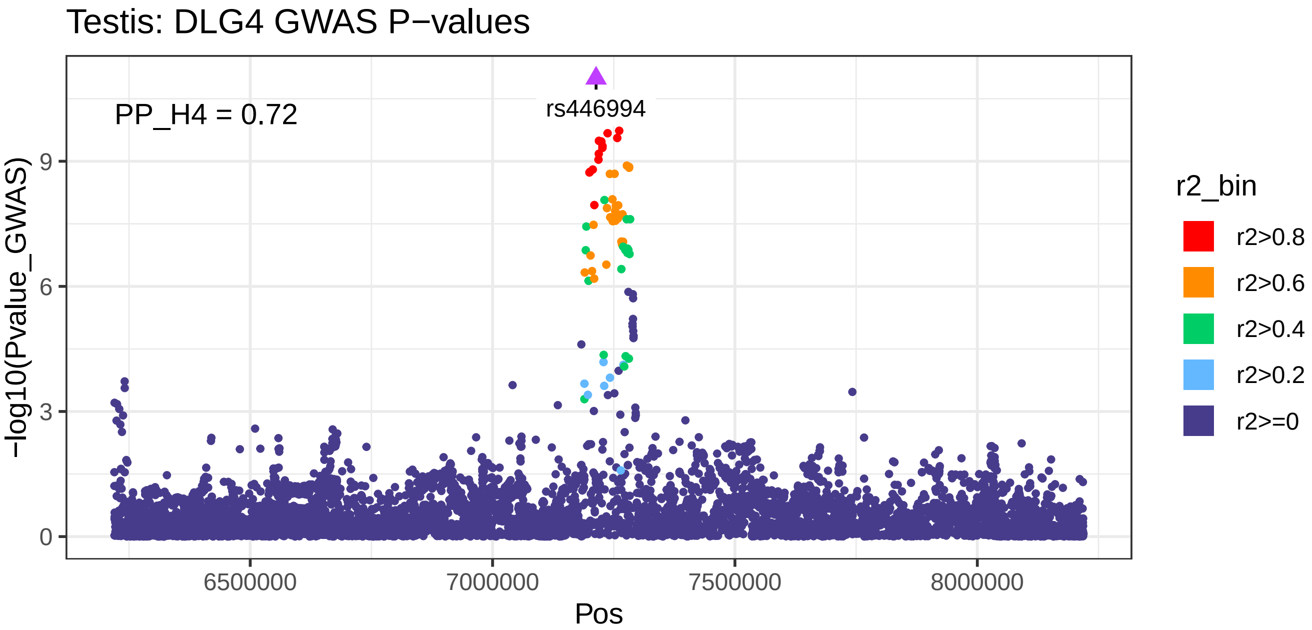
Testis: DLG4 GWAS P−values
PP_H4 = 0.72
rs446994
−log10(Pvalue_GWAS)
0
3
6
9
6500000
7000000
7500000
8000000
Pos
r2_bin
r2>0.8
r2>0.6
r2>0.4
r2>0.2
r2>=0

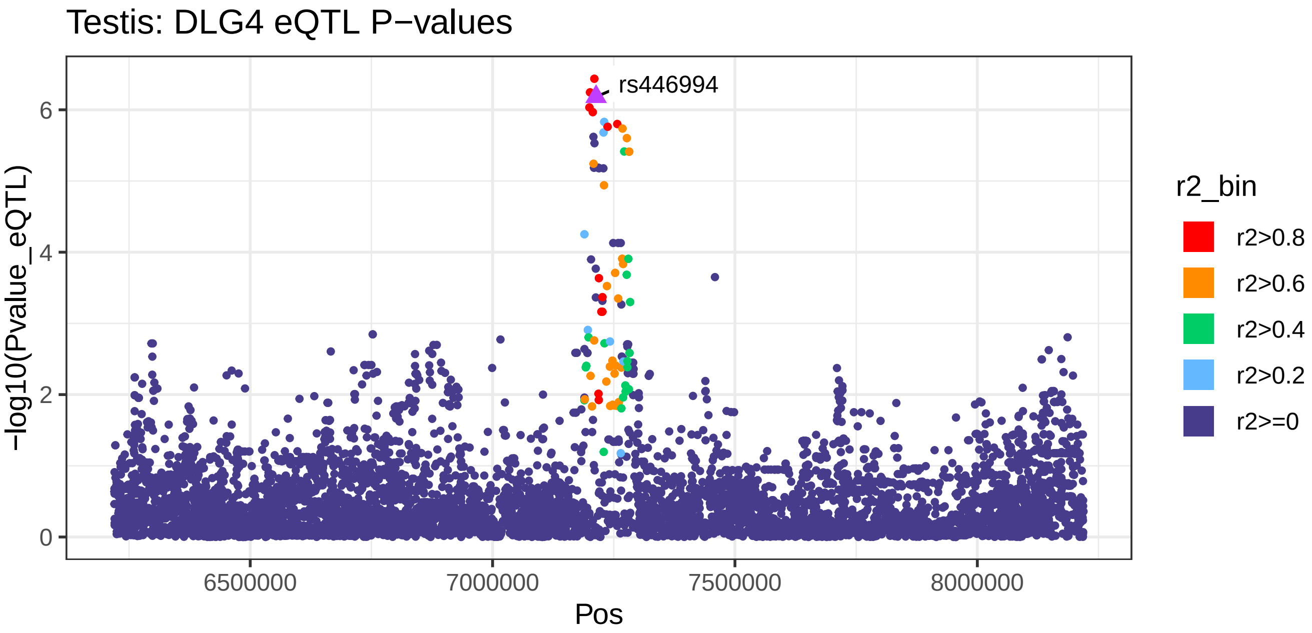
Testis: DLG4 eQTL P−values
rs446994
−log10(Pvalue_eQTL)
0
2
4
6
6500000
7000000
7500000
8000000
Pos
r2_bin
r2>0.8
r2>0.6
r2>0.4
r2>0.2
r2>=0

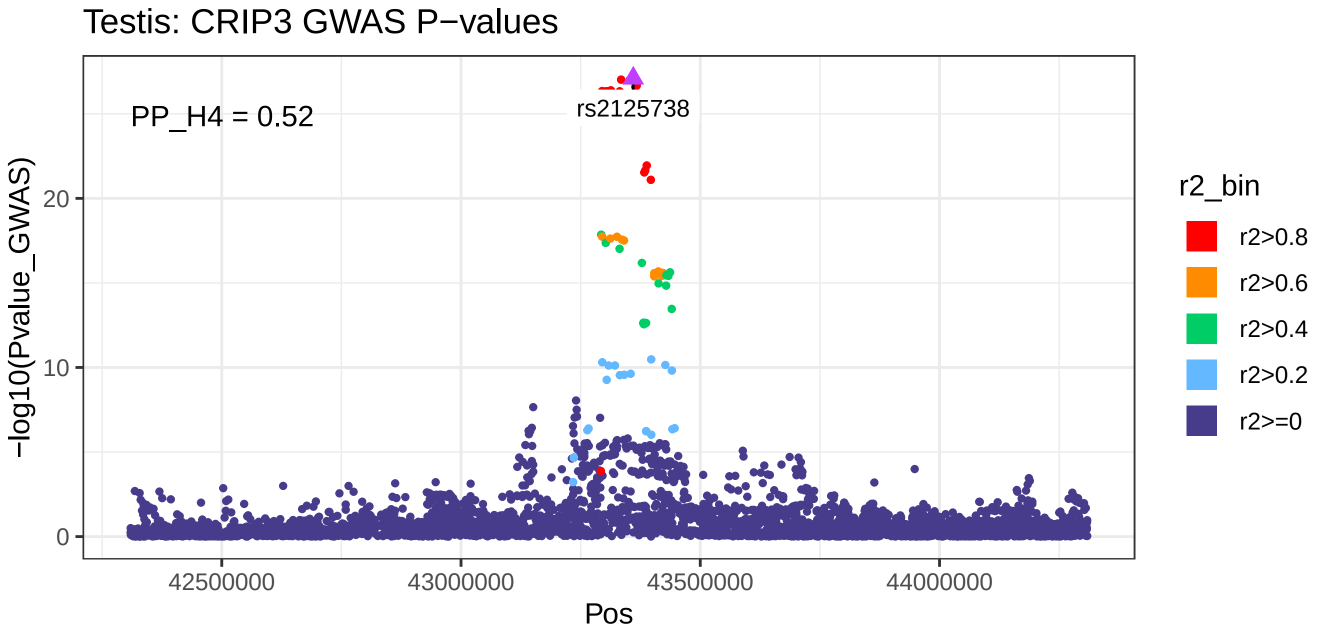
Testis: CRIP3 GWAS P−values
PP_H4 = 0.52
rs2125738
−log10(Pvalue_GWAS)
0
10
20
42500000
43000000
43500000
44000000
Pos
r2_bin
r2>0.8
r2>0.6
r2>0.4
r2>0.2
r2>=0

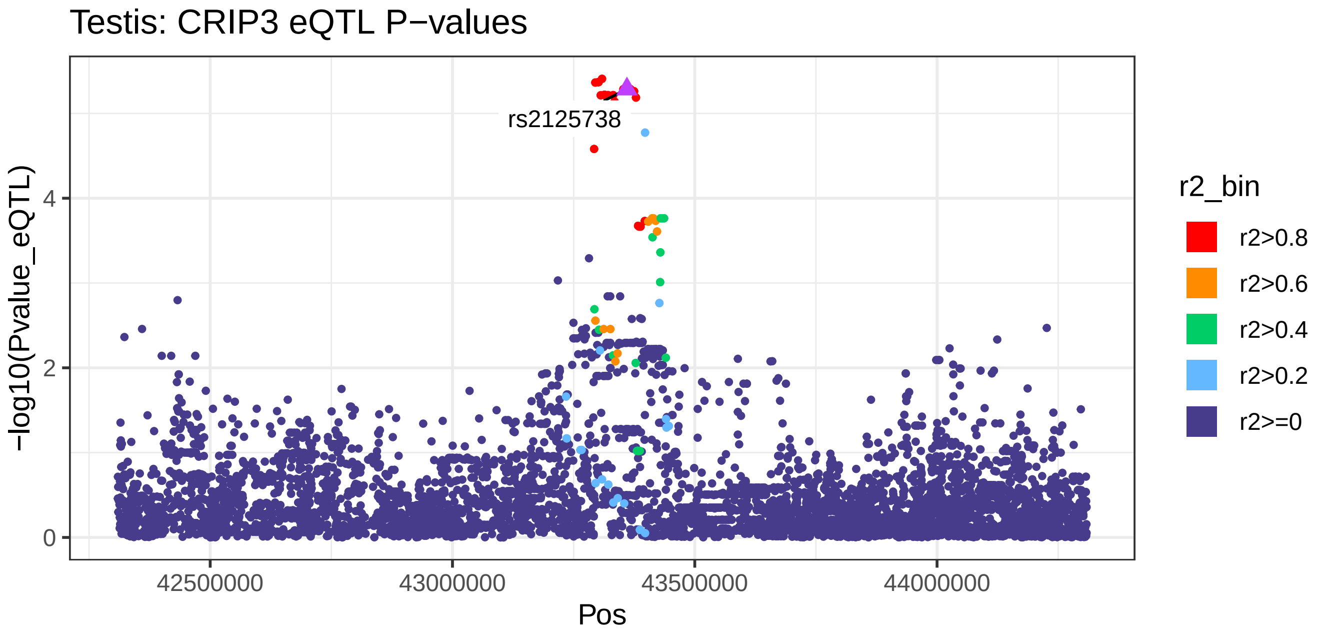
Testis: CRIP3 eQTL P−values
rs2125738
−log10(Pvalue_eQTL)
0
2
4
42500000
43000000
43500000
44000000
Pos
r2_bin
r2>0.8
r2>0.6
r2>0.4
r2>0.2
r2>=0

Thyroid: SPTBN1 GWAS P−values

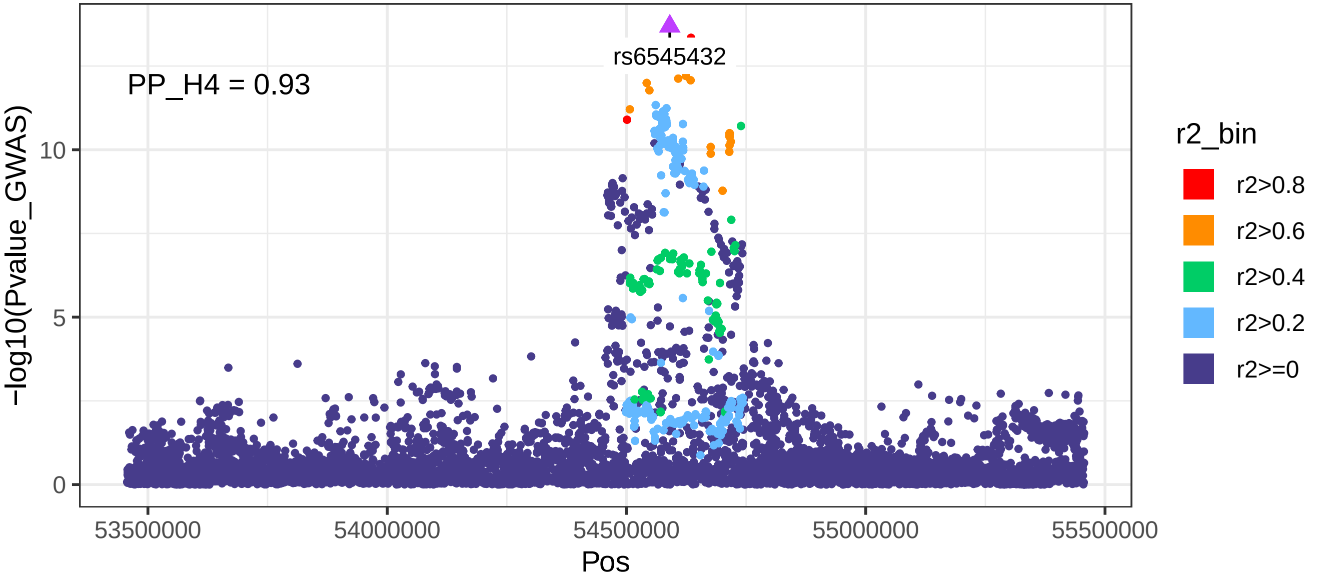


Thyroid: SPTBN1 eQTL P−values

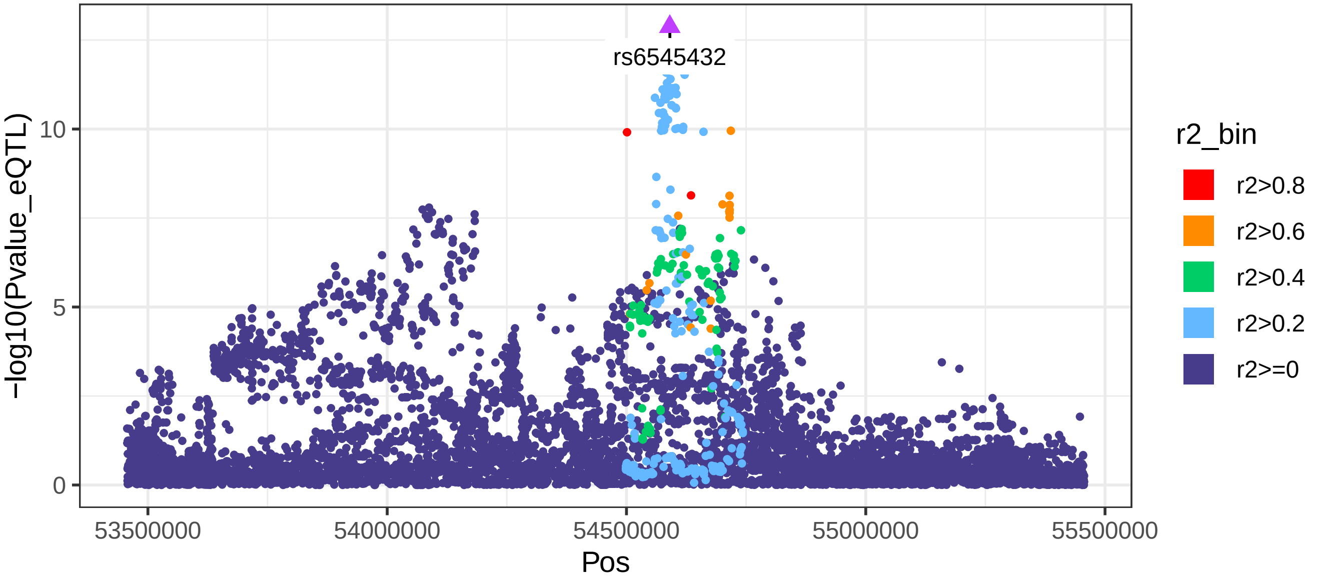

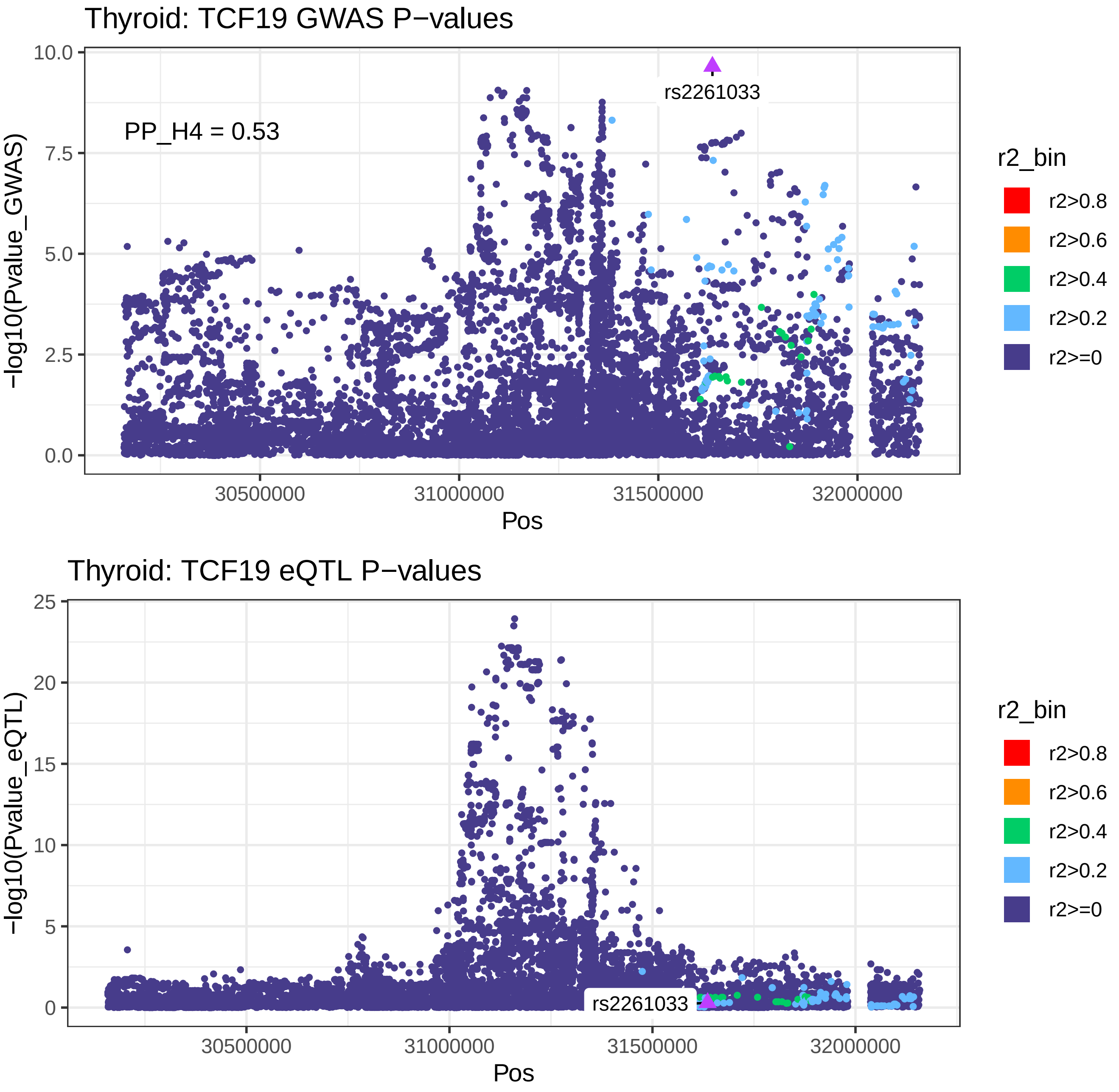
Thyroid: TCF19 GWAS P−values
PP_H4 = 0.53
rs2261033
−log10(Pvalue_GWAS)
10.0
7.5
5.0
2.5
0.0
30500000
31000000
31500000
32000000
Pos
r2_bin
r2>0.8
r2>0.6
r2>0.4
r2>0.2
r2>=0
Thyroid: TCF19 eQTL P−values
rs2261033
−log10(Pvalue_eQTL)
25
20
15
10
5
0
30500000
31000000
31500000
32000000
Pos
r2_bin
r2>0.8
r2>0.6
r2>0.4
r2>0.2
r2>=0

Thyroid: CYP26C1 GWAS P−values

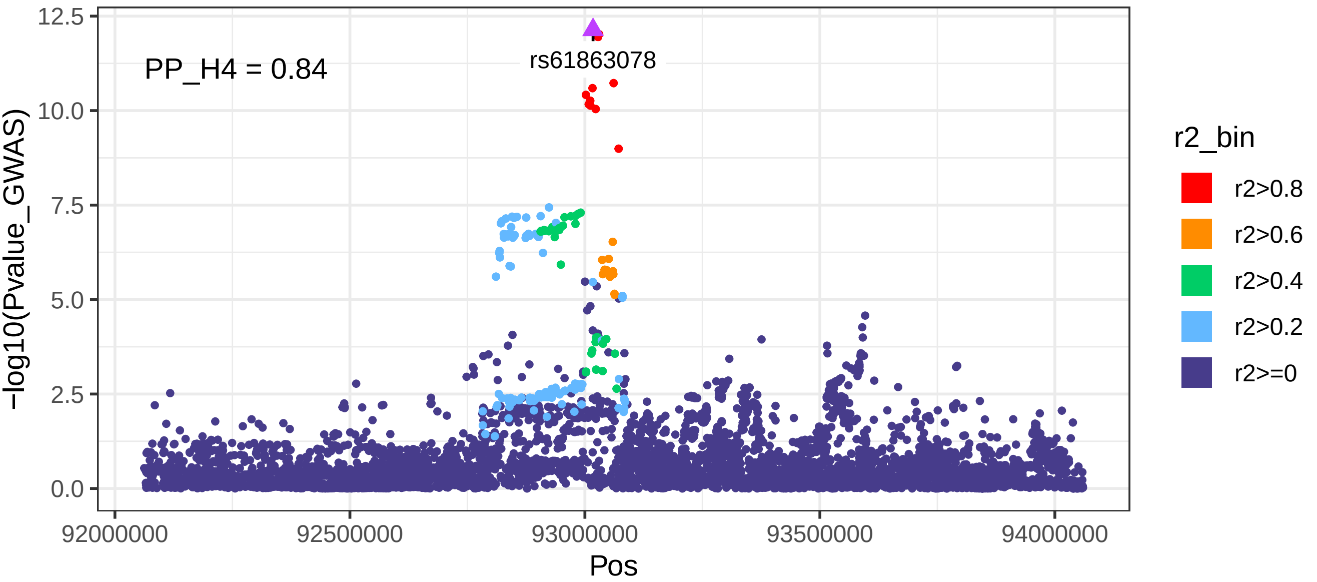


Thyroid: CYP26C1 eQTL P−values

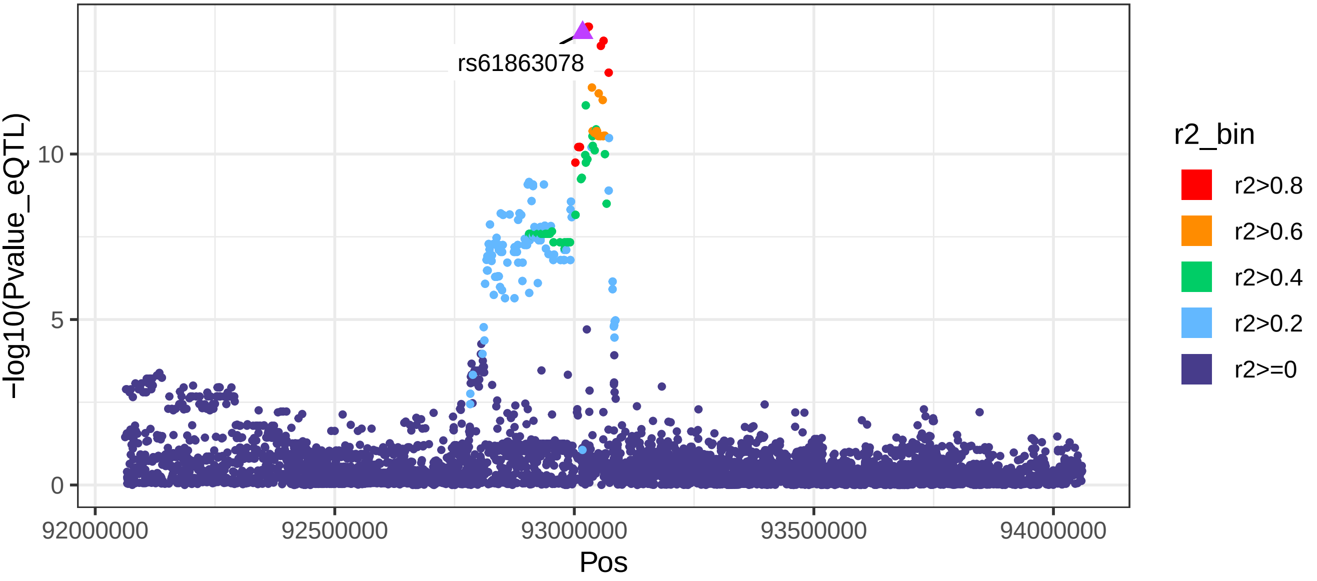

Whole_Blood: ACADVL GWAS P−values

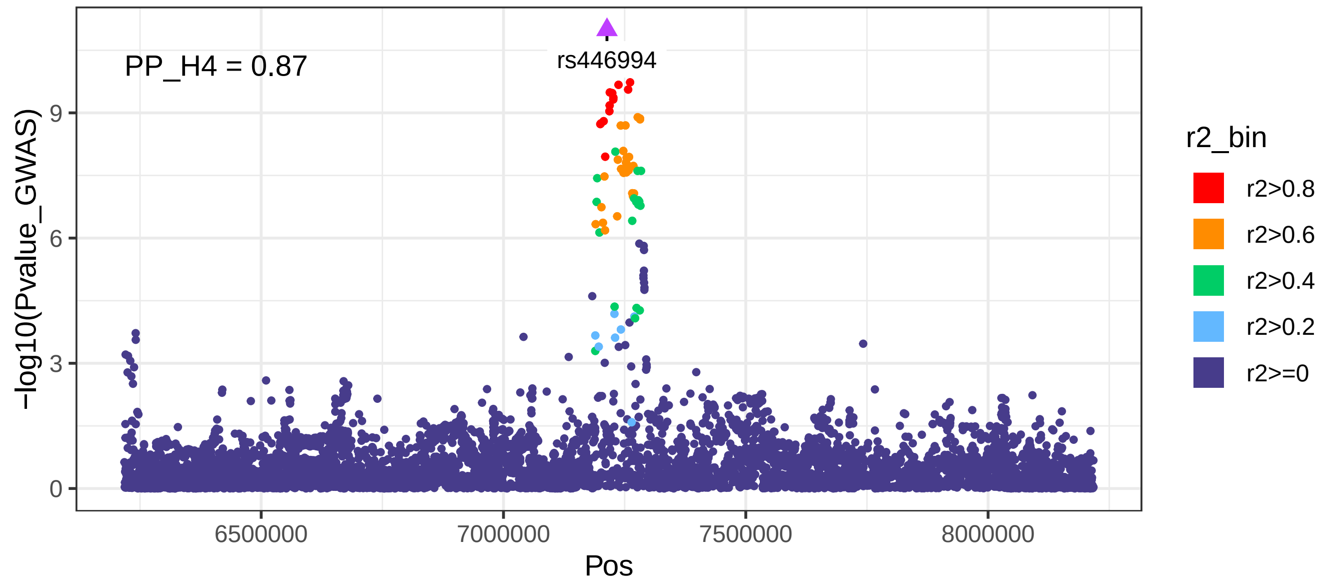


Whole_Blood: ACADVL eQTL P−values

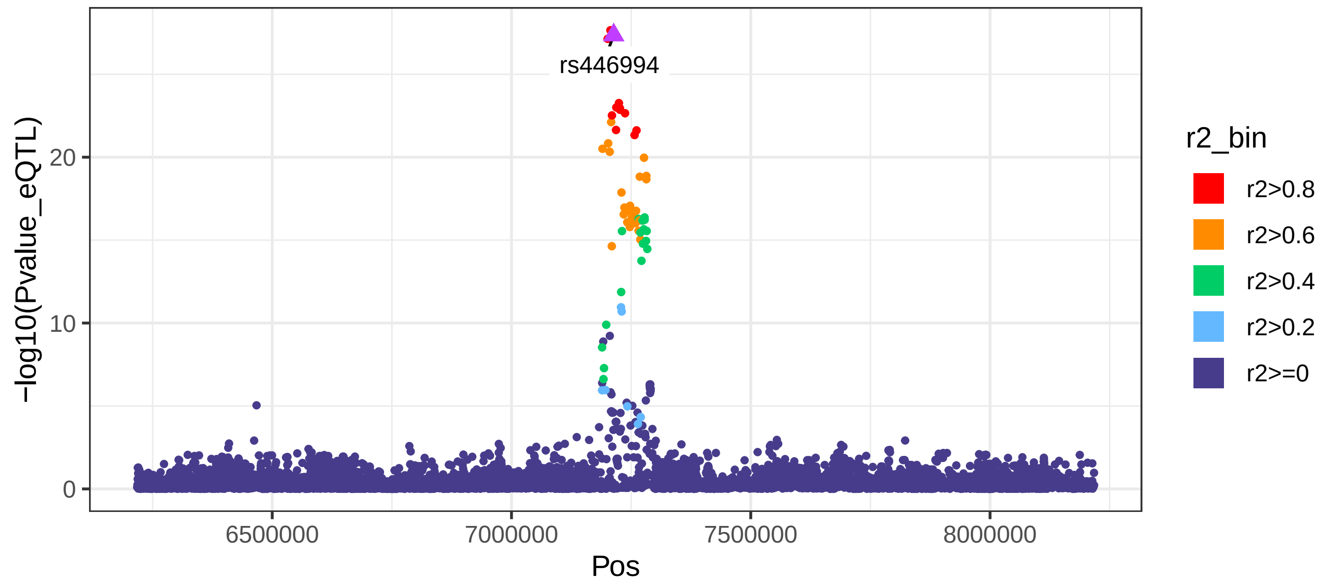

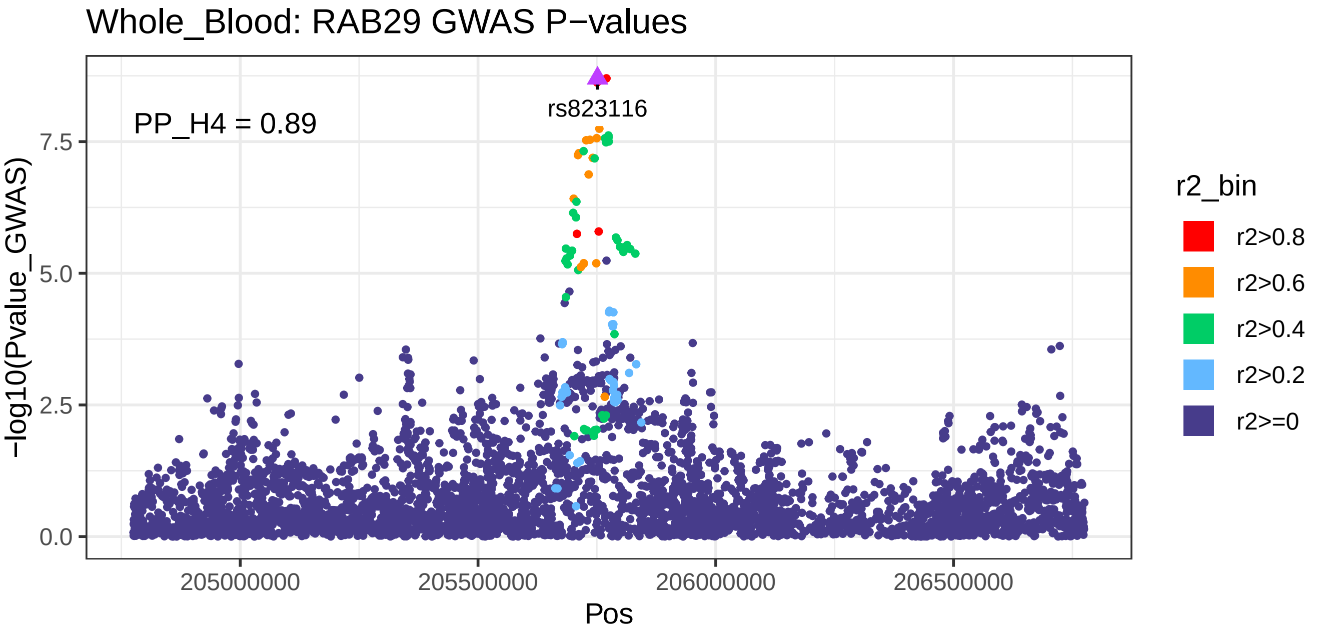
Whole_Blood: RAB29 GWAS P−values
PP_H4 = 0.89
rs823116
−log10(Pvalue_GWAS)
0.0
2.5
5.0
7.5
205000000
205500000
206000000
206500000
Pos
r2_bin
r2>0.8
r2>0.6
r2>0.4
r2>0.2
r2>=0

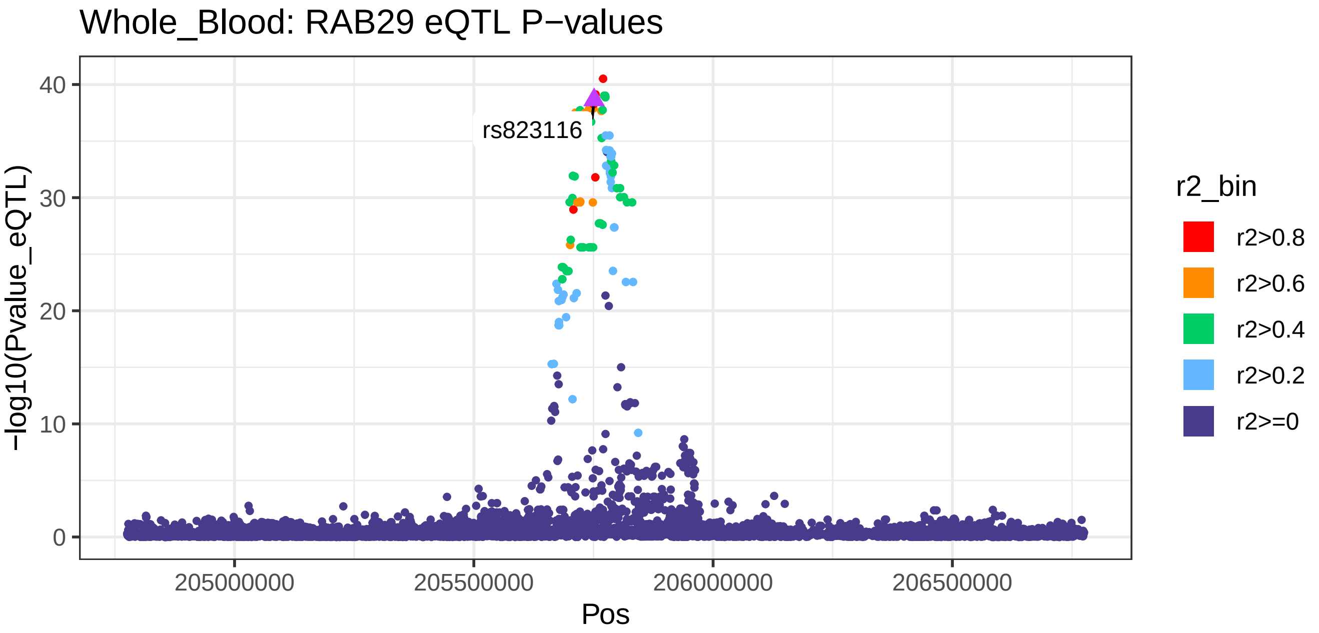
Whole_Blood: RAB29 eQTL P−values
rs823116
−log10(Pvalue_eQTL)
0
10
20
30
40
205000000
205500000
206000000
206500000
Pos
r2_bin
r2>0.8
r2>0.6
r2>0.4
r2>0.2
r2>=0

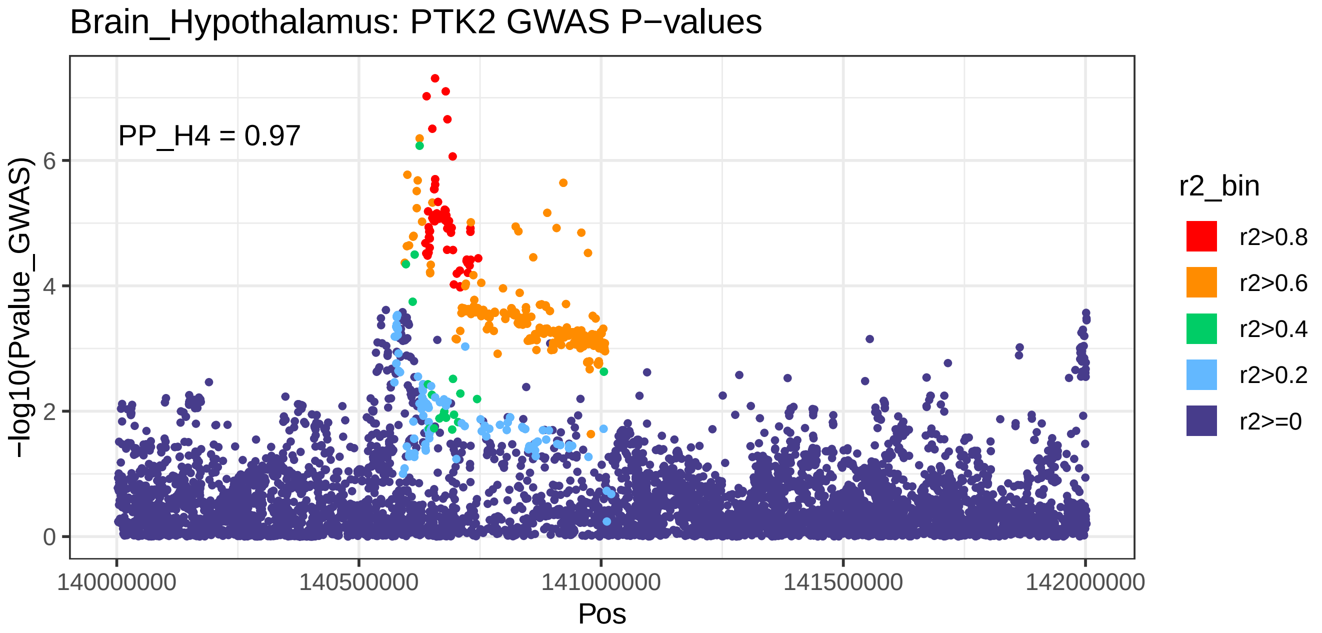
Brain_Hypothalamus: PTK2 GWAS P−values
PP_H4 = 0.97
−log10(Pvalue_GWAS)
0
2
4
6
140000000
140500000
141000000
141500000
142000000
Pos
r2_bin
r2>0.8
r2>0.6
r2>0.4
r2>0.2
r2>=0

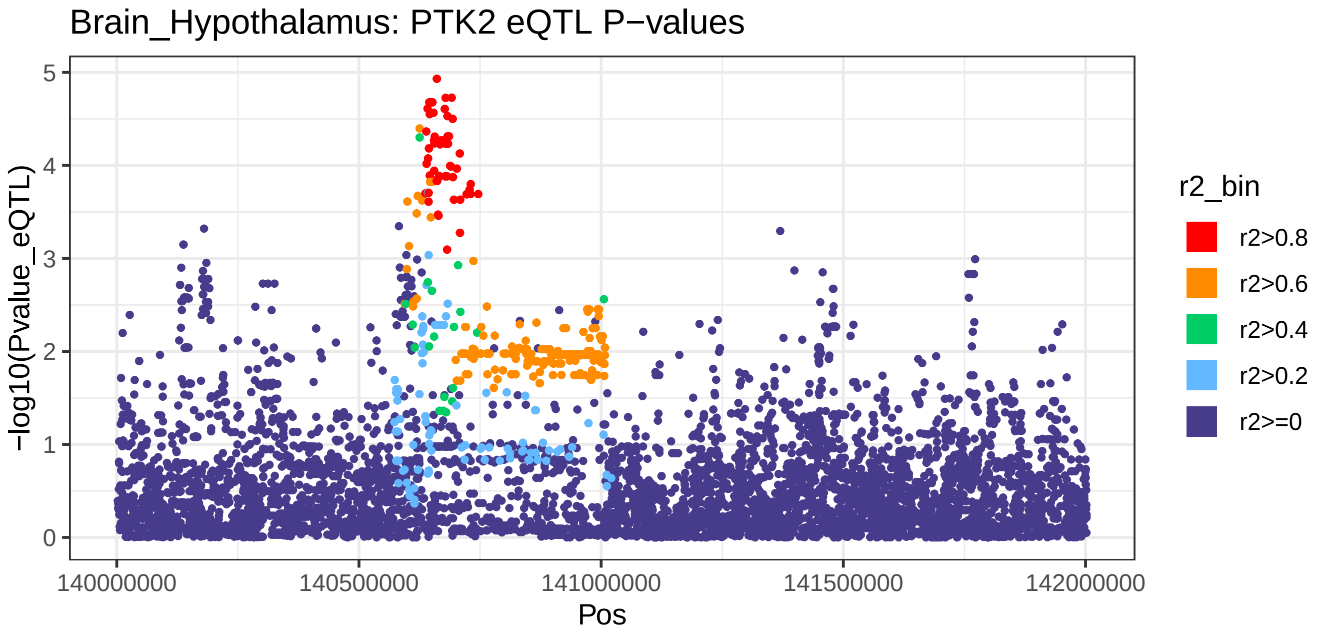
Brain_Hypothalamus: PTK2 eQTL P−values
−log10(Pvalue_eQTL)
0
1
2
3
4
5
140000000
140500000
141000000
141500000
142000000
Pos
r2_bin
r2>0.8
r2>0.6
r2>0.4
r2>0.2
r2>=0
